# Five-year immunogenicity and safety follow-up of the PREVAC randomized Trial of Vaccines for Zaire Ebola Virus Disease

**DOI:** 10.64898/2026.05.29.26354050

**Authors:** Abdoul Habib Beavogui, Seydou Doumbia, Mark Kieh, Bailah Leigh, Samba Sow, Edouard Lhomme, Safaa Ben-Farhat, Natasha Dubois Cauwelaert, Céline Roy, Waly Diouf, Sesay Idrissa, Samba Diarra, Niouma Pascal Millimouno, Fatoumata Abdoulaye Diallo, Michael Kamara, Dudley Pratt, Ilo Dicko, Stephen B. Kennedy, Hélène Espérou, Edward M. Choi, Ange-Marie D. Kpetigo, Eric D’Ortenzio, Alpha Diallo, Solange Lancrey-javal, Benjamin Hamzé, Christine Schwimmer, Aurélie Wiedemann, Ahidjo Ayouba, Martine Peeters, H. Clifford Lane, Elisabeth Higgs, Deborah Watson-Jones, Yazdan Yazdanpanah, Brian Greenwood, Laura Richert, Yves Levy, PREVAC study Team

## Abstract

**Background:** The World Health Organization has expanded its recommendations for prophylactic Ebola vaccination for at-risk populations. Durable vaccine-induced immunity is important for sustaining outbreak preparedness in regions with recurrent Ebola virus disease (EVD). We assessed five-year persistence of vaccine-induced immune responses in adults and children from the PREVAC trial.

**Methods:** Two large randomised phase 2 trials (NCT02876328), in adults and children aged ≥ 1 year, were conducted in four west African countries. Participants were randomly assigned to placebo or to one of three Ebola vaccine strategies: Ad26.ZEBOV followed by MVA-BN-Filo at 56 days; rVSVΔG-ZEBOV-GP followed by placebo; or rVSVΔG-ZEBOV-GP followed by a homologous booster dose at 56 days. After 12 months of follow-up, the primary results were published, participants unblinded to their vaccine assignment, and follow-up continued for 60 months. After Month 24, placebo group recipients were offered active vaccination. Anti–Ebola virus glycoprotein Immunoglobulin G (IgG) concentrations were measured for 5 years.

**Findings:** 1401 adults and 1401 children were initially randomized, and 1315 (93.9%) adults and 1322 (94.4%) children attended at least one long-term visit. Retention was high, with 95% followed beyond 1 year and 83% completion at 5-year follow-up. For the three vaccine strategies, antibody geometric mean concentrations (GMC) declined modestly between Months 12 and 24, followed by a stable plateau from Months 24 to 60. At Month 60, antibody GMC were higher in the rVSV-based groups (1099 and 1216 EU/ml for adults; 1982 and 2347 EU/ml for children) than in the Ad26.ZEBOV, MVA-BN-Filo group (252 adults and 645 EUml children). Antibody persistence at Month 60 was heterogeneous, varying by age, sex, country, and baseline IgG concentration.

**Interpretation:** Licensed Ebola vaccines induced sustained antibody responses in adults and children for up to 5 years. While the protective antibody level is unknown, these data demonstrate long-lasting immune responses from currently employed vaccine strategies.

**Trial registration numbers:** ClinicalTrials.gov number, NCT02876328; EudraCT number 2017-001798-18; and Pan African Clinical Trials Registry number, PACTR201712002760250.

## Introduction

Since its first identification in 1976, Ebola virus disease (EVD) has continued to pose a recurrent threat to global health, reported with increasing outbreak frequency over the past two decades. Major epidemics in West Africa (2013–16) and the Democratic Republic of the Congo (DRC; 2018–20) highlighted the substantial morbidity and mortality associated with EVD and reinforced the need for sustained outbreak preparedness strategies that extend beyond an emergency response. In this context, prophylactic vaccination of populations at risk has emerged as a key component of preparedness, particularly in regions with recurrent outbreaks and among individuals at continuing risk of exposure.

The 2014–16 West African epidemic accelerated Ebola vaccine development, leading to licensure and World Health Organization (WHO) prequalification of two vaccines. The rVSVΔG-ZEBOV-GP vaccine (Ervebo^®^, MSD), a replication-competent recombinant vesicular stomatitis virus expressing the Zaire EBOV glycoprotein, demonstrated clinical efficacy in a cluster-randomised, phase 3 ring-vaccination trial in Guinea.^1^ Complementing this approach, a heterologous two-dose regimen combining Ad26.ZEBOV and MVA-BN-Filo (Zabdeno^®^, Mvabea^®^, Janssen), administered 56 days apart, was licensed based on immunobridging, although no correlate of protection has been established in humans.^2^ While Ad26.ZEBOV, MVA-BN-Filo was approved for adults and children ≥1 year by the European Medicines Agency (EMA) in 2020, rVSVΔG-ZEBOV-GP was initially licensed only for adults by the US Food and Drug Administration (FDA) in 2019. The PREVAC trial one-year results provided pivotal pediatric safety and immunogenicity data, enabling FDA regulatory expansion of rVSVΔG-ZEBOV-GP to individuals ≥12 months in 2023.^3^ Furthermore, the Ad26.ZEBOV, MVA-BN-Filo regimen has demonstrated safety and immunogenicity even in infants younger than 1 year.^4^ In 2024, WHO’s Strategic Advisory Group of Experts on Immunization expanded Ebola vaccine recommendations to include prophylactic use in populations at ongoing risk. However, long-term durability of vaccine-induced immunity remained unknown.

During recent outbreaks, including those in the DRC, an estimated 500,000 to 1 million individuals were vaccinated worldwide.^1,5^ It is noteworthy that the risk of Ebola virus (EBOV) re-exposure remains substantial in endemic areas. EBOV can persist for years in immune-privileged sites, leading to viral relapse and renewed transmission, as documented in several outbreaks.^6,7,8,9^ Consequently, health-care workers, children, and individuals living in endemo-epidemic regions may face repeated exposure over time. These considerations highlight the importance of understanding the durability of vaccine-induced immune responses and their potential role in long-term protection.

Because phase 3 efficacy trials are not feasible outside outbreak settings, evaluation of the durability of vaccine-induced immune responses is central to informing preparedness strategies, including the potential need for booster vaccination. Although vaccine-induced antibody levels have not been formally validated as a correlate of protection in humans, they are being used as surrogate marker of clinical benefit for regulatory approval. Previous studies have shown that both licensed Ebola vaccines studied induce strong humoral responses shortly after vaccination, followed by a decline and then plateau over 12–24 months, although most recipients remain seropositive.^3,10,11,12,13,14,15^ However, comprehensive long-term data comparing vaccine platforms and age groups have been limited.

Here, we report five-year safety and immunogenicity outcomes from two large randomised, placebo-controlled trials conducted in adults and children in West Africa under a unified protocol by the Partnership for Research on Ebola Vaccinations (PREVAC) consortium. These findings extend the previously described 12-month results of the first phase of these studies.^16^

## Methods

### Trial Design and Participants

The PREVAC consortium conducted two randomized phase 2 clinical trials in adults and children, respectively, in Guinea, Liberia, Mali, and Sierra Leone. Each trial was designed as an individually-randomized superiority trial, comparing three different active Ebola vaccine strategies against placebo. Both trials were conducted using a common protocol; the design and results through 12-months of follow-up in adults and children have been published.^16,17^ The full protocol is available in the supplentary appendix.

Enrolment was community-based at four urban and two rural sites in the four West African countries. Briefly, eligible adults 18 years of age or older and children 1 to 17 years of age, all without a history of EVD, who were not pregnant or breast-feeding, were enrolled from April through December 2018. The full inclusion and exclusion criteria and procedures for staggered enrolment according to age group are provided in the supplementary methods (section S2.2).

A schematic overview depicting the randomization groups and blinding is given (section S2.3). Participants were randomly assigned at baseline to receive the Ad26.ZEBOV vaccine (0.5 ml; 5×10^10^ viral particles), followed by the MVA-BN-Filo vaccine (0.5 ml; 1×10^8^ infectious units) 56 days later (the Ad26, MVA group); the rVSVΔG-ZEBOV-GP vaccine (1.0 ml; >=7.2×10^7^ plaque-forming units), followed by placebo 56 days later (the rVSV group); the rVSVΔG-ZEBOV-GP vaccine, followed by a booster dose of the same vaccine 56 days later (the rVSV–booster group); or matching saline placebo at baseline and 56 days later. Children and adults received the same dose of vaccines. Randomization was stratified by site, and the two placebo groups were pooled for analysis.

The trials were double-blinded until all participants completed their 12-month follow-up. After Month 12, the clinical and safety data were locked (April 2020), the data were unblinded (June 2020) as pre-specified in the protocol; participants were informed of their group assignment from July 2021 onwards and long-term follow-up continued without blinding.

During the long-term follow-up, in the context of the risk of recurrent Ebola outbreaks in different African countries, the trial’s steering committee decided to offer open-label Ebola vaccination to all participants who had initially received a placebo. From December 2021 onwards, in consultation with the ministries of health of the concerned countries, participants in the placebo group were offered a single dose of the rVSVΔG-ZEBOV-GP vaccine (in Liberia and Mali) or the Ad26.ZEBOV, MVA-BN-Filo vaccine regimen (in Guinea and Sierra Leone), either at their next scheduled long-term follow-up visit or during an additional visit if their next follow-up visit was not imminent.

The trial protocol was approved by the FDA and the ethics committees of the sponsors and of the trial countries. Informed consent was obtained from all participants or, in the case of children, from a parent or guardian, and assent was also sought for children aged 7 to 17.^17^ An independent data and safety monitoring committee was in place during the blinded part of the trial. Public involvement occurred throughout the trial with the help of representatives of the local communities and social science teams.

### Long-term follow-up visits from 12 to 60 months

After the first 12 months of follow-up from the first dose of vaccine or placebo, follow-up visits occurred at Months 24 (± 6 months), 36 (± 6 months), 48 (± 6 months) and 60 (−6 months, +1 month). Antibody responses were assessed at each follow-up visit. After the initial 12-month follow-up, data on serious adverse events were collected annually via safety surveillance according to regulatory requirements and pharmacovigilance procedures in place through Month 60. In former placebo recipients who were offered active vaccination, safety data was obtained through phone calls at one and six months after vaccination at a minimum.

### Antibody Responses to Ebola Glycoprotein

For the previously published primary analysis covering Day 0 to Month 12,^16^ the Filovirus Animal Nonclinical Group (FANG) enzyme-linked immunosorbent assay (ELISA) was used to quantify antibody responses to the EBOV glycoprotein (GP) Kikwit antigen. However, during the the long-term follow-up (Month 24 to Month 60), a Luminex-based assay was used to assess antibody responses to multiple EBOV antigens, including GP Kikwit, GP Mayinga, nucleoprotein (NP), and VP40 (Supplementary section 2.5.1). This change was driven by several limitations of the FANG ELISA, such as high inter-laboratory variability, low throughput, plate inconsistencies, and complex sample processing algorithms, which questioned its suitability for large scale studies. A comparison of the performance between FANG and Luminex assay has been performed.^18^

Ebola specific antibody responses to GP Kikwit have been used as a primary immunogenicity outcome in both assays throughout the study. For Months 24-60, antibody concentrations were measured with the Luminex assay in one central laboratory (TransVIHMI, Institut de Recherche pour le Développement (IRD), Montpellier, France). In order to ensure comparability between first-year antibody measurement using the FANG ELISA and follow-up measurements using the Luminex assay, results obtained with the Luminex assay for Months 24-60 samples in Net mean fluorescence intensity (MFI) unit, were converted to ELISA Units (EU)/mL. The first-year FANG ELISA data (also expressed in EU/mL) were converted to the same range as the Luminex data based on conversion formulas developed by testing a sample subset with both assays (Supplementary Section S2.5.2).

### Statistical Analysis

Sample sizes for comparing each vaccine regimen with placebo with regard to antibody response and SAEs, separately among adults and children, exceeded those required.^16^ Data from adults and children were analyzed separately, with the use of identical statistical analyses. The analyses of immunogenicity and safety endpoints were conducted as-treated, i.e. the participants having received at least one injection of a given vaccine strategy were included in the as-treated analyses according to the vaccine strategy actually received.

After log_10_ transformation, mean antibody concentrations were compared between each vaccine group versus pooled placebo groups at each follow-up time point by analysis of covariance, with the baseline log_10_ level and site as covariates. For placebo participants who received an active vaccination during follow-up, antibody concentrations were replaced with their last available measurements prior to vaccination. Analyses were based on all available data at each visit. Geometric mean antibody concentrations at each time point, as well as geometric mean ratios (for each vaccine vs. placebo) and 95% confidence intervals are reported.

A linear mixed-effect regression model was used to investigate anti-EBOV GP1,2 Immunoglobulin G (IgG) dynamics during the long-term follow up and to determine any association with participant characteristics. The dependent variable was the log10-transformed anti-EBOV GP1,2 IgG concentrations over time (Months 24 to 60). The model included random effects on the intercept and on the slope. The fixed-effect covariates were vaccine arm, age category, sex, country, and pre-vaccination anti-EBOV GP1,2 IgG concentrations. The effect of each covariate is presented as a ratio of the geometric mean concentrations (GMCs) in a natural scale between the category of interest and its reference.

SAEs were listed and summarized by proportion. The full statistical analysis plan and details regarding analysis methods are provided in the Supplementary Material (Sections S2.6 and S2.7). Statistical analyses were performed with the use of SAS software, version 9.4 (SAS Institute) and R, version 4.1.3 (R Foundation for Statistical Computing).

## Results

### Characteristics of the Participants

A total of 1401 adults and 1401 children were enrolled in the two trials from April through December 2018 (Figures S1 and S2, Table S1). Of these, 1315 (93.9%) adults (369 in the Ad26, MVA group, 372 in the rVSV group, 179 in the rVSV–booster group, and 395 in the Placebo group) and 1322 (94.4%) children (380 in the Ad26, MVA group, 385 in the rVSV group, 190 in the rVSV–booster group, and 367 in the Placebo group) attended at least one long-term follow up visit from Month 24 to Month 60 (March 2020 to December 2023). Baseline characteristics were well balanced across randomized groups (Table 1). The as-treated population for the long-term statistical analysis is described in the Figures S3 and S4 and Table S2.

**Table 1.** Baseline characteristics of the participants participating in the long-term follow up of the PREVAC trial by intervention group.

|  | **Ad26, MVA Group** |  | **rVSV Group** |  | **rVSV–Booster Group** |  | **Placebo Group** |  |
| --- | --- | --- | --- | --- | --- | --- | --- | --- |
|  | **N** | **%** | **N** | **%** | **N** | **%** | **N** | **%** |
| **Adults** |  |  |  |  |  |  |  |  |
| **No. of participants** | 369 | 100.0 | 372 | 100.0 | 179 | 100.0 | 395 | 100.0 |
| **Age** |  |  |  |  |  |  |  |  |
| 18-29 yr | 206 | 55.8 | 197 | 53.0 | 104 | 58.1 | 224 | 56.7 |
| 30-39 yr | 65 | 17.6 | 85 | 22.9 | 39 | 21.8 | 83 | 21.0 |
| >= 40 yr | 98 | 26.6 | 90 | 24.2 | 36 | 20.1 | 88 | 22.3 |
| **Sex** |  |  |  |  |  |  |  |  |
| Male | 209 | 56.6 | 194 | 52.2 | 96 | 53.6 | 215 | 54.4 |
| Female | 160 | 43.4 | 178 | 47.9 | 83 | 46.4 | 180 | 45.6 |
| **Site** |  |  |  |  |  |  |  |  |
| Landreah | 81 | 22.0 | 79 | 21.2 | 35 | 19.6 | 76 | 19.2 |
| Maferinyah | 40 | 10.8 | 49 | 13.2 | 27 | 15.1 | 54 | 13.7 |
| Redemption | 65 | 17.6 | 67 | 18.0 | 33 | 18.4 | 76 | 19.2 |
| Mambolo | 106 | 28.7 | 99 | 26.6 | 45 | 25.1 | 105 | 26.6 |
| UCRC | 40 | 10.8 | 42 | 11.3 | 17 | 9.5 | 43 | 10.9 |
| CVD | 37 | 10.0 | 36 | 9.7 | 22 | 12.3 | 41 | 10.4 |
| **HIV Status** |  |  |  |  |  |  |  |  |
| Negative | 364 | 98.6 | 359 | 96.5 | 177 | 98.9 | 391 | 99.0 |
| Positive | 5 | 1.4 | 13 | 3.5 | 2 | 1.1 | 4 | 1.01 |
| **Children** |  |  |  |  |  |  |  |  |
| **No. of participants** | 380 | 100.0 | 385 | 100.0 | 190 | 100.0 | 367 | 100.0 |
| **Age** |  |  |  |  |  |  |  |  |
| 1-4 yr | 129 | 34.0 | 117 | 30.4 | 65 | 34.2 | 132 | 36.0 |
| 5-11 yr | 120 | 31.6 | 142 | 36.9 | 63 | 33.2 | 121 | 33.0 |
| 12-17 yr | 131 | 34.5 | 126 | 32.7 | 62 | 32.6 | 114 | 31.1 |
| **Sex** |  |  |  |  |  |  |  |  |
| Male | 203 | 53.4 | 214 | 55.6 | 111 | 58.4 | 198 | 54.0 |
| Female | 177 | 46.6 | 171 | 44.4 | 79 | 41.6 | 169 | 46.1 |
| **Site** |  |  |  |  |  |  |  |  |
| Landreah | 72 | 19.0 | 72 | 18.7 | 41 | 21.6 | 75 | 20.4 |
| Maferinyah | 79 | 20.8 | 71 | 18.4 | 32 | 16.8 | 67 | 18.3 |
| Redemption | 62 | 16.3 | 61 | 15.8 | 31 | 16.3 | 51 | 13.9 |
| Mambolo | 76 | 20.0 | 91 | 23.6 | 44 | 23.2 | 88 | 24.0 |
| UCRC | 40 | 10.5 | 38 | 9.9 | 19 | 10.0 | 36 | 9.8 |
| CVD | 51 | 13.4 | 52 | 13.5 | 23 | 12.1 | 50 | 13.6 |
| **HIV Status** |  |  |  |  |  |  |  |  |
| Negative | 380 | 100.0 | 385 | 100.0 | 190 | 100.0 | 367 | 100.0 |
| Positive | .. | .. | .. | .. | .. | .. | .. | .. |
| 480 |  |  |  |  |  |  |  |  |
| 481 |  |  |  |  |  |  |  |  |

### Follow up

Participant retention in the trials from Month 24 to Month 60 was high. Among the initially randomized population, 91.7% of adults and 92.5% of children returned for follow-up visits at Month 24, 90.3% and 90.8% at Month 36, 89.2% and 88.2% at Month 48, and 82.9% and 83.2% at Month 60, respectively (Tables S3, S4 and S5). From Month 36 onwards, between December 2021 and December 2022, 338 adults (82%) and 317 children (82%) in the initial placebo group elected to receive active Ebola vaccination (Table S6). For these participants, antibody concentration after vaccination was censored and imputed with their last available data before vaccination for the long-term analysis of the placebo group (antibody concentrations prior to without censoring are shown in Table S7).

### Antibody Responses

The dynamics of the antibody concentrations between Months 0 and 60 are shown in Figure 1. Geometric mean antibody levels declined between Month 12 and Month 24, followed by a plateau from Month 24 to Month 60 with all three vaccine strategies (Tables S8 to S9 and Figures S5 and S6). Among adults, GMC plateaus ranged from 253 to 224 EU/mL in the Ad26, MVA group, 1160 to 956 EU/mL in the rVSV group, 1243 to 1051 EU/mL in the rVSV–booster group, and 69 to 66 EU/mL in the placebo group during long-term follow up (P<0.001 for all comparisons of each vaccine with placebo) (Table 2). Among children, the GMC plateaus ranged from 672 to 629 EU/mL in the Ad26, MVA group, 2142 to 1695 EU/mL in the rVSV group, 2637 to 2182 EU/mL in the rVSV–booster group, and 72 to 67 EU/mL in the placebo group during long-term follow up (P<0.001 for all comparisons of each vaccine with placebo, at all time points).

**Figure 1.**
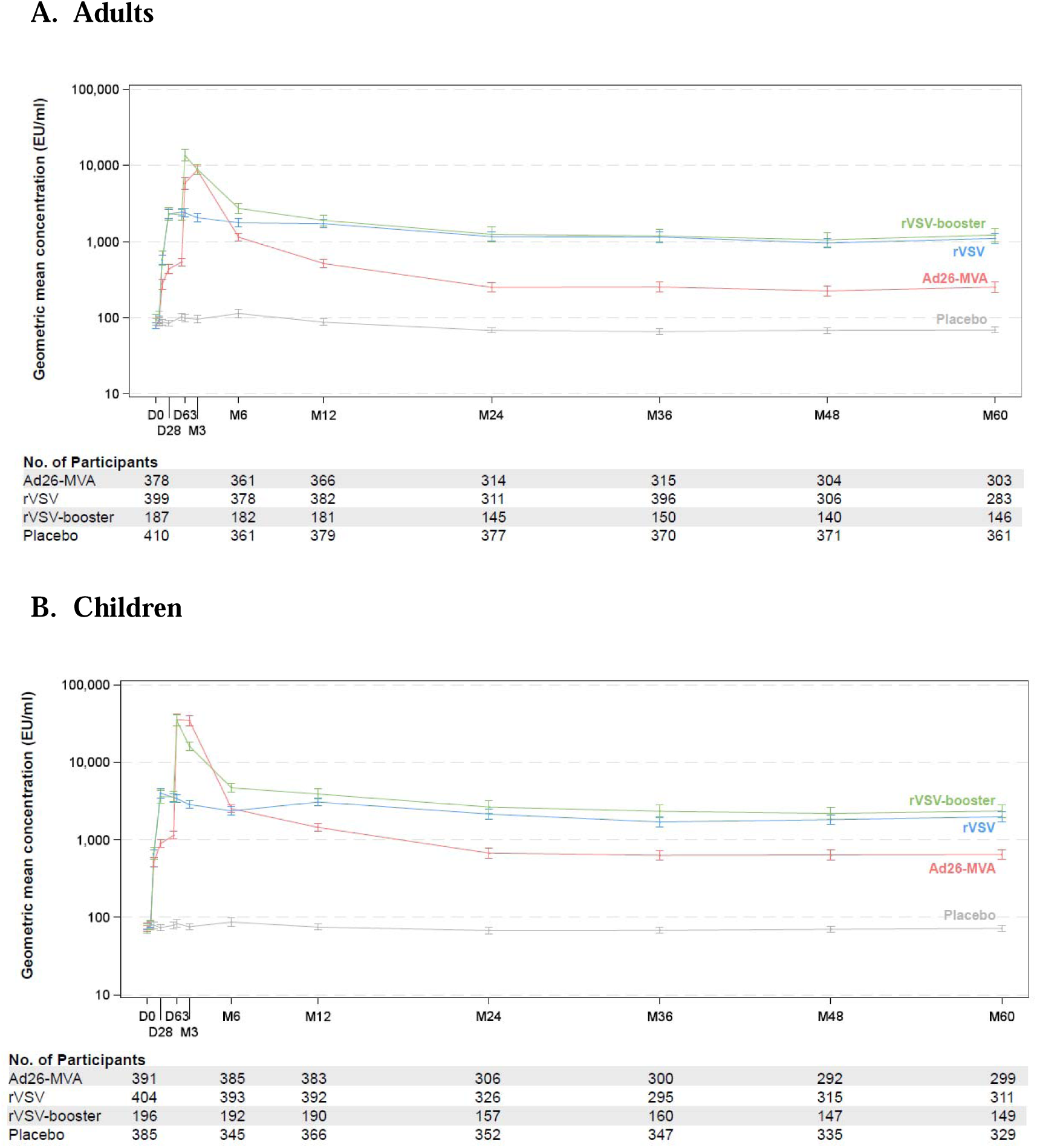
Antibody response in adults and children (Geometric Mean Concentrations), according to trial visit. The geometric mean concentration was based on a log10 concentration. l7 bars indicate 95% confidence intervals. D0 to M12 data were measured by FANG ELISA and converted to the range of Luminex. M24 to M60 data were measured by Luminex.

**Table 2.**
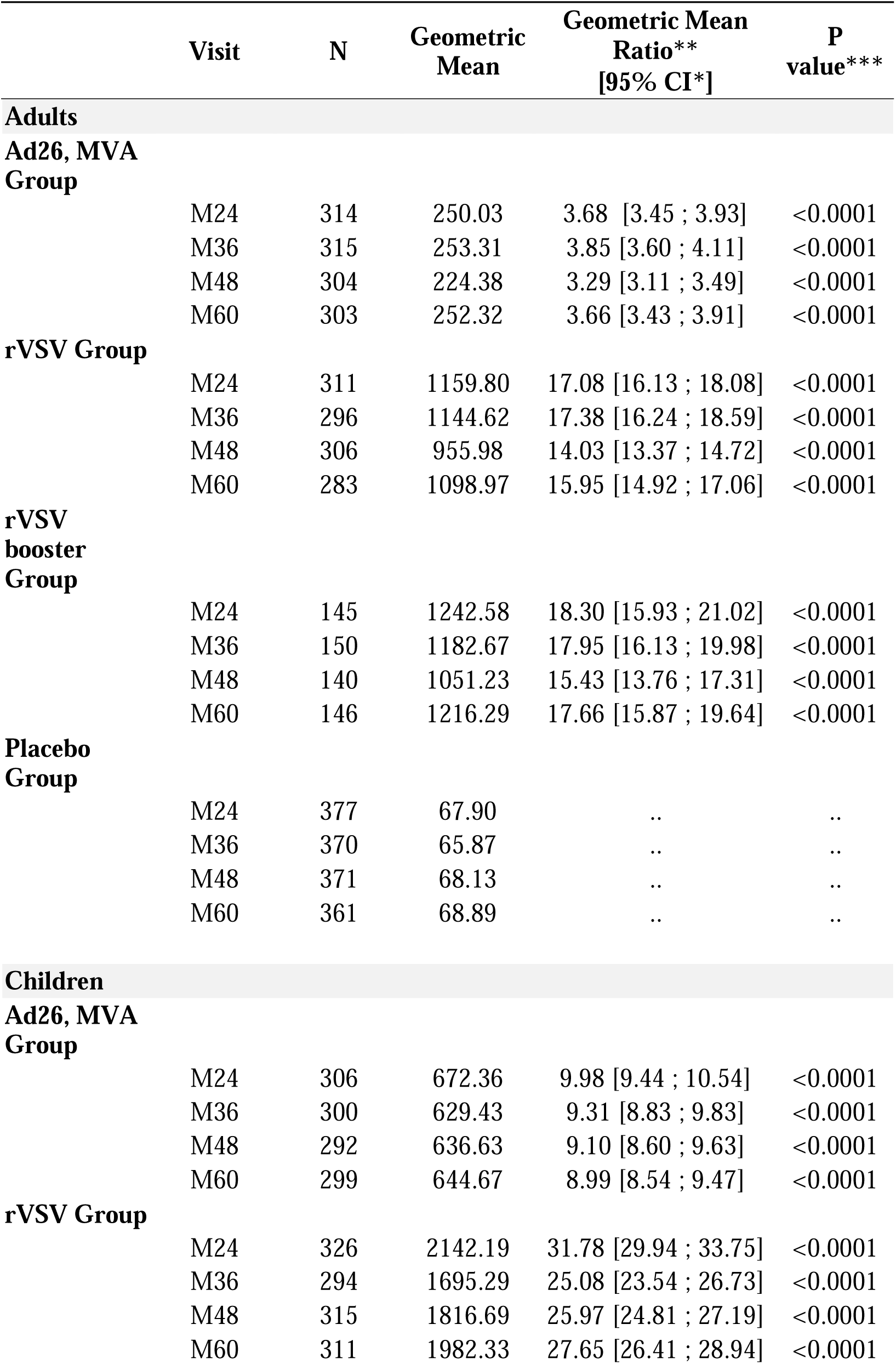
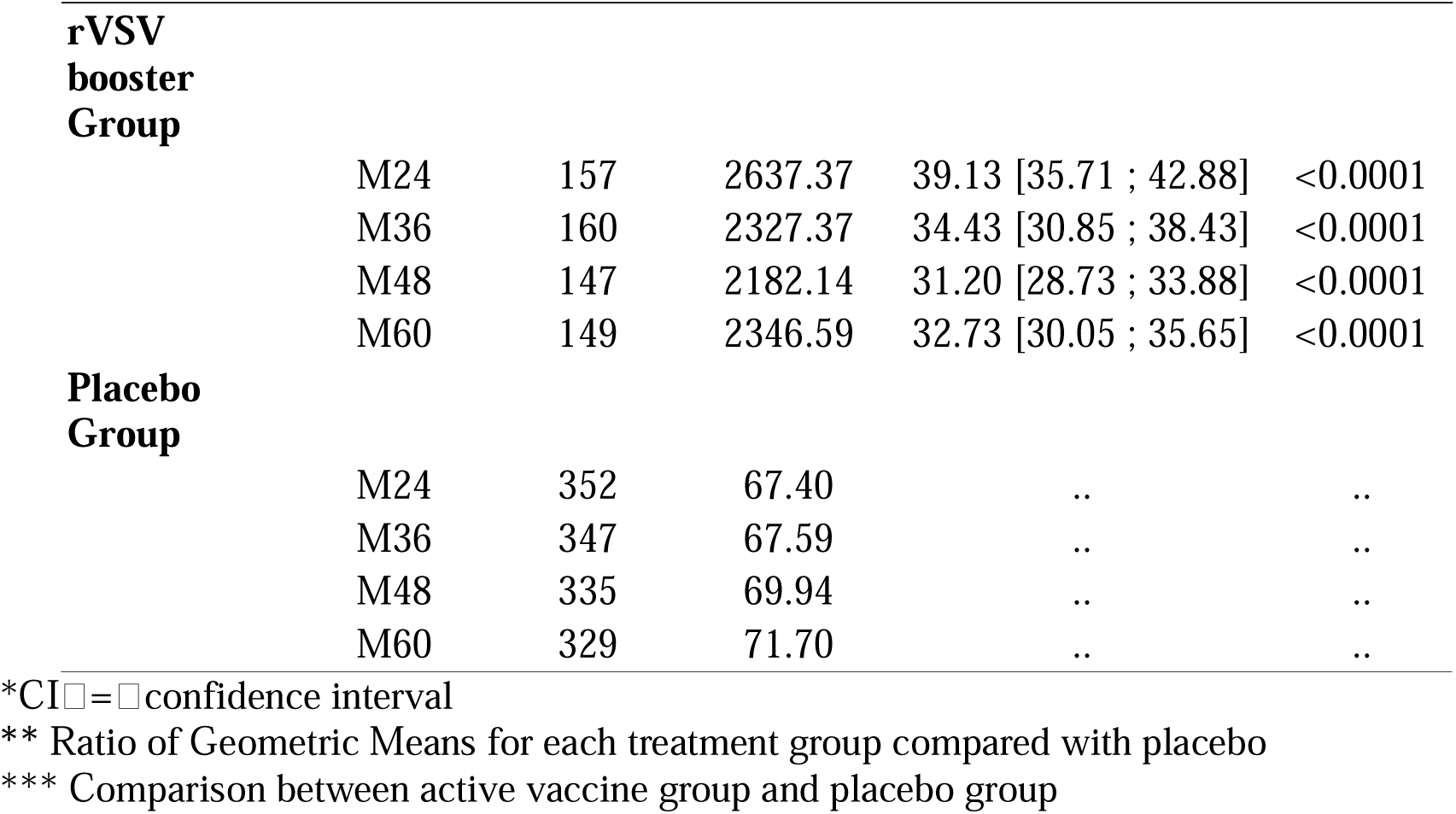
Antibody concentration (geometric mean) per active vaccine group according to trial visits and comparison to the placebo group.

|  | Visit | N | Geometric Mean | Geometric Mean Ratio**<br>[95% CI*] | P value*** |
| --- | --- | --- | --- | --- | --- |
| **Adults** |  |  |  |  |  |
| **Ad26, MVA Group** |  |  |  |  |  |
|  | M24 | 314 | 250.03 | 3.68 [3.45 ; 3.93] | <0.0001 |
|  | M36 | 315 | 253.31 | 3.85 [3.60 ; 4.11] | <0.0001 |
|  | M48 | 304 | 224.38 | 3.29 [3.11 ; 3.49] | <0.0001 |
|  | M60 | 303 | 252.32 | 3.66 [3.43 ; 3.91] | <0.0001 |
| **rVSV Group** |  |  |  |  |  |
|  | M24 | 311 | 1159.80 | 17.08 [16.13 ; 18.08] | <0.0001 |
|  | M36 | 296 | 1144.62 | 17.38 [16.24 ; 18.59] | <0.0001 |
|  | M48 | 306 | 955.98 | 14.03 [13.37 ; 14.72] | <0.0001 |
|  | M60 | 283 | 1098.97 | 15.95 [14.92 ; 17.06] | <0.0001 |
| **rVSV booster Group** |  |  |  |  |  |
|  | M24 | 145 | 1242.58 | 18.30 [15.93 ; 21.02] | <0.0001 |
|  | M36 | 150 | 1182.67 | 17.95 [16.13 ; 19.98] | <0.0001 |
|  | M48 | 140 | 1051.23 | 15.43 [13.76 ; 17.31] | <0.0001 |
|  | M60 | 146 | 1216.29 | 17.66 [15.87 ; 19.64] | <0.0001 |
| **Placebo Group** |  |  |  |  |  |
|  | M24 | 377 | 67.90 | .. | .. |
|  | M36 | 370 | 65.87 | .. | .. |
|  | M48 | 371 | 68.13 | .. | .. |
|  | M60 | 361 | 68.89 | .. | .. |
| **Children** |  |  |  |  |  |
| **Ad26, MVA Group** |  |  |  |  |  |
|  | M24 | 306 | 672.36 | 9.98 [9.44 ; 10.54] | <0.0001 |
|  | M36 | 300 | 629.43 | 9.31 [8.83 ; 9.83] | <0.0001 |
|  | M48 | 292 | 636.63 | 9.10 [8.60 ; 9.63] | <0.0001 |
|  | M60 | 299 | 644.67 | 8.99 [8.54 ; 9.47] | <0.0001 |
| **rVSV Group** |  |  |  |  |  |
|  | M24 | 326 | 2142.19 | 31.78 [29.94 ; 33.75] | <0.0001 |
|  | M36 | 294 | 1695.29 | 25.08 [23.54 ; 26.73] | <0.0001 |
|  | M48 | 315 | 1816.69 | 25.97 [24.81 ; 27.19] | <0.0001 |
|  | M60 | 311 | 1982.33 | 27.65 [26.41 ; 28.94] | <0.0001 |
| **rVSV booster Group** |  |  |  |  |  |
|  | M24 | 157 | 2637.37 | 39.13 [35.71 ; 42.88] | <0.0001 |
|  | M36 | 160 | 2327.37 | 34.43 [30.85 ; 38.43] | <0.0001 |
|  | M48 | 147 | 2182.14 | 31.20 [28.73 ; 33.88] | <0.0001 |
|  | M60 | 149 | 2346.59 | 32.73 [30.05 ; 35.65] | <0.0001 |
| **Placebo Group** |  |  |  |  |  |
|  | M24 | 352 | 67.40 | .. | .. |
|  | M36 | 347 | 67.59 | .. | .. |
|  | M48 | 335 | 69.94 | .. | .. |
|  | M60 | 329 | 71.70 | .. | .. |

\*CI = confidence interval
\*\* Ratio of Geometric Means for each treatment group compared with placebo
\*\*\* Comparison between active vaccine group and placebo group

In each of the vaccine groups, the GMCs were greater among children than among adults. To facilitate consideration in the context of previous studies, we described the proportions of antibody responders for different fold change categories (Figures S7 and S8, Tables S12 and Table S13). The descriptive analyses indicate considerable inter-individual variability in antibody concentrations (large interquartile ranges in Figures S5 and S6, and varying fold changes in Figures S7 and S8).

### Baseline determinants of long-term antibody levels

Description of the dynamics of the mean antibody concentrations according to age group, sex, country and antibody level at baseline are shown in Figures S9 to S20. In the linear mixed effect regression model, the slope of mean antibody concentrations from Month 24 to Month 60 varied according to age, country, and baseline IgG concentration across all three vaccine strategies, and according to sex in the rVSV and rVSV-booster groups (Figures S21-S24, Table S14). At the end of the study (Month 60), children had higher GMCs than adults across all vaccine groups, with GMCs ranging from 1.6- to 3.6-fold higher depending on the strategy and age group. Women had 1.2 and 1.36-fold higher GMCs than men in the rVSV and rVSV-booster groups, respectively. Compared to participants from Mali, GMCs of participants from Guinea, Sierra Leone and Liberia were 0.27 to 0.48-fold lower at Month 60 across the three vaccine strategies (Table 3).

**Table 3.** Baseline variables associated with M60 anti-EBOV GP_1,2_ antibody concentrations after Ad26.ZEBOV, MVA-BN-Filo, rVSVΔG-ZEBOV-GP and rVSVΔG-ZEBOV-GP booster vaccination.

|  | Anti-EBOV GP<sub>1,2</sub> antibody End of study<sup>a</sup> Ratios of geometric mean concentration [95% CI] |  |  |
| --- | --- | --- | --- |
|  | Ad26, MVA Group | rVSV Group | rVSV-booster Group |
| **Age category** |  |  |  |
| **Adults** | Reference | Reference | Reference |
| **Children 1-4 yr** | 3.57 [2.79 ; 4.49] | 1.61 [1.19 ; 2.14] | 1.97 [1.32 ; 2.75] |
| **Children 5-11 yr** | 2.22 [1.63 ; 2.91] | 1.75 [1.34 ; 2.25] | 2.04 [1.48 ; 2.78] |
| **Children 12-17 yr** | 2.50 [1.87 ; 3.31] | 2.18 [1.69 ; 2.77] | 2.00 [1.43 ; 2.70] |
| **Sex** |  |  |  |
| **Male** | Reference | Reference | Reference |
| **Female** | 1.09 [0.88 ; 1.34] | 1.2 [0.97 ; 1.45] | 1.36 [1.06 ; 1.74] |
| **Country** |  |  |  |
| **Mali** | Reference | Reference | Reference |
| **Guinea** | 0.31 [0.24 ; 0.41] | 0.39 [0.30 ; 0.51] | 0.44 [0.31 ; 0.62] |
| **Liberia** | 0.32 [0.23 ; 0.45] | 0.40 [0.29 ; 0.54] | 0.48 [0.31 ; 0.73] |
| **Sierra Leone** | 0.27 [0.20 ; 0.36] | 0.39 [0.30 ; 0.51] | 0.46 [0.30 ; 0.67] |
| **Baseline antibody concentration** |  |  |  |
| **<200 EU/mL** | Reference | Reference | Reference |
| **>=200 EU/mL** | 1.38 [0.94 ; 1.95] | 0.96 [0.73 ; 1.24] | 0.99 [0.65 ; 1.45] |

Estimations from mixed-effect linear regression model using antibody concentrations from M24 to M60. GMC ratios and 95% confidence intervals at the end of the study were estimated by bootstrap.
<sup>a</sup>The end of the study was defined as 60 months after receipt of the prime vaccination, respectively. EBOV = Ebola virus. GP<sub>1,2</sub>: glycoprotein. yr = years. EU/mL = enzyme-linked immunosorbent assay units per milliliter. CI = confidence interval.

### Serious Adverse Events and Deaths during long-term follow-up

During the long-term follow-up, SAEs were reported in 15 of 381 adults (3.9%) in the Ad26, MVA group, in 22 of 405 (5.4%) in the rVSV group, in 10 of 187 (5.4%) in the rVSV–booster group, and in 17 of 412 (4.1%) in the pooled placebo group. Among those originally in the pooled placebo group, SAEs occurred, regardless of the timing of the vaccination, in 13 of 204 (6.4%) participants later vaccinated with Ad26, MVA and in 1 of 134 (0.8%) participants later vaccinated with rVSV and in 3 of 74 (4.1%) who elected not to receive active vaccine (Tables S15 and S17). The majority of those events occurred prior to vaccination (Table S16). One post-vaccination SAE was considered as possibly related to vaccination. This was in a participant aged in their 30s from the former placebo group who experienced an ischaemic stroke 49 days after administration of the Ad26-based vaccine.

SAEs during long-term follow-up were reported in 8 of 396 children (2.0%) in the Ad26, MVA group, in 8 of 410 (2.0%) in the rVSV group, in 5 of 198 (2.5%) in the rVSV–booster group, and in 8 of 388 (2.1%) in the pooled placebo group. Among those originally in the pooled placebo group, SAEs occurred, regardless of the timing of the vaccination, in 4 of 199 (2.0%) participants later vaccinated with Ad26, MVA, in none of 118 (0%) participants later vaccinated by rVSV, and in 4 of 71 (5.6%) who elected not to receive vaccine (Tables S15 and S18). The majority of those events occurred prior to vaccination and none was considere related to vaccination (Table S16).

Overall, 17 adults (6 in the Ad26, MVA group, 6 in the rVSV group, 3 in the rVSV–booster Group, and 2 in the pooled placebo group) and 5 children (2 in the Ad26, MVA group, 2 in the rVSV group, and 1 in the pooled placebo group) died during the long-term follow-up (Tables S19-S20). None of the deaths were judged by the site investigator to be related to administration of Ebola vaccine.

## Discussion

This long-term 5-year follow-up of two large, parallel randomized studies conducted in children and adults in West Africa provides evidence on the long-term safety and humoral immunogenicity of three vaccine strategies using the two licensed anti-EVD vaccines, Ad26.ZEBOV, MVA-BN-Filo and rVSVΔG-ZEBOV-GP. A key strength of these studies is the exceptionally high participant retention over five years. Among the 2,802 volunteers enrolled in the initial phase of the trials,^19^ approximately 90% remained in follow-up until year 3, and 83% completed the full five-year follow-up. This high retention rate strengthens confidence in the validity of the findings and demonstrates the feasibility of sustained long-term follow-up within a multipartner consortium in resource limited settings. It also reflects the effectiveness of the community engagement strategies implemented throughout the study and the strong involvement of geographically diverse adult and pediatric populations across West Africa, which enhances the generalizability of the results.

Our results extend existing knowledge on the long-term safety and immunogenicity of Ad26.ZEBOV, MVA-BN-Filo and rVSVΔG-ZEBOV-GP vaccines. They also provide valuable data to inform vaccination strategies, particularly in paediatric populations for which long term immunogenicity data were not previously available.^19^ These data also confirmed the predictions provided by mathematical models of vaccine responses.^20^ Given the high risk of severe disease and mortality from EVD in young children under five years of age, consideration may be given to vaccinating children living in the households of adult survivors.^5^

Both vaccines were well tolerated over the long-term in children and adults. Longitudinal analyses of humoral immunity showed a modest decline in anti-GP antibody mean concentrations between Months 12 and 24 following vaccination with Ad26.ZEBOV, MVA-BN-Filo or one or two doses of rVSVΔG-ZEBOV-GP, consistent with previous reports for both vaccines.^14,15,21^ Notably, extended follow-up demonstrated a stable mean plateau of antibody concentrations from Month 24 to Month 60 across all vaccine strategies and age groups.

Several differences in immune responses were observed between vaccines and populations. The mean magnitude of humoral responses was consistently higher in children than in adults, irrespective of age subgroup or vaccine regimen. At Month 60, mean antibody concentrations remained increased by more than 16-fold relative to baseline in 35% of children vaccinated with Ad26.ZEBOV, MVA-BN-Filo and in 68-72% of children who received rVSVΔG-ZEBOV-GP, compared with 15% and 47-48% of adults, respectively. These findings are consistent with previous studies reporting stronger vaccine-induced immune responses in children and adolescents than in adults for other vaccines.^22,23^

We also observed higher mean antibody concentrations in vaccinated females than in males in both rVSVΔG-ZEBOV-GP arms. This sex-based difference aligns with findings from studies of Ebola, SARS-CoV-2, and influenza vaccines, which have reported slower antibody waning or higher peak responses in females.^24,25^

Long-term follow-up across participating countries revealed higher mean antibody responses in Mali, particularly from Month 48 onward. Although the underlying causes of these differences remain unclear, this finding with Ebola vaccines accords with previously reported heterogeneity in immune responses to different investigational and licensed vaccines across African settings.^4,26,27^ Environmental exposures, host genetics, gut microbiome composition, and the prevalence of concurrent infections have been proposed as contributors to geographic variability in vaccine immunogenicity.^23,28^

A key remaining question is whether post-vaccination antibody concentrations, even when sustained over the long term, are sufficient to confer protection to individuals at risk of exposure during future Ebola outbreaks. Although all vaccine strategies induced mean durable antibody responses up to 60 months post vaccination, the proportion of high responders (defined as a ≥16-fold increase from baseline) differed by vaccine platform, ranging from approximately 15% among Ad26, MVA adult recipients to up to 70% among rVSV children recipients. Descriptive analyses indicated considerable inter-individual variability in antibody concentrations. Associations between levels of anti-GP antibody concentrations and protection have been shown in non-human primate models and thus antibody concentrations are considered a likely correlate of protection in humans,^2^ but whether this is the titer conferring protection still remains uncertain. This uncertainty reflects the absence of a validated immunological correlate of protection or a minimal threshold of protection for anti-EBOV GP IgG antibodies.^29^

The persistence of antibody responses suggests the induction of long-lived memory B-cell responses and raises questions regarding the potential benefit and optimal timing of booster immunisation.^30^ Robust anamnestic humoral responses have been reported following homologous booster doses administered more than one year after primary vaccination with either rVSVΔG-ZEBOV-GP or Ad26.ZEBOV–MVA-BN-Filo regimens.^31,32,21,33^ Preclinical studies have also shown that protection against EBOV infection is mediated by both humoral and cellular immune mechanisms.^34^ Consistent with these findings, we previously demonstrated in a subgroup of Guinean participants enrolled in PREVAC that both vaccine platforms induced durable EBOV-specific T-cell memory responses persisting for up to 60 months without evidence of waning.^35^ It is of note, that while the additional dose of rVSVΔG-ZEBOV-GP at day 56 did not lead to any long-term changes in antibody concentrations compared to a single dose at baseline, in a separate study in a US population, a booster dose at 18 months led to approximately a 7-fold increase concentration at 36 months when compared to no booster dose.^31^ Given that the level of antibody required to protect against infection or disease is not known, further work is needed to define the optimal strategy for prophylaxis.

A challenge in this study was the change of the primary endpoint antibody assay during long-term follow-up. Substantial efforts were made to standardise the assessment of vaccine-induced humoral responses by using a robust Luminex-based assay, thereby overcoming the known limitations of the FANG assay.^36^ Anti-GP binding antibody responses from Months 24 to 60 were measured in a central laboratory further reducing inter-assay and inter-site variability. This methodological approach supports the conclusion that the observed differences in humoral responses between vaccine groups, age strata, and countries are unlikely to be attributable to assay-related variability.

In addition to the high participant retention rate and the use of a centralized laboratory for Luminex assay, the strengths of this randomized study include the long duration of follow-up. This study would not have been possible without the strong partnership between the investigators and governments of the four West African host countries and the support of European, UK, and US sponsors.

The study had some limitations. Although anti-EBOV GP antibody concentrations were sustained through five years after vaccination, it is not clear whether or not these are adequate levels to provide protection. In addition, neutralising antibody activity was not assessed during the long-term follow-up. Also biological variability related to host or environmental factors across countries that were mot measured cannot be fully excluded.

The demonstration of sustained humoral immune responses over five years provides a biological rationale for evaluation of strategies aimed at boosting immune memory following primary vaccination. Although data on the safety and immunogenicity of heterologous booster regimens for Ebola vaccines remain limited, these findings support the need for further evaluation of an added value of booster strategies, potentially as stratified approaches dependent on demographic characteristics, as part of long-term outbreak preparedness in populations at continued risk of EBOV exposure.

In conclusion, our findings demonstrate durable immune responses induced by licensed Ebola vaccines over five years and help to inform outbreak preparedness and response. The persistence of humoral and cellular immune responses suggests the presence of long-term immunologic memory. However, considerable inter-individual variability was observed with some individuals exhibiting very low antibody levels after five years. These data support the need for even longer term follow up after initial vaccination to assess the benefits of a booster in some individuals. Further work is also warranted to define the correlates of protection and development of strategies to maintain immunity at that protective level for populations at continued risk of EBOV exposure.

## Supporting information

PREVAC protocol

## Data Availability

PREVAC pseudonymized data are sensitive data collected through a clinical trial. Therefore, for participants data protection and privacy, legal and ethical reasons, the PREVAC pseudonymized data cannot be shared publicly. The PREVAC metadata are available on the Recherche Data Gouv repository, DOI: 10.57745/PVXRMC (https://doi.org/10.57745/PVXRMC).
Authorization for the use of the PREVAC/PREVAC-UP pseudonymized clinical and immunological data must be granted by the sponsors and the concerned country PIs. The final decision to either authorize or not the use of the data and data transfer lies with the sponsors in order to ensure the data request is in accordance with legal and ethical regulations. If a data request is granted, a DTA (data transfer agreement) must be established.
For any additional question regarding access to the PREVAC/PREVAC UP pseudonymized data please contact.

https://doi.org/10.57745/PVXRMC

## Notes

The members of the writing committee assume responsibility for the overall content and integrity of this article.

## Trial registration numbers

ClinicalTrials.gov number, NCT02876328 https://clinicaltrials.gov/study/NCT02876328 EudraCT number 2017-001798-18 https://www.clinicaltrialsregister.eu/ctr-search/trial/2017-001798-18/3rd Pan African Clinical Trials Registry number, PACTR201712002760250.

## Open Science

PREVAC pseudonymized data are sensitive data collected through a clinical trial. Therefore, for participants data protection and privacy, legal and ethical reasons, the PREVAC pseudonymized data cannot be shared publicly.

The PREVAC metadata are available on the Recherche Data Gouv repository, DOI: 10.57745/PVXRMC (https://doi.org/10.57745/PVXRMC).

Authorization for the use of the PREVAC/PREVAC-UP pseudonymized clinical and immunological data must be granted by the sponsors and the concerned country PIs. The final decision to either authorize or not the use of the data and data transfer lies with the sponsors in order to ensure the data request is in accordance with legal and ethical regulations. If a data request is granted, a DTA (data transfer agreement) must be established.

For any additional question regarding access to the PREVAC/PREVAC UP pseudonymized data please contact.

## Funding

This research was supported in part by the US National Institutes of Health (NIH), by the French Institut national de la santé et de la recherche médicale (Inserm) and by the London School of Hygiene and Tropical Medicine (LSHTM).

The clinical trial was conducted with the support of Janssen, Bavarian Nordic and Merck & Co., Inc., Rahway, NJ, USA who provided the vaccines according to the EBOVAC 1 grant agreement.

This project is part of the EDCTP2 programme supported by the European Union (grant number RIA2017S-2014 – PREVAC-UP).

This project has received funding from the Innovative Medicines Initiative 2 Joint Undertaking (IMI2JU) under grant agreement No 115854, EBOVAC1. This Joint Undertaking receives support from the European Union’s Horizon 2020 research and innovation program and EFPIA.

Funding provided in part by NCI contract HHSN261201500003I and 75N91019D00024 through the Frederick National Laboratory for Cancer Research. The content of this publication does not necessarily reflect the views or policies of the Department of Health and Human Services, nor does mention trade names, commercial products, or organizations imply endorsement by the U.S. Government.

The project has been funded by a dedicated Inserm allocation on behalf of the French Research Ministry.

The dissemination represents only the authors’ views and IMI2JU is not responsible for any use of the information contained in the dissemination.

## Acknowledgements

We are grateful to the Ministries of Health of Guinea, Liberia, Sierra Leone and Mali who permitted the conduct of the trial. We furthermore thank Alima, GOAL, and all site collaborators for their contribution in the implementation of the trial. The authors and study team wish to thank the participants who consented to the trial.

## Notes

### Competing Interest Statement

The authors have declared no competing interest.

### Clinical Trial

ClinicalTrials.gov number, NCT02876328; EudraCT number 2017-001798-18; and Pan African Clinical Trials Registry number, PACTR201712002760250

### Author Declarations

The study protocol, the informed consent and assent forms, including participants information materials, were approved by ethics committees of the sponsors (INSERM IRB 00003888, LSHTM) and the implementing countries (Guinea, Liberia, Mali, and Sierra Leone) before each version of the protocol was implemented. NIH established an institutional reliance agreement with INSERM to rely upon the INSERM ethics committee.

