## Supplementary material for "Five-year immunogenicity and safety follow-up of the PREVAC randomized Trial of Vaccines for Zaire Ebola Virus Disease": PREVAC protocol

DEPARTMENT OF HEALTH & HUMAN SERVICES
National Institute of Allergy and Infectious Diseases
National Institutes of Health
Bethesda, MD

Inserm
Institut national
de la santé et de la recherche médicale

LONDON SCHOOL of HYGIENE &TROPICAL MEDICINE

### Partnership for Research on Ebola VACcination (PREVAC)

| | |
|---|---|
| **Version:** | 7.0 |
| **Date:** | 13 August 2021 |
| **Sponsors:** | Office of Clinical Research Operations and Regulatory Compliance (OCRPRO) Division of Clinical Research (DCR) National Institute of Allergy and Infectious Diseases (NIAID) National Institutes of Health (NIH)<br><br>Institut National de la Santé et de la Recherche Médicale (INSERM)<br><br>London School of Hygiene & Tropical Medicine (LSHTM) |
| **Executive Committee:** | H. Clifford Lane (NIH), Yves Lévy (INSERM), Peter Piot (LSHTM) |
| **Coordinating Investigator:** | Yazdan Yazdanpanah MD, PhD |
| **Site Investigators:** | See Appendix C |
| **Ethics Review Boards:** | See Appendix C |
| **Protocol Number:** | C15-33 (INSERM), PREVACEBL3005 (LSHTM) |
| **Study Vaccines Provided by:** | Merck & Co., Inc Whitehouse Station, New Jersey, United States<br><br>Janssen Vaccines and Prevention BV Leiden, The Netherlands |

#### LIST OF FIGURES

Figure 1: PREVAC Study Design (Version 4.0) ............................................. 41

### LIST OF ABBREVIATIONS

| | |
|---|---|
| Ad26 | Adenovirus serotype 26 |
| Ad26.ENVA.01 | Adenovirus serotype 26 vector expressing the human immunodeficiency virus type 1, Clade A envelope protein (vaccine) |
| Ad26.ZEBOV | Adenovirus serotype 26 vector expressing the glycoprotein of the Ebola virus Mayinga variant (vaccine) |
| ACHIV | African-Canadian study of HIV-Infected Adults |
| AE | Adverse event |
| AESI | Adverse event of Special Interest |
| AR | Adverse reaction |
| ALT | Alanine transaminase |
| AST | Aspartate transaminase |
| BDBV | Bundibugyo ebolavirus |
| BN | Bavarian Nordic |
| BSL | BioSafety Level |
| CBC | Complete blood count |
| CD | Cluster of differentiation |
| CFR | Code of Federal Regulations |
| ChAd3-EBO Z | Replication deficient Chimpanzee adenovirus type 3-derived vector encoding the Ebola virus Zaire glycoprotein (vaccine) |
| CI | Confidence interval |
| COVID19 | Coronavirus Disease of 2019 |
| CRF | Case report form |
| CVST | Cerebral Venous Sinus Thrombosis |
| DCR | Division of Clinical Research |
| DRC | Democratic Republic of the Congo |
| DSMB | Data and Safety Monitoring Board |
| EBOV | Ebola virus |
| EC | Ethics Committee |
| eCRF | Electronic case report form |
| ELISA | Enzyme-linked immunosorbent assay |
| ELISpot | Enzyme-linked immunospot |
| EMA | European Medicines Authority |
| EU | European Union |
| EU/mL | ELISA Units per milliliter |
| EVD | Ebola virus disease |
| FANG | Filovirus Animal Nonclinical Group |
| FDA | Food and Drug Administration |
| GCP | Good Clinical Practice |
| GP | Glycoprotein |
| GSK | GlaxoSmithKline plc (Brentford, UK) |
| HIC | High-Income Country |

| | |
|---|---|
| HIV | Human Immunodeficiency Virus |
| ICS | Intracellular cytokine staining |
| IFNγ | Interferon gamma |
| Ig | Immunoglobulin |
| IM | Intramuscular |
| IND | Investigational New Drug |
| INRB | Institut National pour la Recherche Biomédicale |
| INSERM | French Institute of Health and Medical Research |
| IRB | Institutional Review Board |
| J&J | Johnson & Johnson |
| LD50 | Median lethal dose |
| LLN | Lower Limit Normal |
| LMIC | Lower and/or Middle-Income Country |
| LSHTM | London School of Hygiene & Tropical Medicine |
| LTFU | Loss-to-follow-up |
| MARV | Marburg virus |
| MCV | Mean corpuscular volume |
| MVA | Modified Vaccinia Ankara |
| MVA-BN-Filo | Modified Vaccinia Ankara Bavarian Nordic vector expressing multiple filovirus proteins (vaccine) |
| NHP | Non-human primate |
| NIAID | National Institute of Allergy and Infectious Diseases |
| NIH | National Institutes of Health |
| NK | Natural killer |
| NP | Nucleoprotein |
| OCRPRO | Office of Clinical Research Policy and Regulatory Operations |
| PBMC | Peripheral blood mononuclear cell |
| PCR | Polymerase chain reaction |
| pfu | Plaque-forming units |
| PHAC | Public Health Agency of Canada |
| PID | Participant Identification Code |
| PREPARE | Pre-Exposure Prophylaxis in People at Potential Occupation Risk for Ebola Virus Exposure |
| PREVAC | Partnership for Research on Ebola VACcination |
| PREVAIL | Partnership for Research on Ebola Virus in Liberia |
| RESTV | Reston ebolavirus |
| RDW | Red cell distribution |
| rHAd26/MVA | Ad26.ZEBOV prime/MVA-BN-Filo boost (vaccine) |
| RNA | Ribonucleic acid |
| RT-PCR | Reverse transcriptase polymerase chain reaction |
| rVSV | Recombinant vesicular stomatitis virus |
| rVSVΔG-ZEBOV-GP | Recombinant vesicular stomatitis virus-Zaire Ebola virus envelope glycoprotein (vaccine) |
| SAE | Serious adverse event |
| SAR | Serious adverse reaction |

| | |
|---|---|
| SD | Standard deviation |
| SFU | Spot forming unit |
| SID | Syringe Identification Number |
| SOP | Standard Operating Procedure |
| STRIVE | Sierra Leone Trial to Introduce a Vaccine Against Ebola |
| SUSAR | Suspected Unexpected Serious Adverse Reaction |
| SUDV | Sudan ebolavirus |
| TAFV | Taï Forest ebolavirus |
| TCID50 | 50% tissue culture infective dose |
| TTS | Thrombosis with Thrombocytopenia Syndrome |
| UK | United Kingdom |
| UP | Unanticipated Problem |
| US | United States |
| vp | Viral particles |
| VSV | Vesicular stomatitis virus |
| WHO | World Health Organization |
| ZEBOV | Zaire ebolavirus |

#### PROTOCOL SYNOPSIS

**Full Title:** Partnership for Research on Ebola VACcinations

**Short Title:** PREVAC

**Clinical Phase:** 2

**Investigational New Drug (IND) Sponsor:** NIAID/OCRPRO

**Sample Size:** N= 2,800 with targets of 1,400 adults and 1,400 children were planned to be enrolled under Version 4.0 which compares the Ad26.ZEBOV (adenovirus serotype 26 [Ad26] vector expressing the glycoprotein of the Ebola virus Mayinga)/MVA-BN-Filo (modified Vaccinia Ankara [MVA] Bavarian Nordic [BN] vector expressing multiple filovirus proteins) vaccine with placebo and the undiluted dose of the rVSVΔG-ZEBOV-GP (recombinant vesicular stomatitis virus [rVSV]-Zaire Ebola virus [ZEBOV] envelope glycoprotein [GP]) vaccine with placebo. No participants were enrolled under Version 1.0; under Version 2.0, 537 participants ≥ 12 years of age were randomized for the comparison of the Ad26.ZEBOV prime/MVA-BN-Filo boost vaccine (rHAd26/MVA) with placebo; and under Version 3.0 1,450 participants were randomized for the comparisons of the rHAd26/MVA vaccine and the diluted dose of the rVSVΔG-ZEBOV-GP vaccine with placebo. Under version 4.0 the target was reached and 2,802 participants were randomized. Thus, the total number randomized in PREVAC is 4,789 participants.

**Study Population:** Adults and children, aged at least 1 year of age, were invited to participate. The trial is conducted in Guinea, Liberia, Mali, and Sierra Leone

**Accrual Period:** 7 months to enroll 2,802 participants under Version 4.0

**Study Design:** Randomized, double-blind, placebo-controlled trial of three vaccine strategies.

**Study Duration:** Following enrolment under Version 4.0, participants will be seen at frequent follow-up visits for 12 months. After the 12 months of follow-up is complete for all participants, the study will be unblinded and all participants will be seen annually through 60 months as per protocol version 5.0.

**Background and Rationale:**

Preliminary data from an open-label, cluster randomized ring vaccination trial conducted in Guinea indicate that the Merck/New Link rVSVΔG-ZEBOV-GP vaccine prevented Ebola virus (EBOV) disease (EVD) events occurring at least 10 days after randomization among close contacts immediately vaccinated as compared to those to be potentially vaccinated 21 days later. In that trial conducted in adults, most EVD events occurred shortly following vaccination; no EVD events occurred more than 6 days after vaccination. The durability of the effect of the vaccine was not assessed.

Two other vaccines, the GlaxoSmithKline (GSK) ChAd3-EBO Z (replication deficient Chimpanzee adenovirus type 3-derived vector encoding the Ebola virus Zaire [EBO Z] GP) vaccine and the Johnson & Johnson (J&J) prime boost vaccine, rHAd26/MVA, have completed phase 1 testing. In addition, safety and immunogenicity after 12 months of the rVSVΔG-ZEBOV-GP and ChAd3-EBO Z vaccine have been evaluated in a phase 2, placebo-controlled trial of adults in Liberia (Partnership for Research on Ebola Virus in Liberia [PREVAIL] I).

By the end of 2015 in West Africa, the incidence of EVD had dramatically decreased. However, multiple questions regarding the safety and effectiveness of EVD vaccines remain unanswered, including the durability and the immediacy of immune responses generated by different vaccine strategies with and without a booster, and the safety of the different vaccines, particularly in children.

In order to answer these questions, a randomized trial that includes adults and children will be carried out. The trial will compare three vaccine strategies with placebo for immunogenicity and safety. The comparisons will be carried out separately for adults and children and for adults and children combined. The three vaccine strategies are: 1) Ad26.ZEBOV (prime) (0.5 mL) followed by an MVA-BN-Filo boost (0.5 mL) at 56 days; 2) rVSVΔG-ZEBOV-GP (prime) (1 mL) with a placebo (1 mL) boost at 56 days; and 3) rVSVΔG-ZEBOV-GP (prime) (1 mL) with a rVSVΔG-ZEBOV-GP boost (1 mL) at 56 days. There will be two placebo groups, one with a syringe fill volume of 0.5 mL to match the fill volumes of the Ad26.ZEBOV and MVA-BN-Filo vaccines and the other with a fill volume of 1 mL to match the fill volume for the rVSVΔG-ZEBOV-GP vaccine. The two placebo groups will be combined for data analyses.

Anti-GP ZEBOV antibodies will be measured and used to compare the immunogenicity profiles of each of the vaccine strategies with placebo until the placebo group is vaccinated. After the vaccination of the initial placebo group, following the outbreak in February 2021 in Guinea and the vaccine licensures, we will assess the durability of immunological response over time within groups. Correlates of protective immunity are not yet known. Thus, blood will be stored from each visit for future use when a correlate of protection is identified.

This randomized trial will provide data for the three vaccine strategies for adults and children on safety and durability (12 months and annually thereafter), immediacy (14 days), and kinetics (7, 14, and 28 days after prime vaccination and 7 and 28 to 35 days after the booster vaccination) of the immune response.

A substudy in adults in Guinea will involve exploratory analyses of T cell and memory B cell responses to the three vaccine strategies (see Appendix D).

A substudy in children will investigate viral shedding after vaccination (see Appendix E).

A substudy in Sierra Leone and Guinea will investigate the effect of malaria and helminth infections on Ebola vaccine responses (see Appendix F).

A substudy on empirical research on barriers and facilitators to study retention will be implemented in PREVAC (see Appendix G).

Under Version 4.0, participants (N=2,800) were randomly allocated to one of the three vaccine strategies or placebo with a repartition of 800 participants assigned to the rHAd26/MVA vaccine strategy and 800 to the rVSVΔG-ZEBOV-GP strategy with the placebo boost at 56 days; 400 participants were assigned to the rVSVΔG-ZEBOV-GP arm that includes an rVSVΔG-ZEBOV-GP boost at 56 days; and 400 participants were assigned to each of the two placebo groups (800 total on placebo).

The enrolment targets for adults and children were each 1,400 participants. The targets were reached. The study is powered to assess immune responses and safety outcomes separately in adults and children.

In the event of a new Ebola epidemic in the PREVAC study country or prospective risk of Ebola to study participants from Ebola, the study design, including the use of placebos, will be re-considered. Under protocol version 6.0 and version 7.0, following the new Ebola outbreak declared in Guinea in February 2021, and the recent vaccine licensures, the participants who received the placebo will be vaccinated with a single dose of rVSVΔG-ZEBOV-GP vaccine in Liberia and Mali and with the Ad26.ZEBOV/MVA-BN-Filo vaccine strategy in Guinea and Sierra Leone during their next long-term follow-up visit or during additional visits if their next scheduled long-term follow-up visit is not approaching soon. The participants who received an incomplete Ad26.ZEBOV/MVA-BN-Filo vaccine strategy will be offered a single dose of the Ad26.ZEBOV or MVA-BN-Filo vaccine in Guinea and Sierra Leone and a single dose of rVSVΔG-ZEBOV-GP in Liberia and Mali.

**Study Agent Descriptions:**

The 3 vaccine strategies to be studied are the rVSVΔG-ZEBOV-GP vaccine, with and without a boost at 56 days, and the rHAd26/MVA vaccine. The rVSVΔG-ZEBOV-GP vaccine requires a 1 mL administration from a 1 or 3 mL syringe for the prime and booster vaccination. The rHAd26/MVA requires a 0.5 mL administration of Ad26.ZEBOV from a 1 or 3 mL syringe for the prime vaccination and a booster vaccination (0.5 mL from a 1 or 3 mL syringe) 56 days later of MVA-BN-Filo.

Each study agent is briefly described below and in more detail in section 6.1.

- The prime-boost rHAd26/MVA vaccine is comprised of an Ad26.ZEBOV vaccine which consists of a single recombinant, replication incompetent human Ad26 vector, constructed to express the Ebola virus Mayinga GP. The MVA-BN-Filo boost at 56 days encodes the GP of Sudan virus (SUDV; formerly known as Ebola Virus Sudan), EBOV (formerly known as Ebola Virus Zaire), and Marburg Virus (MARV) Musoke, and the nucleoprotein of Tai Forest virus (TAFV; formerly known as Côte d'Ivoire ebolavirus) (0.5 mL intramuscular [IM] administration for the Ad26.ZEBOV and MVA-BM-Filo vaccines). The Ad26.ZEBOV vaccine and the MVA-BN-Filo boost were given at the same dose in Versions 2.0, 3.0, and 4.0 of the PREVAC protocol.

- The rVSVΔG-ZEBOV-GP vaccine is comprised of a single rVSV isolate (11481 nt) modified to replace the gene encoding the VSV G envelope GP with the gene encoding the ZEBOV envelope GP (1 mL IM administration). The rVSVΔG-ZEBOV-GP vaccine was not used in Version 2.0. The rVSVΔG-ZEBOV-GP dose was given as a 2-fold diluted dose (approximately 5 x $10^7$ plaque-forming units [pfu]/mL) in Version 3.0 and was given as an undiluted dose (geometric mean of available assays 9.4 x $10^7$ pfu/mL) in Version 4.0 of the PREVAC protocol. The doses of rVSVΔG-ZEBOV-GP used in Versions 3.0 and 4.0 are referred to as the diluted and undiluted doses, respectively. In Version 6.0 and 7.0, the vaccine dose will be consistent with the approved product with a concentration of ≥7.2 x $10^7$ pfu/mL.

**For Version 4.0 of PREVAC, the primary objectives are to compare each of the three vaccine strategies with the pooled placebo group (3 pair-wise comparisons) for safety and immunogenicity. These evaluations will be performed separately in adults and children.**

**Separately for adults and children:**

- To compare each of the three vaccine strategies with the pooled placebo group (3 pair-wise comparisons) for antibody response 12 months after randomization (prime vaccination).

In order to facilitate Merck regulatory filings and bridging of immune responses of the rVSVΔG-ZEBOV-GP vaccine from this study to other studies and between pediatric and adult populations, the following objective will be assessed specifically for those in the two rVSVΔG-ZEBOV-GP vaccine groups:

- To compare the rVSVΔG-ZEBOV-GP vaccine (pooled rVSVΔG-ZEBOV-GP groups) with the matched placebo group for antibody response 28 days after randomization (prime vaccination).

In order to facilitate Janssen regulatory filings of the rHAd26/MVA vaccine, the following objective will be assessed specifically for those in the rHAd26/MVA vaccine group:

- To compare the rHAd26/MVA vaccine group with the matched placebo group for antibody response 3 months after randomization (approximately 28 days after the booster vaccination).

**Primary Endpoints**

- GP-EBOV antibody response at 12 months. The primary analysis for adults and children will be performed for participants who do not have elevated antibody levels at baseline.
- GP-EBOV antibody response at Day 28 after randomization (initial vaccination). For rVSVΔG-ZEBOV-GP arms only, Day 28 post first vaccination will be used for regulatory purposes for comparison to other studies and for

bridging children to adults. The primary analysis for adults and children will be performed for participants who do not have elevated antibody levels at baseline.

- GP-EBOV antibody response at Month 3 after randomization (approximately 28 days after the booster vaccination). For the rHAd26/MVA vaccine arm only, Month 3 will be used for regulatory purposes. The primary analysis for adults and children will be performed for participants who do not have elevated antibody levels at baseline.

**Primary Hypotheses**

The null hypothesis for each of the three comparisons at 12 months is that the vaccine strategy does not differ from the pooled placebo group (0.5 mL and 1.0 mL).

The following hypothesis will only apply to the rVSVΔG-ZEBOV-GP vaccine for the purpose of facilitating Merck regulatory filings:

> The null hypothesis is that the antibody response for the pooled rVSVΔG-ZEBOV-GP group does not differ from the matched placebo group (0.5 mL and 1.0 mL groups) at Day 28 after randomization (initial vaccination).

The following hypothesis will only apply to the rHAd26/MVA vaccine for the purpose of facilitating Janssen regulatory filings:

> The null hypothesis is that the antibody response for the rHAd26/MVA vaccine does not differ from the matched placebo group (0.5 mL and 1.0 mL groups) at Month 3 after randomization.

**Secondary and Exploratory Objectives**

There are a number of secondary objectives that include comparisons of each vaccine strategy versus placebo for: 1) serious adverse events (SAEs; see section 4.1.3 for definition), injection site reactions, targeted symptoms of any grade severity, and grade 3 or 4 adverse events (AEs), in the 28 to 35 days following both the prime and booster vaccinations; and 2) antibody levels measured 14 days after vaccination to assess the immediacy of the response and for up to 5 years to assess the long-term durability of the antibody response.

As a secondary objective, adults and children will be combined for comparisons of each vaccine strategy with placebo.

In the event that the same correlate of protection is identified for each vaccine, exploratory objectives that compare the three vaccine strategies with one another for the correlate will be carried out using measurements on stored blood samples.

These secondary and exploratory objectives are stated in sections 2.4 and 2.5.

**Inclusion Criteria:**

- Informed consent/assent
- Age ≥ 1 year
- Planned residency in the area of the study site for the next 12 months
- Willingness to comply with the protocol requirements

**Exclusion Criteria:**

- Fever > 38º Celsius
- History of EVD (self-report)
- Pregnancy (a negative urine pregnancy test is required for females of child-bearing potential, i.e., females who have experienced menarche or who are aged 14 years and older)
- Positive human immunodeficiency virus (HIV) test for participants < 18 years of age
- Reported current breast-feeding
- Prior vaccination against Ebola (self-report)
- Any vaccination in the past 28 days or planned within the 28 days after randomization (initial vaccination)
- In the judgement of the clinician, any clinically significant acute/chronic condition which would limit the ability of the participant to meet the requirements of the study protocol

The demographics of those enrolled was closely monitored to ensure that the targeted number of adults (N=1,400) and children (N=1,400) were enrolled.
The targets were reached.

**Procedures:**
Vaccination centers were established in West Africa to consent, vaccinate and follow study volunteers. A case report form (CRF) with a unique participant identification code (PID) will be completed for each participant consented and randomized. The CRF may be paper or an electronic CRF (eCRF). The procedures described in this protocol are in terms of a paper CRF.

At the time of prime vaccination, a tear-off label on the syringe that includes a unique syringe identification code (SID) was attached to the baseline CRF. The SID is a unique number from a centrally prepared randomization schedule. This is the primary link between the blinded vaccine assignment and the PID. Study volunteers and clinical staff following the participants for safety and immunogenicity outcomes are fully blinded until the unblinding to be done after the completion of the M12 data collection (see section 2.9).

At 56 days, a syringe with the corresponding PID was prepared for each participant on his/her scheduled 56 day follow-up visit.

At study entry, study volunteers were given an identification card that identifies them as study participants. The identification card will include contact information in the event they experience vaccine side effects or signs and symptoms of Ebola.

Participants were instructed to avoid sharing items during the period of potential virus shedding. Women of childbearing potential were told to use adequate birth control measures following randomization and until at least 3 months following the booster vaccination. Men who are sexually active with women of child-bearing potential were told to use condoms following randomization and until at least 3 months following the booster vaccination.

Following randomization, all participants were seen at 7, 14, 28, 56 and 63 days, and at 3, 6, and 12 months. The study will be unblinded after all participants have completed 12 months of follow-up and after the clinical and safety database lock for the M12 data. All participants will be seen annually through 60 months.

In addition, during the first week following the prime and booster vaccination, there was daily contacts with participants < 18 years of age enrolled to assess injection site reactions and AEs graded for severity, including targeted symptoms, and SAEs.

Participants are encouraged to contact the vaccination center by telephone if they have any AEs. If the participant experiences an AE requiring immediate attention, they will be given instructions on which hospital to go to for evaluation and treatment.

**For Version 5.0 of PREVAC, the secondary objectives are (separately for adults and children):**

- To compare the long-term safety at month 24, 36, 48 and 60 following the three vaccine strategies with the pooled placebo group.
- To determine whether exposure to malaria and helminth infections influences the durability of the humoral and cellular immune responses to Ebola vaccines (see Appendix F).
- To identify multilevel facilitators and barriers to participant retention in PREVAC through retrospective qualitative research with participants after the conclusion of the blinded trial period (first 12 months) (see Appendix G).
- To identify factors that predict participant retention and loss-to-follow-up (LTFU) during the unblinded PREVAC long-term follow-up period (from 12 months up to 5 years) through prospective research (see Appendix G)
- To analyze the early vaccine signature, identify early correlates of durable antibody responses and propose *in silico* methods to optimally allocate vaccine strategies at the individual level.

These secondary and exploratory objectives are stated in sections 2.4 and 2.5

**For Version 6.0 and Version 7.0 of PREVAC, the secondary objectives are (separately for adults and children):**

- To assess the long-term antibody response of three vaccine strategies at 24, 36, 48 and 60 months following randomization.
- To assess the long-term safety at month 24, 36, 48 and 60 following the three vaccine strategies.
- In a subsample of adults in Guinea, to assess the T cell and memory B cell responses for the three vaccine strategies (see Appendix D).
- To determine whether exposure to malaria and helminth infections influences the durability of the humoral and cellular immune responses to Ebola vaccines (see Appendix F).
- To identify multilevel facilitators and barriers to participant retention in PREVAC through retrospective qualitative research with participants after the conclusion of the blinded trial period (first 12 months) (see Appendix G).

- To identify factors that predict participant retention and loss-to-follow-up (LTFU) during the unblinded PREVAC long-term follow-up period (from 12 months up to 5 years) through prospective research (see Appendix G)
- To analyse the early vaccine signature, identify early correlates of durable antibody responses and propose *in silico* methods to optimally allocate vaccine strategies at the individual level.

These secondary and exploratory objectives are stated in sections 2.4 and 2.5

**Data and Safety Monitoring Board:**
An independent Data and Safety Monitoring Board (DSMB) closely monitored accumulating safety data for adults and children in each age group (1-4, 5-11, and 12-17 years). Safety data for children were reviewed at least monthly until the DSMB is satisfied that the vaccines are safe at which time less frequent reviews may be carried out.

The safety reviews and strategy for sequentially enrolling increasingly younger children outlined below were conducted in Version 3.0 of the PREVAC protocol for participants receiving Ad26.ZEBOV, diluted rVSVΔG-ZEBOV-GP, or placebo. A similar strategy for safety reviews and sequentially enrolling younger children was conducted for participants receiving Ad26.ZEBOV, undiluted rVSVΔG-ZEBOV-GP, or placebo under Version 4.0. Initially, in each version, the trial only enrolled adults and children ≥12 to ≤17 years of age. After 70 children aged ≥12 to ≤17 years were enrolled (20 vaccinated with Ad26.ZEBOV, 30 vaccinated with rVSVΔG-ZEBOV-GP, and 20 vaccinated with placebo) and followed for 28 days, the DSMB was asked to consider whether vaccination of children ≥5 to ≤11 years can commence. Similarly, after 70 children aged ≥5 to ≤11 years were enrolled, the DSMB was asked to consider whether vaccination of children at least 1 year of age to ≤ 4 years could commence. Enrollment did not pause after 70 children were accrued in an age group. Once the decision was made that the enrolment of children 1 to 4 years was safe, the goal was to target the enrolment of the 1,400 children to achieve a similar distribution in the 3 age groups.

**Amendment (Version 2.0 of the PREVAC Protocol)**
Under Version 2.0 of the PREVAC protocol 537 participants ≥ 12 years of age were randomized to the prime-boost rHAd26/MVA vaccine strategy or to matching placebo. The decision to begin the trial with the prime-boost rHAd26/MVA vaccine strategy and matching placebo and not include the rVSVΔG-ZEBOV-GP vaccine arms was reached one week before PREVAC was scheduled to begin with 5 randomized groups as originally planned (Version 1.0) because of questions regarding the occurrence of arthritis in 3 of 9 participants in a recent NIH study (described in Section 1.1.3) receiving vaccine from the lot of the rVSVΔG-ZEBOV-GP vaccine which was to be used in PREVAC.

**Amendment (Version 3.0 of the PREVAC Protocol)**

Under Version 3.0 of the PREVAC protocol 1,450 participants were randomized to one of five groups: 1) Ad26.ZEBOV (prime) followed by an MVA-BN-Filo boost at 56 days; 2) rVSVΔG-ZEBOV-GP (prime, diluted dose) with a placebo boost at 56 days; 3) rVSVΔG-ZEBOV-GP (prime, diluted dose) with a rVSVΔG-ZEBOV-GP boost (diluted) at 56 days; 4) placebo for rVSVΔG-ZEBOV-GP; or 5) placebo for the Ad26.ZEBOV and MVA-BN-Filo vaccines.

The rVSVΔG-ZEBOV-GP vaccine was given at a 2-fold dilution in Version 3.0 because the certificate of analysis of the vaccine lot was found to be higher than that used in the PREVAIL I trial in Liberia. Variation in titer/potency in live virus vaccines is common. Vaccine manufacture and release for potency is based upon defined specifications and always encompasses a range, routinely with a lower and upper limit. The lower limit, referred to as the nominal dose, is determined during development and is defined by the lowest dose for which there is demonstrated efficacy. The lower limit for potency must still be valid at the end of shelf-life in order to ensure that the vaccine is still efficacious up until its defined expiry.

Since there were limited numbers of children in previous studies of the rVSVΔG-ZEBOV-GP vaccine, Version 3.0 of PREVAC used a measured approach with dilution of the rVSVΔG-ZEBOV-GP vaccine.

The Version 3.0 amendment also updated the safety information for the rVSVΔG-ZEBOV-GP vaccine, removed language about the initial phase of PREVAC described in Version 2.0, and included minor edits to the Saliva Sample Substudy in Appendix E. The data collection plan for Version 3.0 was the same as that described in Version 1.0 and Version 2.0.

**Amendment (Version 4.0 of the PREVAC Protocol)**

Version 4.0 of the protocol did not begin until at least 70 children in each of the three age groups (1-4, 5-11, and 12-17 years) were enrolled in Version 3.0 and the DSMB reviewed safety data through 28 days for each age group.

Under Version 4.0 of the PREVAC protocol, a target of 2,800 participants (1,400 adults and 1,400 children) to be randomized to one of five groups was reached: 1) Ad26.ZEBOV (prime) followed by an MVA-BN-Filo boost at 56 days; 2) rVSVΔG-ZEBOV-GP (prime, undiluted dose) with a placebo boost at 56 days; 3) rVSVΔG-ZEBOV-GP (prime, undiluted dose) with a rVSVΔG-ZEBOV-GP boost (undiluted) at 56 days; 4) placebo for rVSVΔG-ZEBOV-GP; or 5) placebo for the Ad26.ZEBOV and MVA-BN-Filo vaccines (Figure 1).

In addition to the dosing information for the rVSVΔG-ZEBOV-GP vaccine, the version 4.0 amendment also updated the safety information for the rVSVΔG-ZEBOV-GP vaccine, and amended the Immunological Substudy in Appendix D and the Saliva Substudy in Appendix E. The Immunological Substudy was amended to state that up to 230 participants were to be enrolled in Versions 2.0, 3.0, and 4.0 (in total). The Saliva Substudy was amended to state that the target sample size was 140 children, with approximately an equal distribution of children in each of the three age groups (1-4, 5-11, and 12-17 years), for both Version 3.0 and Version 4.0 of PREVAC.

While the primary objectives of PREVAC will be accomplished with participants enrolled under Version 4.0, the participants randomized under Versions 2.0 and 3.0 will contribute to the evaluation of each of the objectives that compares the rHAd26/MVA vaccine strategy to placebo. In addition, participants enrolled under Version 3.0 of the protocol will provide information on the safety and immunogenicity of the diluted dose of the rVSVΔG-ZEBOV-GP vaccine compared to placebo for adults and children in each of the three age groups (1-4, 5-11, and 12-17 years).

**Amendment (Version 5.0 of the PREVAC Protocol)**

Under Version 5.0 of the PREVAC protocol, all the participants randomised in the previous versions of the protocol (Versions 2.0, 3.0, and 4.0) will be followed annually for 4 additional years.

In addition, the Immunological Substudy in Appendix D is amended to state that the 230 participants enrolled in Versions 2.0, 3.0, and 4.0 will be followed annually for 4 additional years. A substudy on malaria and helminth infections and empirical research on barriers and facilitators to study retention have been added in Appendix F and Appendix G, respectively.

While the long-term objectives of PREVAC will be accomplished with participants enrolled under Version 4.0, the participants randomized under Versions 2.0 and 3.0 will contribute to the evaluation of each of the objectives that compare the rHAd26/MVA vaccine strategy to placebo. In addition, participants enrolled under Version 3.0 of the protocol will provide information on the safety and immunogenicity of the diluted dose of the rVSVΔG-ZEBOV-GP vaccine compared to placebo for adults and children in each of the three age groups (1-4, 5-11, and 12-17 years).

**Amendment (Version 6.0 of the PREVAC Protocol)**

Under Version 6.0 of the PREVAC protocol, as for protocol version 5.0, all the participants randomised in the previous versions of the protocol (Versions 2.0, 3.0, and 4.0) continue to be followed annually for 4 additional years post the M12 visit.

The participants who received the placebo in previous versions will be vaccinated with a single dose of rVSVΔG-ZEBOV-GP vaccine in Liberia and Mali and with Ad26.ZEBOV/MVA-BN-Filo vaccine strategy in Guinea and Sierra Leone. The participants who received an incomplete Ad26.ZEBOV/MVA-BN-Filo vaccine strategy will be offered a single dose of the Ad26.ZEBOV or MVA-BN-Filo vaccine in Guinea and Sierra Leone and a single dose of rVSVΔG-ZEBOV-GP in Liberia and Mali.

While the long-term objectives of PREVAC will be accomplished with participants enrolled under Version 4.0, the participants randomized under Versions 2.0 and 3.0 will contribute to the evaluation of each of the objectives that provide information on the safety and immunogenicity of the rHAd26/MVA vaccine strategy. In addition, participants enrolled under Version 3.0 of the protocol will provide information on the safety and immunogenicity of the diluted dose of the rVSVΔG-ZEBOV-GP vaccine for adults and children in each of the three age groups (1-4, 5-11, and 12-17 years).

**Amendment (Version 7.0 of the PREVAC Protocol)**
Under protocol version 7.0, the same procedure as for protocol version 6.0 will be followed with the addition of two non vaccination criteria.

### 1. INTRODUCTION

#### 1.1. Background

##### 1.1.1. Recent Ebolavirus Zaire outbreaks

The Zaire EVD outbreak In West Africa was first recognised on 22 March 2014 in Guinea. By December 1, 2014, the epidemic had spread to Liberia and Sierra Leone. Through September 2015, over 28,000 confirmed cases of EVD with 11,000 deaths had occurred in these 3 West African countries. By the end of 2015 in West Africa, the incidence of EVD had dramatically decreased. Since the beginning of 2016, there have been flare-ups of Ebola in Sierra Leone, Guinea and Liberia but response teams in each country have minimized transmission.[1]

Since that, four more Ebola outbreaks have been recorded in the Democratic Republic of Congo (DRC). The Equateur outbreak started in May 2018 and lasted 3 months before being declared over. A total of 54 people were infected and 33 lives claimed, resulting in a case fatality rate of 61%[2].

The second and larger outbreak was declared in North Kivu on August 1st, 2018 spanning different health zones across North Kivu and Ituri Provinces.[3] The outbreak was declared over on June 25th, 2020 with 3,470 people diagnosed and 2,287 deaths meaning a 65,9 % fatality rate. This outbreak was the second largest in the world and presented particular challenges due to the fact that it occurred in an area of active conflict[4] Ring vaccination with rVSVΔG-ZEBOV-GP was deployed as part of the outbreak control efforts and more than 303,000 individuals (contacts, contacts of contacts and HCWs/FLWs) were vaccinated.[4,5,6] Preliminary assessment indicates high levels of vaccine efficacy as discussed in section 1.1.3. On June 1st, 2020, seven cases of Ebola virus disease were reported in the city of Mbandaka and the neighbouring health zone of Bikoro in Equateur Province, leading to the declaration of a new EVD outbreak concomitant to the COVID19 pandemic. The outbreak was declared over on November 9th, 2020. 119 confirmed cases were registered, 11 probable, 55 dead and 75 recovered (42,3% mortality). A new outbreak of Ebola virus disease was declared on February 7th, 2021 at Biena and Katwa health zones, in the eastern part of the DRC and is the result of a resurgence of the strain responsible for the previous DRC north Kivu outbreak. As of March 31st, 2021, 12 cases: 6 deaths, 6 recoveries have been reported. A vaccination campaign was started on February 15th, 2021.[7]

Finally, seven years after declaration of the first EVD epidemic in Guinea, a new outbreak emerged on February 14th, 2021 in the sub-prefecture of Gouécké, in the Nzérékoré Region. On May 26th, 2021, 23 cases ont été declares (16 confirmed and 7 probables): 12 deaths have been registered. Vaccination campaigns were launched on February 23rd, 2021. Interestingly the genomic sequencing of EVD positive samples in Guinea showed that this new outbreak is the result of a resurgence of the strain responsible for the 2014-2016 West African outbreak.

The genus Ebolavirus is one of three genera in the family Filoviridae, which along with the genera Marburgvirus and Cuevavirus, are known to induce viral haemorrhagic fever. The 5 distinct species included in the genus Ebolavirus are Bundibugyo (BDBV), Reston (RESTV), SUDV, TAFV, and ZEBOV.

Ebola virus is a large, negative-strand ribonucleic acid (RNA) virus composed of 7 genes encoding viral proteins, including a single GP. The virus is responsible for causing EVD in humans. In particular, BDBV, ZEBOV, and SUDV have been associated with outbreaks of EVD in Africa and reported case fatality rates of up to 90% before vaccination became available.[8] Transmission of EBOV to humans is not yet fully understood, but is likely due to incidental exposure to infected animals. EBOV then spreads through human-to-human transmission, with infection resulting from direct contact with blood, secretions, organs or other bodily fluids of infected people, and indirect contact with environments contaminated by such fluids.

EVD has an incubation period of 2 to 21 days (mean 4 to 10 days). The clinical manifestations have been summarized by Feldmann and Geisbert.[9] They report that infection is followed by an abrupt onset of non-specific symptoms such as fever, chills, malaise, and myalgia. The subsequent signs and symptoms indicate multisystem involvement and include systemic (prostration), gastrointestinal (anorexia, nausea, vomiting, abdominal pain, diarrhoea), respiratory (chest pain, shortness of breath, cough, nasal discharge), vascular (conjunctival injection, postural hypotension, edema), and neurological (headache, confusion, coma) manifestations. Hemorrhagic manifestations consistent with disseminated intravascular coagulation may arise during the peak of the illness and include petechiae, ecchymoses, uncontrolled oozing from venepuncture sites, mucosal haemorrhages, and post-mortem evidence of visceral haemorrhagic effusions. A maculopapular rash associated with varying severity of erythema and desquamation can often be noted by day 5 to 7 of the illness; this symptom is a valuable differential diagnostic feature and is usually followed by desquamation in survivors. Abdominal pain is sometimes associated with hyperamylasemia and true pancreatitis. In later stages, shock, convulsions, severe metabolic disturbances, and, in more than half the cases, diffuse coagulopathy supervene. Laboratory findings include low white blood cell and platelet counts and elevated liver enzymes. In general, the symptoms last for about 7 - 14 days, after which recovery may occur. Death usually occurs 6 to 16 days after the onset of symptoms.

The clinical manifestations of patients with EVD who were treated at an Ebola treatment unit in Monrovia, Liberia between August and October, 2014 have been described.[10] Similar to the review by Feldmann and Geisbert,[9] these authors reported that early symptoms of EVD included high fever (up to 40ºC), malaise, fatigue and body aches. After 3 to 5 days, gastrointestinal symptoms began with epigastric pain, nausea, vomiting and diarrhea. Other signs and symptoms included headache, conjunctival infection, chest pain, abdominal pain, arthralgias, myalgias, and hiccups. Clinically significant hemorrhage from the upper or lower gastrointestinal tract or both occurred in fewer than 5% of patients. Similar symptoms were reported for patients with laboratory confirmed EVD in Conakry, Guinea. For 37 patients who presented to the hospital in March and April 2014, the most common symptoms were fever, fatigue, diarrhea, headache, vomiting and anorexia. These patients were hospitalized a median of 5 days after symptom onset.[11]

Recently published long-term follow up studies from Ebola virus disease survivors from the West African outbreak (PostEboGui cohort study), showed increased blood markers of inflammation, intestinal tissue damage, T cell and B cell activation and a depletion of circulating dendritic cells compared to healthy subjects, suggesting the persistence of a chronic Ebola virus disease.[12]

Among individuals who survive Ebola, the virus may persist in some body fluids for an extended period of time. In 1977, a case report described an investigator who accidently inoculated himself with EBOV and the virus was recovered in his semen 61 days after onset of illness, but not on days 76, 92, and 110.[13] During an outbreak of Ebola in the Congo in 1995, infectious virus was detected in the semen 82 days after disease onset in one patient.[14] In March 2015, it was reported that a woman in Liberia possibly developed Ebola through unprotected sex with an Ebola survivor.[15] Subsequently, genomic evidence confirmed the sexual transmission with an Ebola survivor for whom there was evidence of persistence of infective EBOV in semen for 179 days or more.[16] Recent reports from Liberia and Guinea indicate a large fraction of men have EBOV in their semen after their recovery.[17,18] These results have been further confirmed in a recent study assessing Ebola viral RNA presence in semen, urine, cervicovaginal fluid, saliva, breast milk, and feces from EVD survivors in Guinea (PostEbogui cohort, n = 802) at regular intervals until 40 months after inclusion, showing long-term presence of viral RNA in semen and, unexpectedly, in breast milk.[19]

An overview has summarized the literature on viral persistence in survivors of Ebola.[20]

##### 1.1.2. J&J Ad26.ZEBOV/MVA-BN-Filo vaccine regimen

Janssen Vaccines and Prevention BV (a Janssen pharmaceutical company of Johnson & Johnson), in collaboration with Bavarian Nordic GmbH, is investigating the potential of a prophylactic Ebola vaccine regimen comprised of the following 2 candidate Ebola vaccines:

**Ad26.ZEBOV** is a monovalent vaccine expressing the full length Mayinga GP of the EBOV (formerly known as *Zaire ebolavirus*), and is produced in the human cell line PER.C6®.

**MVA-mBN226B**, further referred to as MVA-BN-Filo®, is a multivalent vaccine expressing the SUDV GP, the EBOV GP, the MARV Musoke GP, and the TAFV nucleoprotein, and is produced in chicken embryo fibroblast cells. The EBOV GP expressed by MVA-BN-Filo has 100% homology to the one expressed by Ad26.ZEBOV.

The EBOV GP at the origin of the West African outbreak had 97% homology to the EBOV GP used in these vaccine regimens.

The PREVAC protocol refers to the Ad26.ZEBOV as prime and to MVA-BN-Filo as boost. These 2 vaccines are also referred to as a heterologous two dose vaccine regimen by J&J.

Ad26.ZEBOV/MVA-BN-Filo vaccine were licensed by European Medicines Authority (EMA) on May 2020 under the commercial names of Zabdeno® and Mvabea® corresponding to the two components. Zabdeno® is given first and Mvabea® is administered approximately eight weeks later as a booster. Zabdeno®/Mvabea® is a vaccine regimen to protect adults and children aged one year and older against Ebola virus disease caused by Zaire ebolavirus.
As per EMA recommendations, this prophylactic 2-dose regimen is not suitable for an outbreak response where immediate protection is necessary. As a precautionary measure, a Zabdeno® booster vaccination should be considered for individuals at imminent risk of exposure to Ebola virus, for example healthcare professionals and those living in or visiting areas with an ongoing Ebola virus disease outbreak, who completed the Zabdeno®, Mvabea® 2-dose primary vaccination regimen more than four months ago.[21]

#### Nonclinical Studies

##### *Immunogenicity and Efficacy*

Immunogenicity and efficacy of the vaccine combination Ad26.ZEBOV and MVA-BN-Filo was evaluated in an non-human primate (NHP) model (i.e., Cynomolgus macaques, *Macaca fascicularis*). The combination was assessed in a multivalent filovirus setting in a small number (2 per regimen) of animals and the study included heterologous prime-boost regimens of Ad26, Ad35 and MVA-BN-Filo vectors expressing different Ebola and Marburg proteins. Full protection from EVD and death after wild-type EBOV Kikwit 1995 challenge was obtained with all heterologous regimens, including the Ad26.ZEBOV and MVA-BN-Filo vaccine regimen. All heterologous prime-boost regimens induced comparable immune responses against the EBOV Mayinga GP. Independently of the vaccine regimen, a strong boost effect was seen after heterologous prime-boost immunization.[22]

Two additional studies involving more animals have been performed, to strengthen the robustness of the nonclinical efficacy data, and also to optimize the prime-boost schedule so as to obtain induction of protective immunity as quickly as possible, to specifically respond to the EVD outbreak in West Africa. Complete survival was observed with 8-week heterologous regimens with Ad26.ZEBOV as prime and MVA-BN-Filo as boost. Partial protection was observed with both MVA-BN-Filo as a prime immunization and a shorter prime/boost interval.[23, 24]

##### *Toxicology*

A repeated-dose toxicity study in rabbits was performed with prime-boost combinations of Ad26.ZEBOV and MVA-BN-Filo. The different dose regimens were well tolerated when administered twice by IM injection to New Zealand White rabbits with a 14-day interval period. All vaccine dosing regimens resulted in detectable EBOV GP-specific antibody titers. No significant toxicological effects (no adverse effects) were observed. The immune response was associated with transient increases in fibrinogen, C-reactive protein, globulin, decreases in hematocrit and hemoglobin, and microscopic findings in draining iliac lymph nodes, spleen and at the injection sites. The findings were noted to be recovering over a 14-day treatment-free period and were considered to reflect a physiological response associated with vaccination. There were no effects noted that were considered to be adverse.

In an embryo fetal and pre- and postnatal development study in female rabbits, there was no maternal or developmental toxicity following maternal exposure to the vaccine regimens during the premating and gestation period. All vaccine regimens elicited detectable EBOV GP-specific maternal antibody titers that were transferred to the fetuses.

##### *Biodistribution*

Single-dose biodistribution studies in rabbits were performed using the MVA-BN vector or the Ad26 vector in combination with another insert (Ad26.ENVA.01: an experimental, prophylactic Ad26 vector expressing the HIV type 1, Clade A envelope protein). MVA-BN distributed to the skin, muscle, blood, spleen, lung, liver, and pooled lymph nodes and was rapidly cleared (within 48 hours following vaccination). Ad26.ENVA.01 was primarily localized in the injection site muscle, the regional lymph nodes and the spleen. Three months after the single IM injection of Ad26.ENVA.01, the vaccine was cleared from most of the examined tissues. As biodistribution is dependent on the vector platform (MVA-BN or Ad26) and not on the insert, it can be assumed that recombinant MVA-BN-Filo or Ad26.ZEBOV is distributed in the same way as the MVA-BN vector or Ad26.ENVA.01 vector, respectively.

#### Clinical Studies

Currently, the clinical program to assess the safety/tolerability and immunogenicity of Ad26.ZEBOV and MVA-BN-Filo include six Phase 1 studies both in LMIC and HIC (all six completed: VAC52150EBL1001, VAC52150EBL1002, VAC52150EBL1003, VAC52150EBL1004, EBL05 and HHSN2722013, four Phase

2 (completed) (VAC52150EBL2001, VAC52150EBL2002, VAC52150EBL2003 and VAC52150EBL2005) and four on going (VAC52150EBL2007, NCT04028349, VAC52150EBL2011 and PREVACEBL3005) and eight Phase 3 clinical studies (VAC52150EBL3002, VAC52150EBL3003, and VAC52150EBL3001 completed; VAC52150EBL3005, VAC52150EBL4001, DRC-EB-001, VAC52150EBL3004, VAC52150EBL3010 ongoing), where monovalent Ad26.ZEBOV vaccine and multivalent MVA-BN-Filo vaccine are combined in homologous or heterologous prime-boost regimens in which each vector is used to prime a filovirus-specific immune response followed by a boost immunization with the same or the other vector 2 to 12 weeks later. As of April 2021, more than 32,000 people have been enrolled in the studies. Two additional Phase 1 studies investigating MVA-BN-Filo have been performed (EBL01 and CVD-Mali Ebola Vaccine #1000).[24]

##### *Phase 1/2/3 studies in adults*

To date, the results of the primary analysis of Phase 1 clinical studies with Ad26.ZEBOV and/or MVA-BN-Filo are available (VAC52150EBL1001, VAC52150EBL1002, VAC52150EBL1003 and VAC52150EBL1004) as well as the results of the phase 2 trial VAC52150EBL2001.[23,24,25 26,27,28]

VAC52150EBL1001, a first-in-human study, is a randomized, placebo-controlled, observer-blind study in healthy adults evaluating the safety, tolerability and immunogenicity of 4 placebo-controlled regimens using MVA-BN-Filo at a dose of $1x10^8$ 50% tissue culture infective dose ($TCID_{50}$) and Ad26.ZEBOV at a dose of $5x10^{10}$ viral particles (vp): 2 regimens with MVA-BN-Filo as prime and Ad26.ZEBOV as boost at a 28- or 56-day interval, and 2 regimens with Ad26.ZEBOV as prime and MVA-BN-Filo as boost at a 28- or 56-day interval. A fifth regimen, with Ad26.ZEBOV at a dose of $5x10^{10}$ vp as prime, and MVA-BN-Filo at a dose of $1x10^8$ $TCID_{50}$ at a 14-day interval was evaluated in an open-label, uncontrolled fashion in healthy adults. VAC52150EBL1001 has enrolled 87 subjects and primary analysis data for safety and immunogenicity are available for 85 subjects (performed when all subjects had completed their 21-day post-boost visit or discontinued earlier). Four SAEs occurred; none of these was considered related to the study vaccines. Two subjects in the 14-day regimen group who experienced grade 3 neutropenia did not receive the boost vaccination as they met criteria for contraindications to boost (specified in the protocol), but continued scheduled assessments as planned. There were no SAEs related to the study vaccines.[25]

In the placebo-controlled regimens, overall frequencies of solicited local and systemic AEs were higher after MVA-BN-Filo and Ad26.ZEBOV, relative to placebo. In addition, there was a trend towards higher overall frequencies of solicited local and solicited systemic events after Ad26.ZEBOV, relative to MVA-BN-Filo. This trend was not observed in the open-label 14-day regimen. The most frequent solicited local AE was injection site pain. The most common solicited systemic AEs were fatigue, headache and myalgia. Grade 3 solicited local AEs occurred in 3 subjects (injection site pain and injection site swelling in 1 subject each, injection site erythema in 2 subjects). These events occurred after

Ad26.ZEBOV vaccination. Grade 3 solicited systemic events were also reported in 3 subjects (nausea and headache in 2 subjects each, myalgia and fatigue in 1 subject each). All these events occurred after Ad26.ZEBOV, except fatigue, which occurred after placebo. The systemic events after Ad26.ZEBOV were considered to be at least possibly related to vaccination by the investigator. In the open-label regimen, no subjects experienced grade 3 solicited local events. Grade 3 solicited systemic events occurred in 3 subjects after Ad26.ZEBOV (chills and fatigue in 3 subjects each, headache in 2 subjects, pyrexia and nausea in 1 subject each). All were considered to be at least possibly related to vaccination by the investigator. No grade 3 solicited systemic events were reported after MVA-BN-Filo. All solicited (local or systemic) AEs were transient in nature and resolved without sequelae. In the placebo-controlled regimens, the overall frequency of unsolicited AEs was generally comparable between Ad26.ZEBOV, MVA-BN-Filo, and placebo recipients. In the open-label regimen, unsolicited AEs were more common after Ad26.ZEBOV than after MVA-BN-Filo.

Overall, the reported AEs following vaccination were mild in the majority of subjects, transient in nature, and resolving without sequelae. These findings are consistent with the safety profiles observed for similar vaccines. The safety profile of the individual vaccines when used as prime was comparable to that when used as boost.

There was no apparent influence of the prime-boost interval (28 days or 56 days) on the safety profiles of the vaccines. The 14-day prime-boost interval seemed to be associated with higher frequencies of solicited AEs relative to the 28-day and 56-day prime-boost intervals; however, due to the open-label design of the 14-day regimen, knowledge of the treatment assignment may have biased subjects' reporting.

Both vaccination sequences tested (i.e., Ad26.ZEBOV prime followed by MVA-BN-Filo boost and MVA-BN-Filo prime followed by Ad26.ZEBOV boost) were highly immunogenic and induced considerable humoral and cellular immune responses. Extending the interval between the prime and boost led to increased antibody responses (magnitude at a 56-day prime-boost interval was about 1.8 times higher than at a 28-day prime-boost interval), while the effect was less pronounced or the reverse for T cell responses. The induced immune responses were functional, as demonstrated by the neutralizing activity of the antibody responses in all subjects. The composition of induced T cell response was favorable, with a high percentage of polyfunctional T cells, which generally are thought to play a role in immunological memory and effector functions.

VAC52150EBL1002 is a randomized, placebo-controlled, observer-blind study in healthy adults evaluating the safety, tolerability and immunogenicity of heterologous and homologous prime-boost regimens using MVA-BN-Filo and Ad26.ZEBOV administered in different doses, sequences and schedules: MVA-BN-Filo ($1x10^8$ $TCID_{50}$) as prime followed by a Ad26.ZEBOV ($5x10^{10}$ vp) as boost at 14, 28, or 56 days after prime; Ad26.ZEBOV ($5x10^{10}$ vp) as prime followed by MVA-BN-Filo ($1x10^8$ $TCID_{50}$) as boost at 28 days after prime; Ad26.ZEBOV

($5x10^{10}$ vp) as prime followed by a high dose of MVA-BN-Filo ($4.4x10^{8}$ $TCID_{50}$) as boost at 14 days after the prime; a high dose of Ad26.ZEBOV ($1x10^{11}$ vp) as prime followed by a high dose of MVA-BN-Filo ($4.4x10^{8}$ $TCID_{50}$) as boost at 28 days after the prime. All planned 128 subjects in VAC52150EBL1002 have received a prime dose, and 126 subjects have received a boost dose (2 subjects were withdrawn). No vaccine-related SAEs have been reported and no safety issues have been identified to date.

VAC52150EBL1003 and VAC52150EBL1004 are randomized, placebo-controlled, observer-blind studies in healthy adults evaluating the safety, tolerability and immunogenicity of 4 regimens using MVA-BN-Filo at a dose of $1x10^{8}$ $TCID_{50}$ and Ad26.ZEBOV at a dose of $5x10^{10}$ vp: 2 regimens had MVA-BN-Filo as prime and Ad26.ZEBOV as boost at a 28- or 56-day interval, and 2 regimens had Ad26.ZEBOV as prime and MVA-BN-Filo as boost at a 28- or 56-day interval.

In VAC52150EBL1003 72 volunteers aged 18-50 years were randomized into 4 groups of 18 (15 received vaccine, and 3 received placebo). The most frequent solicited systemic adverse event was headache (frequency, 50%, 61%, and 42% per dose for MVA-BN-Filo, Ad26.ZEBOV, and placebo, respectively). The most frequent solicited local AE was injection site pain (frequency, 78%, 63%, and 33% per dose for MVA-BN-Filo, Ad26.ZEBOV, and placebo, respectively). No differences in adverse events were observed among the different vaccine regimens. High levels of binding and neutralizing anti–Ebola virus glycoprotein antibodies were induced by all regimens and sustained to day 360 after the first dose.[26]

In VAC52150EBL1004, 72 adults were randomized to receive vaccine (n = 60) or placebo (n = 12). No vaccine-related serious adverse events were reported. The most frequent solicited local and systemic adverse events were injection site pain (frequency, 70%, 66%, and 42% per dose for MVA-BN-Filo, Ad26.ZEBOV, and placebo, respectively) and headache (57%, 56% and 46%, respectively). Adverse event patterns were similar among regimens. Twenty-one days after dose 2, 100% of volunteers demonstrated binding antibody responses against Ebola virus glycoprotein and 87%–100% demonstrated neutralizing antibody responses. Ad26.ZEBOV dose 1 vaccination induced more-robust initial binding antibody and cellular responses than MVA-BN-Filo dose 1 vaccination.[27]

VAC52150EBL2003 is a phase 2 study aiming to assess the safety, tolerability and immunogenicity of different vaccination schedules of Ad26.ZEBOV and MVA-BN-Filo as heterologous prime-boost (15, 29 days after priming) regimens in healthy and in HIV-infected adults. The study was completed in December 2018 (available in clinicaltrial.gov, NCT02598388).

In VAC52150EBL2001 423 healthy adult participants were randomized in a blinded placebo-controlled, parallel-group, multicenter phase 2 study. Prime vaccination with Ad26.ZEBOV was followed by a MVA-BN-Filo boost at 29, 57 or 85 days and 6 months after prime vaccination follow up. After unblinding, only participants who received Ad26.ZEBOV and/or MVA-BN-Filo continued the study until 1 year follow up to assess long-term safety and immunogenicity. The enrolment and follow up of participants was completed in January 2019. Recently published results show that vaccinations were generally well tolerated. Mild or moderate local adverse events (mostly pain) were reported after 206 (62%) of 332 Ad26.ZEBOV vaccinations, 136 (58%) of 236 MVA-BN-Filo vaccinations, and 11 (15%) of 72 placebo injections. Systemic adverse events were reported after 255 (77%) Ad26.ZEBOV vaccinations, 116 (49%) MVA-BN-Filo vaccinations, and 33 (46%) placebo injections, and included mostly mild or moderate fatigue, headache, or myalgia. Unsolicited adverse events occurred after 115 (35%) of 332 Ad26.ZEBOV vaccinations, 81 (34%) of 236 MVA-BN-Filo vaccinations, and 24 (33%) of 72 placebo injections. At 21 days after receiving the MVA-BN-Filo vaccine, geometric mean concentrations of Ebola virus glycoprotein-binding antibodies were 4627 ELISA units (EU)/mL (95% CI 3649–5867) in the 28-day interval group, 10 131 EU/mL (8554–11 999) in the 56-day interval group, and 11 312 EU/mL (9072–14106) in the 84-day interval group, with antibody concentrations persisting at 1149–1205 EU/mL up to day 365.[28]

The study DRC-EB-001 investigates population-level vaccination with the Ad26.ZEBOV, MVA-BN-Filo vaccine. This study was launched during the DRC north Kivu outbreak, and vaccination was offered to communities that neighbored the outbreak area or that were located on transport routes from the edge of the outbreak area to major centers like Goma. The vaccine was given to adults, children and pregnant women.

***Phase 2/3 studies including children***

Ad26.ZEBOV and MVA-BN-Filo are being evaluated as a prime-boost vaccination strategy in children in a phase 2 study (VAC52150EBL2002) and in a phase 3 study (VAC52150EBL3001).

VAC52150EBL2002 is a randomized, observer-blind, placebo-controlled, parallel-group, multicenter, Phase 2 study evaluating the safety, tolerability and immunogenicity of 3 heterologous prime-boost regimens using Ad26.ZEBOV as prime and MVA-BN-Filo as boost vaccination. MVA-BN-Filo boost vaccination was administered at 28-, 56- and 84-day intervals, in healthy and elderly participants and at 28- and 56-day intervals in HIV-infected participants and in children. This study was conducted in Africa and the enrolment took place sequentially in four cohorts: the first cohort consisted of healthy participants (18 - 70 years); the second cohort (2a) included HIV-infected participants (18 to 50 years) and healthy children 12 to 17 years (cohort 2b); the third cohort included children aged 6 to 11 years inclusive and in the fourth cohort young children aged 1 to 5 years were enrolled. Within each cohort, participants were randomized in a 5:1 ratio to receive active vaccine versus placebo. Upon completion of the 42-day post-boost visit those participants who received active vaccine entered long-term follow-up, with visits on days 180 and 365 post-prime vaccinations to assess long-term safety and immunogenicity. Primary outcome measures are the number of participants with AEs up to 42 days post-boost, the number of participants with SAEs in the year following vaccination and the number of participants with solicited local and systemic AEs in the 7 days following prime and boost vaccination. Secondary outcomes include GP-EBOV antibody response and T Cells function. A total of 1075 participants have been enrolled and the study was completed in February 2019

VAC52150EBL2005 is a Phase 2 study to evaluate the safety, reactogenicity, and immunogenicity of a heterologous 2-dose vaccination regimen using Ad26.ZEBOV and MVA-BN-Filo on days 1 and 57 in infants aged 4-11 months in Guinea and Sierra Leone. To date, recruitment and follow up have ended and data analysis is ongoing

VAC52150EBL3001 is a staged study including a double-blinded controlled phase to evaluate the safety and immunogenicity of Ad26.ZEBOV and MVA-BN-Filo. The study takes place in Sierra Leone and consists of a screening phase, an active phase (vaccination) and a follow-up phase. The participants were randomized to Ad26.ZEBOV as prime and MVA-BN-Filo as boost vaccination at day 57 or control vaccination as prime and placebo as boost vaccination at day 57. The control used is MENVEO® World Health Organization (WHO)-prequalified Meningococcal Group A, C, W135 and Y conjugate vaccine. The active phase of the study was conducted in two stages. The first stage is open-label and approximately 40 adults aged 18 years or older were vaccinated with Ad26.ZEBOV as prime and MVA-BN-Filo as boost to gain information about the safety and immunogenicity of the prime-boost regimen. In stage 2 a larger group of approximately 976 individuals were randomized to the Ad26.ZEBOV/MVA-BN-Filo vaccines or MENVEO® vaccines to further evaluate the safety and immunogenicity of the prime-boost regimen across different age groups. In this stage, children aged 1 year or older, adolescents and adults have been included. Safety data was collected in stage 1 and 2, seven days after the initial vaccination and boost vaccination. Safety evaluations included assessment of AEs, which will be monitored throughout the study. For stages 1 and 2, the follow-up was 360 days after prime vaccination. In total 1023 participants have been enrolled and the estimated study completion date is August 2019.

A new study has been launched in 2021 aiming to assess a booster in children previously vaccinated with Ad26.ZEBOV and MVA-BN-Filo (VAC52150EBL2011).

***Studies including pregnant woman***

A Phase 3 study using Ad26.ZEBOV followed by MVA-BN-Filo in healthy pregnant women (INGABO) is currently ongoing in Rwanda. The purpose of this study is to assess adverse maternal/fetal outcomes in pregnant women and adverse neonatal/infant outcomes in neonates/infants born to vaccinated women.

An extension of this trial (VAC52150EBL3005) started in 2019 and aims to evaluate long-term safety and immunogenicity of the candidate Ebola vaccines Ad26.ZEBOV and MVA-BN®-Filo in participants who were exposed to these vaccines in the VAC52150EBL3001 trial.

Safety data generated with the 2 vaccines are provided below:

##### *Safety Profile of Ad26.ZEBOV*

Ad26.ZEBOV is a monovalent recombinant, replication-incompetent Ad26-based vaccine. Only limited clinical data are available for Ad26.ZEBOV. Thrombosis with thrombocytopenia syndrome has been very rarely reported with the Ad26.COV2.S vaccine (see clinical safety below). Adenovirus vaccine programs with other gene inserts have revealed no other significant safety issues.[23,24]

##### *Safety Profile of MVA-BN*

MVA-BN is a further attenuated version of the MVA virus, which in itself is a highly attenuated strain of the poxvirus Chorioallantois Vaccinia Virus Ankara. MVA-BN induces strong cellular activity as well as a humoral (antibody) immune response and has demonstrated an ability to stimulate a response even in individuals with pre-existing immunity against Vaccinia. One of the advantages of MVA-BN is the virus' inability to replicate in a vaccinated individual. The replication cycle is blocked at a very late stage, which ensures that new viruses are not generated and released. This means that the virus cannot spread in the vaccinated person and none of the serious side effects normally associated with replicating Vaccinia viruses have been seen with MVA-BN.

MVA-BN (MVA-BN®, trade name IMVAMUNE® outside the European Union [EU], invented name IMVANEX® in the EU) has received marketing authorization in the EU for active immunization against smallpox in adults, and in Canada for adults who have a contraindication to the first or second generation smallpox vaccines including people with immune deficiencies and skin disorders**.** A Phase 3 clinical study has been performed in the United States (US) (POX-MVA-013). Results of completed and ongoing clinical studies of MVA-BN-based vaccines in more than 8,100 individuals, including elderly, children and immunocompromised subjects in whom replicating vaccines are contraindicated, have shown that the platform displays high immunogenicity and a favorable safety profile.[24] Across all clinical studies, no trends for unexpected or SAEs due to the product were detected. Nonclinical studies in macaques with different levels of immune suppression support the safety profile of the MVA-BN strain.[29] Biosafety aspects of MVA-BN strains used for vaccinations have been reviewed.[30]

<u>Relevant Safety Information</u>

One subject in the study VAC52150EBL2001 experienced a serious and very rare condition called “Miller Fisher syndrome”. This condition consists of double vision, pain on moving the eye, and difficulty with balance while walking. Miller Fisher syndrome most commonly occurs following a recent infection. The subject experienced these symptoms about a week after suffering from a common cold and fever. The event happened about a month after boost vaccination with either MVA-BN-Filo or placebo. This subject had to go to the hospital for treatment and has recovered. After an extensive investigation, the event has been considered to be doubtfully related to vaccine and most likely related to the previous common cold.

In addition, there have been a few reports of mild transient paresthesia, especially in the hands and feet, or a sensation of mild muscle weakness of unknown cause in subjects vaccinated with Ad26.ZEBOV or placebo. These symptoms have been observed to last no more than 24-48 hours in the majority of cases but can last for several weeks to several months before going away on their own. One serious case of peripheral sensory neuropathy (nerve damage that may cause tingling, numbness, pain or loss of pain sensation) of moderate severity has occurred and has been ongoing for several months, interfering with some of the participant’s daily activities. These types of symptoms have also been reported following administration of other licensed vaccines and following acute viral infections of various types.

Few cases of convulsions/seizures (with or without observed fever) were reported following post-approval use of Ad26.ZEBOV in young children, for which a causal relation to vaccination is plausible. The frequency of (febrile) seizure with Ad26.ZEBOV in pediatrics (1 to 17 years of age) is rare. The use of prophylactic antipyretic medicinal products soon after vaccination is recommended for children with seizure disorders or with a prior history of febrile seizures. Antipyretic treatment should be initiated according to local treatment guidelines.

Clinical Safety Experience With Ad26-based Vaccines

Thrombosis in combination with thrombocytopenia (thrombosis with thrombocytopenia syndrome [TTS]), in some cases accompanied by internal bleeding, has been observed very rarely following vaccination with the Janssen COVID-19 (Ad26.COV2.S) vaccine. Reports include severe cases of venous thrombosis at unusual sites such as cerebral venous sinus thrombosis (CVST), splanchnic vein thrombosis and arterial thrombosis, in combination with thrombocytopenia. The associated symptoms began approximately 1 to 2 weeks after vaccination, mostly in women under 60 years of age. Thrombosis in combination with thrombocytopenia can be fatal. The exact pathophysiology of TTS is unclear. This event has not been observed to date with any other Janssen Ad26-based vaccines. Participants should be instructed to seek immediate medical attention if they develop symptoms such as shortness of breath, chest pain, leg swelling, persistent abdominal pain, severe or persistent headaches, blurred vision, skin bruising or petechiae beyond the site of vaccination.

##### 1.1.3. Merck VSV-based vaccine

The VSV-based vaccine was originally developed by the Public Health Agency of Canada (PHAC) and advanced in development with the support of a number of partners including the US Department of Health and Human Services and the Department of Defense.

The rVSVΔG-ZEBOV-GP vaccine is comprised of a single recombinant replicating VSV isolate (11481 nt) modified to replace the gene encoding the G envelope GP with the gene encoding the envelope GP from ZEBOV (Kikwit, 1995 strain).

A brief summary of the vaccine, non-clinical, and clinical safety, immunogenicity, and efficacy data is presented. Detailed information may be found in the Investigator's Brochure.[31]

The rVSVΔG-ZEBOV-GP vaccine was licensed as ERVEBO® by the European Medicines Authority (EMA) and the United States Food and Drug Administration (FDA) in December 2019. ERVEBO® (rVSVΔG-ZEBOV-GP) is a vaccine indicated for the prevention of disease caused by Zaire ebolavirus in individuals 18 years of age and older.

**Non-clinical studies**

***Immunogenicity and Efficacy***

The initial studies of the rVSVΔG-ZEBOV-GP vaccine demonstrated 100% protection against lethal ZEBOV challenge in 4 cynomolgus macaques following a single intramuscular immunization of $1 \times 10^7$ pfu.[32] At the same dose, this vaccine was also able to protect cynomolgus macaques from homologous aerosol challenge.[33] Protection from aerosol challenge after IM vaccination has not been replicated in more recent studies in which vaccinated animals experienced pulmonary inflammation with significant lung pathology; the reasons for this difference are being studied further.

Delivering $2 \times 10^7$ pfu of laboratory-grade vaccine by the oral (n=4), intranasal (n=40), or IM (n=2) route 28 days prior to challenge protected cynomolgus macaques from subsequent ZEBOV challenge.[34] Interestingly, the level of neutralizing antibodies was low by all routes of immunization, highlighting the uncertainty regarding immunologic correlates of protection to Ebola virus infection.

rVSVΔG-ZEBOV-GP vaccine also protected 4 of 8 (50%) rhesus macaques when administered 30 minutes following challenge.[35] While all macaques developed fever and lymphopenia, 50% of the animals did not progress to more severe disease. VSV viremia was detected on day 3 post-immunization in most animals. ZEBOV viremia in survivors was 2 to 4 logs lower than in non-survivors. Of note, animals treated with rVSVΔG-ZEBOV-GP did not show a decrease in natural killer (NK) cells following challenge but instead showed a substantial increase.

The rVSVΔG-ZEBOV-GP vaccine has undergone challenge testing in murine and non-human primate models of pre- and post-exposure prophylaxis. In the murine model, the pre-exposure prophylaxis study include mice that were immunized with 20,000, 200, 2 and 0.02 pfu (n=10 per group). The mice were challenged 28 days after immunization with 1000 median lethal dose ($LD_{50}$) of mouse-adapted ZEBOV. All mice immunized with 200 or 20,000 pfu survived. In the post-exposure prophylaxis study, mice were challenged with 1000 $LD_{50}$ of mouse-adapted ZEBOV and then treated 30 minutes and 2 hours later with graded doses of $2x10^5$, $2x10^4$, $2x10^3$ and $2x10^2$ pfu (10 per group). When treated 2 hours after the challenge, doses of $2x10^5$, $2x10^4$, and $2x10^3$ were 80-100% effective in preventing death, Treatment within 30 minutes was not effective, with the majority of mice dying at each dose.

In cynomolgus macaques, one pre-exposure prophylaxis study included 4 animals who were inoculated IM with $1x10^8$ pfu and 2 controls. All animals were challenged IM 28 days later. The two controls died 6 days after challenge and all 4 vaccinated animals survived.

A second study included 8 cynomolgus macaques who were inoculated IM with $3x10^6$ pfu, $2x10^7$ pfu, and $1x10^8$ pfu each as well as 3 controls. The animals were challenged IM 42 days later. 0/3 of the animals that received placebo, 7/8 of animals that received $3x10^6$ pfu, 8/8 animals that received $2x10^7$ pfu and 8/8 animals that received $1x10^8$ pfu vaccination survived.

A third study included 4-5 cynomolgus macaques who were inoculated IM with $3x10^2$ pfu, $3x10^3$ pfu, $3x10^4$ pfu, $3x10^5$ pfu, and $3x10^6$ pfu each as well as 2 controls. The animals were challenged IM 42 days later. 0/2 of the animals that received placebo and all animals that received vaccination survived.

A durability study in cynomolgus macaques is in progress to assess immunogenicity and protection from challenge at 42 days, three months, and one year following IM vaccination with $3x10^4$ pfu (2 animals in the one year group) or $3x10^6$ pfu (the rest of the animals). Preliminary results indicate that 0/2 of the animals that received placebo, 4/4 animals challenged 42 days after vaccination, 2/6 animals challenged 3 months after vaccination, and 3/7 animals challenged one year after vaccination survived. Additional studies are being planned to assess vaccine dose, boost, and origin of cynomolgus macaques.

In the post-exposure prophylaxis study 12 rhesus macaques were challenged with 3000 $TCID_{50}$ of ZEBOV Kikwit. At 30 minutes or 24 hours after challenge, groups of 6 macaques were vaccinated with $2x10^8$ pfu of clinical grade rVSVΔG-ZEBOV-GP. Two of 6 (33%) treated 30 minutes after challenge survived but none of the animals treated at 24 hours survived. Two controls died within 10 days. [31]

The kinetics of rVSVΔG-ZEBOV-GP vaccine replication *in vitro* suggest it replicates substantially faster than wild-type ZEBOV and interference with ZEBOV replication via competition for receptor and/or for intracellular resources is a potential mechanism for its effects as a strategy for post-exposure prophylaxis.[36] In contrast to control animals, which undergo a reduction in lymphocytes during infection, vaccinated animals either showed a stable or small increase in CD3+, CD4+, and CD8+ T cells.

##### *Toxicology*

Repeated dose toxicology studies have been performed to evaluate the toxicity and local tolerance of VSVΔG-ZEBOV-GP in mice and monkeys. In these studies, VSVΔG-ZEBOV-GP was administered via IM injection on Study Days 1 and 14 followed by a 30-day observation period. BALB/c (Bagg albino) mice and cynomolgus monkeys received VSVΔG-ZEBOV-GP at up to $2.0\times10^7$ pfu/animal and $1.0\times10^8$ pfu/animal, respectively. There was no evidence of systemic toxicity based on clinical pathology and necropsy data. Developmental and reproductive toxicology studies are ongoing.[31] The nonclinical safety of rVSV-HIV vaccines has also been assessed in non-Good Laboratory Practice (GLP) NHP studies, indicating that rVSV is highly attenuated and supports the continued use of rVSVΔG-ZEBOV-GP in clinical trials.[37] Particularly noteworthy was the absence of neurovirulence in NHP following intrathalamic inoculation.[38]

##### Clinical Studies

Due to the public health emergency of the 2014-2016 West African outbreak, rVSV∆G-ZEBOV-GP vaccine was administered to over 16,000 adults in eight phase 1 and four phase 2-3 clinical trials.[39] The “Ebola Ça Suffit” open label cluster randomized vaccination trial in Guinea evaluated vaccine effectiveness in case contacts, where clusters of contacts of Ebola cases were randomised for immediate or delayed vaccination with rVSV-ZEBOV. Vaccine efficacy was estimated to be 100% (95% CI 68·9–100, p=0·0045) 10 days or more after randomisation in individuals vaccinated in the immediate group compared with those eligible and randomised to the delayed group.[49]

Additional safety and immunogenicity data on rVSV-ZEBOV have been generated in the Sierra Leone Trial to Introduce a Vaccine Against Ebola (STRIVE) study, which enrolled more than 8000 health-care and frontline workers in Sierra Leone, and in a trial in Guinea (the Front Line Worker trial). There have also been eight phase 1 trials and a phase 3 safety and manufacturing-consistency trial of the rVSV-ZEBOV vaccine conducted in North America, Europe, and Africa. Collectively, trial data indicate that the rVSV-ZEBOV vaccine has an acceptable safety profile, and that it induces immunity that is durable for at least 24 months in adults (antibody responses for 89–100% of recipients depending on the doses administered).[40,42]

Among other studies of this vaccine, the Partnership for Research on Ebola Vaccines in Liberia I (PREVAIL I) randomised, double-blind, placebo-controlled trial assessed the safety and immunogenicity of the rVSV-ZEBOV vaccine and another candidate vaccine in 1500 adults, showed that adverse effects were generally mild and time-limited and satisfied safety and immunogenicity profiles.[52]

The ACHIV (study African-Canadian Study of HIV-Infected Adults and a Vaccine for Ebola) trial is a randomized placebo-controlled trial evaluating the safety and immunogenicity of rVSVΔG-ZEBOV-GP in HIV-positive adults and adolescents and includes the evaluation of one and two doses. This trial is being conducted in Burkina Faso, Canada, and Senegal with ongoing enrolment. Given that Ebola outbreaks may overlap with areas of high HIV prevalence, understanding the safety and immunogenicity of rVSVΔG-ZEBOV-GP in these populations is very important.

PREPARE (Pre-Exposure Prophylaxis in People at Potential Occupation Risk for Ebola Virus Exposure) is a randomized controlled trial being conducted in the US and Canada to evaluate the durability of immune responses to rVSVΔG-ZEBOV-GP in individuals who are at occupational risk of exposure to Ebola (e.g., through working in BSL-4 laboratories, etc.) and includes randomization to assess the impact of a booster dose given at 18 months.

##### *Safety Profile of rVSVΔG-ZEBOV-GP*

Preliminary safety data collected to date have demonstrated rVSVΔG-ZEBOV-GP to be generally well-tolerated when administered to healthy adults. Injection-site reactions following vaccination are generally mild or moderate and self-limited. Thereafter, there is a predictable period of early reactogenicity, including fever and a flu-like syndrome, which is common after receipt of other live-virus vaccines, and resolves within 7 days. During this time, transient decreases in white blood cells and platelets have also been observed but bleeding or increased risk of infection has not been reported. The early reactogenicity is likely to be a result of viremia with the vaccine virus which has been reported in the majority of subjects and resolves within 2 weeks. Mild and self-limited oral ulcers have been observed in a few subjects. No rVSV has been detected in oral ulcers to date. Vaccine virus has been found in saliva, urine, and skin-vesicles from those vaccinated. The largest proportion of adult subjects that had vaccine virus detected in saliva or urine was 5 of 20 in $2x10^7$ pfu recipients with positive saliva on Day 7 and this decreased to 1 of 10 on Day 14. Person-to-person transmission has not been documented.

Joint pain (arthralgia) has been reported in the first week after vaccination in up to 35% of vaccinees (range 5 to 35%, blinded data). In a small proportion of subjects (<5% at most study sites), joint swelling (arthritis) has been observed in the 2nd to 3rd week after vaccination.[31]

In one Phase 1 clinical trial in which 25 of the 51 subjects that received the study vaccine were vaccinated in an open-labeled manner because they were deployable to the Ebola outbreak, the proportion of subjects with arthritis was higher (11 of 51; 22%) for reasons that have not been fully elucidated. The arthritis is thought to be virally-mediated, and a vast majority of cases have resolved within several weeks.[41,42] However, several subjects have reported recurrent or persistent arthralgia and arthritis up to 6 months of vaccination, the longest time followed to date.

Arthritis was also reported in a second study being conducted at the NIH in the US. In this open-label study of healthcare workers called the Multicenter Study of the Immunogenicity of Recombinant Vesicular Stomatitis Vaccine for Ebola-Zaire (rVSVΔG-ZEBOV-GP) for Pre-Exposure Prophylaxis in Individuals at Potential

Occupational Risk for Ebola Virus Exposure (PREPARE), among 9 participants enrolled, 3 developed signs and symptoms of arthritis in the knee 10-14 days following vaccination. Rheumatologic evaluation (including imaging and, in one case, diagnostic arthrocentesis) of these 3 individuals supported the diagnosis of what was deemed likely to be arthritis, and which took 3-4 months to resolve to baseline. Of note, while the complaint of arthralgia was commonly reported in an earlier phase 1 study conducted jointly by NIH and the Walter Reed Army Institute of Research,[43] arthritis had not been seen (although observed in other phase I/II trials conducted elsewhere)[41]. Also, 6 of the 9 participants reported moderate to severe pain in the arm during the injection of the vaccination (R. Davey, personal communication, April 7, 2017), a complaint that had also not been observed in the earlier phase 1 experience.

In a Phase 1b dose response randomized trial of the rVSVΔG-ZEBOV-GP vaccine with doses ranging from $3 \times 10^3$ to $1 \times 10^8$ pfu/mL given to adults in the U.S., there was no difference in the percent with post-vaccination arthritis among those vaccinated with one of the doses of rVSVΔG-ZEBOV-GP (19/418; 4.5%) compared to placebo (3/94; 3.2%) (p=0.78). The percent with arthritis among those given the $2 \times 10^7$ dose of rVSVΔG-ZEBOV-GP was 4.3% (2/47); for those given the highest dose of rVSVΔG-ZEBOV-GP studied, $1 \times 10^8$ pfu/mL, 1 of 48 (2.1%) had arthritis post-vaccination.[44]

In a double-blind placebo controlled multicenter Phase 3 lot consistency clinical trial[45] of adults in the US, Canada and Spain, the incidence of arthritis within 42 days post-vaccination was statistically significantly higher in 40/790 (5.1%) vaccine recipients who received a dose of $2 \times 10^7$ pfu/mL when compared to 0/133 (0%) placebo recipients (p-value=0.008). Among 260 participants given a higher dose of rVSVΔG-ZEBOV-GP ($1 \times 10^8$ pfu/mL), 11 (4.2%) developed arthritis, a percentage similar to those given $2 \times 10^7$ pfu/mL). The cases of arthritis in this clinical trial were mostly mild to moderate in intensity and generally resolved within a week, with some participants experiencing longer duration.

Rash (with and without vesicles) has been observed in a few subjects mostly in the second week after vaccination, and vaccine virus has been detected in several subjects in whom vesicular fluid or skin biopsy specimens were collected. The reported skin lesions also appear to be consistent with a virally-mediated process. The safety profile of rVSVΔG-ZEBOV-GP in preliminary reports from the Ebola epidemic region has been generally similar to that observed outside the region.[31]

##### *Data from children, HIV-positive subjects, and pregnant women*

To date, there have been a small number of patients less than 18 years of age or HIV-positive individuals exposed to the rVSVΔG-ZEBOV-GP vaccine in ongoing trials in Africa. Preliminary open-label data in 20 adolescents between 13 and 17 years of age and 20 children between 6 and 12 years of age in Gabon suggests a similar reactogenicity profile to the reactogenicity profile observed in adult subjects. Viremia and shedding after vaccination appear to be higher in children and adolescents compared to adults. Peak viremia occurs on Day 2 and decreases by Day 7. Median copies/mL detected by reverse transcriptase polymerase chain reaction (RT-PCR) in $2x10^7$ pfu recipients are 530 in adults, 1,592 in adolescents between 13 and 17 years, 1,109 in children between 6 and 12 years of age (Kruskal Wallis test P<0.05). The largest proportion of pediatric subjects that had vaccine virus detected in saliva or urine was 14 of 18 adolescents with positive saliva on Day 7 (shedding on Day 14 was not assessed in this study).

Preliminary unblinded data from one Phase 2 trial suggests that approximately 5% (22 subjects out of 500 total) of the volunteers that were vaccinated with rVSVΔG-ZEBOV-GP were HIV-positive and the safety profile appeared generally similar to HIV-negative participants, although the small numbers limit the conclusions that can be drawn at this time.

Pregnant women were excluded from the vaccine trials to date. However, a small number of women have been found to be pregnant after vaccination and are being followed for the outcome of their pregnancies. Of note, a recent systematic review evaluating the risk-benefit balance of the rVSV-ΔG-ZEBOV-GP vaccine showed that data from ring efficacy trials yielded a good risk-benefit balance of the vaccine not only in adults, but also in children and pregnant and lactating women and HIV-infected people.[46]

##### *Immunogenicity of rVSVΔG-ZEBOV-GP*

Anti-GP antibodies are detectable by enzyme-linked immunosorbent assay (ELISA) at 14 days post vaccination in a majority of vaccine recipients and to date, approximately 96% seroconversion has been observed by 28 days post-vaccination. Persistence of EBOV-GP-specific antibody responses is strong at least 2 years after vaccination..[47] As the ELISA and neutralization assays are validated, more data will be emerging from the larger, Phase 3 studies in the clinical development program.

##### *Efficacy of rVSVΔG-ZEBOV-GP*

An open-label, cluster randomized ring vaccination Phase 3 trial was conducted in Guinea in which clusters of contacts and contacts of contacts of confirmed cases of Ebola were identified and randomly assigned to receive a single $2x10^7$ pfu dose of rVSVΔG-ZEBOV-GP immediately or after 21 days. Efficacy and effectiveness have been assessed in an interim analysis and the results were published by Henao-Restrepo and colleagues.[48] In the publication of the interim analysis, 7,651 subjects were included in the planned interim analysis which included 48 clusters (4,123 subjects) randomly assigned to immediate vaccination and 42 clusters (3,528 subjects) randomly assigned to delayed vaccination. In the immediate vaccination group, there were no cases of EVD with symptom onset at least 10 days after randomization, whereas in the delayed vaccination group there were 16 cases of EVD from seven clusters reported, demonstrating a vaccine efficacy of 100% (95% confidence interval [CI]: 74.7, 100.0; p=0.0036). At the cluster level, with the inclusion of all eligible adults, vaccine effectiveness was 75.1% (95% CI: 7.1, 94.2; p=0.1791). No new cases of EVD were diagnosed in vaccinees from the immediate or delayed groups from 6 days post-vaccination. Final analyses involving 9,096 participants in 98 randomized clusters have been published. Similar to the interim analysis results, no cases of EVD with symptom onset at least 10 days after randomization occurred among those randomized to immediate vaccination, whereas in the delayed vaccination group there were 16 cases of EVD from seven clusters reported, demonstrating a vaccine efficacy of 100% (95% CI: 68.9, 100.0; p=0.0045).[49]

The rVSVΔG-ZEBOV-GP vaccine has been used during the last Ebola outbreaks in DRC and Guinea under a compassionate use protocol using ring vaccination first (Equateur and North Kivu and Ituri outbreaks), and once authorized, as a prophylaxis for vaccinating subjects at risk with more than 300 000 people vaccinated. A study was conducted during the compassionate use protocol in Ituri and North Kivu by the Institut National pour la Recherche Biomedicale (INRB) and WHO on data collected from a ring vaccination cohort protocol confirms the high efficacy of rVSV-ZEBOV-GP Ebola vaccine against disease. Seventy-one cases of Ebola occurred among the 93,965 people at risk vaccinated in 679 rings. Conversely, during the same period, 880 cases occurred among individuals at risk of Ebola who were not vaccinated. Of the, 71 Ebola cases occurring among vaccinated individuals, 15 cases had onset of symptoms 10 days or more days post- vaccination (this is the time after which vaccinees are assumed to be protected) and 57 cases had onset of symptoms 0-9 days post vaccination (it is assumed that during this period the vaccinees are not protected or partially protected). No deaths were reported among vaccinees who developed Ebola with onset 10 or more days after vaccination and the overall case fatality rate was reduced among all vaccinees who developed Ebola.[50]

##### 1.1.4. Placebo

The placebo vials were sterile normal saline (sodium chloride).

#### 1.2. Rationale

As described above, in an open-label, cluster randomized ring vaccination trial of adults conducted in Guinea, the Merck rVSVΔG-ZEBOV-GP vaccine prevented EVD events diagnosed 10 days or more after randomization among close contacts immediately vaccinated as compared to those eligible to be vaccinated 21 days later. In that trial many EVD events occurred within 10 days of randomization. Immunogenicity testing was not performed. The rVSVΔG-ZEBOV-GP vaccine was licensed as ERVEBO® by the EMA and the FDA in December 2019.

The J&J prime boost vaccine regimen, rHAd26/MVA, has completed phase 1 testing in adults and is currently being evaluated in phase 2 and 3 clinical trials.

Ad26.ZEBOV/MVA-BN-Filo vaccines were licensed by EMA on May 2020 under the commercial names of Zabdeno® and Mvabea® corresponding to the two components. Zabdeno® is given first and Mvabea® is administered approximately eight weeks later as a booster.

Multiple questions regarding the safety and effectiveness of EVD vaccines remain unanswered, including the immediacy of immune responses generated by the Ad26.ZEBOV and rVSVΔG-ZEBOV-GP vaccines, the durability of the immune response following a booster at 56 days (MVA-BN-Filo following Ad26.ZEBOV prime and repeat rVSVΔG-ZEBOV-GP following rVSVΔG-ZEBOV-GP prime), and the safety of the different vaccine strategies.

During the EVD epidemic in West Africa a considerable proportion of patients were children and the mortality rate in those aged < 5 years was very high: 89.5% (CI 95%: 75.9, 95.8).[51]

There is a need for additional effective vaccines for adults and children as the risk of re-emergence of EVD is real, as shown by the recent outbreaks in DRC and Guinea. Although many studies completed or ongoing have shed light on questions regarding vaccine safety and immunogenicity, there are a number of unknowns regarding vaccination against Ebola virus. For example, additional data in children, the durability and rapidity of immune responses or the correlation between immune response and clinical protection remain important areas of investigation.

Thus, this trial is addressing questions related to safety and immunogenicity and, long term protection in adults and children under three vaccine strategies

The trial includes a placebo group and is powered to compare each vaccine strategy with placebo separately for adults and children for the antibody response at 12 months. Consideration was given to using an active vaccine as the control in children but in order to provide the best safety data of each vaccine strategy, a “true” placebo (saline) is used as the comparator.

The trial is double-blind until 12 months of follow-up is completed for all participants. After the clinical and safety database lock for the M12 data, the study will be unblinded. Randomized participants will not know their randomized assignment until the M12 study results are available. The exception to this is if an Ebola outbreak occurs in one of the participating countries, in which case participants will be informed of their randomized assignment after their M12 visit but prior to the M12 study results being available. Clinical staff carrying out safety evaluations are blinded to the participants’ randomized assignment. Laboratory staff carrying out immunogenicity testing and biochemical testing are also blinded to the vaccine/placebo that the participant received. To accomplish the blinding in both adults and children, participants randomized to the rVSVΔG-ZEBOV-GP vaccine without the boost received placebo at 56 days, while participants randomized to rHAd26/MVA received the MVA-BN-Filo boost at 56 days, and those randomized to the rVSVΔG-ZEBOV-GP vaccine with a boost received a repeat rVSVΔG-ZEBOV-GP vaccination at 56 days. A placebo with two fill volumes, 0.5 mL and 1.0 mL, was used to blind the prime vaccination.

During the long-term follow-up, post M12 visit, and following the current outbreak in Guinea and vaccine licensures, the participants who received the placebo will be vaccinated with a single dose of rVSVΔG-ZEBOV-GP vaccine in Liberia and

Mali and with the Ad26.ZEBOV/MVA-BN-Filo vaccine strategy in Guinea and Sierra Leone during their next long-term follow-up visit, or during additional visits if their next scheduled long-term follow-up visit not approaching soon. The participants who received an incomplete Ad26.ZEBOV/MVA-BN-Filo vaccine strategy will be offered a single dose of the Ad26.ZEBOV or MVA-BN-Filo vaccine in Guinea and Sierra Leone and a single dose of rVSVΔG-ZEBOV-GP in Liberia and Mali. The participants who have been offered active vaccine will continue their follow-up until M60 visit following their enrolment in PREVAC. Safety data will continue to be collected for this newly vaccinated group.

### 2. METHODOLOGY

#### 2.1. Study design

After obtaining informed consent, participants were randomized to the following 5 groups in a 2:1:2:1:1 allocation 1) Ad26.ZEBOV (prime) (0.5 mL) followed by an MVA-BN-Filo boost (0.5 mL) at 56 days; 2) placebo (prime and boost at 56 days) (0.5 mL); 3) rVSVΔG-ZEBOV-GP (prime) (1 mL) followed by placebo boost (1 mL) at 56 days; 4) rVSVΔG-ZEBOV-GP (prime) (1 mL) followed by rVSVΔG-ZEBOV-GP boost (1 mL) at 56 days; and 5) placebo (prime and boost at 56 days) (1 mL) (Figure 1). At 56 days, participants assigned to the rVSVΔG-ZEBOV-GP vaccine without a boost and the two placebo groups received a placebo vaccination. Those initially given the Ad26.ZEBOV vaccine received the MVA-BN-Filo vaccine at 56 days. Those assigned to the boosted rVSVΔG-ZEBOV-GP arm received a booster vaccination with the rVSVΔG-ZEBOV-GP vaccine at 56 days.

In each of the groups receiving the rVSVΔG-ZEBOV-GP vaccine (prime or boost), the undiluted dose of the rVSVΔG-ZEBOV-GP vaccine was used.

The trial is conducted at sites in Guinea, Liberia, Mali, and Sierra Leone.

In the event of a new Ebola epidemic in these countries or prospective risk of Ebola to study participants from Ebola, the study design, including the vaccination of subjects that received placebo with Ebola vaccine, will be re-considered. Under protocol version 6.0 and version 7.0, and following the new Ebola outbreak declared in Guinea in February 2021 and the vaccine licensures, the participants who received the placebo will be vaccinated with a single dose of rVSVΔG-ZEBOV-GP vaccine in Liberia and Mali and with the Ad26.ZEBOV/MVA-BN-Filo vaccine strategy in Guinea and Sierra Leone during their next long-term follow-up visit, or during additional visits if their next scheduled long-term follow-up visit is not approaching soon. The participants who received an incomplete Ad26.ZEBOV/MVA-BN-Filo vaccine strategy will be offered a single dose of the Ad26.ZEBOV or MVA-BN-Filo vaccine in Guinea and Sierra Leone and a single dose of rVSVΔG-ZEBOV-GP in Liberia and Mali. Safety data will continue to be collected for this newly vaccinated group.

Figure 1: PREVAC Study Design (Version 4.0)

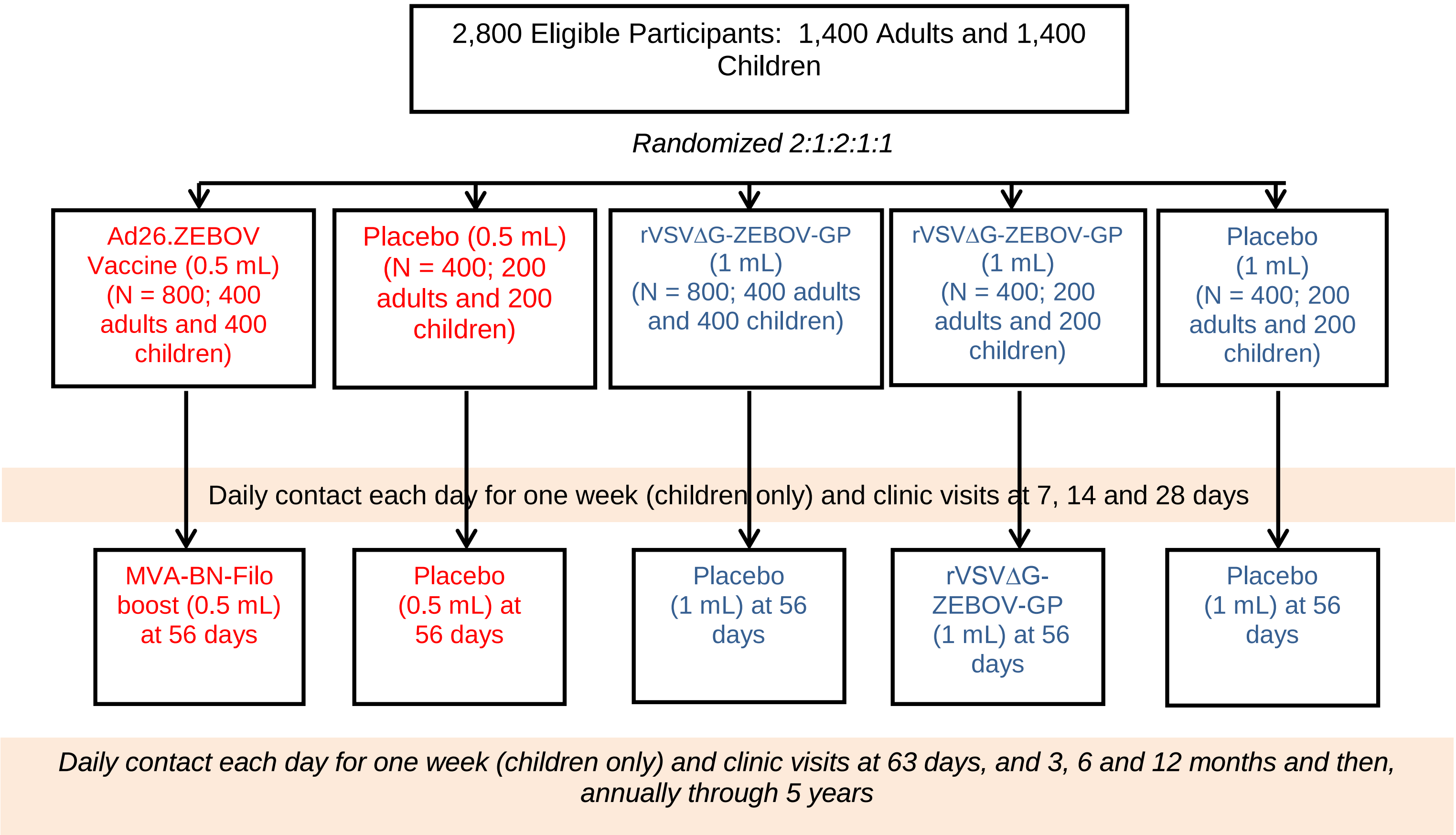

#### 2.2. Primary objectives

Separately for adults and children:

- To compare each of the three vaccine strategies with the pooled placebo group (3 pair-wise comparisons) for antibody response 12 months after randomization (prime vaccination).

In order to facilitate Merck regulatory filings and bridging of immune responses of the rVSVΔG-ZEBOV-GP vaccine from this study to other studies and between pediatric and adult populations, the following objective will be assessed specifically for those in the two rVSVΔG-ZEBOV-GP vaccine groups:

- To compare the rVSVΔG-ZEBOV-GP vaccine (pooled rVSVΔG-ZEBOV-GP groups) with the matched placebo group for antibody response 28 days after randomization (prime vaccination).

In order to facilitate Janssen regulatory filings of the rHAd26/MVA vaccine, the following objective will be assessed specifically for those in the rHAd26/MVA vaccine group:

- To compare the rHAd26/MVA vaccine group with the matched placebo group for antibody response 3 months after randomization (approximately 28 days after the booster vaccination).

#### 2.3. Primary hypotheses

The null hypothesis for each of the three comparisons at 12 months is that the antibody response for the vaccine strategy does not differ from the pooled placebo arm (0.5 mL and 1.0 mL groups)

The following hypothesis will only apply to the rVSV∆G-ZEBOV-GP vaccine for the purpose of facilitating Merck regulatory filings:

> The null hypothesis is that the antibody response for the pooled rVSVΔG-ZEBOV-GP group does not differ from the matched placebo group (0.5 mL and 1.0 mL groups) at Day 28 after randomization (initial vaccination).

The following hypothesis will only apply to the rHAd26/MVA vaccine for the purpose of facilitating Janssen regulatory filings:

> The null hypothesis is that the antibody responses for the rHAd26/MVA vaccine does not differ from the matched placebo group (0.5 mL and 1.0 mL groups) at Month 3 after randomization.

#### 2.4. Secondary objectives

The following secondary objectives will be assessed. Unless otherwise stated, the objectives will be addressed separately for adults and children:

- To compare the groups given the Ad26.ZEBOV prime vaccine and the rVSVΔG-ZEBOV-GP prime vaccine (both rVSVΔG-ZEBOV-GP groups combined) each with the pooled placebo group for the antibody response 14 days after randomization (prime vaccination) (immediacy of response).
- To compare each of the vaccine groups versus the pooled placebo group for the antibody response profile using measurements at 7, 14, 28, 56, 63 days and at 3, 6 and 12 months after randomization.
- To compare each of the vaccine groups with the pooled placebo group for SAEs at 12 months (see section 4.1.3).
- To compare the groups given the Ad26.ZEBOV prime vaccine and the rVSVΔG-ZEBOV-GP prime vaccine (both rVSVΔG-ZEBOV-GP groups combined) each with the pooled placebo group for the percent with injection site reactions and AEs graded for severity, including targeted symptoms, during the first week following randomization (including the daily contacts for children only).
- To compare the groups given the Ad26.ZEBOV prime vaccine and the rVSVΔG-ZEBOV-GP prime vaccine (both rVSVΔG-ZEBOV-GP groups combined) each with the pooled placebo group for percent reporting injection site reactions and AEs graded for severity, including targeted symptoms, following prime vaccination at the vaccination visit, and through 7, 14, and 28 days after the prime vaccination.

- To compare each of the vaccine groups with the pooled placebo group for changes from baseline in biochemical markers and complete blood count (CBC) measurements at 7 days after randomization (children only).
- To compare the rHAd26/MVA and rVSVΔG-ZEBOV-GP boost strategies with the pooled placebo group for changes from baseline in biochemical markers and CBC measurements at 63 days after randomization (children only).
- To compare the rHAd26/MVA and rVSVΔG-ZEBOV-GP boost strategies with the pooled placebo for percent with injection site reactions and AEs graded for severity, including targeted symptoms, immediately following the booster vaccination and through month 3 (approximately 35 days after the booster vaccination).
- To assess the long-term antibody response of three vaccine strategies at 24, 36, 48 and 60 months following randomization.
- To assess the long-term safety at month 24, 36, 48 and 60 following the three vaccine strategies.
- To compare antibody responses and safety outcomes of each of the vaccination strategies versus the pooled placebo group in subgroups defined by age, gender, country, whether the volunteer is a close contact of an Ebola case, the presence of laboratory abnormalities at baseline, and has specific co-morbidities (in particular HIV and nutritional status as measured by body mass index).
- For adults and children combined, to assess antibody responses and safety outcomes for each of the vaccination strategies.
- To carry out operational research which will include ethnographic, participatory and/or qualitative (i.e., focus groups and individual interviews) studies to: 1) identify issues relevant to understanding and acceptability of the trial, the social issues surrounding informed consent, with the primary goal of informing efforts to ensure autonomous fully informed individual consent and assent for minors; 2) describe participants' and caregivers' experience in the trial, and identify barriers and develop solutions to support trial adherence in a culturally sensitive and ethically appropriate way; and 3) understand prevailing representations and affects surrounding the epidemic (including rumors), Ebola and other vaccines, the trial and other relevant phenomena in order to ensure effective communication around the trial.
- In a subsample of adults in Guinea, to assess the T cell and memory B cell responses for the three vaccine strategies (see Appendix D).
- In a subsample of children, to compare the rVSVΔG-ZEBOV-GP vaccine strategies with the pooled placebo group for shedding of rVSV-ZEBOV-GP RNA (see Appendix E).

- To determine whether exposure to malaria and helminth infections influences the durability of the humoral and cellular immune responses to Ebola vaccines (see Appendix F).

- To identify multilevel facilitators and barriers to participant retention in PREVAC through retrospective qualitative research with participants after the conclusion of the blinded trial period (first 12 months) (see Appendix G).

- To identify factors that predict participant retention and loss-to-follow-up (LTFU) during the unblinded PREVAC long-term follow-up period (from 12 months up to 5 years) through prospective research (see Appendix G).

- To analyze the early vaccine signature, identify early correlates of durable antibody responses and propose in silico methods to optimally allocate vaccine strategies at the individual level.

#### 2.5. Exploratory objectives to be assessed only if and when the same correlate of protection has been established for both vaccine candidates

Separately for adults and children:

- To compare the three vaccine strategies with one another for the correlate of protection response at 28 days and 12, 24, 36, 48 and 60 months after randomization. For the comparison at 28 days, the two rVSVΔG-ZEBOV-GP groups will be combined.
- To compare the group randomly assigned to the Ad26.ZEBOV prime with the two groups (pooled) assigned the rVSVΔG-ZEBOV-GP prime vaccine for the correlate of protection response 14 days after randomization (immediacy of response).
- To compare the three vaccine strategies with one another for the correlate of protection response 3 months after randomization.

For pooled analysis of children and adults:

- To compare the three vaccine strategies with one another for the correlate of protection response 28 days and 12, 24, 36, 48 and 60 months after randomization (prime vaccination) (durability of response). For the comparison at 28 days the two rVSVΔG-ZEBOV-GP groups will be combined.
- To compare the Ad26.ZEBOV prime vaccination with the rVSVΔG-ZEBOV-GP prime vaccination (2 rVSVΔG-ZEBOV-GP groups combined) for the correlate of protection response 14 days after randomization (immediacy of response).
- To compare the group randomly assigned to the Ad26.ZEBOV prime with the two groups (pooled) assigned the rVSVΔG-ZEBOV-GP prime vaccine for the correlate of protection response 28 days after randomization.

- To compare the three vaccine strategies with one another for the correlate of protection response 3 months after randomization.
- To compare the three vaccine strategies with one another for percent of participants who experience SAEs through 60 months after randomization.
- To compare the three vaccine strategies with one another for the correlate of protection response and safety outcomes by age.

#### 2.6. Endpoints

The primary immunogenicity endpoint is the GP-EBOV antibody response 12 months after randomization. This endpoint will be used to compare the immunogenicity of the three vaccine strategies with placebo. The primary analysis for adults and children will be performed for participants who do not have elevated antibody levels at baseline.

GP-EBOV antibody response at Day 28 after randomization. For rVSVΔG-ZEBOV-GP arms only, Day 28 post first vaccination will be used for regulatory purposes for comparison to other studies and for bridging children to adults. The primary analysis for adults and children will be performed for participants who do not have elevated antibody levels at baseline.

GP-EBOV antibody response at Month 3 after randomization (approximately 28 days after the booster vaccination). For the rHAd26/MVA vaccine arm only, Month 3 will be used for regulatory purposes. The primary analysis for adults and children will be performed for participants who do not have elevated antibody levels at baseline.

Antibodies to the Ebola virus GP will be measured with the Filovirus Animal Nonclinical Group (FANG) ELISA assay if available. Other assays may also be used. If a correlate of protection is identified, stored sera will be used to measure the correlate and carry out comparisons of the three vaccine strategies with placebo and with one another. The precise definition of antibody responders will be defined by the trial steering committee prior to their unblinding of the study results. The primary analysis will exclude participants with elevated antibody levels at entry.

There are a number of secondary endpoints. Antibody levels will be determined at multiple time points to assess the immediacy and long-term durability of the response. SAEs through 60 months will be collected. AEs of lesser severity, including injection site reactions and targeted symptoms, will be collected following the initial vaccination and following the booster. In addition, substudies will examine a) T cell and memory B cell changes, b) viral shedding and c) the relationship between the levels of exposure to malaria and helminth infections and the rate of decline in antibody titre to Ebola.

#### 2.7. Sample size

Sample size was established for the 5-arm study to provide power to compare safety and immunogenicity separately for adults (N=1,400) and children (N=1,400) (see Figure 1). The target was reached and 2,802 participants were enrolled under protocol Version 4.0. Under Versions 2.0 and 3.0 of the PREVAC protocol 1,987 participants (537 in Version 2.0 and 1,450 in Version 3.0) were randomized. Thus, the total number randomized cumulatively in Version 2.0, 3.0 and 4.0 is 4,789 participants. A statistical analysis plan describes how data from Version 2.0 and Version 3.0 will be combined with data from Version 4.0 for selected objectives.

Sample size for adults for Version 4.0 has been reduced from what was originally planned (from 3,500 to 1,400). In part, this is because of the delays, and resulting costs, incurred before Version 4.0 could be implemented; in part, this is because a very conservative estimate of sample size was specified for adults in earlier versions of the PREVAC protocol; and in part, this is because of the plan to use data collected in Version 2.0 and 3.0 with data collected in Version 4.0 to address some objectives. Even with this reduction in sample size for adults, power for comparing the major efficacy and safety outcomes for adults and children separately is excellent.

Sample size is greater than what is required to address the primary objectives because if a correlate of protection is identified, the vaccine strategies will be compared with one another for that correlate using an intention to treat analysis. Expected differences between vaccine groups may be smaller than comparisons with placebo and the correlate may have greater variability than the assay which will be used to measure antibody levels to address the primary, secondary and exploratory objectives. The larger sample size will also permit the exploration of subgroups and preserve power in the event there are more participants with elevated antibodies at baseline than anticipated.

For the planned antibody comparisons of each vaccine strategy versus placebo at 12 months, the planned sample is based on data from PREVAIL I.[52] In PREVAIL I antibody levels measured at baseline, one week and at one month are briefly summarized below.

- 4.0% of participants had elevated antibodies to the EBOV at baseline. An elevated baseline antibody level in PREVAIL I was defined using the distribution of antibody levels measured from sera collected in 2004 and 2001 from 92 adults in Mali, a region where there had been no reports of ZEBOV transmission or disease during this time period. Baseline antibody levels were considered elevated in PREVAIL I if they were greater than 3 standard deviations (SD) above the mean (607 enzyme-linked immunosorbent assay units/mL based on the FANG ELISA assay).[52]
- Among participants without elevated antibodies at baseline (96%), the percent with an antibody response at week 1 was 2.5% for those given the rVSVΔG-ZEBOV-GP vaccine and 1.5% for those given placebo (p=0.36).

By month 1 antibody levels had increased substantially and 83.7% of participants given the rVSVΔG-ZEBOV-GP vaccine had an antibody response as compared to 2.8% of placebo participants (p<0.001). At 12 months, 79.5% of participants given the rVSVΔG-ZEBOV-GP vaccine had an antibody response versus 6.8% of participants in the placebo group (p<0.001). Responders during follow-up were defined as having a change in log-transformed antibody levels greater than 2 SD of the change in the placebo group at month 1.

With type 1 error = 0.0167 (2-sided) to adjust for the three comparisons, separately for adults and for children in all age groups combined, and power = 0.90, even if the percent with a positive antibody response at 12 months is 50% in a vaccine group, with equal allocation, approximately 30 participants per group (60 participants total) are needed assuming the percent in the placebo group with a positive antibody response is approximately 7%. With unequal allocation as for the rVSVΔG-ZEBOV-GP with the booster versus placebo comparison, a total of 63 participants (21 vaccinated with rVSVΔG-ZEBOV-GP with a boost and 42 vaccinated with placebo), a difference of 50% versus 7% can also be detected at 12 months with 87% power. These sample size estimates indicate that power for the planned subgroup analysis by age is also appropriate.

The planned sample size is also adequate for the comparisons with placebo if more than 4% of participants are antibody positive at baseline and excluded from the primary analysis, and if there is some missing data 12 months.

If the same correlate of protection is identified for both vaccine strategies, the vaccine strategies will be compared with one another at multiple time points. For the contrast with the smallest sample size (200 adults or children assigned rVSVΔG-ZEBOV-GP with a boost versus 400 vaccinated with rHAd26/MVA or rVSVΔG-ZEBOV-GP without a boost), power is 0.83 to detect an absolute difference of 15% in the antibody response (50% versus 35%) at the 0.0167 (2-sided) level of significance, assuming 10% of participants are excluded due to a positive antibody level at entry or for missing data. If the percent with a positive antibody response at 12 months is 75%, power is 0.87 to detect a 15% lower antibody response for this planned contrast.

For SAEs, we estimate that 5% of participants in the placebo group will experience an SAE by 12 months. This estimate is based on data from PREVAIL I where after 12 months of follow-up 11.8% of participants assigned to placebo developed an SAE. In PREVAIL I most of the SAEs were due to malaria events which did not require hospitalization.[52] Thus, with the exclusion of laboratory confirmed malaria events that did not require hospitalization, the 12 month percent with SAEs in this study is expected to be lower. Among adults and children, considered separately, with type 1 error = 0.0167 (2-sided), power is 0.88 to detect an increase of 7% (to 12%) for the rHAd26/MVA and for the rVSVΔG-ZEBOV-GP without a boost strategies compared to placebo assuming 400 per group. For the rVSVΔG-ZEBOV-GP with a boost (N=200) versus placebo (N=400) comparison for adults and children, power is 0.86 to detect an increase of 9% (to 14%).

Sample size is also sufficient for comparing the vaccine strategies with the pooled placebo group for the antibody response at day 14 (an important secondary objective that address the immediacy of the immune response). For example, if the antibody response at 14 days is 25% in a vaccine group and 7% in the placebo group, fewer than 250 participants total are needed.

Sample size is also sufficient for assessing the three vaccine strategies for long-term antibody response until 60 months following randomization. With type 1 error = 0.0167 (2-sided) to adjust for the three comparisons, separately for adults and for children in all age groups combined, power = 0.90, with equal allocation of 400 participants per arm (for the rHAd26/MVA-BN-Filo and for the rVSVΔG-ZEBOV-GP without a boost strategies compared to placebo), assuming the percent in the placebo group with a positive antibody response is approximately 7% at 60 months, a difference of at least 8% in the active vaccine arm can be demonstrated.

With unequal allocation as for the rVSVΔG-ZEBOV-GP with the booster (n=200) versus placebo comparison (n=400), a difference of at least 10% in the active vaccine arm can be demonstrate at 60 months with 90% power and type 1 error = 0.0167 (2-sided).

#### 2.8. Study cohort

This phase 2 study is conducted in Guinea, Liberia, Mali, and Sierra Leone.

As part of the public health campaign to prevent EVD, there was widespread communication about the trial to encourage volunteers to go to a vaccination center near where they live.

Our goal was to include 1,400 adults and 1,400 children. This goal was reached in December 2018. The gender and age distributions of those enrolled were closely monitored.

Eligibility for enrolment in the trial was assessed on a first-come first-serve basis; in case of oversubscription we ensured that access was made on an equitable basis that did not compromise the external validity of the trial.

##### 2.8.1. Inclusion criteria

The inclusion criteria for the study are broad reflecting the target population that would eventually receive efficacious vaccines.

- Informed consent/assent
- Age ≥ 1 year
- Planned residency in the area of the study site for the next 12 months
- Willingness to comply with the protocol requirements

##### 2.8.2. Exclusion criteria

- Fever > 38º Celsius

- History of EVD (self-report)
- Pregnancy (a negative urine pregnancy test is required for females of child-bearing potential, i.e., females who have experienced menarche or who are aged 14 years and older)
- Positive HIV test for participants < 18 years of age
- Reported current breast-feeding
- Prior vaccination against Ebola (self-report)
- Any vaccination in the past 28 days or planned within the 28 days after randomization (initial vaccination)
- In the judgement of the clinician, any clinically significant acute/chronic condition that would limit the ability of the participant to meet the requirements of the study protocol

#### 2.9. Randomization and blinding

Eligible participants were randomized in a 2:1:2:1:1 allocation to rHAd26/MVA (prime and boost), 0.5 mL of placebo (prime and boost), rVSVΔG-ZEBOV-GP without a boost, rVSVΔG-ZEBOV-GP with a boost, or, 1mL of placebo (prime and boost) (Figure 1). Syringes were pre-labelled with a unique SID number according to a centrally prepared randomization schedule. The tear-off label also included a bar code identifier. For each vaccination center, the randomization schedule was prepared using block randomization to ensure the desired allocation ratio for the five arms of the study for each vaccination center.

It was possible that the person carrying out the vaccination was able to differentiate syringes by the fill volume. The rVSVΔG-ZEBOV-GP vaccine has a 1 mL fill volume and the rHAd26/MVA vaccines have a 0.5 mL fill volume. The person carrying out the vaccination did not know whether vaccine or matched placebo is given.

At the time of vaccination, a tear-off label on the syringe that includes the SID was attached to the baseline CRF (or an eCRF will be used). This was the primary link used between the vaccine administered and the PID, and PIDs associated with SIDs are available to the pharmacy.

With this approach, randomization did not occur until the participant is vaccinated with the prime vaccine.

Participants randomized to the rVSVΔG-ZEBOV-GP vaccine without a boost for the initial vaccination received placebo (1.0 mL fill) at 56 days. Those randomized to receive rHAd26/MVA initially received the MVA-BN-Filo boost at 56 days in a syringe with a 0.5 mL fill; and those randomized to rVSVΔG-ZEBOV-GP with a boost received the rVSVΔG-ZEBOV-GP vaccine again at 56 days in a syringe with a 1 mL fill. Two groups received placebo (saline) for the prime and booster vaccination. For one group, a 0.5 mL fill syringe was prepared for each vaccination and for the other a 1.0 mL fill syringe was prepared for each vaccination. The syringe used at 56 days was labeled with the volunteer’s PID.

Study participants and clinical staff assessing the study participants for safety and laboratory outcomes are fully blinded until all participants complete 12 months of follow-up. The laboratories carrying out the safety and immunogenicity analyses are blinded to the vaccine assignment. Proceduresare put into place to break the blind if necessitated by a medical emergency. If unblinding of a participant for safety reasons is required, it is documented.

Following the Guinea outbreak occurring in 2021 and the vaccine licensures, the participants who received the placebo will be vaccinated with a single dose of rVSVΔG-ZEBOV-GP vaccine in Liberia and Mali and with the Ad26.ZEBOV/MVA-BN-Filo vaccine strategy in Guinea and Sierra Leone during their next long-term follow-up visit, or during additional visits if their next scheduled long-term follow-up visit is not approaching soon. The participants who received an incomplete Ad26.ZEBOV/MVA-BN-Filo vaccine strategy will be offered a single dose of the Ad26.ZEBOV or MVA-BN-Filo vaccine in Guinea and Sierra Leone and a single dose of rVSVΔG-ZEBOV-GP in Liberia and Mali.

After 12 month follow-up visits have been completed for all participants, and after the clinical and safety database lock for the M12 data, the study will be unblinded. Annual follow-up visits will continue for 4 additional years from the 12 month visit.

### 3. STUDY PLAN

Vaccination centers were established in West Africa to consent, vaccinate and follow study participants. A CRF with a unique PID is completed for each participant consented and screened. The CRF is an eCRF

At the time of vaccination, study volunteers were given an identification card that identifies them as a study participant. The identification card includes contact information in the event they experience vaccine side effects, or signs and symptoms of Ebola. Volunteers are encouraged to contact the vaccination center by telephone at any time of day if they have any AEs. If the participant experiences an AE requiring immediate attention, they will be given instructions on which hospital to go to for evaluation and treatment.

Women of childbearing potential were told to use adequate birth control measures for at least 3 months following the initial vaccination and booster vaccination. Men who are sexually active with women of child-bearing potential were told to use condoms for at least 3 months following the initial vaccination and booster vaccination. Participants were also instructed to avoid sharing items during the period of potential virus shedding.

Following randomization, all participants were seen at 7, 14, 28, 56 and 63 days, and at 3, 6 and 12 months. After 12 months, participants are seen annually at 24, 36, 48, and 60 months. In addition to these study visits, volunteers are encouraged to contact the vaccination center by telephone if they experience AEs.

Participants who received the placebo will be vaccinated with a single dose of rVSVΔG-ZEBOV-GP vaccine in Liberia and Mali and with Ad26.ZEBOV/MVA-BN-Filo vaccine strategy in Guinea and Sierra Leone during their next long-term follow-up visit, or during additional visits if their next scheduled long-term follow-up visit is not approaching soon. The participants who received an incomplete Ad26.ZEBOV/MVA-BN-Filo vaccine strategy will be offered a single dose of the Ad26.ZEBOV or MVA-BN-Filo vaccine in Guinea and Sierra Leone and a single dose of rVSVΔG-ZEBOV-GP in Liberia and Mali.

#### 3.1. Screening/Baseline data collection

Prior to randomization, children must have an HIV test. There is no specific time period within which the HIV test must have been performed. If the test is negative, they will be eligible for randomization

Baseline data will include:

- Informed consent type and date
- Birth month/year and gender
- Urine pregnancy test result for females who are of childbearing potential, i.e., females who have experienced menarche or who are aged 14 years and older
- High risk occupation (e.g., work involved or might involve contact with living or dead persons with Ebola such as a nurse, physician, ambulance driver, burial crew member or other healthcare worker or not a healthcare worker)
- History of past contact with a person with EVD
- Name and contact information for participant (to be kept locally)
- Names and contact information of 2 alternate contact people (to be kept locally)
- Body temperature
- Height and weight
- Mid-upper arm circumference (for children 1-5 years).
- Serum for aspartate transaminase (AST), alanine transaminase (ALT), creatinine, and potassium
- Blood for CBC with differential, platelet count, hemoglobin, hematocrit, mean corpuscular volume (MCV), and red cell distribution width (RDW)
- Blood for assessing HIV and syphilis serostatus
- Stored serum for immunogenicity testing and future research
- Stored cells for assessing T cell and memory B cell function (subsample of adults enrolled at a single site in Guinea)
- Stored saliva for assessing viral shedding (subsample of children)
- Antipyretics prescribed/offered

#### 3.2. Vaccine administration

Following informed consent and the collection of baseline data, the study vaccines were administered to participants who met all of the eligibility criteria via intramuscular injection in the upper, outer aspect of the arm (deltoid region).

Children may be vaccinated in the thigh. Prior to injection, the area to be injected was prepared using an alcohol swab. Vaccine was administered using a 1 or 3 mL syringe with a sterile needle. While holding the needle at an approximate 90-degree angle to the skin, the needle was directed through the skin, into the muscle of the arm (or thigh). The location of vaccination was recorded on the case report form.

At the time of prime vaccination administration, the tear-off portion of the SID label on the syringe was placed on the baseline CRF.

Study participants/family members were provided with information on how to minimize risk of infection with Ebola. After vaccination, participants were watched closely for at least 30 minutes. Injection site reactions, targeted symptoms of any grade severity, and grade 3 and 4 AEs that may have occurred during this observation period were recorded.

Participants have been offered medication post-vaccination for fever and pain relief depending on each site’s policy. If participants received or took their own medication for fever and pain following vaccination, it was recorded.

The same procedures were followed for the MVA-BN-Filo and rVSVΔG-ZEBOV-GP booster/placebo vaccination at 56 days. As stated below, the window for the booster vaccination at 56 days is 53 to 66 days (-3,+10 days).

Post M12 visit, during the long-term follow-up, the participants who received the placebo will be vaccinated with a single dose of rVSVΔG-ZEBOV-GP vaccine in Liberia and Mali and with  the Ad26.ZEBOV/MVA-BN-Filo vaccine strategy in Guinea and Sierra Leone. The participants who received an incomplete Ad26.ZEBOV/MVA-BN-Filo vaccine strategy will be offered a single dose of the Ad26.ZEBOV or MVA-BN-Filo vaccine in Guinea and Sierra Leone and a single dose of rVSVΔG-ZEBOV-GP in Liberia and Mali.

Temporary non-vaccination criteria:

- Fever > 38º Celsius
- Pregnancy (a negative urine pregnancy test is required for females of child-bearing potential, i.e., females who have experienced menarche or who are aged 14 years and older)
- Reported current breast-feeding (self-report)
- Any vaccination in the past 28 days or planned within the 28 days after trial vaccination

Non-vaccination criteria:

- EVD notified in the eCRF
- For minor participants: change in HIV status since enrolment (self-report)
- Previous Ebola vaccination outside of the study including incomplete vaccines strategies

- Known medical history or significant risk factors for a thrombotic and/or thrombocytopenic event (for participants who will receive the Ad26.ZEBOV or MVA-BN-Filo vaccine)

#### 3.3. Follow-up data collection

Following randomization, all participants are seen at the following visits (visit window): 7 (± 3 days), 14 (± 3 days), 28 (± 7 days), 56 (53 to 66 days) and 63 days (7 ± 3 days after the booster vaccination), and at 3 (± 14 days), 6 (± 1 month), and 12 (± 1 month) months. Participants will continue to be seen at 24 (± 6 months), 36 (± 6 months), 48 (± 6 months) and 60 (- 6 months, +1 month) months. The site will be instructed to schedule the follow-up visits as close as possible as to the theoretical date. Also, every effort should be made to follow-up participants who missed visits.

At each follow-up visit, volunteers will be asked a few questions to assess the development of any serious medical conditions (e.g., malaria), the possibility of unreported SAEs, receipt of Ebola vaccination outside of the protocol procedure (in particular in the context of Ebola outbreak response), receipt of COVID19 vaccination outside of the protocol procedure, and for women whether they are pregnant. Temperature will be recorded, and blood will be drawn for immunogenicity testing and future research.

The following additional information was collected at 7, 14, 28, and 63 days and at 3 months (approximately 35 days after the booster) following randomization:

- Grade 3 and 4 AEs and targeted symptoms since vaccination
- Injection site reactions since vaccination

The following additional specimens and information were/will be obtained for children: 1) during the first week following the prime and booster vaccination, there was daily contacts with children to assess injection site reactions, targeted symptoms and SAEs, and to measure temperature; 2) on day 7 and 63, serum for AST, ALT, creatinine, and potassium and blood for CBC with differential, platelet count, hemoglobin, hematocrit, MCV, and RDW were obtained; 3) on day 28, at months 3 and 6 and annually weight will be recorded; 4) annually height will be measured; and 5) mid-upper arm circumference (children aged 1-5 years) will be measured at months 3, 6, 12, 24, 36, 48, and 60.

For a subsample of adults, at baseline and days 7, 14, 56, 63, 70, and month 12, cells for assessing T cell and memory B cell function were obtained. Cells will also be collected at 24, 36, 48 and 60 months (see Appendix D).

At one site saliva was collected for a subsample of children for measurement of viral shedding (see Appendix E).

On day 56 prior to the booster vaccination, a urine pregnancy test was performed for females of child-bearing potential, i.e., females who have experienced menarche or who are aged 14 years and older. If a woman is pregnant, she could not receive the booster vaccination. However, she remained under follow-up.

Following the booster vaccination, participants were watched closely for at least 30 minutes. Injection site reactions, targeted symptoms of any grade severity, and grade 3 and 4 AEs that may have occurred during this observation period were recorded.

Some additional quantitative and qualitative data will be harvested through specific questionnaires for the substudy on barriers and facilitators to PREVAC retention (see Appendix G)

After unblinding, all participants will be informed of which vaccine they received in a specific newsletter and prior to any participant new vaccination. An additional participant visit may be conducted for unblinding if required.

After the consenting procedure, participants who received the placebo or an incomplete Ad26.ZEBOV/MVA-BN-Filo vaccine strategy and who have no (temporary) non-vaccination criteria will be vaccinated during their long-term follow-up visit, or during additional visits if their next long-term follow-up visit is not approaching soon. Following the new vaccination, participants will be watched closely for at least 30 minutes. SAE that may occurr during this observation period will be  recorded. The participant will be informed to continue to report SAEs and that there will be no additional blood sample required for this vaccination visit.

#### 3.4. Laboratory Tests

Safety laboratory tests will be performed by a clinical laboratory at the site.

Safety laboratory reports will be generated and returned to the vaccination center(s) prior to the next scheduled visit.

At one site peripheral blood mononuclear cells (PBMCs) are collected for a subsample of adults and stored for the investigation of T cell and memory B cell responses (see Appendix D).

At one site saliva was collected for a subsample of children for measurement of viral shedding (see Appendix E).

At two sites, one in Sierra Leone one in Guinea, blood samples are collected for the parasitology assays for the malaria substudy (see Appendix F).

Immunogenicity testing are performed using a validated FANG GP-ELISA assay on baseline, Day 28, and Months 3, 12, 24, 36, 48, and 60 specimens. The participating pharmaceutical companies are provided samples from these visits and have the primary responsibility for obtaining this testing. A non-validated FANG GP-ELISA assay or another assay may also be used at these and other time points.

##### 3.5. Biometric Data

Digital finger prints and iris scans may also be collected, digitized and stored in a secure database. These data would be used to ensure appropriate matching of study participants with PIDs.

#### 4. ASSESSMENT OF SAFETY

Safety SOPs describe procedures for reporting deaths, Ebola events, pregnancies, pregnancy outcomes, and SAEs, including assessments of severity and the relationship of the vaccine to events. Below a brief summary is provided.

##### 4.1. SPECIFICATION OF SAFETY PARAMETERS

###### 4.1.1. Definition of AE

An AE is any untoward medical occurrence in a randomized volunteer who is administered the trial product (vaccine or placebo), whether or not considered related to the vaccination.

###### 4.1.2. Definition of an Adverse Reaction (AR)

An AR is any untoward and unintended response related to an investigational medicinal product (in this case, vaccine or placebo).

###### 4.1.3. Definition of SAE

An AE is considered "serious" if, in the view of either the investigator or sponsor, it results in any of the following outcomes:

a. Death
b. Events that are life-threatening

   Note: The term ‘life-threatening’ in the definition of ‘serious’ refers to an event in which the participant was at risk of death at the time of the event. It does not refer to an event, which hypothetically might have caused death, had it been more severe.

c. Events requiring hospitalisation
d. Events requiring prolongation of existing hospitalisation,

   Note: In general, hospitalization signifies that the participant has been admitted at the hospital or emergency ward for observation and/or treatment that would not have been appropriate in the physician’s office or in an out-patient setting. Complications that occur during hospitalization are also considered as SAEs. If a complication prolongs hospitalization or fulfils any other serious criteria, the event will also be considered serious. When in doubt as to whether ‘hospitalization’ occurred or was necessary, the AE should be considered serious.

Hospitalization for elective treatment of a pre-existing condition (known or diagnosed prior to informed consent signature) that did not worsen from baseline is NOT considered an SAE.

e. Events resulting in a persistent or significant incapacity or substantial disruption of the ability to conduct normal life functions

Note: The term disability means a substantial disruption of a person's ability to conduct normal life functions. This definition is not intended to include experiences of relatively minor medical significance such as uncomplicated headache, nausea, vomiting, diarrhea, influenza-like illness, and accidental trauma (e.g. sprained ankle) which may interfere or prevent everyday life functions but do not constitute a substantial disruption.

f. Congenital anomaly or birth defects in the offspring of a study participant

g. Important medical events, that may not result in death, be life-threatening, or require hospitalization may be considered serious when, based upon appropriate medical judgment, they may jeopardize the participant and may require medical or surgical intervention to prevent one of the outcomes listed in this definition. Examples of such medical events include:

- Any suspected transmission of an infectious agent by a medicinal product
- Allergic bronchospasm requiring intensive treatment in an emergency room or at home
- Blood dyscrasias or convulsions that do not result in inpatient hospitalization
- Development of drug dependency or drug abuse.
- Cranial nerve disorders, including paralyses/paresis (e.g. Bell's palsy)
- Optic neuritis
- Multiple sclerosis
- Transverse myelitis
- Guillain-Barré syndrome, including Miller Fisher syndrome, Bickerstaff's encephalitis and other variants
- Acute disseminated encephalomyelitis, including site specific variants: e.g. non-infectious encephalitis, encephalomyelitis, myelitis, myeloradiculomyelitis
- Myasthenia gravis and Lambert-Eaton myasthenic syndrome
- Immune-mediated peripheral neuropathies and plexopathies, (including chronic inflammatory demyelinating polyneuropathy, multifocal motor neuropathy and polyneuropathies associated with monoclonal gammopathy).
- Narcolepsy
- Isolated paresthesia of >7 days duration

Note: For this protocol, EVD events and laboratory confirmed malaria events that do not require hospitalization will not be considered as SAEs. Laboratory diagnostic criteria for confirmation of malaria should be based on local criteria.

##### 4.1.4. Serious Adverse Reaction (SAR)

Any AR which is also considered serious.

##### 4.1.5. Suspected Unexpected Serious Adverse Reaction (SUSAR)

An AR that is both unexpected (not consistent with the applicable product information) and also meets the definition of a SAR.

##### 4.1.6. Definition of Unanticipated Problems (UP)

An UP is any incident, experience or outcome that meets all of the following criteria:

1. Unexpected in terms of nature, severity, or frequency in relation to:
   a. the research risks that are described in the ethical committee/institutional review board (IRB)-approved research protocol and informed consent document; Investigator's Brochure or other study documents; and
   b. the characteristics of the population being studied; and

2. Possibly, probably, or definitely related to participation in the research ("possibly related" means there is a reasonable possibility that the incident, experience, or outcome may have been caused by the procedures involved in the research); and

3. Places study participants or others at a greater risk of harm (including physical, psychological, economic, or social harm) than was previously known or recognized.

Furthermore, a UP could be an expected event that occurs at a greater frequency in one of the vaccine groups than the placebo group.

##### 4.1.7. Adverse Events of Special Interest

AESIs (including potential AESIs) are significant AEs that are judged to be of special interest because of clinical importance, known or suspected class effects, or based on nonclinical signals. AESIs and potential AESIs will be carefully monitored during the study by the sponsor.

AESIs and potential AESIs must be reported to the sponsor within 24 hours of awareness irrespective of seriousness (i.e., serious and non- serious AEs) or causality following the procedure described above for SAEs.

Specific requirements for the AESI are described below.

###### 4.1.7.1. Thrombosis with Thrombocytopenia Syndrome

TTS has been observed very rarely following vaccination with Janssen COVID19 vaccine and is considered to be an AESI in this study.[53] TTS is a syndrome characterized by a combination of both a thrombotic event and thrombocytopenia.[54]

Because this syndrome is rare and not completely understood, all cases of thrombosis and/or symptomatic thrombocytopenia will be considered a potential case of TTS and should be reported to the sponsor within 24 hours of awareness. Each potential AESI will be reviewed to identify a TTS case. A potential TTS case is defined as:

- Thrombotic events: suspected deep vessel venous or arterial thrombotic events as detailed in Appendix I

and/or

- Symptomatic thrombocytopenia, defined as platelet count below LLN for the testing lab

Symptoms, signs, or conditions suggestive of a thrombotic event or symptomatic thrombocytopenia should be recorded and reported as a potential AESI even if the final or definitive diagnosis has not yet been determined, and alternative diagnoses have not yet been eliminated or shown to be less likely. Follow-up information and final diagnoses, if applicable, should be submitted to the sponsor as soon as they become available.

In the event of symptomatic thrombocytopenia, study site personnel should report the absolute value for the platelet count and the reference range for the laboratory test used.

For either a thrombotic event or symptomatic thrombocytopenia, it is recommended to test for anti-Platelet Factor 4 antibodies at the local laboratory or substitute local laboratory; repeat testing may be requested for confirmation upon sponsor discretion.

AESIs, including potential AESIs, will require enhanced data collection and evaluation. Every effort should be made to report as much information as possible about the event to the sponsor in a reasonable timeframe.

###### 4.1.7.2. Treatment and Follow-up Recommendation

The medical management of thrombotic events with thrombocytopenia is different from the management of isolated thromboembolic diseases. Study site personnel and/or treating physicians should follow the algorithm for testing and treatment of participants presenting with thrombosis and thrombocytopenia 5 to 30 days after vaccination stated in the PREVAC procedure SOP_SAF_01 “Safety Reporting”.

#### 4.2. CLASSIFICATION OF AN AE

##### 4.2.1. Relationship to study product

The relationship assessment is one of the criteria used when determining regulatory reporting requirements. This procedure will also apply for the new vaccination under protocol v6.0 and v7.0.

Relationship of SAEs should be assessed by the site physician using the following question:

Is there a reasonable possibility that the AE may have been caused by the study product (vaccine)?

YES : The AE is known to occur with the study product, there is a reasonable possibility that the study product caused the AE, or there is a temporal relationship between the study product and event. Reasonable possibility means that there is evidence to suggest a causal relationship between the study product and the AE.

NO : There is not a reasonable possibility that the administration of the study product caused the event, there is no temporal relationship between the study product and event onset, or an alternate etiology has been established.

##### 4.2.2. Unexpectedness

Only SAEs will be assessed as expected or unexpected. An SAE will be considered unexpected if the nature, severity, or frequency of the event is not consistent with the risk information previously described for the vaccine.

An SAE is considered "unexpected" if it is not listed in the Investigator’s Brochure or is not listed at the specificity or severity that has been observed; or, if an Investigator’s Brochure is not required or available, is not consistent with the risk information described in the general investigational plan or elsewhere in the protocol, as amended.

*For example, under this definition, hepatic necrosis would be unexpected (by virtue of greater severity) if the* Investigator’s Brochure *referred only to elevated hepatic enzymes or hepatitis. Similarly, cerebral thromboembolism and cerebral vasculitis would be unexpected (by virtue of greater specificity) if the* Investigator’s Brochure *listed only cerebral vascular accidents.*

"Unexpected," as used in this definition, also refers to AEs or suspected ARs that are mentioned in the Investigator's Brochure as occurring with a class of drugs or as anticipated from the pharmacological properties of the drug, but are not specifically mentioned as occurring with the particular drug under investigation.

#### 4.3. TIME PERIOD AND FREQUENCY FOR EVENT ASSESSMENT AND FOLLOW-UP

##### 4.3.1. Time Period and Frequency Assessment

#### 4.3.1.1. AE

Injection site reactions, targeted symptoms of any grade severity, and unsolicited Grade 3 or 4 AEs will be collected and reported on CRFs as defined by the protocol (see Appendix A). AEs of grade 1 or 2 severity other than injection site reactions and targeted symptoms will not be collected. Grade 3 or 4 signs or symptoms associated with laboratory abnormalities and clinically significant grade 3 and 4 laboratory abnormalities documented by the investigators as AEs should be reported on the CRFs. All other laboratory abnormalities will be summarized using the protocol required laboratory data that is in the central database. Laboratory abnormalities that are considered clinically significant should be reported as AEs or SAEs irrespective of the presence of symptoms.

Sites will be notified of laboratory abnormalities identified as part of the protocol required testing (baseline for all participants and also 7 days after the prime and booster vaccination for children). Laboratory abnormalities will be summarized for each DSMB review. Details for documenting laboratory abnormalities in the participants medical record are provided in the manual of operations.

Injection site reactions will include redness of skin, swelling/induration, pain/tenderness with activity, and itching at injection site.

The targeted symptoms will include reduced activity/somnolence, fatigue, vomiting, chills, abnormal sweating. skin lesions (macules, papules, purpura, petechiae), mouth ulcers, decreased appetite, feverishness, diarrhea, nausea, headache, dizziness, abdominal pain, muscle pain, joint swelling, and joint pain. For non-verbal children, subjective symptoms may not be assessed and irritability/fussiness, crying, and screaming will be assessed.

Injection site reactions and symptoms will be graded as follows: Grade 1 – symptoms causing no or minimal interference with usual social & functional activities; Grade 2 - symptoms causing greater than minimal interference with usual social & functional activities; Grade 3 - symptoms causing inability to perform usual social & functional activities; and Grade 4 - symptoms causing inability to perform basic self-care functions OR medical or operative intervention indicated to prevent permanent impairment, persistent disability, or death.

###### 4.3.1.2. SAEs, EVD events, Malaria Events

SAE and EVD events will be assessed and reported on CRFs as soon as the site becomes aware of the event. For EVD events, where possible, a sample of blood will be stored for future research. Malaria events will be assessed at each follow-up visit. Malaria events that meet the SAE definition (e.g., result in hospitalization) will be reported on the SAE form.

###### 4.3.1.3. Pregnancy

Women who reported being pregnant in the first 3 months of follow-up after the prime vaccination are followed for the outcome of their pregnancy. This procedure will also apply for the new vaccination under protocol v6.0 and v7.0.

The medical surveillance of the women and their children is reinforced: a particular attention was given to serious pathology occurring during pregnancy. An SAE form was completed if an AE during pregnancy or birth outcome meets the SAE definition. Congenital anomalies or birth defects should have also been reported as SAE.

#### 4.3.1.4. UP

UPs that are not related to the study vaccines, for example, a breach of subject confidentiality, is reported to the respective Ethical/Institutional Review Committees in accordance with the requirements of the Committee for oversight of the particular site where the UP occurs. UPs related to the study vaccines will be reported per AE/SAE reporting requirements in 4.3.1.1 and 4.3.1.2. Details for UP reporting are provided in the SOPs.

##### 4.3.2. Time of collecting information

All SAE events will be collected from the signing of informed consent until the end of the study of the participant.

##### 4.3.3. Follow-up

All SAE events must be followed until clinical recovery is complete and laboratory results have returned to normal or baseline, or until the event has stabilized. Follow-up should continue regardless of when this occurred in relation to the study conclusion.

#### 4.4. GENERAL REPORTING, RESPONSIBILITIES AND TIMELINES

Events Reported on CRFs by the investigators:

AEs (injection site reactions, targeted symptoms of any grade of severity and unsolicited grade 3 and 4 AEs), UPs, SAEs, deaths, Ebola events, malaria events, and pregnancy outcomesare reported on CRFs.

Events to be notified by the investigators to the Medical Officer and to Sponsors:

SAEs and Ebola events will be reported within 24 hours of their knowledge of the event.

The Medical Officer will assess the expectedness and relationship of SAE to the study vaccines. All events will be coded using the Medical Dictionary for Regulatory Activities (MedDRA®).

The plan for DSMB reporting is described in section 7.2 and in the statistical analysis plan.

Events to be notified to US Food and Drug Administration (FDA):

Only SUSARs will be notified to the FDA by the NIH within the legal timeframe:

- 7 calendar days in case of death or life-threatening events after the sponsor's initial receipt of the information; relevant complementary information should be collected and notified within 8 extra-days
- 15 calendar days for all other events after the sponsor's initial receipt of the information; relevant complementary information should also be collected and notified within 8 extra-days

Events to be notified to IRB/ethics committee (EC): Each sponsor will be responsible for the SUSAR and other events notification to their respective ECs or IRBs in compliance with local regulations.

Safety Reports:

The Safety Reports will be sent to the sponsors for review and validation. NIH is responsible for the Safety Reports submission to the FDA and each sponsor is responsible for the Safety Reports submission to their respective Health Authority and IRB/EC in compliance with local regulations. NIH will also forward Safety Reports to industry partners in accordance with provisions identified in the respective clinical trial agreements.

#### 4.5. HALTING RULES

##### 4.5.1. Study halting rules

The DSMB for this study meets frequently to assess the overall safety of study participants. The DSMB may pause enrolment in the event of vaccine-related deaths or SAEs that are considered vaccine-related. The DSMB may also pause enrolment or request that participants be notified if there is an increased frequency of unanticipated adverse effects.

##### 4.5.2. Participant halting rules

If a participant experienced an SAR (that is attributed to initial vaccination), the booster vaccination is not given. These participants will continue to be followed until the end of the study. This procedure will also apply for the new vaccination under protocol v6.0 and v7.0.

### 5. CLINICAL MANAGEMENT

#### 5.1. AEs and symptoms and signs of EVD

Study participants are given information on whom to call or see in the event they experience AEs and symptoms of EVD. If the participant experiences an AE requiring hospitalization, they will be given instructions on which hospital to go to for evaluation and treatment. The management of suspected and confirmed EVD are according to local standards.

#### 5.2. HIV and syphilis counseling

Participants who test positive for HIV or syphilis at study entry were provided with counselling and referred for treatment according to local standards.

#### 5.3. Study withdrawal

Every effort should be made to follow participants for blood draws and SAEs until the end of the study. Following vaccination, participants may withdraw from follow-up in the study at any time. They may resume participation at any time. Reasons for withdrawal will be recorded.

#### 5.4. Pregnancy

Women who reported being pregnant in the first 3 months of follow-up after the prime vaccination were followed for the outcome of their pregnancy. The outcome was reported on a CRF. Women who reported being pregnant prior to the booster vaccination or who have a positive pregnancy test at 56 days did not receive the booster vaccination at 56 days. This procedure will also apply for the new vaccination under protocol v6.0 and v7.0.

#### 5.5. Childhood vaccinations

Children were not enrolled if they received a vaccination in the 28 days before randomization. Children were advised not to receive another vaccination in the 28 days after the prime vaccination as well as prior to and after the booster vaccination unless the benefit of that vaccination was judged to be greater than the potential risk. For example, if a child had an unexpected exposure that places them at high risk of a disease (e.g., rabies, meningitis), they have been vaccinated.

#### 5.6. Concomitant participation in other clinical trials during the long-term follow-up

Participants are not authorized to participate in other Ebola vaccine clinical trials during their long-term follow-up.

### 6. PHARMACY PROCEDURES

#### 6.1. Study agents

##### 6.1.1. Ad26.ZEBOV and MVA-BN-Filo vaccines

The drug products, Ad26.ZEBOV and MVA-BN-Filo are provided by Johnson & Johnson. Ad26.ZEBOV are supplied in single dose glass vials with an extractable volume of 0.5 mL at a dose of $5x10^{10}$ vp. Ad26.ZEBOV is colorless to slightly yellowish or brownish, clear to slightly opalescent suspension, practically free of particles.

The MVA-BN®-Filo vaccine are supplied in single dose glass vials with an extractable volume of 0.5 mL at a dose of $1x10^8$ infectious units. MVA-BN®-Filo is a milky, light yellow suspension with no visible extraneous particles.

##### 6.1.2. rVSV∆G-ZEBOV-GP

The drug product, rVSV∆G-ZEBOV-GP, is provided by Merck. The vaccine is a sterile, aqueous, buffered solution composed of rVSV∆G-ZEBOV-GP drug product filled into single use vials with an extractable volume of 1 mL at a labelled dose of $\geq 7.2 \times 10^7$ pfu/mL with release specifications from $7.8x10^7$ pfu/mL to $2.3x10^8$ pfu/mL. The labelled dose of $\geq 7.2 \times 10^7$ pfu/mL is equivalent to the nominal dose of $2 \times 10^7$ pfu/mL and reflects a change in assay used to quantify potency. The product is a clear, colorless aqueous suspension free from particles.

In Version 2.0 of PREVAC, the rVSVΔG-ZEBOV-GP vaccine was not used. In Version 3.0, the rVSVΔG-ZEBOV-GP vaccine was given after 2-fold dilution for prime and boost. In Version 4.0, the undiluted dose was used for both the prime and boost vaccinations.

##### 6.1.3. Sterile Normal Saline

The placebo is sterile normal saline (sodium chloride 0.9 percent for injection, United States Pharmacopeia, preservative free).

#### 6.2. Study agent presentation and storage

##### 6.2.1. Study agent labels

Clinical supplies will be affixed with a clinical label in accordance with regulatory requirements.

##### 6.2.2. Study agent storage

Storage requirements as well as procedures for temperature deviations for the investigational vaccines are described in the Pharmacy Procedures.

Normal saline was stored at room temperature in accordance with the manufacturer label.

If deviations in storage temperature occur from the normal allowance for the pharmacy freezer, the affected investigational products will be quarantined and the storage temperature excursion will be promptly reported to the Principal Investigators, the IND sponsor, and product manufacturer: Johnson & Johnson for Ad26.ZEBOV and MVA-BN-Filo, Merck for rVSVΔG-ZEBOV-GP. The excursion will be evaluated and investigated and action will be taken to restore and maintain the desired temperature limits. Pending the outcome of the investigation, the manufacturer will notify the Principal Investigator, sponsor, and pharmacist if continued clinical use of the product is acceptable.

##### 6.3. Preparation of study agents for injection

For the initial vaccination, products were prepared in individual syringes at the vaccination site pharmacies in each country. The syringes for the 56 day vaccination were prepared by these pharmacies.

Products remained under the manufacturer required temperature range prior to preparation of syringes detailed in the Pharmacy Procedures.

Examples of syringe labels are included in the Pharmacy Procedures as well as details related to product receipt, accountability, storage (shelf-life), preparation and disposition.

##### 6.4. Study agent accountability

Study pharmacist in each country is responsible for maintaining an accurate record of vaccine supplies for this study, as well as documenting the destruction of unused syringes according to local standards for biohazardous waste disposal. Electronic documentation as well as paper copies may be used. Accountability instructions are included in the Pharmacy Procedures.

#### 7. EVALUATION

##### 7.1. Definition of the study completion

For regulatory purpose, the study completion for PREVAC is defined as the time when database locks are achieved for both the M12 non-validated and validated FANG GP-ELISA assay results.

The study completion for the long term follow-up of PREVAC is defined as the time when database locks are achieved for both the M60 non-validated and validated FANG GP-ELISA assay results.

#### 7.2. Planned data analyses

A first database lock will be performed after all the required M12 data will be collected.

A second database lock will performed after all the required M60 data will be collected.

Final M12 data analyses will be carried out according to a statistical analysis plan that is updated prior to unblinding. The planned analyses below only describe those planned for participants enrolled under Version 4.0. Separate analyses will be carried out for participants enrolled under Version 2.0 and 3.0. These analyses and analyses which will be pooled across protocol versions are described in the statistical analysis plan.

Final M60 data analyses will be carried out according to a statistical analysis plan that is updated prior to M60 database lock. All the participants randomized in PREVAC will take part of the long term follow-up (M24 – M60).

Prior to unblinding, the analysis plan for the immunogenicity substudy (Appendix D) and the viral shedding substudy (Appendix E) will also be finalized.

For the purposes of regulatory filings there is a specific primary objective for each participating pharmaceutical company involving a single follow-up time point and the use of a validated assay for immunogenicity testing. Thus, the two companies may also prepare a statistical analysis plan prior to unblinding and regulatory filing that describes analysis plans for their primary objective (e.g., comparisons with matched placebo for immunogenicity due to gamma irradiation of a subset of specimens) as well as plans for controlling type 1 error for investigations of secondary endpoints and subgroups.

The following plan outlines data analyses that are carried out during the trial by the unblinded statisticians for the independent DSMB.

Adults and children will be analyzed separately and combined. Baseline characteristics and follow-up completeness will be summarized by vaccine group.

Prior to carrying out the endpoint analyses, which pool the two placebo groups, the two placebo groups will be compared for the immunogenicity and safety outcomes. The primary analyses for adult and children will be carried out at the 0.0167 (2-sided) level of significance and will compare the three vaccine strategies with the pooled placebo group for the percent with a positive antibody response at 12 months. Each pair-wise comparison will be carried out using Mantel-Haenszel chi-square statistics which consider stratification by vaccination center. Similar methods will also be used to compare groups for the immediacy of the antibody response at 14 days. These analyses will exclude those participants with elevated antibody levels at baseline. Participants were not screened at study entry for elevated antibody levels. These exclusions will be carried out during analysis.

Analyses will also be carried out for all randomized participants irrespective of antibody level at baseline.

Similar methods will be used to compare antibody responses of each vaccine strategy with the pooled placebo group at other time points, including 28 days after randomization and 28 days following the booster vaccination.

Similar methods will be used until vaccination of the placebo group during long-term follow-up (24, 36, 48 and 60 months after randomization). After that point comparisons to placebo will require consideration of the decreasing placebo sample size and the durability of vaccine responses within groups will be assessed, using appropriate statistical models for repeated longitudinal data (mixed effect regression models).

The two additional primary objectives for immune responses, pooled rVSVΔG-ZEBOV-GP groups versus pooled placebo groups 28 days after randomization for Merck, and rHAd26/MVA versus pooled placebo groups approximately 28 days following booster vaccination for Janssen, will be carried out at the 0.05 (2-sided) level of significance.

All comparisons (for adults, children, and for adults and children combined) of immune responses and safety outcomes at other time points will be carried out at the 0.0167 (2-sided) level of significance when there are three pair-wise comparisons made (e.g., each vaccine strategy versus pooled placebo) and at the 0.025 (2-sided) level of significance when there are two pair-wise comparisons made (e.g., for comparisons at days 7, 14, 28 and 56 with the pooled placebo group in which the two rVSVΔG-ZEBOV-GP strategies are pooled).

SAEs will be classified by system organ class according to MedDRA®. The total number of events and the number of participants with at least one event will be reported. Mantel-Haenszel chi-square statistics will be used for each pair-wise comparison of the number of participants with an SAE through 12 months of follow-up; time-to-event methods, e.g., Kaplan-Meier, Cox regression, will also be used to compare groups for SAEs over the 12 month follow-up period. Analyses will also be carried out for SAEs judged to be related to the vaccine. Similar analyses will be performed for SAEs reported through 60 months.

Each vaccine group will be compared with the pooled placebo group using Mantel-Haenszel chi-square tests and time-to-event methods for diagnoses of laboratory-confirmed malaria through 12 and 60 months of follow-up.

For safety and efficacy outcomes assessed before the booster at 56 days, the two groups given the rVSVΔG-ZEBOV-GP prime vaccination will be combined for comparisons with pooled placebo group For outcomes that are assessed after 56 days, each of the three vaccination strategies will be compared with the pooled placebo group.

Mantel-Haenszel chi-square tests will be used to compare each vaccine with placebo for the percent with grade 3 or 4 AEs and with targeted symptoms reported at 7, 14, and 63 days. Similar methods will be used to compare injection site reactions reported immediately following prime vaccination and at 7 and 14 days. Likewise, treatment differences in injection site reactions reported immediately following booster vaccination and through day 63 (7 days after booster vaccination) will be summarized with similar methods. In the event that there is evidence that safety outcomes differ between the two placebo groups, analyses will also be performed for each vaccine versus its matched placebo.

Longitudinal mixed models will be used to compare log-transformed antibody levels collected at multiple follow-up time points during follow-up and to estimate slopes of antibody evolution during long-term follow-up. Results will be summarized as geometric means and geometric mean ratios. These analyses will be carried out among participants who did not have elevated antibody levels at baseline. Baseline titer level will be included as a covariate. These analyses will be supplemented with scatter plots that display fold increases in antibody titers, line plots, box plots and reverse cumulative distributions of antibody levels at each visit for each vaccine group.

For children, changes in biochemical and CBC test values after 7 and 63 days will be compared for each vaccine versus the pooled placebo group using analysis of covariance with the baseline laboratory level as a covariate. Laboratory test results will be graded for severity according to the Division of AIDS (DAIDS) AE Table and local laboratory norms, and the proportion with grade 3 or 4 laboratory abnormalities will be compared.

For children, findings based on contacts during the 7 days following the prime and booster vaccination will be summarized each day. Trends over the 7 days will be examined for each vaccine versus placebo comparison.

Subgroup analyses for the primary endpoints and major secondary outcomes, including SAEs and other safety data, will be performed to determine whether vaccine versus placebo differences vary qualitatively across various baseline-defined subgroups. Subgroup analyses will be performed using regression analysis by age, gender, country, vaccination center, health care worker or job involving close contact with EVD cases, baseline laboratory abnormalities, HIV status, and nutritional status as determined by body mass index. An overall test of heterogeneity will provide evidence of whether the magnitude of the vaccine pair-wise comparison for efficacy/safety outcomes varies across these baseline subgroups.

In addition to the separate analysis for adults and children for each vaccine group versus placebo for antibody levels and safety outcomes, the endpoints will also be summarized pooling the data for adults and children. Methods similar to those described above will be used after stratification on age.

Analyses which compare each vaccine group with placebo for immunogenicity and safety outcomes and adjust for age, gender, vaccination center, health care worker or job involving close contact with EVD cases, baseline laboratory abnormalities, HIV status, and nutritional status as determined by body mass index will also be carried out with regression.

#### 7.3. Independent DSMB

The statistical analysis plan will be reviewed with the independent DSMB prior to beginning the study and after any changes are made to it. It will be updated by the investigators prior to unblinding the data or if a correlate of protection is found.

The DSMB closely monitors accumulating safety and immunogenicity data for adults and children in each age group (1-4, 5-11, and 12-17 years). The DSMB reviews data for participants enrolled under Versions 2.0, 3.0 and 4.0.

After 70 children aged ≥12 to ≤17 years have been enrolled (20 vaccinated with Ad26.ZEBOV, 30 vaccinated with rVSVΔG-ZEBOV-GP, and 20 vaccinated with placebo) and followed for 28 days, the DSMB were asked to consider whether vaccination of children ≥5 to ≤11 years can commence. The decision to begin enrolling younger children was based on the severity of AEs experienced and the likely relationship of AE to a vaccine, the frequency of adverse effects, and whether the AEs were time-limited.

If judged safe to enrol younger children, 70 children aged ≥5 to ≤11 years were enrolled and after they have been followed for 28 days the DSMB was asked to consider whether vaccination of children at least 1 year of age to ≤ 4 years could commence. The decision to begin enrolling children at least 1 year of age to ≤ 4 years was based on AE severity, frequency and time-limitedness.

Once the decision was made that the enrolment of children 1 to 4 years was safe, the enrolment target of the 1,400 children was reached to achieve a similar distribution in the 3 age groups.

Safety data for children are reviewed at least monthly until the DSMB is satisfied that the vaccines are safe at which time less frequent reviews may be carried out.

The DSMB is immediately notified of SAEs considered vaccine-related. The DSMB may pause enrolment in the event of vaccine-related deaths or SAEs that are considered vaccine-related. The DSMB may also pause enrolment or request that participants be notified if there is an increased frequency of unanticipated adverse effects.

In addition to monitoring safety and immunogenicity outcomes, the DSMB will also review the completeness of follow-up and other aspects of study conduct.

After each meeting the DSMB will recommend continuing the study as planned, modifying the study, or terminating the study.

There is no plan to terminate the trial early based on the immunogenicity findings.

Any DSMB meeting after all M12 visits completed will be left to the discretion of the DSMB itself.

##### 7.4. Steering Committee

The trial will be conducted under the direction of a steering committee. Members of the steering committee will be blinded to interim safety and immunogenicity results. The steering committee monitors the enrolment and follow-up of participants, including the collection of AEs and blood for immunogenicity testing. The steering committee may request that more men or women or participants in specific age groups be enrolled to ensure broad generalizability of the findings.

The steering committee is also responsible for identifying new information for participants that arises during the trial that may relate to the willingness of participants to continue participating in the trial. Members of the steering committee will meet with the DSMB during an open session to discuss study conduct.

#### 8. PROTECTION OF HUMAN SUBJECTS AND OTHER ETHICAL CONSIDERATIONS

##### 8.1. Local review of protocol and informed consent

Prior to initiation, the protocol, the informed consent and assent forms, and participant information materials were submitted to and approved by the INSERM IRB/ EC; and LSHTM EC; and University of Maryland EC; and the ECs in Guinea, Liberia, Mali, and Sierra Leone. Any future amendments to the study protocol, consent, or participant materials will likewise be submitted and approved by these ECs.

##### 8.2. Ethical conduct of the study

The trial are conducted according to the Declaration of Helsinki in its current version and any subsequent revisions; US regulations governing protection of human subjects such as 21 US Code of Federal Regulations (CFR) 50; the requirements of Good Clinical Practice (GCP) as defined in published guidelines; the US Office for Human Research Protections; and with local law and regulation, whichever affords greater protection of human subjects.

A team of social anthropologists will undertake a longitudinal survey to collect the experiences of both participants and health professionals within the trials. They will inform in real time the investigators of issues that may be expressed by the persons carrying out the field work.

##### 8.3. Informed consent of study participants

Participating adults must sign/mark the appropriate approved informed consent form prior to any study-related procedures being conducted. This includes provision of consent for the long term follow-up. Informed consent for children will be given by one of the parents or legal guardian of the child. A video and/or picture booklets that describe the study may be used to ensure that illiterate volunteers understand the study requirements and risks and benefits. A witness will document that subjects who are illiterate reviewed the booklet/video and were provided the opportunity to ask questions about the consent information. Details of consent and assent procedures for minors who are able to provide assent will be described in the manual of operations.

Minors who decline participation in the study after reviewing the assent materials will not be enrolled even if their parent(s) or legal guardian consent to their participation.

During the long-term follow-up and in accordance with protocol version 6.0 and version 7.0, the participants who received the placebo or incomplete Ad26.ZEBOV/MVA-BN-Filo vaccine strategy will consent/assent before being offered the new vaccination.

Community mobilization efforts will help identify local stakeholders to ensure that the informed consent process is locally acceptable and takes into account background health, scientific and basic literacy levels, and is effective.

#### 8.4. Confidentiality of study participants

The confidentiality of all study participants will be protected in accordance with GCP Guidelines and national regulations.

Data generated as result of this study are anonymized and are to be available for inspection on request by IRBs and the regulatory health authorities, including external site auditors and inspectors. For example, monitors may visit the clinical research sites to verify the existence of signed informed consent documents and the prompt and accurate recording of SAEs. The monitors may also inspect the clinical site regulatory files to ensure applicable guidelines are being followed.

#### 8.5. Compensation

Participants in the study will be compensated for their time and inconvenience. Compensation will be provided at each regularly scheduled study visit/contact. The governments of each country will recommend the amount of compensation to be provided.

#### 8.6. Dissemination of trial results

Participants in each study and representatives of the community will be kept informed of enrolment and follow-up statistics. When the trial has been completed, the results will be shared with participants and community representatives.

#### 8.7. Risk/Benefit Assessment

The sponsors evaluated the risks of the respective interventions to the benefit from enrollment in the study. In accordance with US regulation 45CFR46, minimal risk is defined as the probability and magnitude of harm or discomfort anticipated in the research are not greater in and of themselves than those ordinarily encountered in daily life or during the performance of routine physical or psychological examinations or tests. As such, consistent with the administration of any vaccine, approved or investigational, and considering the safety profile of the study vaccines described in sections 1.1.2 and 1.1.3, the study intervention represents greater than minimal risk to both adults and children. The direct benefit to both adults and children having received the study vaccines result from the immunogenic response observed from the study vaccines in previous studies. Evidence indicates a reservoir of Ebola remains in West Africa that is capable of generating sporadic outbreaks in the near future.[55] The immunogenic response generated by these vaccines has the potential to protect vaccinated individuals from Ebola acquisition. The rVSVΔG-ZEBOV-GP and Ad26.ZEBOV/MVA-BN-Filo vaccines are now licensed. Additionally, access to healthcare is limited in West Africa.[56] During the 2015 PREVAIL I study, participants included in the study were provided laboratory testing not available through the public health system resulting in the identification of a number of underlying conditions that allowed individuals to enter the health care system for treatment.[52] For PREVAC, the baseline and continued follow-up of volunteers will provide the opportunity for early identification of unrecognized illnesses and referral to the health care system for treatment.

The sponsors believe the research in adults is approvable as research involving greater than minimal risk to subjects but with a prospect of direct benefit to the individual subjects. In children, the investigators believe the research is approvable under Subpart D 45 CFR 46.405 as the risk is greater than minimal risk but may provide prospect of direct benefit. This request is based on evidence from a Guinean study reporting that children, especially those under age 5, were particularly vulnerable and highly susceptible to death.[57] In their paper describing the impact of Ebola on children in Guinea, the authors found an overall case fatality rate of 62.9% for children between 1 month and 20 years. For those between one month and five years, the case fatality was 82.9%. The WHO statistics for Guinea for all patients in the 2014-2016 outbreak was 67%.[8] Because 45 CFR 46.408 requires that the consent of both parents be obtained unless there is the potential for benefit, the sponsors have identified that there is a potential for benefit due to the vulnerability of children to Ebola and therefore request under provisions permitted under 45 CFR 46.408(b) that consent from one parent to enroll a child be sufficient.

### APPENDIX A

#### Time and Events Schedule for Baseline and Regularly Scheduled Follow-up Visits

| Events | Adults | Children |
|---|---|---|
| **Screening** (children must have a documented negative HIV test prior to randomization) | | X |
| **Baseline** | | |
| Informed consent/assent | X | X |
| Initial vaccination | X | X |
| Demographics [a)] | X | X |
| Contact information [b)] | X | X |
| Indicators of increased risk [c)] | X | X |
| Height, weight and temperature | X | X |
| Mid-upper arm circumference for children 1-5 years | | X |
| Pregnancy test for females of child-bearing potential, i.e., females who have experienced menarche or who are aged 14 years and older | X | X |
| Blood sample for immunogenicity testing and future research | X | X |
| Blood sample for chemistries and CBC | X | X |
| Blood sample for HIV/syphilis testing | X | X |
| Saliva sample for assessing viral shedding (subsample) | | X |
| Stored blood for assessing T cell and memory B cell function in a subset of participants in Guinea | X | |
| Injection site reactions, targeted symptoms of any grade severity, and grade 3 or 4 AEs following prime vaccination | X | X |
| HIV pre-counseling | X | X |
| **Days 1 to 6 following prime and booster vaccination** | | |
| Contact for injection site reactions, targeted symptoms of any grade severity, grade 3 or 4 AEs, measurement of temperature, and possible SAEs in children | | X |
| **Day 7 and Day 63** | | |
| Injection site reactions, targeted symptoms of any grade severity, and grade 3 or 4 AEs | X | X |
| Blood sample for immunogenicity testing and future research | X | X |
| Saliva sample for assessing viral shedding (subsample) | | X |
| Blood sample for assessing T cell and memory B cell function in a subset of adults in Guinea | X | |
| HIV and syphilis post-counseling referral | X | X |

| Events | Adults | Children |
|---|---|---|
| Blood sample for chemistries and CBC | | X |
| Temperature | X | X |
| **Day 14** | | |
| Injection site reactions, targeted symptoms of any grade severity, and grade 3 or 4 AEs | X | X |
| Temperature | X | X |
| Blood sample for immunogenicity testing and future research | X | X |
| Saliva sample for assessing viral shedding (subsample) | | X |
| Blood sample for assessing T cell and memory B Cell function in a subset of adults in Guinea | X | |
| **Day 28** | | |
| Injection site reactions, targeted symptoms of any grade severity, and grade 3 or 4 AEs | X | X |
| Temperature | X | X |
| Weight | | X |
| Blood sample for immunogenicity testing and future research | X | X |
| Saliva sample for assessing viral shedding (subsample) | | X |
| **Day 56** | | |
| Booster vaccination | X | X |
| Blood sample for immunogenicity testing and future research | X | X |
| Pregnancy test for females of childbearing potential, i.e., females who have experienced menarche or who are aged 14 years and older | X | X |
| Temperature | X | X |
| Injection site reactions, targeted symptoms of any grade severity, and grade 3 or 4 AEs following booster vaccination | X | X |
| Saliva sample for assessing viral shedding (subsample) | | X |
| Blood sample for assessing T cell and memory B cell function in a subset of adults in Guinea | X | |
| **Day 70** | | |
| Blood sample for assessing T cell and memory B cell function in a subset of adults in Guinea | X | |
| **Months 3 and 6** | | |
| Injection site reactions, targeted symptoms of any grade severity, and grade 3 or 4 AEs (Month 3 only) | X | X |
| Temperature | X | X |
| Weight | | X |

| **Events** | **Adults** | **Children** |
|---|---|---|
| Mid-upper arm circumference for children 1-5 years | | X |
| Blood sample for immunogenicity testing and future research | X | X |
| Saliva sample for assessing viral shedding (subsample at Month 3 only) | | X |
| **Months 12, 24, 36, 48, and 60** | | |
| Temperature | X | X |
| Weight | | X |
| Height | | X |
| Mid-upper arm circumference for children 1-5 years | | X |
| Blood sample for immunogenicity testing and future research | X | X |
| Survey data for the study on barriers and facilitators to PREVAC retention | X | X |
| Blood sample for the parasitology assays for the malaria substudy in Sierra Leone and in Guinea (Landreah site) | X | X |
| Blood sample for assessing T cell and memory B cell function in a subset of adults in Guinea | X | |
| **New vaccination visit during the long-term follow-up for participants who received the placebo or incomplete rHAd26/MVA vaccine strategy and who are going to be vaccinated during the long term follow-up**<br>(During a long-term follow-up visit or at an additional visit) | | |
| Informed consent/assent | X | X |
| Temperature | X | X |
| Pregnancy test for females of child-bearing potential, i.e., females who have experienced menarche or who are aged 14 years and older | X | X |
| New vaccination | X | X |
| **SAEs throughout follow-up** | X | X |
| **EVD events (reported as soon as aware) throughout follow-up and where possible a stored blood sample for future research** | X | X |
| **Death (reported as soon as aware) throughout follow-up** | X | X |
| **Pregnancy outcome for women who become pregnant in the first three months of follow-up after prime vaccination and in the first three months of follow-up after the new vaccination for participants who received the placebo or incomplete Ad26.ZEBOV/MVA-BN-Filo vaccine strategy** | X | X |
| **Recording on COVID19 or Ebola vaccination outside of the PREVAC protocol** | X | X |

a) Birth month and year, gender
b) Contact information for self and 2 contacts who will know how to locate the volunteer
c) Role as health care worker, contact with persons known to have EVD

### APPENDIX B

#### Reference Documents on PREVAC Study Website

The PREVAC website will maintain updated links to the following documents referenced in the PREVAC protocol and to other information pertinent to the study:

- PREVAC Procedures
- Pharmacy Manual of Operations
- Investigator's Brochure for each vaccine
- The Division of AIDS (DAIDS) Table for Grading the Severity of Adult and Pediatric Adverse Events, Version 2.0, November 2014.

### APPENDIX C

### Site Investigators and Ethics Review Boards

**Site Investigators:**

| | |
|---|---|
| Abdoul Habib Beavogui | Guinea (Landreah and Maferinyah) |
| Cheick Mohamed KEITA | Guinea (Landreah) |
| Pascal Niouma Millimouno | Guinea (Maferinyah) |
| Mark Kieh | Liberia |
| Stephen B. Kennedy | Liberia |
| Bailah Leigh | Sierra Leone |
| Mohamed H. Samai | Sierra Leone |
| Gibrilla Fadlu Deen | Sierra Leone |
| Samba Sow | Mali Center for Vaccine Development |
| Milagritos Tapia | Mali Center for Vaccine Development |
| Seydou Doumbia | Mali University Clinical Research Center |
| Mahamadou Diakite | Mali University Clinical Research Center |

**Ethics Review Boards:**

INSERM IRB (IRB00003888 – FWA00005831)
Paris, France

LSHTM Research EC
London, United Kingdom

National Research Ethics Board (NREB)
Monrovia, Liberia

Comité National d’Ethique pour la Recherche en Santé (CNERS)
Conakry, Guinea

Sierra Leone Ethics and Scientific Review Committee (SLESRC)
Freetown, Sierra Leone

Faculty of Medicine, Pharmacy, and Odonto-Stomatology (FMPOS) and IRB of the University of Maryland, Mali

**Regulatory Sponsor in Liberia:**

Project PREVAIL will be the Regulatory Sponsor In Liberia to the Liberian Medicines and Health Products Regulatory Authority (LMHRA).

INSERM has entered into an institutional authorization agreement with the NIH to allow the NIH to rely upon the ethical review of the INSERM EC. The INSERM EC will conduct initial and annual continuing review of the research in accordance with the requirements identified in the Federal-Wide Assurance granted by the Office for Human Research Protections of the US Department of Health and Human Services.

### APPENDIX D

### PREVAC Immunological Substudy Protocol

##### (To Be Performed in Guinea Only)

Mondor Immunomonitoring Center, MIC (C. Lacabaratz)/Vaccine Research Institute, VRI (Head: Pr. Y. Lévy)

#### 1. Rationale

A variety of vaccine platforms against Ebola have been generated in mice, guinea pigs and NHPs. Different types of vaccines against EBOV have been proposed in clinical trials, including in majority recombinant vaccines expressing the EBOV surface GP. But question regarding their efficiency remains unanswered.

Among these vaccines, the Ad26.ZEBOV vaccine is used in prime boost strategy in combination with MVA-BN-Filo in phase I study: EBOVAC1 trial is actually conducted in the United Kingdom (UK), the US and African countries to assess the safety and immunogenicity of the candidate prime-boost regimen in healthy volunteers. Enrolment of the first human trial started in December 2015, preliminary post-prime safety and immunogenicity data are expected in March 2015 and the full safety and humoral/cellular immunogenicity data in May/June 2015. Phase II trials (EBOVAC2) are actually ongoing in healthy volunteers in Europe (France and UK) and non-epidemic African countries.

Alternatively, some data from an open-label, cluster randomized ring vaccination trial conducted in Guinea, showed that the rVSVΔG-ZEBOV-GP vaccine prevented EVD events occurring at least 10 days after randomization in close contacts who were immediately vaccinated, but duration of the vaccine effect was not assessed. Two other phase 1 dose-escalation trials using rVSVΔG-ZEBOV-GP vaccine were conducted in US. No short-term safety concerns were identified after a single administration of the rVSVΔG-ZEBOV-GP vaccine, and anti-Ebola humoral responses (titers of antibodies against ZEBOV GP assessed by ELISA) were identified in all the volunteers 28 days after vaccination.

Up to now, very few data on the quality of cellular immune responses induced by the vaccines and no data on their middle/long-term duration are available. Moreover, no comparison of immunogenicity profiles between different vaccine strategies with and without a booster has been assessed.

#### 2. Objectives

The objective of the immunological sub-study is to analyze the immunogenicity of the 2 prophylactic vaccine candidates (rVSVΔG-ZEBOV-GP from Merck and rHAd26/MVA from Janssen) used in the PREVAC study in a population at risk of EBOV infection. We will focus on T cell responses induced by the vaccines in adult population and on their middle and long term persistence until 5 years after vaccination.

This hypothesis will be addressed with both the diluted (Version 3.0) and undiluted (Version 4.0) rVSVΔG-ZEBOV-GP vaccine.

#### 3. Endpoints

In addition to the analysis of humoral response (neutralizing antibodies and antibodies mediating antibody-dependent cellular cytotoxicity), analysis of the immunogenicity of the study vaccine regimens will include the characterization of specific cellular response. The quality, the precocity as well as the durability of the T cell responses induced by the different vaccines will be raised, and we can propose the following assays.

The primary endpoints will focus on quantitative and qualitative aspects of specific T cell response.

- Activation of cluster of differentiation CD4+ and CD8+ T cell subsets and their cytokine expression patterns will be determined by flow cytometry after EBOV-specific stimulation (including, but not limited to, IFNγ, interleukin IL2, and tumor necrosis factor alpha (TNF-alpha) using intracellular cytokine staining (ICS) before (D0), 14 days after each vaccination (D14 and D70) and at M12, M24, M36, M48 and M60. The polyfunctionality of specific CD4+ and CD8+ T cells (capacity to produce more than 1 cytokine) will be analyzed as well.

The secondary endpoints will include:

- Phenotypic analysis of B and T cell subsets before (D0), 14 days after vaccination (D14 and 70) and M12, M24, M36, M48 and M60. The frequency of B (naive, memory, switched, plasma cells) and T cell subsets (naive, memory, effector, cytotoxic capacity and T follicular helper [Tfh] cells) will be assessed by flow cytometry combining a panel of different markers: e.g. CD19, CD21, CD27, CD38, immunoglobulin (Ig) D, IgM and CD3, CD4, CD8, CD45RA, CC-chemokine receptor 7 (CCR7), CD27, Perforin, Granzyme B, CXCR5, programmed cell death protein 1 (PD-1), respectively for B and T cell panels.

- Measurement of serum cytokine/chemokine levels (inflammation/activation markers) before (D0) and after vaccination (D7 and D63) using Luminex technology allowing detection up to 30 analytes in a small sample volume.

- Ex vivo gene expression profile in whole blood before and 3h after each vaccination (D0, D56), D7 and D63 to evaluate gene abundance by microarray in order to find the components of the vaccine signature and to compare them between Ebola vaccines. Highly sensitive and quantitative assays were developed to process samples including quality controls at each step of processing, from sampling to the results report. The technology used is based on the BeadChip Illumina platform which consists of the direct hybridization assay that offers the highest level of multiplexing for whole-genome expression profiling available, with the most up-to-date expression content and high-throughput processing. Each array on the HumanHT-12 v4 Expression BeadChip targets more than 34,000 annotated genes.

#### 4. Sub-study eligibility criteria

Adult participants from Guinea
Participation approval in immunological sub-study (additional blood draws, specific consent)

#### 5. Sub-study components and procedures

For the evaluation of the different immunogenicity endpoints, samples will be collected at the time points and in volumes as indicated in the Table below:

<u>Blood volumes (ml)</u>

| Type of tube | Assays | D0 Prime | D7 | D14 | D56 Boost | D63 | D70 | M12,24, 36, 48, 60 |
|---|---|---|---|---|---|---|---|---|
| EDTA (PBMC) | ICS | 24 | | 24 | | | 24 | 24 |
| | B / T phenotype | 6 | | 6 | | | 6 | 6 |
| Dry tube (serum) | Luminex | 4 | 4 | | | 4 | | |
| Tempus (RNA) | Gene profile | 6* | 3 | | 6* | 3 | | |
| | **Blood total vol.** | **40** | **7** | **30** | **6** | **7** | **30** | **30** |

*: 3 ml before and 3 hours after vaccine injection

<u>Number of tubes</u>

| Type of tube | D0 Prime | D7 | D14 | D56 Boost | D63 | D70 | M12, 24, 36, 48, 60 |
|---|---|---|---|---|---|---|---|
| EDTA<br>7 ml tube | 4 | | 4 | | | 4 | 4 |
| Dry<br>5 ml tube | 1 | 1 | | | 1 | | |
| Tempus | 2* | 1 | | 2* | 1 | | |

*: 1 tube before and 3 hours after vaccine injection

**6. Sample size calculation**

According to methodologist’s calculation respecting allocation ratio of randomization 2:1:2:1:1, 230 participants were enrolled in the substudy under Versions 2.0, 3.0, and 4.0 (in total).

Separate analyses will be carried out for participants enrolled under Versions 3.0 and 4.0.

**7. Sites**

Whole blood samples for this sub-study will be collected at Conakry (Guinea) site on 3 different types of tubes (tubes without anticoagulant, EDTA and Tempus$^{TM}$ Blood RNA tubes), and then:

- sent immediately to the immunological sites (2 "selected" sites in Conakry for PBMC isolation: to Landréah and Institut National de Santé Publique from D0 to M12, and to Landréah from M24 to M60)

- processed at the immunological sites according to current versions of approved Standard Operating Procedure,

- and stored according to a specific procedure before shipping to Mondor Immunomonitoring Platform, Créteil, France.

Different Standard Operating Procedures will contain all details regarding the collection, handling, labeling, and shipment of blood samples to the laboratory in charge of immunomonitoring.

### APPENDIX E

### PREVAC Saliva Sample Substudy

##### (To Be Performed in Liberia Only)

#### Rationale

This substudy of viral shedding of rVSVΔG-ZEBOV-GP is being conducted to estimate the proportion of children who have detectable vaccine virus by RT-PCR and to quantify vaccine virus levels shed after a prime and a boost vaccine dose. In Phase 1 studies that collected specimens to monitor shedding of rVSVΔG-ZEBOV-GP, the largest proportion of adult subjects that had vaccine virus detected in saliva or urine was 5/20 in $2x10^7$ pfu North American recipients with positive saliva on Day 7. This decreased to 1 of 10 on Day 14.[43] Preliminary open-label data in African adults, adolescents between 13 and 17 years of age and children between 6 and 12 years of age suggests that shedding after vaccination is higher in children and adolescents compared to adults. While no shedding was detected in adults, 14/18 adolescents and 7/20 children had positive saliva on Day 7. Shedding in children beyond Day 7 has not been assessed to date and will be assessed in this substudy.

#### Objective

The objective of the saliva substudy is to estimate the proportion of children who shed vaccine virus and to quantify the rVSVΔG-ZEBOV-GP vaccine shed in children (participants aged < 18 years) after the prime and boost vaccinations. For estimates on days 7, 14, 28, and 56 (prior to booster), the two rVSVΔG-ZEBOV-GP groups will be pooled. On day 63 and at 3 months (after the booster), the two rVSVΔG-ZEBOV-GP groups will be considered separately. Our hypothesis is that viral shedding will be greatest 7 days after vaccination (prime or booster) and will decline afterwards.

This hypothesis will be addressed with both the diluted (Version 3.0) and undiluted (Version 4.0) rVSVΔG-ZEBOV-GP vaccine.

#### Procedures and Measurements

At a single vaccination center, approximately 0.5-1.0 mL of saliva were collected from children for whom informed consent was obtained at baseline (prior to prime vaccination), days 7, 14, 28, 56 (prior to booster), and 63 (7 days following booster), and at 3 months (approximately 35 days after booster). The saliva was divided into at least 2 x 200 µL aliquots in RNA microfuge tubes. 600 µL of Trizol LS or equivalent transport media was added to each aliquot, the tubes were sealed, mixed and stored at -70° C or lower, prior to shipment for quantitative RT-PCR (RT-qPCR) testing to determine levels of virus present in the samples (threshold value will be determined based on assay characteristics). Testing will be performed on all rVSVΔG-ZEBOV-GP vaccine and placebo group substudy participants blinded to vaccine group.

**Statistical Considerations**

126 children were enrolled in the substudy under protocol Version 3.0 and 139 children were enrolled under protocol Version 4.0.

With approximately 140 children enrolled from the 5 randomized groups, there will be approximately 60 participants who are given the rVSVΔG-ZEBOV-GP vaccine as prime and 40 participants in the pooled placebo group. Approximately 40 participants who are enrolled in the substudy would receive the rHAd26 /MVA vaccine,  The 95% confidence interval half-widths are presented for several percentages for 60 children receiving the rVSVΔG-ZEBOV-GP prime vaccine: 10% ±8%, 30% ±12% and 50% ±13%.

There will be approximately 20 participants  who are given the rVSVΔG-ZEBOV-GP prime vaccine and booster vaccine at 56 days.  40 participants will receive a placebo booster. The 95% confidence interval half-widths are presented for several percentages receiving the rVSVΔG-ZEBOV-GP prime and booster vaccine for 20 children: 10% ±13%, 25% ±19%, and 30% ±20%.

Counts and percentages of children with vaccine virus detected in saliva samples as well as mean, and medians ($25^{th}$, $75^{th}$ percentiles) for virus levels (copies/mL) will be presented for days 7, 14, 28, 56 (prior to the booster vaccination) and for day 63 and 3 months.  For the first four time points, the rVSVΔG-ZEBOV-GP prime vaccine with and without the rVSVΔG-ZEBOV-GP boost will be pooled; for the last 2 time points (after the booster vaccination), these groups will be considered separately.

Separate analyses will be carried out for participants enrolled under Versions 3.0 and 4.0.

### APPENDIX F

### MALARIA AND HELMINTH INFECTIONS SUBSTUDY

### (To Be Performed in Sierra Leone and Guinea)

### STUDY OF THE IMPACT OF EXPOSURE TO MALARIA AND HELMINTH INFECTIONS ON THE DURABILITY OF THE IMMUNE RESPONSE TO TWO EBOLA VACCINES

#### RATIONALE

There is good evidence that the presence of malaria and some helminth infections at the time of vaccination reduces the immune response to some vaccines.and increasing evidence that exposure to malaria interferes with B cell memory.[58,59,60,61,62,63] However, the influence of malaria and other parasitic infections on the duration of the antibody response to vaccination has been little studied. This component of the long term follow-up of the PREVAC study will investigate whether sustained exposure to malaria and helminth infections influences the duration of the antibody response to immunization with J&J and Merck Ebola vaccines .

#### STUDY OBJECTIVE

The objective of the study is to determine whether the rate of decline in antibody titre to the EBOLA virus glycoprotein induced by immunisation with the J&J or Merck Ebola vaccine is influenced by exposure to malaria or helminth infections, measured by serology, over a period of 4-5 years.

#### STUDY OUTLINE

Seven hundred and eight volunteers over the age of one year (397 adults and 311 children) have been recruited to the PREVAC study at the Mambolo site in Sierra Leone and vaccinated with either the rVSVΔG-ZEBOV-GP Ebola vaccine, two dose Ad26.ZEBOV/ MVA-BN-Filo vaccine or a placebo and are being followed for 12 months after vaccination. On completion of the PREVAC study, volunteers in the PREVAC study will be invited to join a long-term follow-up study, which will investigate the longer term course of the immune response to immunisation with an Ebola vaccine. Participants who agree to join the long-term follow-up study, will be invited to attend the clinic approximately 24, 36, 48 and 60 months after vaccination and asked about any serious illnesses during the preceding year.

Serological tests will be used to measure exposure to malaria and helminth infections as these assays provide a more reliable measure of exposure than occasional, point-prevalence cross-sectional surveys. There is already strong evidence that malaria is highly prevalent in Mambolo but less is known about the prevalence of other parasites as the village has been involved in recent years in several mass drug administration programs with ivermectin and albedazole. Thus, an initial, age-stratified cross-sectional survey will be undertaken in 1,000 villagers during which blood, stool and urine samples will be collected and risk factors for malaria and helminth infections recorded. Blood samples will tested for malaria by microscopy and PCR and for filariasis by a rapid diagnostic test. Urine samples will be tested for schistosomiasis by microscopy and a rapid diagnostic test and stool samples examined for intestinal helminths using the Kato-Katz technique. Based on the results of these findings a multiplex bead assay will be developed which contains malaria antigens that detect antibodies associated with recent or past exposure to malaria and helminth antigens whose selection will be influenced by the initial parasitological survey. Assays will be undertaken either by the London School of Hygiene and Tropical Medicine either in London or in The Gambia.

Base-line samples, samples collected at the initial 12 months' follow-up, available in the PREVAC biobank, and samples collected at each subsequent annual follow-up survey will be tested for antibodies to malaria and helminth infections using the multiplex assay. Currently, only samples from Guinea are used to study the cell mediated immune responses to the Ebola vaccine. Hence, the serological assays for malaria and helminth infection will also be performed on the samples from the Guinean participants participating in the immunological substudy, thus enabling to investigate the impact of exposure to malaria and helminth infections on this component of the immune response to Ebola vaccines.

**ENDPOINT**

The rate of decline in antibody titre to the Ebola virus glycoprotein will be determined for each individual during the period of follow-up and compared with the level of exposure to malaria and helminth infections. A detailed analytical plan as to how correlations will be determined will be developed following analysis of the initial base-line samples.

### APPENDIX G

### EMPIRICAL RESEARCH ON BARRIERS AND FACILITATORS TO STUDY RETENTION IN THE PREVAC TRIAL

#### 1. RATIONALE

This sub-study aims to identify multi-level determinants of participant retention in the PREVAC trial to develop recommendations that will benefit future clinical research in resource limited settings affected by Ebola and other infectious diseases.

Early clinical trials testing Ebola virus disease vaccine strategies have achieved high rates of participant retention under highly challenging conditions. For example, the PREVAIL I Ebola vaccine trial was conducted during a public health emergency in Liberia and achieved an exceptionally high level of participant retention, with 97.8% of the participants retained through 12 months of follow-up. Although many factors may have contributed to this achievement, including disease acuity, study tracking and remuneration procedures, and community-level social mobilization activities[64], no systematic data collection or analysis has been conducted to provide an empirical understanding of this success. Similarly, the Sierra Leone Trial to Introduce a Vaccine Against Ebola (STRIVE) conducted with adult health care workers retained 92.5% of participants following enrolment, although lower numbers of participants completed every appointment visit. Lower retention was observed among older participants and frontline healthcare workers[65].

The PREVAC randomized trial will evaluate the safety, immunogenicity, and durability of three Ebola vaccine strategies for adults and children at 12 months and annually thereafter for up to 5 years. The PREVAC trial will joins an expanding set of Ebola vaccine trials and studies in the PREVAC and PREVAIL partnerships which involve long-term follow-up with research participants over 12 months or longer. The extended follow-up periods on these trials can present challenges for maintaining participant retention and preventing loss-to-follow-up. The exceptionally long duration of follow-up in PREVAC presents an important opportunity to advance scientific understanding of factors related to participant retention in Ebola vaccine clinical trials, and clinical trials conducted in East Africa more generally.

This sub-study proposes to leverage PREVAC trial operations to conduct a mixed-methods sub-study (qualitative and quantitative methods) which is designed to better understand multi-level predictors of participant retention and loss-to-follow-up (LTFU) in the trial. Advancing a scientific understanding of multi-level influences on participant retention in PREVAC could strengthen the design and rigor of future clinical trials conducted within resource limited settings affected by Ebola and emerging infectious diseases.

#### 2. OBJECTIVES

Objective 1: Identify multilevel facilitators and barriers to participant retention in PREVAC through retrospective qualitative research with participants after the conclusion of the blinded trial period (first 12 months).

Hypothesis: Both individual and site-level factors foster participant retention in PREVAC (e.g., individual participant age, motivation, and beliefs about arm assignment; as well as site-level renumeration, tracking, and community mobilization procedures)

Objective 2: Identify factors that predict participant retention, loss-to-follow-up (LTFU), and timeliness of appointment attendance during the unblinded PREVAC long-term follow-up period (from 12 months up to 5 years) through prospective research.

Hypothesis: Both individual and site-level factors will longitudinally predict participant retention and loss-to-follow-up during the PREVAC long-term follow-up period.

#### 3. ENDPOINTS

The primary endpoints for the sub-study will be:

- Retention (defined as the percentage of study appointments kept) during:
  - the blinded PREVAC trial period (first 12 months)
  - the unblinded PREVAC long-term follow-up period (12 months up to 5 years)

- Participants lost-to-follow-up (never returning) over the unblinded PREVAC long-term follow-up period (12 months up to 5 years)

- Timeliness of study appointment attendance (number of days between study appointment target date and appointment completion date) across the PREVAC long-term follow-up period

The primary endpoints will be examined in relation to the following trial process measures:

- Perceived barriers/facilitators to study retention by trial participants and trial staff, as identified through qualitative interviews and focus groups.

- Individual participant factors in four domains hypothesized to affect study retention: demographic factors, trial-related perceptions, Ebola-related perceptions, and other psychosocial factors. These factors will be quantified through brief survey questions administered to trial participants.

- Site-level procedures to support participant retention, including appointment visit incentives, participant tracking, digital and community-based outreach, and use of community champions and community mobilization techniques. These site-level factors will be documented via a survey completed by site coordinators.

#### 4. SUB-STUDY ELIGIBILITY CRITERIA

Objective 1: Adult participants enrolled in sub-study participating PREVAC sites who demonstrate perfect vs. imperfect visit attendance during their first 12 months on study and who are not known to have died; also site/study staff and community engagement/outreach personnel.

Objective 2: Participants enrolled in PREVAC who enter long-term follow-up. Participants will include all adult and adolescent (12-17) PREVAC enrolees. Participation of any younger individuals is at the discretion of sites.

#### 5. SUB-STUDY COMPONENTS AND PROCEDURES

<u>Objective 1 methods</u>: This will be a retrospective research study conducted at two or more PREVAC sites which collects two forms of qualitative data:

1. Semi-structured qualitative interviews regarding barriers/facilitators to study retention will be conducted with individual adult PREVAC participants after 12 months on study, via a case-control design.

- <u>Sample</u>: Each participating site will enroll a sample of up to 20 adult participants with perfect retention and up to 20 adult participants with imperfect retention in the first 12 months of PREVAC.

- <u>Recruitment</u>: Participating sites will generate a list of eligible participants in each category. Candidates for the qualitative interviews will be randomly selected from the list. Site staff will approach the candidates during PREVAC-related study appointments, mobile phone contact, or community outreach to request their participation in the interview sub-study.

- <u>Procedure</u>: Those who agree to participate will be consented and will complete a brief, in-person, semi-structured qualitative interview regarding factors that assisted or impeded their study appointment attendance. To facilitate recall, the interviews will be conducted with participants as close to their 12-month completion date as possible. The interviews will be conducted in a private space by social scientists or staff members affiliated with PREVAC or the local sites. All interviewers will receive training and certification on the interview protocol from NIAID/NIH. Interviewers will record brief notes on each interview, and all interviews will be audiotaped, with the consent of the participant. The recorded interviews will be transcribed and translated for thematic analysis.

2. Focus groups with PREVAC site/study staff and community engagement/outreach personnel regarding barriers/facilitators to study retention.

- Sample: Each participating site will convene up to 3 focus groups with site/study staff and community engagement/outreach personnel after the site has completed the initial 12-month follow-up period with all PREVAC participants. Focus group participants may include PREVAC participant trackers, community mobilizers, community health workers, site/study staff, and investigators/co-investigators. Each focus group would have approximately 6-8 participating individuals.

- Recruitment: Participating sites will circulate an announcement about the focus groups to relevant team members. It will indicate that focus group participation is voluntary and not a condition of employment, and that individuals are free to decline participation. Interested individuals will be scheduled for participation in a focus group.

- Procedure: The focus groups will explore site/staff perspectives on factors which facilitated or impeded PREVAC participant retention, as well as the feasibility and impact of site retention support procedures and community engagement activities. The focus groups will be conducted in a private space by social scientists or staff members affiliated with PREVAC or the local sites. All focus group facilitators will receive training and certification on the focus group protocol from NIAID/NIH. A separate staff member will record notes on the focus group, and the focus groups will be audiotaped. The recorded focus groups will be transcribed and translated for thematic analysis.

Thematic analysis of the qualitative data will be overseen by NIAID/NIH. For the qualitative interviews, notes and transcripts from five interviews will be reviewed to prepare analytic memos describing initial themes regarding retention facilitators/barriers, and to develop an initial qualitative data codebook. The initial themes and codes will be applied to another five interviews and edited as necessary; this process will continue in an iterative fashion until the codes are both cohesive and distinct. The finalized codebook will be applied to all interview transcripts using qualitative analysis software. The focus group data will receive a similar analytic approach. Integrative analyses of all qualitative data would subsequently identify multilevel facilitators and barriers to participant retention in PREVAC during the blinded trial period (first 12 months), and they would compare/contrast the most salient factors identified by each data source and participating site.

Objective 2 methods: This would be a prospective research study conducted at all PREVAC sites using two forms of quantitative survey data:

1. Quantitative survey data from PREVAC participants.

- Sample: The quantitative survey assessment would be conducted with participants who enter long-term follow-up.

- Recruitment: There is no separate recruitment or consent procedure; the survey assessment is integrated into the consent form and administered alongside the existing CRF.

- Procedures: A quantitative survey (in the form of a CRF) will be developed to provide a quantitative assessment of factors hypothesized to influence trial retention. The survey will be developed in partnership with the PREVAC investigators/co-investigators and social scientists. It will be designed to be brief (~5-10 minutes to administer) and would gather data on domains including:

    - (1) additional demographic information (e.g., educational level, income, marital and parent/guardian status);
    - (2) individual trial-related information and perceptions (e.g., satisfaction with trial, disclosure of trial participation to others, transport time to site);
    - (3) Ebola-related perceptions (e.g., personal experience with Ebola, Ebola risk perception, perceived Ebola stigma);
    - (4) psychosocial factors (e.g., social support; brief measures of anxiety and depression symptoms).

  The questionnaire under the CRF format would be administered to participants by study staff during annual study appointments. Where indicated the assessment may be administered to a parent or caregiver of a youth participant. This survey data would be combined with existing data on PREVAC participants including age, gender, trial arm assignment, and site for statistical analysis.

2. Site-level data on PREVAC sites.

- Sample: Participating PREVAC sites.

- Recruitment: None.

- Procedures: A checklist will be developed to assess current site-level strategies to support participant retention. It would gather regarding key strategies (e.g., participant tracking, digital and community-based outreach, community champions and community mobilization). It would additionally assess amount of incentives that participants receive for appointment attendance. This site-level form would be completed by the site coordinator when the site commences the long-term follow-up and annually thereafter.

Statistical analysis for the quantitative survey data will be performed by NIAID/NIH. The analysis would identify longitudinal determinants of participant retention and LTFU over the long-term PREVAC follow-up period. The potential to conduct hierarchical linear models which nest individual participant factors within site-level factors will be explored.

#### 6. SAMPLE SIZE CALCULATION

For the Objective 1 qualitative interviews with PREVAC participants, the sample size at each site participating in this sub-study will be up to 20 participants with perfect visit attendance and up to 20 participants with imperfect visit attendance during their first 12 months of trial participation. We specify up to 20 interviews in each category because this number is generally understood as satisfactory for achieving thematic saturation in qualitative research. Each participating site will additionally conduct up to 3 focus groups with site/study staff and community engagement/outreach personnel; each focus group would include approximately 6-8 participating individuals.

For the Objective 2 quantitative survey research, the sample size will be all participants in the long term follow-up of PREVAC to maximize the available data and statistical power to predict long-term participant retention and LTFU.

#### 7. SITES

For Objective 1, all PREVAC sites are eligible to participate. Participation may be limited to a subset of the PREVAC sites to enhance the feasibility of the qualitative data collection and analysis.

For Objective 2, all PREVAC sites are eligible participate, thereby enabling analyses of how differences in site or country-specific retention-related practices may affect observed participant retention rates.

### APPENDIX H

### NIH Roles in the Conduct of PREVAC

This appendix is required by NIH Office of Human Subjects Research Protections in order to establish an institutional reliance agreement for NIH to rely upon the INSERM Ethical Review Committee.

NIH roles in the conduct of PREVAC are described below:

- Dr. Lane is a member of the tripartite executive committee for the trial along with Yves Levy of INSERM and Peter Piot of LSHTM. The INSERM representative chairs the executive committee.
- PREVAC will be conducted in Guinea, Mali, Sierra Leone, and Liberia with the respective partnership of INSERM, LSHTM, and NIAID. Trial execution in Liberia will be conducted under the auspices of Project PREVAIL.
- NIAID resources will be involved in supporting the clinical trial infrastructure in Liberia and Mali.
- NIAID staff will be on site to oversee the proper conduct of the research in Liberia and Mali but the actual execution of the research will be done by Liberians and Malians (e.g., preparing vaccines, administering consent, obtaining lab samples, vaccine administration, assessing vaccine reactions, completing case report forms, follow-up visits).
- NIAID staff will be involved in overseeing the clinical lab as well as the research lab at the Liberian Institute of Biomedical Research.
- NIAID funds support a contract with Leidos Biomedical who has hired staff in Liberia and also sub-contracted additional staff for the conduct of the study.
- NIAID funds support a contract with Leidos Biomedical who provides sub-contract funding to INSERM for the conduct of the study in Guinea. Specific functions in which NIAID collaborates with the other partners include:
    - Trial governance
    - Protocol development
    - Coordination with industry partners for provision of test articles
    - Data and safety management board support
    - Site development
    - Laboratory management
    - Staff training
    - Clinical trial monitoring
- NIAID is the regulatory sponsor of record for the IND to the US FDA through the DCR OCRPRO.

### APPENDIX I:

#### THROMBOTIC EVENTS TO BE REPORTED AS POTENTIAL ADVERSE EVENTS OF SPECIAL INTEREST

At the time of protocol writing, the list of thrombotic events to be reported to the sponsor as potential AESIs is provided below. Further guidance may become available on thrombotic events of interest.

- MedDRA prefer terms for large vessel thrombosis and embolism
  - Aortic embolus, aortic thrombosis, aseptic cavernous sinus thrombosis, brain stem embolism, brain stem thrombosis, carotid arterial embolus, carotid artery thrombosis, cavernous sinus thrombosis, cerebral artery thrombosis, cerebral venous sinus thrombosis, cerebral venous thrombosis, superior sagittal sinus thrombosis, transverse sinus thrombosis, mesenteric artery embolism, mesenteric artery thrombosis, mesenteric vein thrombosis, splenic artery thrombosis, splenic embolism, splenic thrombosis, thrombosis mesenteric vessel, visceral venous thrombosis, hepatic artery embolism, hepatic artery thrombosis, hepatic vein embolism, hepatic vein thrombosis, portal vein embolism, portal vein thrombosis, portosplenomesenteric venous thrombosis, splenic vein thrombosis, spontaneous heparin-induced thrombocytopenia syndrome, femoral artery embolism, iliac artery embolism, jugular vein embolism, jugular vein thrombosis, subclavian artery embolism, subclavian vein thrombosis, obstetrical pulmonary embolism, pulmonary artery thrombosis, pulmonary thrombosis, pulmonary venous thrombosis, renal artery thrombosis, renal embolism, renal vein embolism, renal vein thrombosis, brachiocephalic vein thrombosis, vena cava embolism, vena cava thrombosis, truncus coeliacus thrombosis
- MedDRA prefer terms for more common thrombotic events
  - Axillary vein thrombosis, deep vein thrombosis, pulmonary embolism, MedDRA PTs for acute myocardial infarction[66], MedDRA PTs for stroke[67]

[2] World Health Organization Ebola Virus Disease DRC External Situation Report 17, in Équateur. 2018, World Health Organization.

[3] World Health Organization Ebola Virus Disease DRC External Situation Report 40, in North Kivu. 2019, World Health Organization.

[4] https://apps.who.int/iris/bitstream/handle/10665/328264/OEW39-2329092019.pdf

[5] https://www.who.int/csr/resources/publications/ebola/ebola-ring-vaccination-results-12-april-2019.pdf

[6] https://www.who.int/fr/news/item/25-06-2020-10th-ebola-outbreak-in-the-democratic-republic-of-the-congo-declared-over-vigilance-against-flare-ups-and-support-for-survivors-must-continue

[7] https://africacdc.org/disease-outbreak/outbreak-brief-6-ebola-virus-disease-evd-outbreak/

[17] Fallah M. A cohort study of survivors of Ebola virus infection in Liberia (PREVAIL III). Conference on Retroviruses and Opportunistic Infections, 2016, Abstract 74LB, Boston, MA.

[18] Etard JF, Salious-Sow M, Leroy S, et al. Sequelae of Ebola virus disease in surviving patients in Guinea: Postebogui Cohort. Conference on Retroviruses and Opportunistic Infections, 2016, Abstract 73LB, Boston, MA.

[19] Keita A, Vidal N, Toure A, Diallo M, Magassouba N, Baize S, Mateo M, Raoul H, Mely S, Subtil F, Kpamou C, Koivogui L, Traore F, Sow M, Ayouba A, Etard JF, Delaporte E, Peeters M, PostEbogui Study Group. A 40-Month Follow-Up of Ebola Virus Disease Survivors in Guinea (PostEbogui) Reveals Long-Term Detection of Ebola Viral Ribonucleic Acid in Semen and Breast Milk. *Open Forum Infect Dis* 2019 Nov 08; 6 (12): 482.

[20] Thorson A, Formenty P, Lofthouse C, Broutet N. Systematic review of the literature on viral persistence and sexual transmission from recovered Ebola survivors: evidence and recommendations. *BMJ Open* 2016 6: doi: 10.1136/bmjopen-2015-008859.

[21] https://www.ema.europa.eu/en/news/new-vaccine-prevention-ebola-virus-disease-recommended-approval-european-union

[22] Stanley DA, Honko AN, Asiedu C, et al. Chimpanzee adenovirus vaccine generates acute and durable protective immunity against ebolavirus challenge. *Nature Medicine* 2014; 20:1126-1129.

[23] Investigator's Brochure for Ad26.ZEBOV.

[24] Investigator's Brochure for MVA-BN-Filo.

[25] Milligan ID, Gibani MM, Sewell R, et al. Safety and immunogenicity of novel adenovirus type26-and modified vaccinia Ankara-vectored Ebola vaccines. A randomized clinical trial. *JAMA* 2016; 315: 1610-1623

[26] Mutua G, Anzala O, Luhn K, Robinson C, Bockstal V, Anumendem D, Douoguih M. Safety and Immunogenicity of a 2-Dose Heterologous Vaccine Regimen With Ad26.ZEBOV and MVA-BN-Filo Ebola Vaccines: 12-Month Data From a Phase 1

Randomized Clinical Trial in Nairobi, Kenya. *J Infect Dis* 2019; doi.org/10.1093/infdis/jiz071

[27] Anywaine Z, Whitworth H, Kaleebu P, et al. Safety and Immunogenicity of a 2-Dose Heterologous Vaccination Regimen With Ad26.ZEBOV and MVA-BN-Filo Ebola Vaccines: 12-Month Data From a Phase 1 Randomized Clinical Trial in Uganda and Tanzania. *J Infect Dis* 2019; doi.org/10.1093/infdis/jiz070

[28] Pollard AJ, Launay O, Lelievre JD, Lacabaratz C, Grande S, Goldstein N, Robinson C, Gaddah A, Bockstal V, Wiedemann A, Leyssen M, Luhn K, Richert L, Bétard C, Gibani MM, Clutterbuck EA, Snape MD, Levy Y, Douoguih M, Thiebaut R; EBOVAC2 EBL2001 study group. Safety and immunogenicity of a two-dose heterologous Ad26.ZEBOV and MVA-BN-Filo Ebola vaccine regimen in adults in Europe (EBOVAC2): a randomised, observer-blind, participant-blind, placebo-controlled, phase 2 trial. *Lancet Infect Dis.* 2020 Nov 17.

[29] Stittelaar KJ, Kuiken T, de Swart RL, et al. Safety of modified vaccinia virus Ankara (MVA) in immune-suppressed macaques. *Vaccine* 2001; 3700-3709.

[30] Verheust C, Goossens M, Pauwels K, Breyer D. Biosafety aspects of modified vaccinia virus Ankara (MVA)-based vectors used for gene therapy or vaccination. *Vaccine* 2012; 30:2623-2632.

[31] Investigator's Brochure for rVSVΔG-ZEBOV-GP.

[32] Jones SM, Feldmann H, Stroher U, et al. Live attenuated recombinant vaccine protects nonhuman primates against Ebola and Marburg viruses. *Nature Medicine*1:786-790.

[65] Carter RJ, Senesi RGB, Dawson P, Gassama I, Kargbo SAS, Petrie CR, Rogers MH, Samai M, Luman ET. Participant retention in a randomized clinical trial in an outbreak setting: Lessons from the Sierra Leone Trial to Introduce a Vaccine Against Ebola (STRIVE). *J Infect Dis*. 2018 May 18;217(suppl_1):S65-S74.

[66] Source: Shimabukuro T. CDC COVID-19 Vaccine Task Force. Thrombosis with thrombocytopenia syndrome (TTS) following Janssen COVID-19 vaccine. Advisory Committee on Immunization Practices (ACIP). April 23, 2021. https://www.cdc.gov/vaccines/acip/meetings/slides-2021-04-23.html.

[67] Vaccine Adverse Event Reporting System (VAERS)Standard Operating Procedures for COVID-19 (as of29 January 2021)https://www.cdc.gov/vaccinesafety/pdf/VAERS-v2-SOP.pdf
