## Supplementary Appendix for "Five-year immunogenicity and safety follow-up of the PREVAC randomized Trial of Vaccines for Zaire Ebola Virus Disease"

Figure S16. Antibody concentrations (Geometric Mean Concentrations) over time in the rVSV group (adults and children combined), according to category of antibody at baseline . 53

### Section 1. PREVAC Study Team

Jamila Aboulhab[2] M.D., Michelle Aguirre-MacKenzie[12] B.S., Pauline Akoo[3] M.B., Ch.B., Esther Akpa[2] M.S.N., M.P.H., R.N., Robert Akpata[1] M.D., Sara Albert[17] B.A., M.P.H, Boni Maxime Ale[4] M.D., M.Sc., M.P.H., Serry Alimamy-Bangura[11] M.B., Ch.B., Benetta C. Andrews[10] M.D., Stephane Anoma[6] M.D., Negin Atri[2] M.P.H., C.P.H., Augustin Augier[6] M.Com., Ken Awuondo[3] M.Sc., Ahidjo Ayouba[30] Ph.D., Moses Badio[10] M.Sc., Aminata Bagayoko[1] M.D., Abby Balde[17] M.P.H., Joséphine Balssa[1,8] Pharm.D., Lamin Molecule Bangura[11] B.Sc., Kesha Barrington[17] M.P.A., Eric Barte de Saint Fare[6] B.A., Beth Baseler[17] M.S., Ali Bauder[12] B.A., P.M.P., Claire Bauduin[4] M.Sc, Luke Bawo[10] M.Sc., Abdoul Habib Beavogui[9] M.D., Ph.D., Michael Belson[2] B.S., Safaa Ben-Farhat[4] M.Eng., Marion Bererd[6] B.S., Nicolas Bernaud[1,18] M.Sc., Teedoh Beyslow[10] Pharm.D., Neirade Biai[30] M.Sc., Jeanne Billioux[2] M.D., Shere Billouin-Frazier[17] M.Sc., Blandine Binachon[4] M.D., M.P.H., Julie Blie[10] M.Sc., Patricia Boison[17] M.S., Fatorma Bolay[10] Ph.D., Aliou Boly[6] M.M., Rachael Elizabeth Bonawitz[12] M.D., M.S., Anne-Gaëlle Borg[6] M.C.M., Samuel Bosompem[2] Pharm.D. M.Sc., Courtney Bozman[28] M.Sc., Tyler Brady[2] M.P.H., Sarah Browne[10] R.N., B.S.N., Ryan Bullis[12] Ph.D., Gabrielle Caberia[1] Pharm.D. candidate, Barbara Cagniard[1] Ph.D., Kelly Cahill[2] R.N, M.Sc., C.C.R.C., R.A.C., Yingyun Cai[28] Ph.D., Aissata Abdoulaye Camara[6] M.Sc., Aboubacar Keira Camara[1] M.D., M.Sc., Alseny Modet Camara[6] M.D., Cécilia Campion[4] M.Sc., Alexandre Cantan[4,29] B.Sc., Jennifer Cash[17] B.S., Michael Chea[10] B.Sc., Geneviève Chêne[4] M.D., Ph.D., Edward Choi[3] Ph.D., Michelle Chouinard[6] M.S.W., Florence Chung[1] Ph.D., Lucy Chung[2] Pharm.D., Papa Ndiaga Cisse[14] Ph.D., Elfrida Cline-Cole[17] M.A., Céline Colin[4] M.Sc., Beth-Ann Coller[12] Ph.D., Djélikan Siaka Conde[1] M.D., Katherine Cone[2] M.S.W., LCSW-C, C-SWHC, Laurie Connor[12] M.S., Nicholas Connor[3] M.Sc., Joseph Boye Cooper[10] M.Sc., Sandrine Couffin-Cadiergues[1] Ph.D., Mariam Coulibaly[20] Pharm.D., M.Sc., Page Crew[2] Pharm.D., M.P.H, B.C.P.S., Sandrine Dabakuyo-Yonli[4] Pharm.D., Ph.D., Djeneba Dabitao[20] Pharm.D., Ph.D., Bionca Davis[5] M.P.H., Gibrilla Fadlu Deen[11] M.D., M.Sc., Jean-François Delfraissy[1] M.D., Ph.D., Christelle Delmas[1] M.Sc., Mahamadou Diakite[20] Pharm.D., D.Phil, Alpha Diallo[1,8] M.D., M.P.H., Fatoumata Abdoulaye Diallo[6], M.D., Mamadou Saliou Diallo[6] M.D., M.P.H., Ayouba Diarra[20] M.Sc., Samba Diarra[20] M.Sc., Ph.D., Oualy Diawara[19] B.S., Ilo Dicko [20] M.D, M.P.H., Bonnie Dighero-Kemp[28] B.Sc., Samba Diop[21] M.Sc., Ph.D., Waly Diouf[14] Ph.D., Saurabh Dixit[2] Ph.D., Barry Djenabou[6] M.Sc., Laurie Doepel[2] B.A., Eric D'Ortenzio[1,7,8] M.D., M.P.H., Seydou Doumbia[20] M.D., Ph.D., Moussa Moise Doumbia[19] M.D., Nelson Dozier[28] M.Sc., Natasha Dubois Cauwelaert[1,8] Ph.D., Alain DuChêne[5] B.S., Michael Duvenhage[17] N.DIP.IT., Risa Eckes[2] R.N., Elizabeth Elliott[2] M.Sc., Luisa Enria[3] Ph.D., Hélène Espérou[1] M.D., Cécile Etienne[1] M.Sc., Allison Eyler[17] H.S.D., Lawrence Fakoli[10] M.Sc., Mosoka Fallah[10] Ph.D., Marie-Alix Fauvel[1] M.Sc., Sylvain Faye[14] Ph.D., John Fayiah[10] M.Sc., Suzanne Fleck[3] Ph.D., Vemy Fofana[6] B.Comp, Karine Fouth Tchos[2] M.D., M.P.H., Kokulo Franklin[10] M.Phil., M.Sc., Daniela Fusco[1] Ph.D., Auguste Gaddah[13] Ph.D., Marylène Gaignet[1] M.Sc., Katherine Gallagher[3] Ph.D., Julie Gardner[12], B.S., Harrison Gichini[28] M.Sc., Julia Garcia

Gozalbes[1] M.D., Greg Grandits[5] M.S, Maima Gray[10] B.Pharm., Brian Greenwood[3] M.D., Robin Gross[28] M.Sc., Louis Grue[17] R.N., B.S., B.S.N., Birgit Grund[27] Ph.D., Oumar Guindo[20] M.Sc., Pharm.D., Swati Gupta[12] Dr.P.H., M.P.H., Fadima Haidara[19] M.D., Benjamin Hamzé[1] Pharm.D., Emma Hancox[3] M.Sc., Patricia Hensley[3] M.Ph., Lisa Hensley[28] Ph.D., M.S.P.H., Betsey Herpin[2], M.S.N, Elisabeth Higgs[2] M.D., D.T.M.H., M.I.A., Trudi Hilton[3] B.Pharm., M.Sc., Mickael Hneino[1], Ph.D., Tracey-Ann Hoeltermann[28], B.Sc., M.P.H., , Horace Preston Holley[17] M.D., Marie Hoover16 Ph.D., Natasha Howard3 Ph.D., Melissa Hughes12 B.A., M.B.A., C.P.M., P.M.P., Sesay Idrissa[32] B.Sc, Skip Irvine[12] B.S., David Ishola[3] M.D., Ph.D., Yvonne Jato[2] M.P.H., Madison Joe[10] M.Sc., Melvin Johnson[10] M.Sc., Aboubacar Sidiki Kaba[6] M.D., Jonathan Kagan[2] Ph.D., Kade Kallon[17], M.Sc., Michael Kamara[3] MB.ChB, M.Sc., Myriam Kante[4] B.S., Judith Katoudi6 M.D., M.P.H., Cheick Mohamed Keita[6] M.D., Sakoba Keita[15] M.D., Seykou Keita[22] M.D., Stephen B. Kennedy[10]M.D., M.Sc., Babajide Keshinro[13] M.B.B.S., F.W.A.C.P., Hassan Kiawu[10] M.Sc., Mark Kieh[10] M.D., M.S.M.H.C., Brent Killinger[12] B.A., Moumouni Kinda[6] M.D., M.B.A., Matthew Kirchoff[2] Pharm.D., M.Sc., M.B.A., Gregory Kocher[28] M.Sc., Mamoudou Kodio[19] Pharm.D., Brian Kohn[3] B.Sc., Lamine Koivogui[23] Pharm.D., Ph.D., Richard Kojan[6] M.D., Cece Francis Kolié[6] Pharm.D., Jacques Seraphin Kolié[6] M.D., David Kollie[10] B.Sc., Stacy Kopka[17] M.S., Bockarie Koroma[11] B.Pharm., Dickens Kowuor[3] B.Sc., M.Sc., Ph.D., Catherine Kpayieli-Freeman[10] M.Sc., Ange-Marie D. Kpetigo[31] M.Sc., Christine Lacabaratz[1,18] Ph.D., Boris Lacarra[1] M.D., Laurie Lambert[17] B.S., Courtney Lambeth[12] B.S., Solange Lancrey-javal[1,8] Pharm.D., H. Clifford Lane[2] M.D., Shadrach Langba[10] B.Sc., Bolarinde Lawal[3] M.Sc., Andrew Wen-Tseng Lee[12] M.D., Shona Lee[3] Ph.D., Shelley Lees[3] Ph.D., Annabelle Lefevre[1] M.D., Bailah Leigh[11] M.D., M.Sc., Frederic Lemarcis[1] Ph.D., Yves Lévy[1,18] M.D., Ph.D., Edouard Lhomme[26] M.D., Ph.D., Janie Liang[28] M.Sc., Mameni Linga[10] M.Sc., Ken Liu[12] Ph.D., Brett Lowe[3] M.Phil., Julia Lysander[10] M.Sc., Ibrah Mahamadou[6] Pharm.D., Irina Maljkovic-Berry[28] Ph.D., Marvington Mambiah[10] A.Sc., Daniela Manno[3] M.D., Ph.D., Jonathan Marchand[2,17] M.S., Lindsay Marron[28] M.Sc., Moses B.F. Massaquoi[10] M.D., M.Sc., , Charly Matard[4] B.S., Steven Mazur[28] B.S., John McCullough[16] B.S., Chelsea McLean[13] Ph.D., Noémie Mercier[1] Pharm.D., Pauline Michavila[6] B.Bus., Tracey Miller[17] R.N., B.S.N., Niouma Pascal Millimouno [6] M.D., Alejandra Miranda[17] M.S., Soumaya Mohamed[6] B.J., Tom Mooney[3] B.A., Dally Muamba[6] M.D., James Mulbah[11] B.Pharm., Rita Lukoo Ndamenyaa[6] M.D., M.Sc., James Neaton[5] Ph.D., Désiré Neboua[1] M.D., Micki Nelson[12] B.S.N., M.S., Kevin Newell[17] M.P.H., M.Ed., Vinh-kim Nguyen[24] M.D., Yusupha Njie[3] B.Sc., Wissedi Njoh[17] M.S.N., Matthew Onorato[12] B.S., Uma Onwuchekwa[22] B.Sc., Susan Orsega[2] M.S.N., FNP-BC, Inmaculada Ortega-Perez[1,8] Ph.D., M.P.H., Cynthia Osborne[17] B.S., Tuda Otieno[3] M.SC., Davy Oulaï[4] M.D., Sushma Patel[12] M.S., P.M.P., Danielle Peart[2] B.S., Martine Peeters[30] Ph.D., James Pettitt[28] M.Sc., Nathan Peiffer-Smadja[1] M.D., Ph.D., Robert Phillips[3] M.Sc., Jerome Pierson[2] Ph.D., Peter Piot3 M.D., Ph.D., Micheal Piziali2 J.D., M.Sc., Stéphany Pong[1,8] Pharm.D., Elena Postnikova[2] Ph.D., Dudley Pratt[3] M.D., Calvin Proffitt[17] M.A., Alexandre Quach[1] M.D., Sinead Quigley[1] M.S.Sc., Nadeeka Randunu[2,17] B.Sc., M.B.A., Laura Richert[26] M.D., Ph.D., Priscille Rivière[1] M.Sc., Céline Roy[4,29], Ph.D., Amy Falk Russell[12] M.S., Philip Sahr[10] M.D., Katy Saliba[2], M.Sc., Ph.D., Mohamed Samai[11] M.B.B.S., Ph.D., Sibiry Samake[20] Pharm.D., M.Sc., Jen Sandrus[17] A.A., Ibrahim Sanogo[20] Ms.P., M.D., Yeya Sadio Sarro[20] Pharm.D., Ph.D., Serge Sawadogo[6] M.D., M.Sc., Sani Sayadi[6] M.D., M.P.H., Maxime Schvartz[1] M.D., Christine Schwimmer[4] Ph.D., Fatou Secka[3] B.Sc., M.Sc., MB.ChB, Heema

Sharma[28]M.Sc., Denise Shelley[17] M.S., Bode Shobayo[10] M.Sc., Sophia Siddiqui[2] M.D., M.P.H., Jakub Simon[12] M.D., Shelly Simpson[17] M.S., Billy Muyisa Sivahera[6] M.D., , Mary Smolskis[2] B.S.N., M.A., Elizabeth Smout[3] M.D., M.Sc., Emily Snowden[3] M.A., Anne-Aygline Soutthiphong[4,29] M.Sc., Amadou Sow[6] M.Sc., Samba O. Sow[22] M.D., M.Sc., Ydrissa Sow[2] M.D., M.P.H., Michael Stirratt[25] Ph.D., Léa Surugue[1] M.J., Sienneh Tamba[10] R.N., B.S.N., Cheick Tangara[20] M.Sc., Milagritos D. Tapia[22] M.D., Julius Teahton[10] M.Sc., Jemee Tegli[10] M.Sc., Monique Termote[4] M.Sc., Guillaume Thaurignac[30] M.Sc., Rodolphe Thiebaut[4] M.D., Ph.D., Greg Thompson[5] B.S., John Tierney[2] B.S.N., M.P.M., Daniel Tindanbil[3] M.Sc., Abdoulaye Touré[23] Pharm.D., M.P.H., Ph.D., Elvis Towalid[10] B.Pharm, Stacey Traina[12] B.S., Awa Traore[19] Pharm.D., Tijili Tyee[10] Pharm.D., David Vallée[1] Pharm.D., Renaud Vatrinet[1] Ph.D., Corine Vincent[4] M.Sc., Susan Vogel[2] R.N, B.S.N., Cedrick Wallet[4] M.Sc., Travis Warren[2] Ph.D., Deborah Watson-Jones[3] M.D., Ph.D., Wade Weaver[28] M.Sc., Deborah Wentworth[5] M.P.H., Cecelia Wesseh[10] B.Sc., Hilary Whitworth[3] Ph.D., Jimmy Whitworth[3] Ph.D., Aurelie Wiedemann[1,18] Ph.D., Wouter Willems[13] Ph.D, Barthalomew Wilson[10] M.Sc., Jayanthi Wolf[12] Ph.D., Alie Wurie[11] M.D., M.Sc., Delphine Yamadjako[17] M.S., Marcel Yaradouno[6] M.Sc., Quiawiah Yarmie[10] M.Sc., Yazdan Yazdanpanah[1,7,8] M.D., Ph.D., Shuiqing Yu[28] B.S., Zara Zeggani[6] M.Sc., Huanying Zhou[28] B.S.

**Affiliations**

[1] French Institute for Health and Medical Research (Inserm), 75013 Paris, France

[2] National Institute of Allergy and Infectious Diseases, Bethesda, MD, USA or under contract/subcontract to NIAID

[3] London School of Hygiene & Tropical Medicine, London, UK

[4] Univ. Bordeaux, INSERM, Institut Bergonié, CHU de Bordeaux, CIC-EC 1401, Euclid/F-CRIN Clinical trials platform, F-33000 Bordeaux, France

[5] School of Public Health, University of Minnesota, Minneapolis, MN, USA

[6] The Alliance for International Medical Action, Alima, B.P.15530 Dakar, Sénégal

[7] AP-HP, Hôpital Bichat-Claude Bernard, Service de Maladies Infectieuses et Tropicales, Paris F-75018, France

[8] ANRS Emerging Infectious Diseases, Paris, France

[9] Centre National de Formation et de Recherche en Santé Rurale de Maferinyah, Maferinyah, Guinea

[10] Partnership for Research on Ebola Virus in Liberia (PREVAIL), Monrovia, Liberia

[11] College of Medicine and Allied Health Sciences (COMAHS), University of Sierra Leone, Freetown, Sierra Leone

[12] Merck Sharp & Dohme Corp, Inc., Kenilworth, NJ, USA

[13] Janssen Vaccines and Prevention BV Leiden, The Netherlands

[14] Département de Sociologie, FLSH, Université Cheikh Anta DIOP, Dakar, Sénégal

[15] Agence Nationale de Sécurité Sanitaire, Conakry, Guinea
[16] Advanced BioMedical Laboratories, L.L.C., 1605 Industrial Hwy, Cinnaminson, NJ, USA
[17] Clinical Monitoring Research Program Directorate, Frederick National Laboratory for Cancer Research.
[18] Vaccine Research Institute, Univ. Paris Est Créteil, Henri Mondor Hospital, Créteil, France
[19] Centre pour le Développement des Vaccins, Ministère de la Santé, Bamako, Mali
[20] University Clinical Research Center (UCRC), University of Sciences, Techniques and Technologies of Bamako (USTTB), Bamako, Mali
[21] Liberia Institute for Biomedical Research Ethics Committee/National, Monrovia, Liberia
[22] Center for Vaccine Development and Global Health, University of Maryland School of Medicine, 685 West Baltimore Street Baltimore, MD 21201-1509, USA
[23] INSP (Institut Nationale de Santé Publique), Conakry, Guinea
[24] École de santé publique de l'Université de Montréal, Montréal, Canada
[25] National Institute of Mental Health, Bethesda, MD, USA
[26] Univ. Bordeaux, INSERM, Institut Bergonié, CHU de Bordeaux, CIC-EC 1401, Euclid/F-CRIN clinical trials platform and U1219 BPH Inria Sistm, F-33000 Bordeaux, France
[27] School of Statistics, University of Minnesota, Minneapolis, MN, USA
[28] Integrated Research Facility at Fort Detrick (IRF-Frederick), National Institute of Allergy and Infectious Diseases, National Institutes of Health (NIH), Fort Detrick, Frederick, MD, USA
[29] Univ. Bordeaux, INSERM, MART, UMS 54, F-33000 Bordeaux, France
[30] Recherche Translationnelle Appliquée au VIH et aux Maladies Infectieuses, University of Montpellier, Institut de Recherche pour le Développement, INSERM, 34090, Montpellier, France
[31] Bordeaux Population Health Research Centre, Université de Bordeaux, Inserm, and Inria, Bordeaux, France
[32] University of Management and Technology UNIMTECH, FreeTown, Sierra Leone.

### Section 2: Supplementary methods

##### S2.1. Design of the PREVAC trial

Partnership for Research on Ebola VACcination (PREVAC) comprises two randomized, double-blind, parallel-arm phase 2 trials which have evaluated the immunogenicity and safety of three Ebola vaccine strategies compared to placebo. The trial was designed to evaluate the safety and immunogenicity of the three vaccine strategies separately in adults and children. The plan from the outset, as stated in the PREVAC protocol, was to report the results for adults and children as separate trials. One strategy utilized the 2-dose Ad26.ZEBOV, MVA-BN-Filo vaccine regimen manufactured and provided by Janssen Vaccines & Prevention B.V., and the other two strategies used the rVSVΔG-ZEBOV-GP vaccine manufactured and provided by Merck & Co.

Both adults and children were enrolled at six sites in four West African countries: two sites in Guinea (Landreah, located in an urban area in Conakry, and Maferinyah, located in a rural area in the Forecariah region); one site in Liberia (Redemption Hospital in Monrovia); two sites in Bamako, Mali (the Center for Vaccine Development (CVD) and the University Clinical Research Center (UCRC) both located in an urban area in Bamako); and one site in Sierra Leone (Mambolo, a rural community in Kambia District, northern Sierra Leone). The primary results for adults and children enrolled in the latest version (4.0) of the protocol were previously published.[1]

##### S2.2. Inclusion and Exclusion Criteria and staggered enrolment

The inclusion and exclusion criteria specified in the protocol were.
Inclusion Criteria:

- Informed consent/assent
- Age ≥ 1 year
- Planned residency in the area of the study site for the next 12 months
- Willingness to comply with the protocol requirements

Exclusion Criteria:

- Fever > 38º Celsius
- History of Ebola virus disease (EVD) (self-reported)
- Pregnancy (a negative urine pregnancy test was required for females of childbearing potential, i.e., females who have experienced menarche or who are aged 14 years and older)
- Positive Human Immunodeficiency Virus (HIV) test for participants < 18 years of age
- Reported current breast-feeding
- Prior vaccination against Ebola (self-reported)
- Any vaccination in the past 28 days or planned within the 28 days after randomization (initial vaccination)
- In the judgement of the clinician, any clinically significant acute/chronic condition which would limit the ability of the participant to meet the requirements of the study protocol

All adult participants provided written informed consent. Written informed consent of the parent/guardian was obtained for children of all age groups with written assent if the child was aged 7 to 17 years old.

Enrollment was staggered according to age group, starting with adults and with adolescents 12 to 17 years of age, followed by children 5 to 11 years of age, and finally children 1 to 4 years of age.

**S2.3. Randomisation and masking**

Participants were randomly assigned to one of the following five groups in a 2:1:2:1:1 allocation. Participants in group 1 received Ad26.ZEBOV (0.5 mL; $5\times10^{10}$ viral particles) at day 0, followed by MVA-BN-Filo (0.5 mL; $1\times10^{8}$ infectious units) at day 56; group 2 received matched placebo (0.5 mL) at days 0 and 56; group 3 received rVSVΔG-ZEBOVGP (two rVSVΔG-ZEBOV-GP lots were used in the trial, both with titers $\geq 7.2 \times 10^{7}$ pfu/mL) (1 mL) at day 0, followed by placebo (1 mL) at day 56; group 4 received rVSVΔG-ZEBOVGP (1 mL) at day 0, followed by rVSVΔG-ZEBOV-GP (1 mL) at day 56; and group 5 received matched placebo (1 mL) at days 0 and 56 (Figure SM1). The placebo used in the trial was normal saline;

two placebo groups were used due to the different fill volumes of the two vaccines. Data from placebo groups 2 and 5 were pooled for analyses. The four resulting randomized groups are referred to as Ad26, MV; rVSV; rVSV–booster; and placebo.

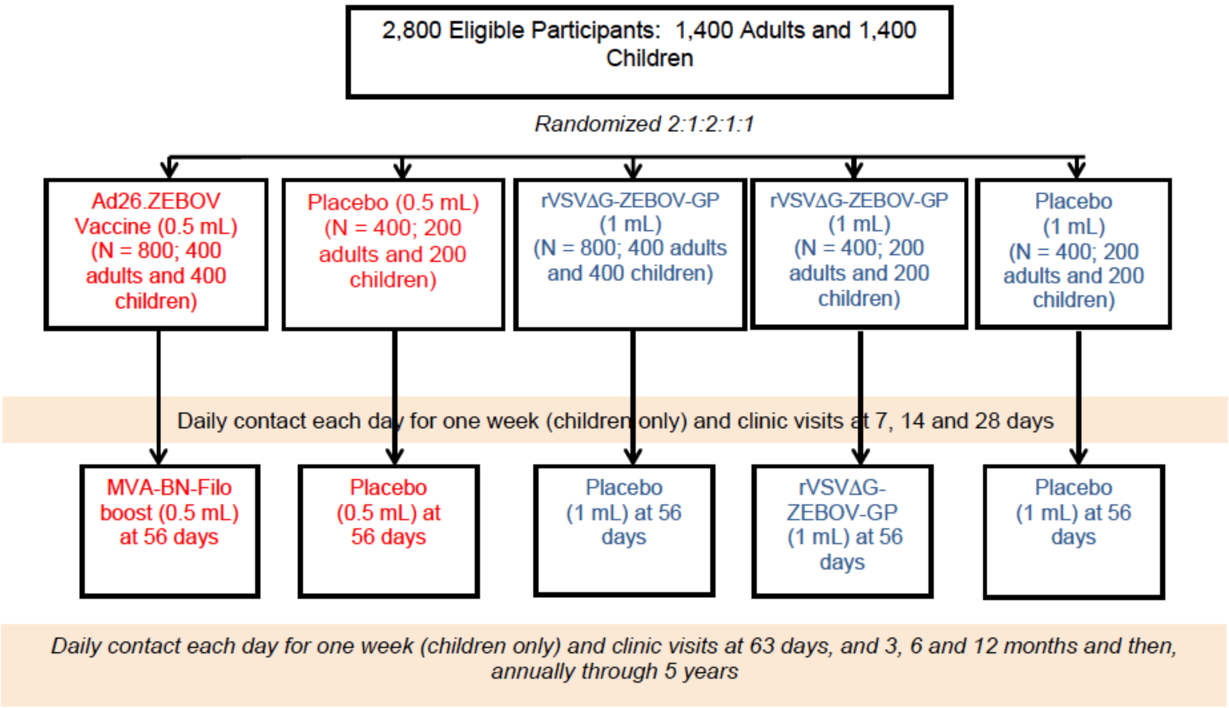

**Figure SM1**: PREVAC Study Design (Version 4.0)

Study participants and clinical and laboratory staff assessing the study participants for safety and laboratories carrying out safety and immunogenicity analyses were blinded to whether vaccine or matched placebo was given. Trial investigators were also blinded to interim data reviewed by the independent data safety monitoring board (DSMB). After completion of the first-year follow-up and the lock of interim Month-12 (M12) database the study was unblinded. The long-term annual visit and long-term safety assessment were carried out unblinded.

##### S2.4. Data circuit and dissemination

The trial was designed by the University of Minnesota in collaboration with Inserm-Euclid (Inserm/Univ Bordeaux); LSHTM; Partnership for Research on Ebola Virus in Liberia (PREVAIL), Monrovia, Liberia; Centre National de Formation et de Recherche en Santé Rurale de Maferinyah, Maferinyah, Guinea; University Clinical Research Center (UCRC), University of Sciences, Techniques and Technologies of Bamako (USTTB), Bamako, Mali; College of Medicine and Allied Health Sciences (COMAHS), University of Sierra Leone, Freetown, Sierra Leone.

Data were collected by the three sponsors of the trial, the NIH, Inserm and LSHTM and by the principal investigators of the four participating countries: Liberia, Guinea, Sierra Leone and Mali.

The manuscript was written by the writing group (the full list is detailed in the manuscript). The Trial Steering Committee (TSC) of the trial authorized the publication of the results.

All authors had full access to the data.

**S2.5. Measurement of antibodies to Ebola Glycoprotein**

For the M12 results of the PREVAC trial (PREVAC study team, NEJM 2022), the Filovirus Animal Nonclinical Group (FANG) enzyme-linked immunosorbent assay (ELISA) assay was used to measure the Ebola-specific antibody response to the glycoprotein (GP) Kikwit antigen for the primary day 0 (D0) to Month 12 (M12) analysis. Antibody concentrations were determined in two laboratories: The Liberian Institute for Biomedical Research (LIBR) in Charlesville, Liberia (analyzes of samples from Guinea and Sierra Leone participants), and the National Institute of Allergy and Infectious Diseases (NIAID) Integrated Research Facility (IRF) in Frederick, Maryland (analyzed samples from Liberia and Mali participants). More information on the FANG ELISA assay and its inter- and intra-lab variability is detailed in supplementary appendix of the PREVAC M12 primary analysis published in the New England Journal of Medicine in December 2022. All results of the antibody responses were expressed in ELISA unit (EU)/mL.

For the PREVAC trial long-term analysis (Month 24 to Month 60), the consortium opted to transition to the Luminex assay to determine the Ebola-specific antibody response to the same antigen (GP Kikwit). All analyses for the long-term follow-up were conducted in a centralized laboratory at TransVIHMI, Institut de Recherche pour le Développement (IRD) in Montpellier, France. Intrinsic parameters of the Luminex test have been assessed prior to this decision by the PREVAC TSC and are documented in the minutes of a PREVAC TSC Meeting held on 06/04/2023 and in the published manuscript "Comparison of Four Assays That Measure Antibodies to Ebola Virus Glycoprotein".[2]".

S2.5.1. Luminex assay laboratory protocol

All instructions for carrying out the multiplex serological test using xMAP technology (Luminex®) are described in the Luminex Assay Standard Operating Procedure.

Luminex® technology works according to the combined principles of ELISA and flow cytometry using fluorescent beads. The beads have a different color to identify the antigen to which they have been coupled. A biotinylated secondary antibody (directed against human IgG for example), associated with streptavidin coupled with phycoerythrin (SAPE), makes it possible to quantify the presence or absence of interaction and is subsequently expresses as an MFI (Median Fluorescence Intensity) value.

A batch corresponds to recombinant proteins to be coupled to several magnetic COOH beads. Every batch of coupled beads allows the analysis of 2700 samples tested in duplicate on 60 different plates. Overall, 8 batches of beads were used to test the M24 to M60 samples from 14/09/2023 to 24/10/2024. Samples were diluted 1/200 and tested in duplicate on the same plate.

The dilution was done by depositing 2 µL of the sample in 398 µL dilution buffer, then 200 µL of this dilution were taken to prepare the duplicate.

%CV and ULOQ management for Luminex assay

If the results (Net MFI) between duplicates presented an intra-assay coefficient of variation (CV) <20%, results were validated. Otherwise, if a CV>20% was observed and a mean result above a threshold (100 MFI for GPKik and 300 MFI for GPmay), results were considered invalid and retested in a second-round test in duplicate. An inter-assay CV between the four MFI values was calculated and pairs of duplicates with inter-assay CV<30% were considered for downstream analysis.

The Luminex Lower Limit of Quantification (LLOQ) and Upper Limit of Quantification (ULOQ) were estimated by the IRD laboratory at 10.47 and 4,154 MFI, respectively.

Antibody responses above the ULOQ were retested in duplicates with higher dilution factors (1,000 or 2,000) until the level obtained was below the ULOQ.

Determination of valid and invalid results is summarized in Figure SM2.

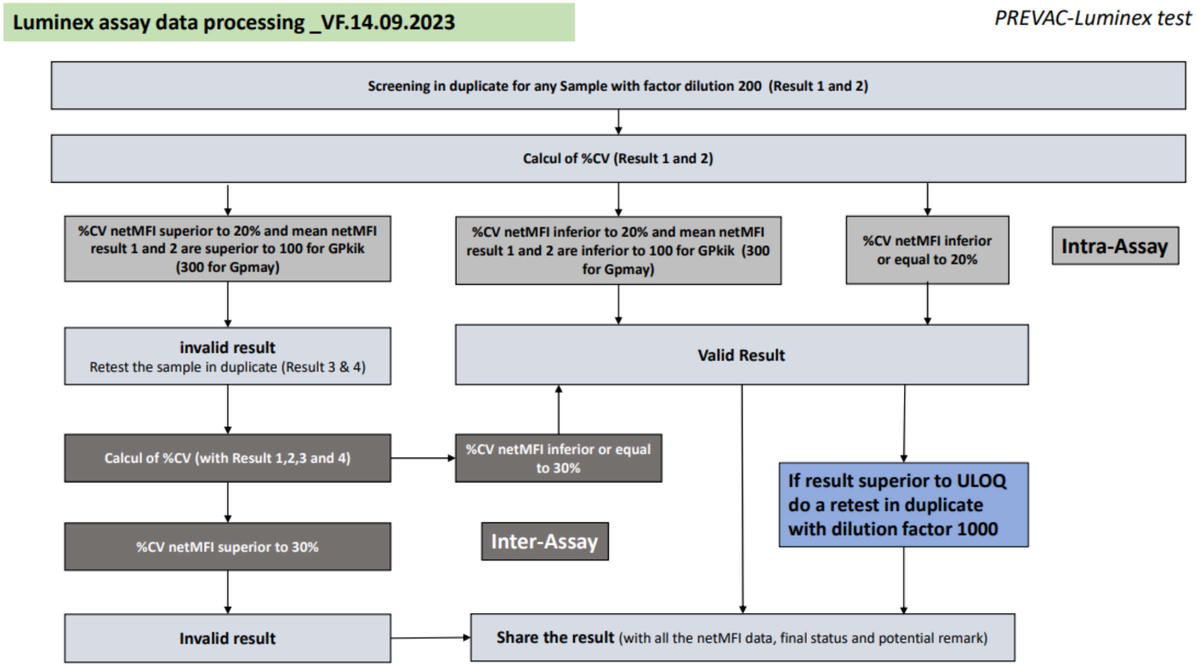

**Figure SM2**: Data processing algorithm

<u>Conversion of Luminex antibody responses from net MFI to International Unit (IU)/mL</u>

To ensure comparability between D0-M12 data and the long-term follow-up immunological results, the Luminex assay results for M24-M60 samples, initially expressed in net MFI units, were converted to IU/mL by using a conversion formula.

The conversion formula was obtained using Partnership for Research on Ebola VACcination-extended follow-up (PREVAC-UP) antibody samples calibrated with the World Health Organization (WHO) reference standard (WHO standards sera [15/220 and 15/262], which have known concentrations in IU/mL. The calibration was performed on batch 2 of the Luminex assay and controlled on batches 7 and 8, to determine the concentrations in IU/mL. A standard curve was derived for conversion from net MFI to IU/mL on log10 scale based exclusively on batch 2 data (Figure SM3). The calibration obtained in batches 7 and 8 gave similar conversion formulas.

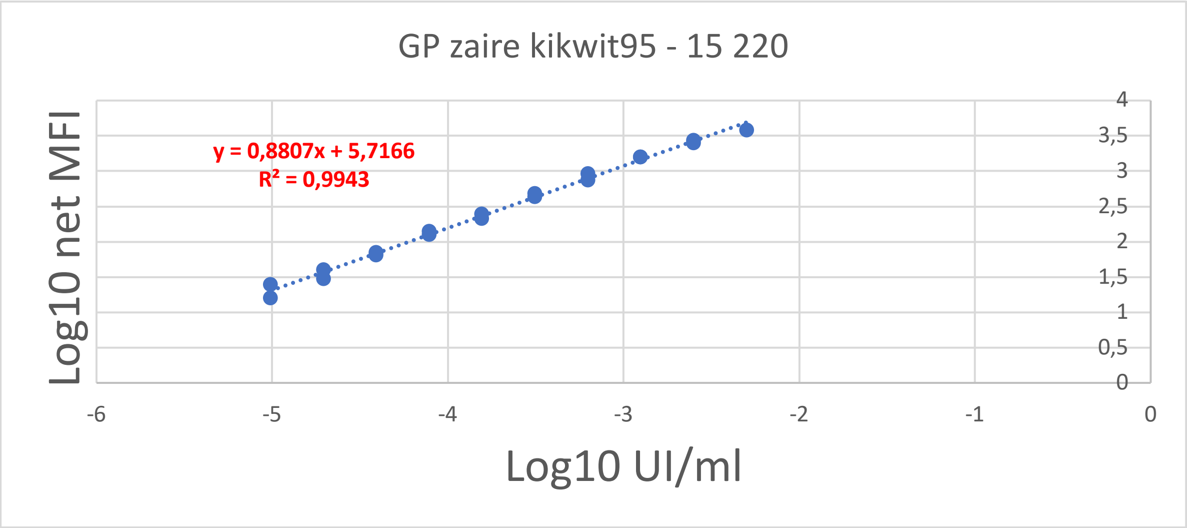

**Figure SM3**: Batch 2 WHO standards sera (15/220 and 15/262) standard curve

<u>Conversion of Luminex antibody responses from IU/mL to EU/mL</u>

The correspondence between IU/mL and EU/mL had been previously established at 1 IU/mL=27,135.90 EU/mL (Rudge TL Jr, Sankovich KA, Niemuth NA, Anderson MS, Badorrek CS, Skomrock ND, Cirimotich CM, Sabourin CL. Development, qualification, and validation of the Filovirus Animal Nonclinical Group anti-Ebola virus glycoprotein immunoglobulin G enzyme-linked immunosorbent assay for human serum samples. PLoS One. 2019;14(4):e0215457. doi: 10.1371/journal.pone.0215457). Following this conversion, all Luminex data were multiplied by the appropriate dilution factor and were expressed in EU/mL.

<u>S2.5.2. Bridging between FANG and Luminex assays</u>

To adjust for the impact of the change in technique on the measured antibody concentrations in PREVAC/PREVAC-UP, a subset of 2,467 D0-M12 samples was selected to be retested with the Luminex assay. These samples were selected to represent the full range of the FANG antibody concentrations obtained at each FANG ELISA laboratory (LIBR: Guinea and Sierra Leone samples and IRF: Liberia and Mali samples), by selecting randomly +/- 10% of the samples in each decile and adding the minimum and the maximum concentrations. In addition, all samples of immunological (Guinea samples) and Malaria sub-studies (Sierra Leone samples) (at D0, D63, M3, M6, M12) were selected to be retested with the Luminex assay.

The analysis of samples by both the Luminex and the FANG ELISA showed that the Luminex assay gives higher antibody concentrations in EU/mL than those obtained with the FANG ELISAparticularly for higher values (Figure SM4).

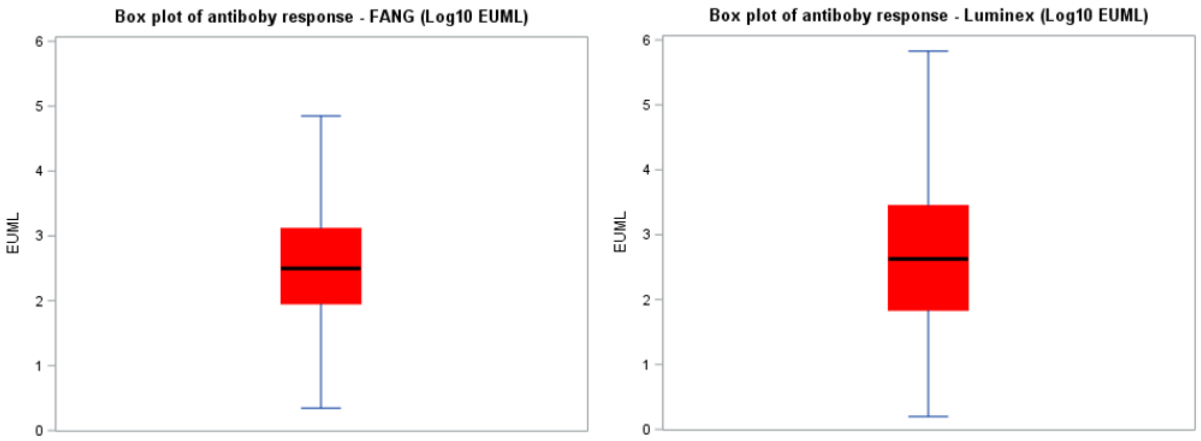

**Figure SM4**: Box plots of log10 antibody responses measured with the FANG (left) and the Luminex (right - EU/mL converted)

This may be explained in part by the Hook effect, which has been observed in the FANG ELISA assay readouts in samples with high antibody concentrations, resulting in a decrease of positive signal. The Hook effect is usually observed in undiluted samples due to too many antibodies and too few targets and complexes being built, with the overflow blocking the signal. Here, the FANG ELISA samples were diluted. However, a Hook effect was still suspected, indicating interference with the signal at higher dilutions in the FANG ELISA. For the longitudinal analyses from Month 0 to Month 60 in PREVAC, conversion formulas were thus established to bring the initial FANG ELISA data (M0 to M12) to the Luminex scale (M24-M60).

Due to the inter-laboratory variability of the FANG ELISA data that had been previously detected (PREVAC Study group, NEJM 2022), all steps described below are stratified on the FANG ELISA laboratory: LIBR laboratory, and IRF laboratory:

Data from samples measured with both assays were used to derive conversion formulas for all Day 0–Month 12 FANG ELISA values by FANG ELISA laboratory (IRF vs. LIBR), to bring them to the Luminex assay scale through Deming regression. This regression is used, particularly for method comparison studies to look for systematic differences between two measurement methods as it accounts for errors in both the dependent and independent variables.

It assumes that the available data (yi, xi) are measured observations of the "true" values (yi*, xi*), which lie on the regression line:

$$y_i = y_i^* + \varepsilon_i$$

$x_i = x_i^* + \sigma_i$

where errors ε and σ are independent and the ratio of their variances is assumed to be known.

The Deming Regression seeks to find the line of "best fit" $y^* = \beta_0 + \beta_1 x^*$ , such that the weighted sum of squared residuals of the model is minimized.

Because of a non-linear association between FANG ELISA and Luminex (EU/mL) antibody responses at low concentration levels that could not be fitted using the Deming regression, the decision was made to exclude all Luminex/FANG ELISA paired samples with either an antibody response below 100 EU/mL for the calibration of the Deming regression (Figures SM5 and SM6). Figures SM7 and SM8 show responses measured with the FANG ELISA and the Luminex (EU/mL converted) techniques and the better fit of the Deming regression line after censoring.

Note that the SAS macro wrapper for an efficient Deming regression algorithm via PROC IML (The Wicklin Method[1] by Jesse A. Canchola and Ben Wang) was used for estimates and graphs.

[1] https://pharmasug.org/proceedings/2022/SA/PharmaSUG-2022-SA-004.pdf

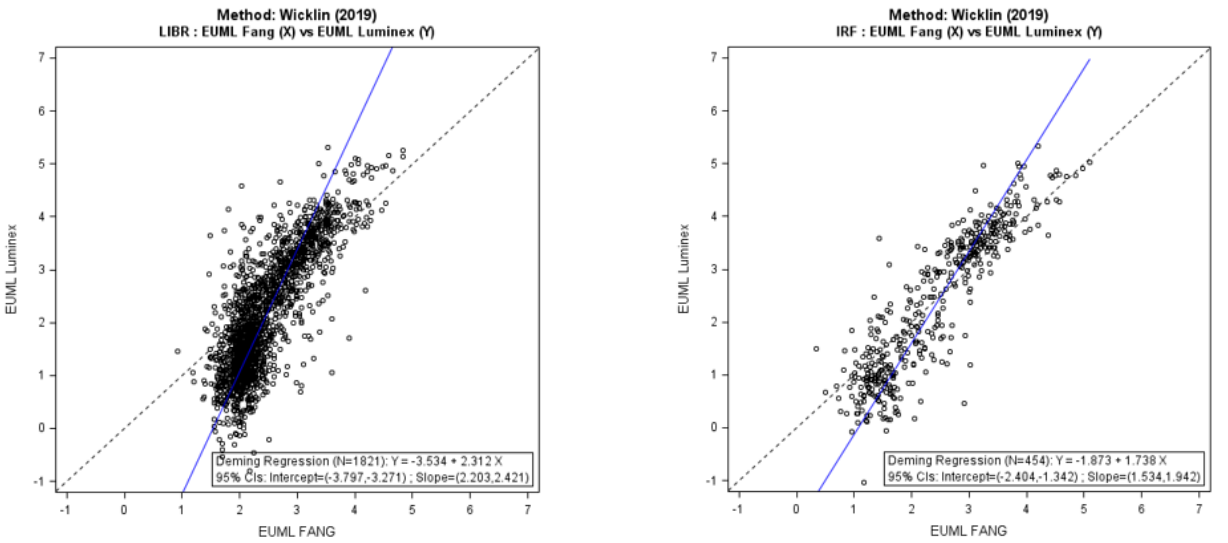

**Figure SM5**: Scatter Plot of log10 antibody responses measured with the FANG ELISA and the Luminex (EU/mL converted) assay and Deming regression line, for LIBR (left) and IRF (right) FANG ELISA laboratory, before censoring of values < 100 EU/mL

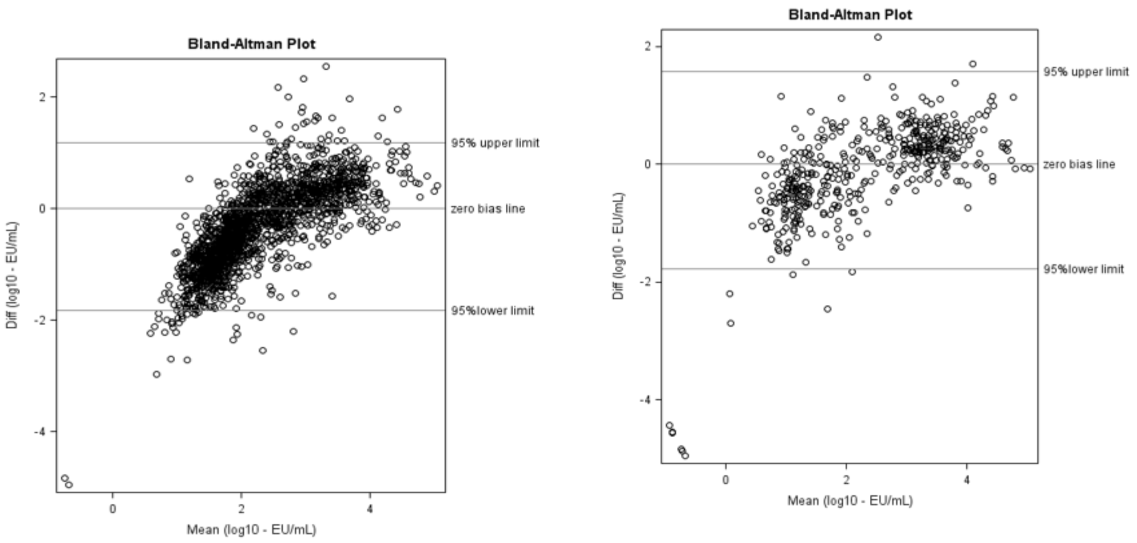

**Figure SM6**: Bland Altman of log10 antibody responses measured with the FANG ELISA and the Luminex (EU/mL converted) assay, for LIBR (left) and IRF (right) FANG ELISA laboratory, before censoring of values < 100 EU/mL

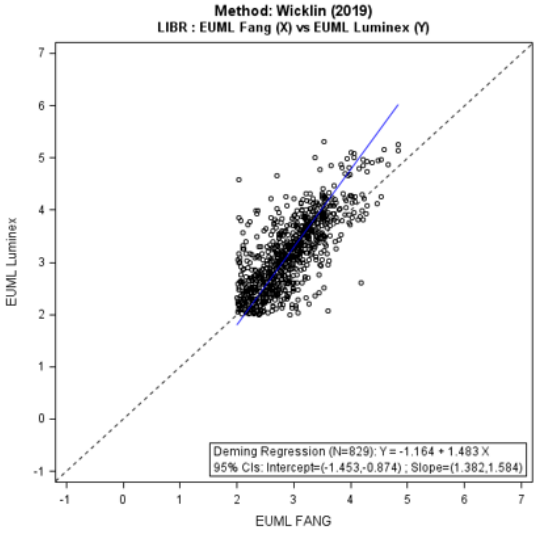

**Figure SM7**: Scatter Plot of log10 antibody responses measured with the FANG ELISA and the Luminex (EU/mL converted) assay and Deming regression line, for LIBR (left) and IRF (right) FANG ELISA laboratory, after censoring of values < 100 EU/mL

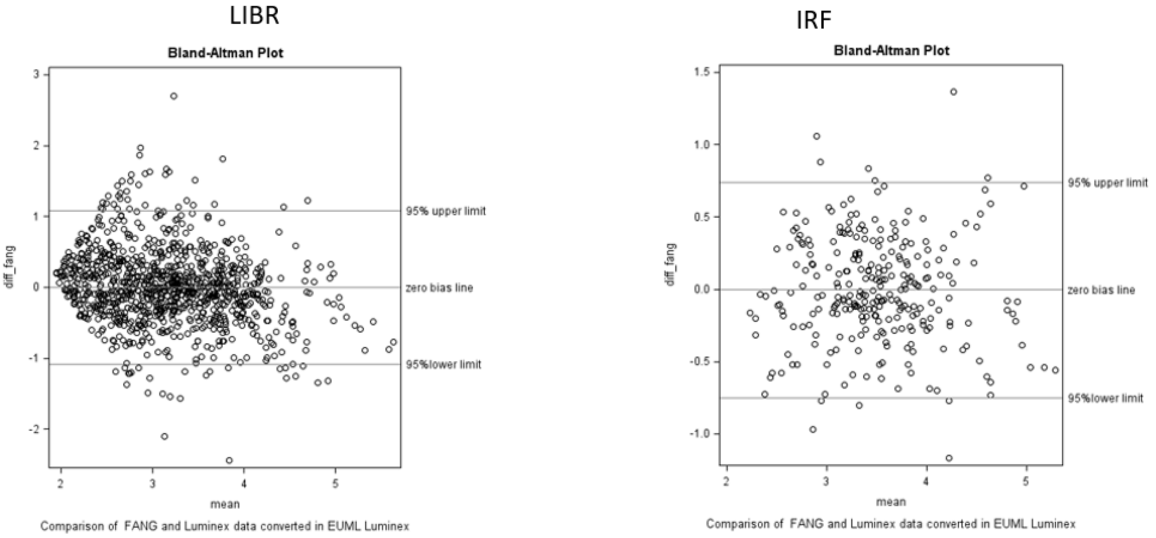

**Figure SM8**: Bland-Altman Plot of log10 antibody responses measured with the FANG ELISA and the Luminex (EU/mL converted) assay, for LIBR (left) and IRF (right) FANG ELISA laboratory, after censoring of values < 100 EU/mL

The resulting Deming regression equations used for conversion are shown in the box below.

LIBR Laboratory

$Log10(FANG_{LuminexScale}) = -1.163747 + 1.4830533 * Log10(FANG_{FangScale})$

IRF Laboratory

$Log10(FANG_{LuminexScale}) = 0.0157716 + 1.1004654 *Log10(FANG_{FangScale})$

Each formula was applied to the corresponding D0-M12 FANG ELISA results that had not been analyzed with the Luminex technique (n=21466 samples).
After this step, all PREVAC data were expressed in EU/mL on the Luminex scale.

##### S2.6. Full statistical analysis plan (in a separate document)

##### S2.7. Summary of statistical analyses

The analysis of immunogenicity endpoints was conducted as treated: this means that the participants who had received at least one injection of a given vaccine strategy were included in the as-treated analyses according to the true vaccine strategy received. Participants initially randomized to the rVSV–Booster group but who received Placebo as booster or no injection were analyzed in the rVSV group (n=10). Other participants who had received at least one injection of a wrong vaccine strategy (i.e. different "mix-and match" of vaccines than those planned in the protocol) were excluded from the analysis.

Following the new Ebola outbreak declared in Guinea in February 2021 and the vaccine licensures, most participants who received the placebo were vaccinated during the long-term follow-up. Hence, for comparisons at Months 36, 48, and 60, antibody values for vaccinated placebo participants were replaced with their last available measurements prior to vaccination.

###### S2.7.1. Antibody concentrations: descriptive analyses

The descriptive summaries of antibody levels were carried out separately for children and adults. The following were described:

- the Geometric Mean Concentration (GMC) and its 95% confidence interval (CI) at each time point, per vaccine group, from D0 to M60 and the fold increase from D0 to 24, 36, 48 and 60 Months, respectively, per vaccine group (< 4, 4 to < 8, 8 to <16, ≥16).
- box plots of antibody levels at each time point, per vaccine group, from D0 to M60.
- the antibody responses rates, per vaccine group, with different fold change categories from baseline at each long-term follow-up time point. With this definition, participants with a missing baseline or follow-up result were excluded from the response rate analyses.

*Linear mixed-effects regression model*

To describe the evolution of the $\log_{10}$ anti-Ebola virus (anti-EBOV) $GP_{1,2}$ immunoglobulin G (IgG) antibody concentrations between month 24 and month 60 in the Ad26, MVA; rVSV; and rVSV-booster groups, a linear mixed effects regression model was used, in which the fixed intercept was adjusted for the following: vaccine strategy (with Ad26.ZEBOV, MVA-BN-Filo as reference), sex (with male as reference), country (Guinea, Liberia, and Sierra Leone (SL), with Mali as reference), age category (children 1-4, 5–11, and 12-17 years, with adults as reference), and pre-vaccination EBOV $GP_{1,2}$ IgG antibody concentration (M0), as single effects; and sex, country, age category, and pre-vaccination EBOV $GP_{1,2}$ IgG antibody concentrations in interaction with vaccine strategy.

The fixed slope was adjusted for country, vaccine strategy and an interaction term between these two co-variates. In addition to covariate effects, random effects on both intercept and slope were considered. The resulting model is defined by the following equation:

$$Ab_{ij}$$
$$= \beta 0 + \gamma 0_i + \beta 1 t_{ij} + \gamma 1 t_{ij} + \beta 2 Arm1_i$$
$$+ \beta 3 Arm2_i + \beta 4 Arm1_i \times t_{ij} + \beta 5 Arm2_i$$
$$\times t_{ij} + \beta 6 Age_{(1-4)i} \ + \beta 7 Age_{(5-11)i} + \beta 8 Age_{(12-17)i}$$
$$+ Arm1_i(\beta 9 Age_{(1-4)i} + \beta 10 Age_{(5-11)i} + \beta 11 Age_{(12-17)i}) + Arm2_i(\beta 12 Age_{(1-4)i} + \beta 13 Age_{(5-11)i} + \beta 14 A$$ [illegible]
$$+ \beta 15 Sex_{Femalei} + \beta 16 Sex_{Femalei} \times Arm1_i$$
$$+ \beta 17 Sex_{Femalei}$$
$$\times Arm2_i \ + \beta 18 Country_{Guineai} \ + \beta 19 Country_{Liberiai} + \beta 20 Country_{SLi}$$
$$+ Arm1_i(\beta 21 Country_{Guineai} + \beta 22 Country_{Liberiai} + \beta 23 Country_{SLi})$$
$$+ Arm2_i(\beta 24 Country_{Guineai} + \beta 25 Country_{Liberiai} + \beta 26 Country_{SLi})$$
$$+ t_{ij}(\beta 27 Country_{Guineai} + \beta 28 Country_{Liberiai} + \beta 29 Country_{SLi}) + \beta 30 Country_{Guineai}$$
$$\times Arm1_i \times t_{ij} + \beta 31 Country_{Guineai} \times Arm2_i \times t_{ij} + \beta 32 Country_{Libériai}$$
$$\times Arm1_i \times t_{ij} + \beta 33 Country_{Libériai} \times Arm2_i \times t_{ij} \ + \beta 34 Country_{SLi}$$
$$\times Arm1_i \times t_{ij} + \beta 35 Country_{SLi} \times Arm2_i \times t_{ij} + \beta 36 Abb_i$$
$$+ \ \beta 37 Abb_i \times Arm1_i + \ \beta 38 Abb_i \times Arm2_i + \ \varepsilon_i$$

This model was used to fit data from the Ad26, MVA; rVSV; and rVSV-booster groups from the month 24 ($t$=0) to month 60 ($t$=3).

$Ab_{ij}$ represents the $\log_{10}$ IgG antibody concentration of the participant $i$ at timepoint $j$. Thus, $t_{ij}$ characterizes the variable of time rescaled to consider 24-month post-vaccination as the baseline ($t$=0). The variable $Abb_i$ is the continuous covariate pre-vaccine IgG antibody concentration and describes the anti-EBOV $GP_{1,2}$ IgG antibody concentration of the participant $i$ measured at inclusion, before vaccine injection. The vector of parameters $\beta = (\beta_0, \ldots, \beta_{38})$ represents the fixed effects related to the different covariates, and the parameters $\gamma_{0i}$ and $\gamma_{1i}$ are the random effects on the intercept and the slope, respectively. Random effects were assumed as normally distributed with mean 0, and covariance matrix $\Omega$ was structured. Finally, the variable $\varepsilon_{ij}$ represents the residual error assumed to be normally distributed with mean 0 and variance $\sigma^2$.

Model parameter estimates and the q-q plots of random effects and residuals are provided below (Figure SM8), indicating a satisfactory model fit and no major deviation from normality assumptions.

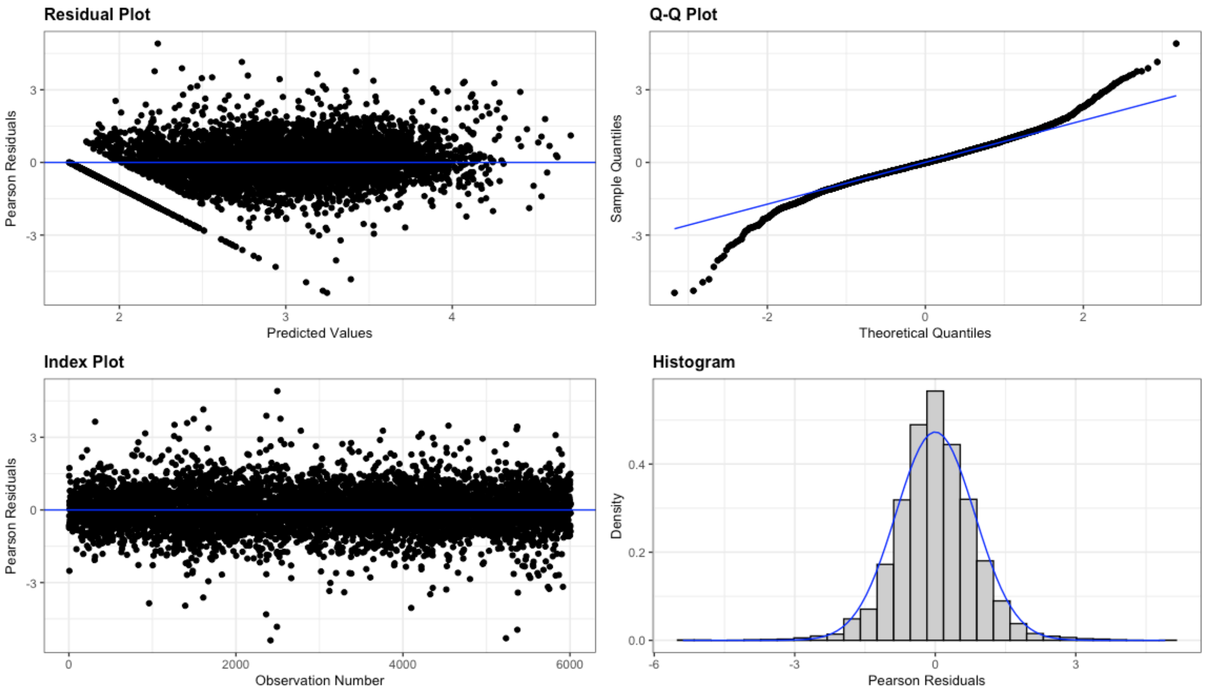

**Figure SM8:** Linear Model with mixed effect Residual Diagnostics: Graphical Analysis Including Residuals vs Fitted Values, Q-Q Plot, Residuals Index Plot, and Histogram of Residuals with Density Curve.

*Internal predictive quality of the model*

To evaluate the performance of the linear mixed effect model to predict antibody concentration, we performed a leave-one-out cross-validation (LOOCV).

The mixed model developed on the complete dataset was then estimated on the training dataset and used to predict the observation of the testing dataset (i.e., the observation at month 24-60 of the kth participant). This procedure was repeated as many times as the number of participants in the complete dataset and two criteria were then calculated on predictions to evaluate how accurate they are: (1) the percent coverage (the higher the better) and (2) the root mean squared error (RMSE, the lower the better). Results are displayed in the following figure representing the corresponding LOOCV predictions (Figure SM9). The model showed good LOOCV prediction quality with a percent of coverage of 97.3% and an RMSE of 0.243 EU/mL.

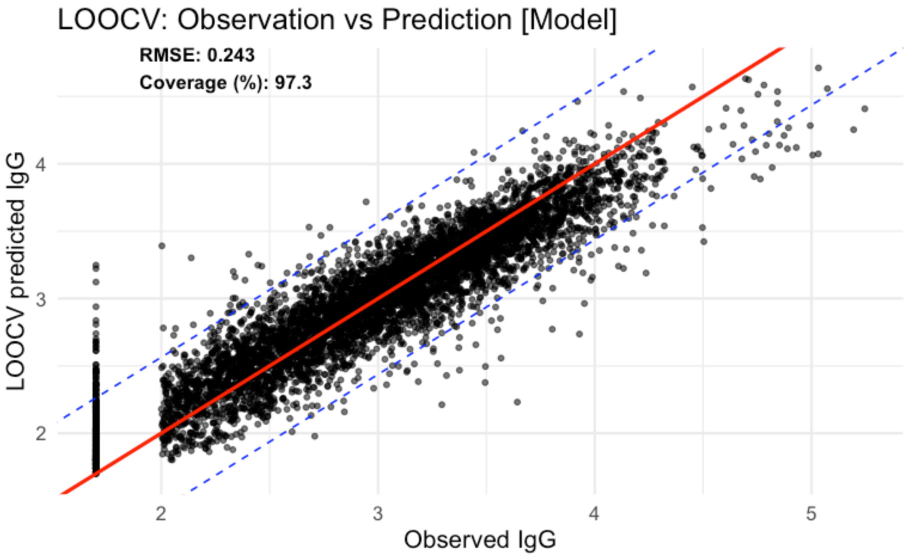

**Figure SM9: Results of LOOCV: observations vs. predictions plot [Model].**

The x axis represents individual observations at month 24-60 (successively removed from the training dataset), and the y axis represents predictions of IgG at month 24-60 made by models at each iteration. Each black dot represents the result of one iteration of the LOOCV (prediction made for the observation removed from the dataset).

The red solid line represents the relationship Observation = Prediction. Dashed blue lines represent the 95% confidence interval built on the distribution of the residuals of the model estimated on complete data (i.e., observation ± 1.96*SD, with SD being the standard error of the error model of the linear mixed-effect model estimated on complete data). LOOCV = leave-one-out cross-validation. IgG = immunoglobulin G. SD = standard deviation.

*Estimation of geometric mean concentration (GMC) ratios*

For the estimated linear mixed-effect model, the effect of each categorical covariate (age, sex, country, and pre-vaccination IgG antibody concentration) was evaluated as the ratio of the GMCs of anti-EBOV $GP_{1,2}$ IgG antibody between the category of interest and its reference. Ratios of GMCs and their 95% CI at the end of the study (5 years post-vaccination) were estimated with a classic bootstrap procedure:

1) we sampled with replacement 1,837 individuals;

2) the linear mixed effects model was estimated, and computed prediction individual trajectories;

3) we calculated the predicted trajectories of ratios of GMCs, on the original scale of antibody concentration, for each category of each covariate at the end of the study.

We repeated these steps 2,000 times, which enabled us to derive the mean and the 95% CI of each variable of interest depending on vaccination arm.

S2.7.2. Antibody concentrations: comparative analyses

For each long-term timepoint (months 24-60), each active vaccine strategy was compared with the pooled placebo group, at the 0.0167 (2-sided) level of significance.

After $\log_{10}$-transformation, mean antibody concentrations were compared at each long-term M24 and M60 follow-up timepoint, respectively, for each vaccine group versus placebo, for adults and children separately, using analysis of covariance models with the baseline $\log_{10}$ concentration and site as covariates. GMCs after back-transformation are reported. In addition, geometric mean ratios and 95% CIs are reported for each vaccine versus placebo comparison for each long-term follow-up timepoint.

S2.7.3. Safety analyses

The safety analysis was descriptive and conducted as-treated. The safety endpoint was the occurrence of a serious adverse event (SAEs) at any time during the 12 and 60 months follow-up period. Count of participants with events, overall and by system organ class according to Medical Dictionary for Regulatory Activities (MedDRA), and percents (with 95% CIs) are provided by vaccine group.

### Section 3: Supplementary results

#### S3.1. Population

##### Table S1. Participants characteristics at baseline (PREVAC trials population included at baseline, intent-to-treat population)

| | Ad26, MVA Group | | rVSV Group | | rVSV–Booster Group | | Placebo Group | |
|---|---|---|---|---|---|---|---|---|
| | N | % | N | % | N | % | N | % |
| **Adults** | | | | | | | | |
| **No. of participants** | 397* | 100,0 | 395 | 100,0 | 197 | 100,0 | 412 | 100,0 |
| **Age** | | | | | | | | |
| 18-29 yr | 228 | 57,0 | 213 | 54,0 | 119 | 60,0 | 238 | 58,0 |
| 30-39 yr | 67 | 16,88 | 89 | 22,5 | 41 | 20,8 | 84 | 20,4 |
| >= 40 yr | 102 | 26,0 | 93 | 24,0 | 37 | 19,0 | 90 | 22,0 |
| **Sex** | | | | | | | | |
| Male | 225 | 57,0 | 213 | 54,0 | 110 | 56,0 | 225 | 55,0 |
| Female | 172 | 43,3 | 182 | 46,1 | 87 | 44,2 | 187 | 45,4 |
| **Site** | | | | | | | | |
| Landreah | 82 | 20,7 | 81 | 20,5 | 38 | 19,3 | 80 | 19,4 |
| Maferinyah | 40 | 10,0 | 49 | 12,0 | 28 | 14,0 | 55 | 13,0 |
| Redemption | 73 | 18,4 | 70 | 17,7 | 35 | 17,8 | 81 | 19,7 |
| Mambolo | 121 | 30,0 | 111 | 28,0 | 55 | 28,0 | 110 | 27,0 |
| UCRC | 43 | 10,8 | 45 | 11,4 | 19 | 9,6 | 45 | 10,9 |
| CVD | 38 | 10,0 | 39 | 10,0 | 22 | 11,0 | 41 | 10,0 |
| **HIV Status** | | | | | | | | |
| Negative | 391 | 98,0 | 382 | 97,0 | 195 | 99,0 | 408 | 99,0 |
| Positive | 6 | 1,5 | 13 | 3,3 | 2 | 1,0 | 4 | 1,0 |
| **Children** | | | | | | | | |
| **No. of participants** | 403 | 100,0 | 407 | 100,0 | 202 | 100,0 | 389 | 100,0 |
| **Age** | | | | | | | | |
| 1-4 yr | 137 | 34,0 | 123 | 30,0 | 71 | 35,0 | 136 | 35,0 |
| 5-11 yr | 127 | 31,5 | 146 | 35,9 | 65 | 32,2 | 129 | 33,2 |
| 12-17 yr | 139 | 34,0 | 138 | 34,0 | 66 | 33,0 | 124 | 32,0 |
| **Sex** | | | | | | | | |
| Male | 217 | 54,0 | 222 | 55,0 | 117 | 58,0 | 207 | 53,0 |
| Female | 186 | 46,2 | 185 | 45,5 | 85 | 42,1 | 182 | 46,8 |
| **Site** | | | | | | | | |
| Landreah | 80 | 19,9 | 81 | 19,9 | 43 | 21,3 | 81 | 20,8 |
| Maferinyah | 83 | 21,0 | 75 | 18,0 | 34 | 17,0 | 69 | 18,0 |
| Redemption | 64 | 15,9 | 66 | 16,2 | 33 | 16,3 | 55 | 14,1 |
| Mambolo | 81 | 20,0 | 91 | 22,0 | 46 | 23,0 | 93 | 24,0 |
| UCRC | 43 | 10,7 | 41 | 10,1 | 23 | 11,4 | 41 | 10,5 |

| | | | | | | | | |
|---|---|---|---|---|---|---|---|---|
| CVD | 52 | 13,0 | 53 | 13,0 | 23 | 11,0 | 50 | 13,0 |
| **HIV Status** | | | | | | | | |
| Negative | 403 | 100,0 | 407 | 100,0 | 202 | 100,0 | 389 | 100,0 |
| Positive | - | - | - | - | - | - | - | - |

* One participant of the Ad26, MVA group, which was not analyzed in D0-M12 main analysis, has been analyzed in the long term analysis, after resolution of informed consent issues.
**CVD: Centre pour le Développement des Vaccins
***UCRC: University Clinical Research Center

#### Figure S1. CONSORT intention-to-treat Diagram: Adults

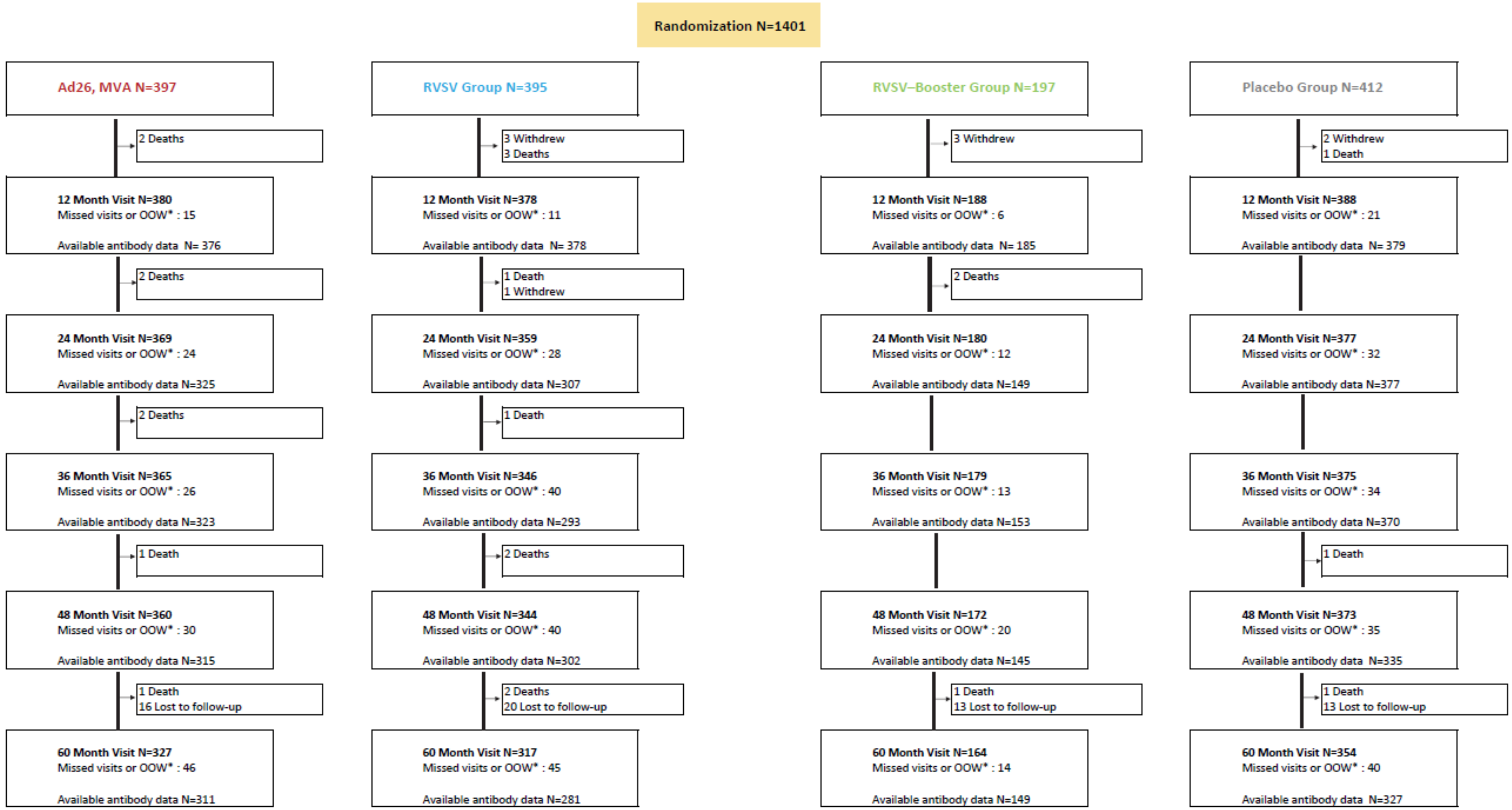

*OOW: out of window

**Figure S2. CONSORT intention-to-treat Diagram: Children**

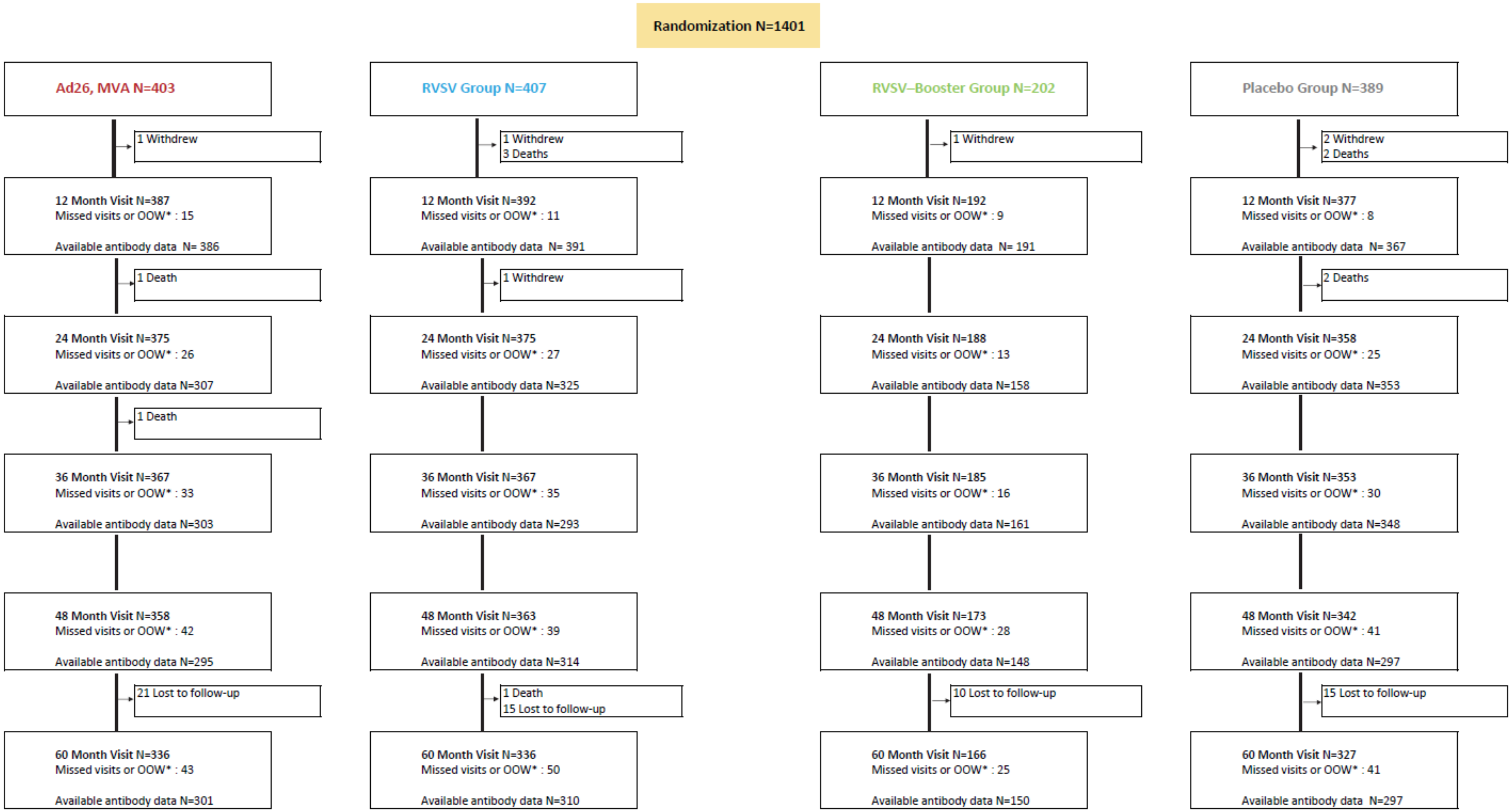

*OOW: out of window

**Figure S3. CONSORT as-treated Diagram: Adults**

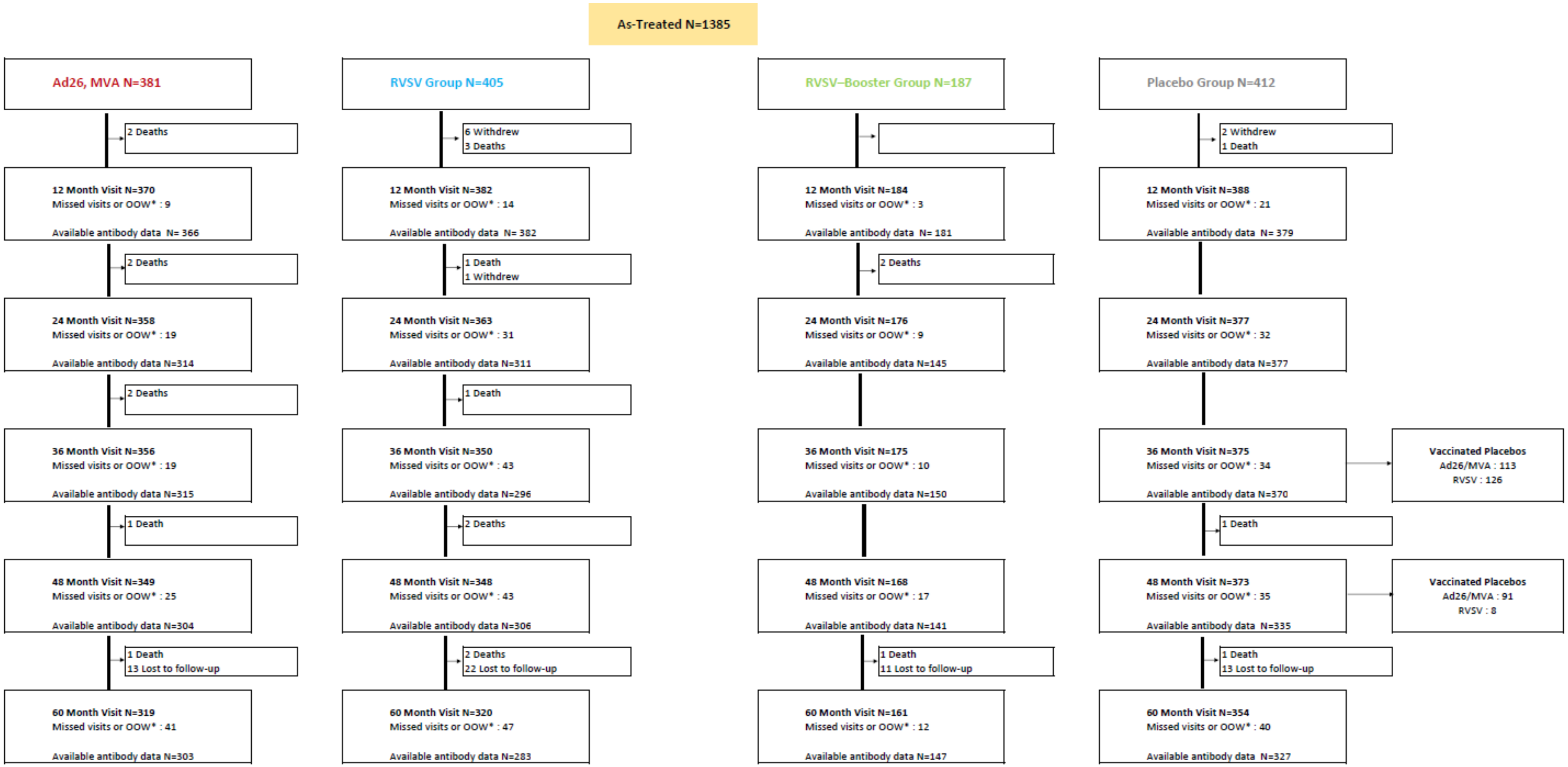

#### Figure S4. CONSORT as-treated Diagram: Children

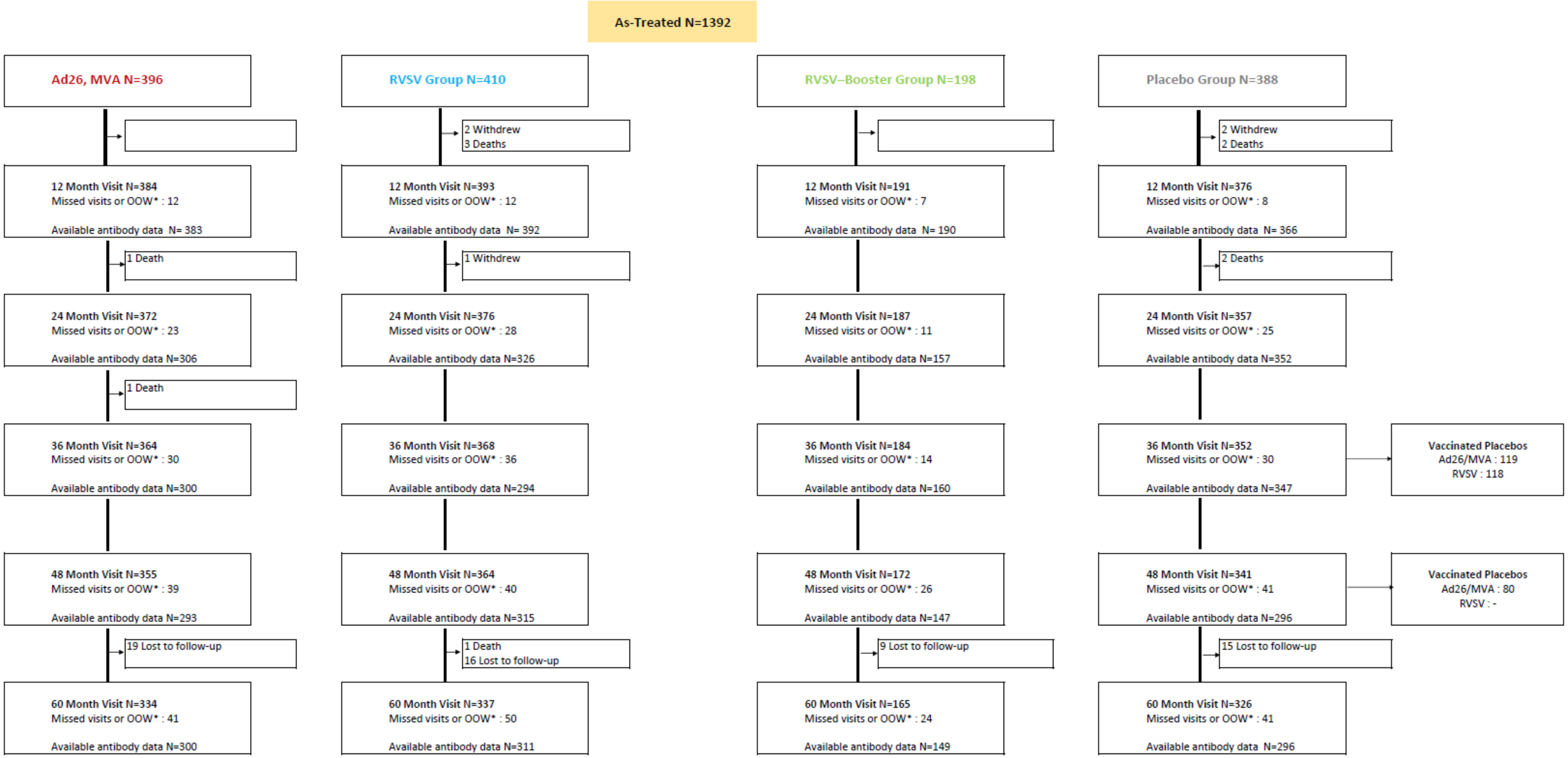

**Table S2. Description of number and reasons of incorrect vaccination and exclusions from the as-treated analysis**

| | Not included in their initial vaccine arm | Excluded from as-treated analysis |
|---|---|---|
| **Adults** | | |
| Ad26, MVA | .. | 16** |
| RVSV Group | .. | .. |
| rVSV–Booster Group | 10* | .. |
| Placebo Group | .. | .. |
| **Children** | | |
| Ad26, MVA | .. | 7** |
| RVSV Group | .. | 1*** |
| rVSV–Booster Group | 4* | .. |
| Placebo Group | .. | 1*** |

* Participants from RVSV-Booster Group who did not receive the booster vaccination and who were included in the RVSV Group in the as-treated-analysis.
** Participants from Ad26, MVA group who did not receive the MVA vaccination.
***Participants from the RVSV and Placebo groups who received a wrong vaccination.

#### Table S3. Visit attendance during long term follow up: Adults

| | Landreah | | Maferinyah | | Redemption | | Mambolo | | UCRC | | CVD | | Total | |
|---|---|---|---|---|---|---|---|---|---|---|---|---|---|---|
| **M24** | | | | | | | | | | | | | | |
| **Visits attended, n (%)** | 281 | | 172 | | 259 | | 397 | | 152 | | 140 | | 1401 | |
| No | 17 | (6.0) | 2 | (1.2) | 24 | (9.3) | 52 | (13.1) | 13 | (8.6) | 8 | (5.7) | 116 | (8.3) |
| Yes | 264 | (94.0) | 170 | (98.8) | 235 | (90.7) | 345 | (86.9) | 139 | (91.4) | 132 | (94.3) | 1285 | (91.7) |
| **Visits attended in window, n (%)** | 281 | | 172 | | 259 | | 397 | | 152 | | 140 | | 1401 | |
| No | 17 | (6.0) | 2 | (1.2) | 24 | (9.3) | 52 | (13.1) | 13 | (8.6) | 8 | (5.7) | 116 | (8.3) |
| Yes | 264 | (94.0) | 170 | (98.8) | 235 | (90.7) | 345 | (86.9) | 139 | (91.4) | 132 | (94.3) | 1285 | (91.7) |
| **M36** | | | | | | | | | | | | | | |
| **Visits attended, n (%)** | 281 | | 172 | | 259 | | 397 | | 152 | | 140 | | 1401 | |
| No | 20 | (7.1) | 13 | (7.6) | 24 | (9.3) | 48 | (12.1) | 23 | (15.1) | 6 | (4.3) | 134 | (9.6) |
| Yes | 261 | (92.9) | 159 | (92.4) | 235 | (90.7) | 349 | (87.9) | 129 | (84.9) | 134 | (95.7) | 1267 | (90.4) |
| **Visits attended in window, n (%)** | 281 | | 172 | | 259 | | 397 | | 152 | | 140 | | 1401 | |
| No | 20 | (7.1) | 13 | (7.6) | 24 | (9.3) | 48 | (12.1) | 23 | (15.1) | 8 | (5.7) | 136 | (9.7) |
| Yes | 261 | (92.9) | 159 | (92.4) | 235 | (90.7) | 349 | (87.9) | 129 | (84.9) | 132 | (94.3) | 1265 | (90.3) |
| **M48** | | | | | | | | | | | | | | |
| **Visits attended, n (%)** | 281 | | 172 | | 259 | | 397 | | 152 | | 140 | | 1401 | |
| No | 27 | (9.6) | 17 | (9.9) | 26 | (10.0) | 49 | (12.3) | 28 | (18.4) | 4 | (2.9) | 151 | (10.8) |
| Yes | 254 | (90.4) | 155 | (90.1) | 233 | (90.0) | 348 | (87.7) | 124 | (81.6) | 136 | (97.1) | 1250 | (89.2) |
| **Visits attended in window, n (%)** | 281 | | 172 | | 259 | | 397 | | 152 | | 140 | | 1401 | |
| No | 27 | (9.6) | 17 | (9.9) | 26 | (10.0) | 50 | (12.6) | 28 | (18.4) | 4 | (2.9) | 152 | (10.8) |
| Yes | 254 | (90.4) | 155 | (90.1) | 233 | (90.0) | 347 | (87.4) | 124 | (81.6) | 136 | (97.1) | 1249 | (89.2) |
| **M60** | | | | | | | | | | | | | | |
| **Visits attended, n (%)** | 281 | | 172 | | 259 | | 397 | | 152 | | 140 | | 1401 | |
| No | 43 | (15.3) | 24 | (14.0) | 37 | (14.3) | 63 | (15.9) | 39 | (25.7) | 4 | (2.9) | 210 | (15.0) |
| Yes | 238 | (84.7) | 148 | (86.0) | 222 | (85.7) | 334 | (84.1) | 113 | (74.3) | 136 | (97.1) | 1191 | (85.0) |
| **Visits attended in window, n (%)** | 281 | | 172 | | 259 | | 397 | | 152 | | 140 | | 1401 | |
| No | 44 | (15.7) | 24 | (14.0) | 38 | (14.7) | 75 | (18.9) | 44 | (28.9) | 14 | (10.0) | 239 | (17.1) |
| Yes | 237 | (84.3) | 148 | (86.0) | 221 | (85.3) | 322 | (81.1) | 108 | (71.1) | 126 | (90.0) | 1162 | (82.9) |

#### Table S4. Visit attendance during the long term follow up: Children

| | Landreah | | Maferinyah | | Redemption | | Mambolo | | UCRC | | CVD | | Total | |
|---|---|---|---|---|---|---|---|---|---|---|---|---|---|---|
| **M24** | | | | | | | | | | | | | | |
| **Visits attended, n (%)** | 285 | | 261 | | 218 | | 311 | | 148 | | 178 | | 1401 | |
| No | 31 | (10.9) | 14 | (5.4) | 19 | (8.7) | 21 | (6.8) | 15 | (10.1) | 5 | (2.8) | 105 | (7.5) |
| Yes | 254 | (89.1) | 247 | (94.6) | 199 | (91.3) | 290 | (93.2) | 133 | (89.9) | 173 | (97.2) | 1296 | (92.5) |
| **Visits attended in window, n (%)** | 285 | | 261 | | 218 | | 311 | | 148 | | 178 | | 1401 | |
| No | 31 | (10.9) | 14 | (5.4) | 19 | (8.7) | 21 | (6.8) | 15 | (10.1) | 5 | (2.8) | 105 | (7.5) |
| Yes | 254 | (89.1) | 247 | (94.6) | 199 | (91.3) | 290 | (93.2) | 133 | (89.9) | 173 | (97.2) | 1296 | (92.5) |
| **M36** | | | | | | | | | | | | | | |
| **Visits attended, n (%)** | 285 | | 261 | | 218 | | 311 | | 148 | | 178 | | 1401 | |
| No | 34 | (11.9) | 21 | (8.0) | 28 | (12.8) | 23 | (7.4) | 16 | (10.8) | 6 | (3.4) | 128 | (9.1) |
| Yes | 251 | (88.1) | 240 | (92.0) | 190 | (87.2) | 288 | (92.6) | 132 | (89.2) | 172 | (96.6) | 1273 | (90.9) |
| **Visits attended in window, n (%)** | 285 | | 261 | | 218 | | 311 | | 148 | | 178 | | 1401 | |
| No | 34 | (11.9) | 21 | (8.0) | 28 | (12.8) | 23 | (7.4) | 16 | (10.8) | 7 | (3.9) | 129 | (9.2) |
| Yes | 251 | (88.1) | 240 | (92.0) | 190 | (87.2) | 288 | (92.6) | 132 | (89.2) | 171 | (96.1) | 1272 | (90.8) |
| **M48** | | | | | | | | | | | | | | |
| **Visits attended, n (%)** | 285 | | 261 | | 218 | | 311 | | 148 | | 178 | | 1401 | |
| No | 46 | (16.1) | 26 | (10.0) | 34 | (15.6) | 27 | (8.7) | 20 | (13.5) | 5 | (2.8) | 158 | (11.3) |
| Yes | 239 | (83.9) | 235 | (90.0) | 184 | (84.4) | 284 | (91.3) | 128 | (86.5) | 173 | (97.2) | 1243 | (88.7) |
| **Visits attended in window, n (%)** | 285 | | 261 | | 218 | | 311 | | 148 | | 178 | | 1401 | |
| No | 46 | (16.1) | 26 | (10.0) | 35 | (16.1) | 27 | (8.7) | 20 | (13.5) | 11 | (6.2) | 165 | (11.8) |
| Yes | 239 | (83.9) | 235 | (90.0) | 183 | (83.9) | 284 | (91.3) | 128 | (86.5) | 167 | (93.8) | 1236 | (88.2) |
| **M60** | | | | | | | | | | | | | | |
| **Visits attended, n (%)** | 285 | | 261 | | 218 | | 311 | | 148 | | 178 | | 1401 | |
| No | 64 | (22.5) | 34 | (13.0) | 38 | (17.4) | 42 | (13.5) | 25 | (16.9) | 4 | (2.2) | 207 | (14.8) |
| Yes | 221 | (77.5) | 227 | (87.0) | 180 | (82.6) | 269 | (86.5) | 123 | (83.1) | 174 | (97.8) | 1194 | (85.2) |
| **Visits attended in window, n (%)** | 285 | | 261 | | 218 | | 311 | | 148 | | 178 | | 1401 | |
| No | 66 | (23.2) | 34 | (13.0) | 38 | (17.4) | 54 | (17.4) | 25 | (16.9) | 19 | (10.7) | 236 | (16.8) |
| Yes | 219 | (76.8) | 227 | (87.0) | 180 | (82.6) | 257 | (82.6) | 123 | (83.1) | 159 | (89.3) | 1165 | (83.2) |

#### Table S5. Description of the observed long-term follow-up duration in months, per protocol-defined visit *

| | Mean | SD | Min | Max |
|---|---|---|---|---|
| **M24** | 26.5 | 1.9 | 21.4 | 30.1 |
| **M36** | 35.7 | 1.7 | 30.4 | 43.3 |
| **M48** | 47.1 | 2.6 | 41.6 | 54.5 |
| **M60** | 58.4 | 1.9 | 53.7 | 63.6 |

*The observed duration can vary from the protocol-defined visit nomenclature due to the window allowed for each visit

#### Table S6. Vaccination rates in the initial placebo group by study sites at the time of the long-term visit

| | Vaccinated participants in initial placebo group, N (%) | | | | | | | |
|---|---|---|---|---|---|---|---|---|
| | Ad26, MVA | | | | rVSV | | | |
| | M24 | M36 | M48 | M60 | M24 | M36 | M48 | M60 |
| **Adults** | | | | | | | | |
| Landreah | .. | .. | 63/80 (79%) | | .. | .. | | |
| Maferinyah | .. | .. | 47/55 (86%) | 1/55 (2%) | .. | .. | | |
| Redemption | .. | .. | | | .. | .. | 66/81 (82%) | 7/81 (9%) |
| Mambolo | .. | .. | 3/110 (3%) | 90/110 (82%) | .. | .. | | |
| UCRC | .. | .. | .. | .. | .. | .. | 25/45 (56%) | |
| CVD | .. | .. | .. | .. | .. | .. | 35/41 (85%) | 1/41 (2%) |
| **Children** | | | | | | | | |
| Landreah | .. | .. | 58/81 (72%) | 2/81 (3%) | .. | .. | .. | .. |
| Maferinyah | .. | .. | 58/69 (84%) | 4/69 (6%) | .. | .. | .. | .. |
| Redemption | .. | .. | .. | .. | .. | .. | 47/55 (86%) | |
| Mambolo | .. | .. | 3/92 (3%) | 74/92 (80%) | .. | .. | .. | .. |
| UCRC | .. | .. | .. | .. | .. | .. | 29/41 (71%) | |
| CVD | .. | .. | .. | .. | .. | .. | 42/50 (84%) | |

#### Table S7. Description of antibody concentration (log10) by visit for the Placebo group prior to censoring due to vaccination

| Median [Q1 - Q3] | M12 | M24 | M36 | M48 | M60 |
|---|---|---|---|---|---|
| **All participants** | 3.91 [3.91 - 4.71] | 3.91 [3.91 - 3.91] | 3.91 [3.91 - 3.91] | 6.36 [3.91 - 8.10] | 6.25 [4.91 - 7.79] |
| **Non-vaccinated participants** | 3.91 [3.91 - 3.91] | 3.91 [3.91 - 3.91] | 3.91 [3.91 - 3.91] | 3.91 [3.91 - 3.91] | 3.91 [3.91 - 3.91] |
| **Vaccinated participants** | 3.91 [3.91 - 4.75] | 3.91 [3.91 - 3.91] | 3.91 [3.91 - 3.91] | 6.83 [3.91 - 8.26] | 6.45 [5.29 - 7.86] |

#### S3.2. Antibody response

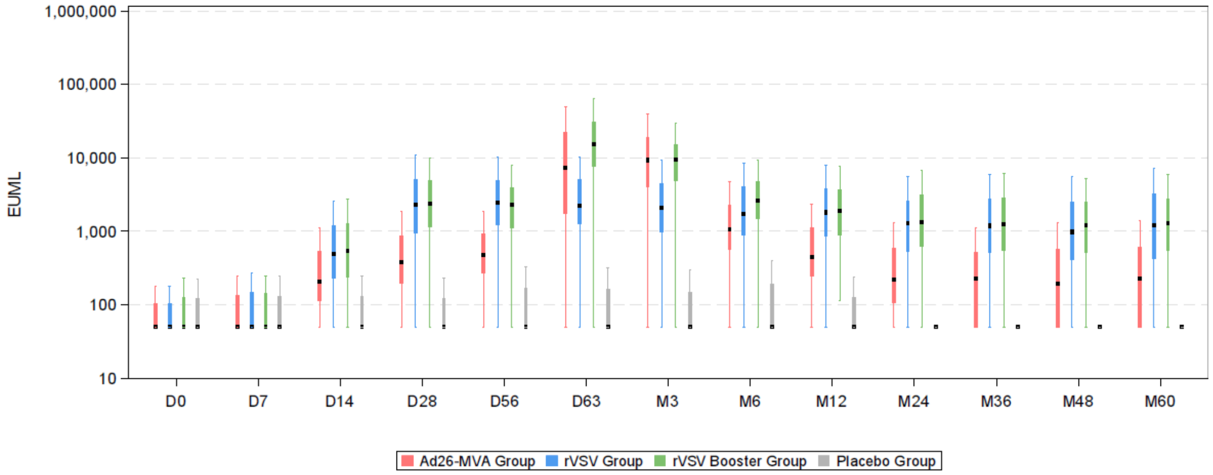

**Figure S5. Distribution of antibody response (box plots) according to vaccine strategy and trial visit: Adults**

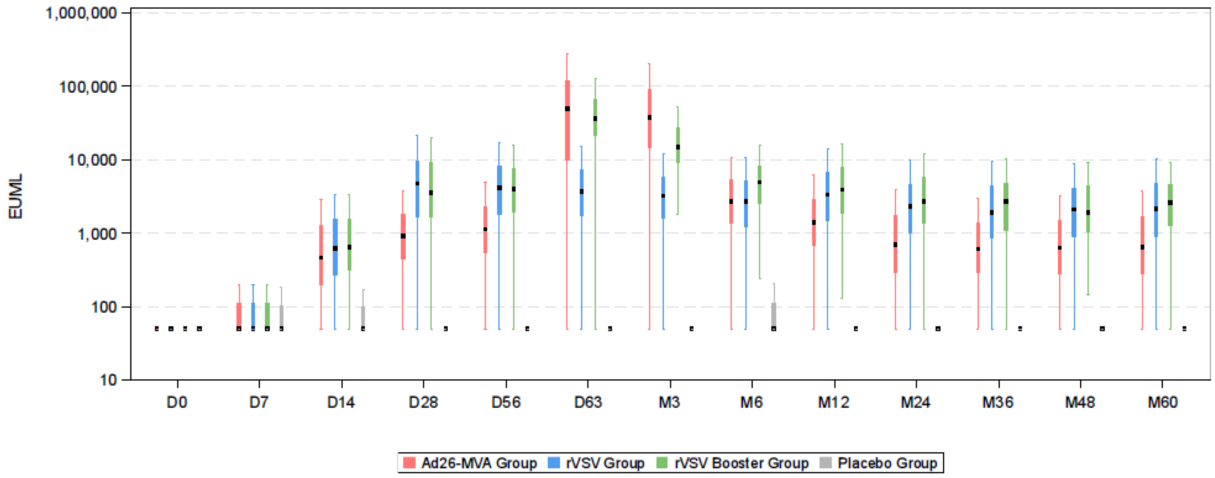

**Figure S6. Distribution of antibody response (box plots) according to vaccine strategy and trial visit: Children**

#### Table S8. Description of the antibody concentration according to vaccine strategy and trial visit: Adults

| | Ad26, MVA Group | | rVSV Group | | rVSV—Booster Group | | Placebo Group | | Total | |
|---|---|---|---|---|---|---|---|---|---|---|
| **Ebola IgG concentration in EU/mL at D0, n** | 394 | | 389 | | 197 | | 410 | | 1390 | |
| Mean (SD) | 228.1 | (1004.5) | 334.6 | (2179.8) | 347.8 | (1182.9) | 587.7 | (7504.9) | 380.9 | (4291.3) |
| Median (IQR) | 50.0 | (50.0;103.3) | 50.0 | (50.0;103.3) | 50.0 | (50.0;122.3) | 50.0 | (50.0;121.5) | 50.0 | (50.0;110.5) |
| Range [min;max] | | [50.0;14112.8] | | [50.0;34809.7] | | [50.0;9751.6] | | [50.0;151542.0] | | [50.0;151542.0] |
| Missing data | 3 | | 6 | | 0 | | 2 | | 11 | |
| **Ebola IgG concentration in EU/mL at D7, n** | 381 | | 376 | | 189 | | 399 | | 1345 | |
| Mean (SD) | 995.2 | (14351.3) | 456.1 | (3307.0) | 855.9 | (5966.2) | 170.3 | (419.4) | 580.2 | (8150.4) |
| Median (IQR) | 50.0 | (50.0;132.7) | 50.0 | (50.0;144.3) | 50.0 | (50.0;141.3) | 50.0 | (50.0;129.3) | 50.0 | (50.0;135.8) |
| Range [min;max] | | [50.0;278396.4] | | [50.0;46115.1] | | [50.0;78578.9] | | [50.0;4704.6] | | [50.0;278396.4] |
| Missing data | 16 | | 19 | | 8 | | 13 | | 56 | |
| **Ebola IgG concentration in EU/mL at D14, n** | 378 | | 381 | | 187 | | 389 | | 1335 | |
| Mean (SD) | 3163.6 | (29565.0) | 2046.4 | (6333.3) | 9598.1 | (94651.3) | 176.4 | (367.7) | 2875.7 | (38939.8) |
| Median (IQR) | 206.4 | (113.5;537.3) | 493.6 | (229.2;1155.4) | 551.0 | (235.8;1285.1) | 50.0 | (50.0;131.0) | 225.9 | (50.0;675.4) |
| Range [min;max] | | [50.0;506474.6] | | [50.0;55829.3] | | [50.0;1283984] | | [50.0;2584.5] | | [50.0;1283984] |
| Missing data | 19 | | 14 | | 10 | | 23 | | 66 | |
| **Ebola IgG concentration in EU/mL at D28, n** | 377 | | 375 | | 188 | | 385 | | 1325 | |
| Mean (SD) | 3116.6 | (35136.5) | 5932.5 | (15641.9) | 11347.0 | (69080.6) | 290.6 | (1780.5) | 4260.2 | (33279.5) |
| Median (IQR) | 380.3 | (192.9;869.3) | 2323.8 | (944.8;5022.8) | 2464.7 | (1181.9;5076.9) | 50.0 | (50.0;121.4) | 531.6 | (116.6;2268.9) |
| Range [min;max] | | [50.0;663234.4] | | [50.0;222234.3] | | [50.0;893704.8] | | [50.0;31277.4] | | [50.0;893704.8] |
| Missing data | 20 | | 20 | | 9 | | 27 | | 76 | |
| **Ebola IgG concentration in EU/mL at D56, n** | 372 | | 364 | | 184 | | 368 | | 1288 | |
| Mean (SD) | 3331.2 | (39445.8) | 5110.4 | (10774.7) | 7413.0 | (48098.5) | 278.3 | (1055.2) | 3544.9 | (28573.6) |
| Median (IQR) | 476.1 | (268.3;916.7) | 2458.8 | (1238.2;4888.0) | 2280.5 | (1119.0;3878.3) | 50.0 | (50.0;164.9) | 669.6 | (165.4;2297.6) |
| Range [min;max] | | [50.0;754383.4] | | [50.0;113583.6] | | [50.0;651003.2] | | [50.0;15123.8] | | [50.0;754383.4] |
| Missing data | 25 | | 31 | | 13 | | 44 | | 113 | |

| | Ad26, MVA Group | | rVSV Group | | rVSV−Booster Group | | Placebo Group | | Total | |
|---|---|---|---|---|---|---|---|---|---|---|
| **Ebola IgG concentration in EU/mL at D63, n** | 364 | | 361 | | 182 | | 357 | | 1264 | |
| Mean (SD) | 19025.2 | (48501.1) | 4833.9 | (9472.7) | 25307.0 | (37852.9) | 287.3 | (993.4) | 10584.4 | (31598.7) |
| Median (IQR) | 7272.3 | (1720.8;21544.2) | 2229.2 | (1260.7;4997.7) | 15474.5 | (7558.8;30943.1) | 50.0 | (50.0;161.9) | 2148.1 | (230.5;9251.7) |
| Range [min;max] | | [50.0;787573.7] | | [50.0;100655.3] | | [50.0;360953.9] | | [50.0;12614.8] | | [50.0;787573.7] |
| Missing data | 33 | | 34 | | 15 | | 55 | | 137 | |
| **Ebola IgG concentration in EU/mL at M3, n** | 361 | | 367 | | 184 | | 363 | | 1275 | |
| Mean (SD) | 18650.9 | (45925.6) | 4174.1 | (7452.3) | 15149.2 | (24319.5) | 402.0 | (2585.2) | 8782.9 | (27538.8) |
| Median (IQR) | 9256.2 | (3890.2;18505.6) | 2071.8 | (988.3;4413.4) | 9485.5 | (4934.5;14935.1) | 50.0 | (50.0;149.7) | 2644.5 | (234.5;8955.5) |
| Range [min;max] | | [50.0;727601.5] | | [50.0;81083.3] | | [50.0;246580.3] | | [50.0;46013.7] | | [50.0;727601.5] |
| Missing data | 36 | | 28 | | 13 | | 49 | | 126 | |
| **Ebola IgG concentration in EU/mL at M6, n** | 370 | | 376 | | 184 | | 361 | | 1291 | |
| Mean (SD) | 4736.2 | (53566.8) | 3335.3 | (4811.3) | 5609.3 | (16614.7) | 458.2 | (1695.3) | 3256.4 | (29514.1) |
| Median (IQR) | 1062.9 | (518.4;2186.5) | 1744.7 | (905.1;4060.0) | 2564.9 | (1481.8;4720.0) | 50.0 | (50.0;192.5) | 994.3 | (224.5;2578.2) |
| Range [min;max] | | [50.0;1030908] | | [50.0;43014.0] | | [50.0;201588.5] | | [50.0;17351.5] | | [50.0;1030908] |
| Missing data | 27 | | 19 | | 13 | | 51 | | 110 | |
| **Ebola IgG concentration in EU/mL at M12, n** | 376 | | 378 | | 185 | | 379 | | 1318 | |
| Mean (SD) | 1150.0 | (2450.5) | 3385.5 | (5325.2) | 3432.9 | (5520.2) | 252.1 | (1294.2) | 1853.4 | (4058.2) |
| Median (IQR) | 434.6 | (234.4;1072.9) | 1825.3 | (876.5;3789.3) | 1894.2 | (867.8;3634.8) | 50.0 | (50.0;126.0) | 551.9 | (126.0;1896.8) |
| Range [min;max] | | [50.0;28205.8] | | [50.0;42677.4] | | [112.9;58602.4] | | [50.0;23789.8] | | [50.0;58602.4] |
| Missing data | 21 | | 17 | | 12 | | 33 | | 83 | |
| **Ebola IgG concentration in EU/mL at M24, n** | 325 | | 307 | | 149 | | 377 | | 1158 | |
| Mean (SD) | 744.3 | (2941.7) | 2184.6 | (2630.6) | 2782.3 | (5726.8) | 139.2 | (425.0) | 1191.4 | (3084.2) |
| Median (IQR) | 208.2 | (50.0;535.4) | 1311.3 | (532.8;2603.0) | 1321.3 | (618.9;3155.2) | 50.0 | (50.0;50.0) | 245.4 | (50.0;1193.6) |
| Range [min;max] | | [50.0;48762.6] | | [50.0;19288.2] | | [50.0;63643.0] | | [50.0;4607.1] | | [50.0;63643.0] |
| Missing data | 72 | | 88 | | 48 | | 35 | | 243 | |
| **Ebola IgG concentration in EU/mL at M36, n** | 323 | | 293 | | 153 | | 370 | | 1139 | |
| Mean (SD) | 809.1 | (2468.3) | 2738.9 | (6271.0) | 2199.3 | (2748.8) | 172.7 | (860.4) | 1285.5 | (3766.6) |
| Median (IQR) | 221.4 | (50.0;505.7) | 1191.8 | (520.4;2724.3) | 1243.6 | (559.1;2884.9) | 50.0 | (50.0;50.0) | 263.9 | (50.0;1134.9) |
| Range [min;max] | | [50.0;33556.3] | | [50.0;78305.5] | | [50.0;18446.3] | | [50.0;13536.7] | | [50.0;78305.5] |

| | Ad26, MVA Group | | rVSV Group | | rVSV−Booster Group | | Placebo Group | | Total | |
|---|---|---|---|---|---|---|---|---|---|---|
| Missing data | 74 | | 102 | | 44 | | 42 | | 262 | |
| **Ebola IgG concentration in EU/mL at M48, n** | 315 | | 302 | | 145 | | 335 | | 1097 | |
| Mean (SD) | 625.1 | (1502.5) | 1830.1 | (2216.3) | 2020.9 | (2381.4) | 2018.5 | (4142.1) | 1566.9 | (2886.7) |
| Median (IQR) | 192.3 | (50.0;561.4) | 986.8 | (416.9;2479.8) | 1210.0 | (506.1;2483.5) | 271.6 | (50.0;2392.5) | 510.1 | (132.8;1767.0) |
| Range [min;max] | | [50.0;19221.2] | | [50.0;17333.1] | | [50.0;12534.8] | | [50.0;49501.2] | | [50.0;49501.2] |
| Missing data | 82 | | 93 | | 52 | | 77 | | 304 | |
| **Ebola IgG concentration in EU/mL at M60, n** | 311 | | 281 | | 149 | | 327 | | 1068 | |
| Mean (SD) | 806.8 | (1869.6) | 2426.3 | (3385.2) | 2821.8 | (7359.6) | 1787.8 | (5794.6) | 1814.4 | (4725.6) |
| Median (IQR) | 229.3 | (50.0;608.5) | 1200.7 | (430.4;3150.5) | 1280.3 | (547.6;2788.8) | 364.4 | (50.0;1267.7) | 518.9 | (157.6;1861.0) |
| Range [min;max] | | [50.0;17239.0] | | [50.0;29186.9] | | [50.0;85012.0] | | [50.0;87205.2] | | [50.0;87205.2] |
| Missing data | 86 | | 114 | | 48 | | 85 | | 333 | |

**Table S9. Description of the antibody concentration according to vaccine strategy and trial visit: Children**

| | Ad26, MVA Group | | rVSV Group | | rVSV– Booster Group | | Placebo Group | | Total | |
|---|---|---|---|---|---|---|---|---|---|---|
| **Ebola IgG concentration in EU/mL at D0, n** | 398 | | 401 | | 200 | | 386 | | 1385 | |
| Mean (SD) | 159.9 | (643.0) | 242.1 | (1402.8) | 463.3 | (4482.8) | 219.5 | (958.1) | 244.1 | (1959.9) |
| Median (IQR) | 50.0 | (50.0;50.0) | 50.0 | (50.0;50.0) | 50.0 | (50.0;50.0) | 50.0 | (50.0;50.0) | 50.0 | (50.0;50.0) |
| Range [min;max] | | [50.0;10238.2] | | [50.0;24250.9] | | [50.0;63193.4] | | [50.0;15587.8] | | [50.0;63193.4] |
| Missing data | 5 | | 6 | | 2 | | 3 | | 16 | |
| **Ebola IgG concentration in EU/mL at D7, n** | 382 | | 390 | | 189 | | 368 | | 1329 | |
| Mean (SD) | 5849.6 | (111307.7) | 371.6 | (2953.3) | 420.9 | (3673.9) | 451.8 | (3712.4) | 1975.4 | (59739.6) |
| Median (IQR) | 50.0 | (50.0;110.4) | 50.0 | (50.0;111.3) | 50.0 | (50.0;111.7) | 50.0 | (50.0;102.9) | 50.0 | (50.0;109.8) |
| Range [min;max] | | [50.0;2175628] | | [50.0;55862.3] | | [50.0;50193.7] | | [50.0;65776.8] | | [50.0;2175628] |
| Missing data | 21 | | 17 | | 13 | | 21 | | 72 | |
| **Ebola IgG concentration in EU/mL at D14, n** | 387 | | 398 | | 196 | | 374 | | 1355 | |
| Mean (SD) | 7209.4 | (116316.3) | 2066.3 | (7413.2) | 1783.5 | (5523.2) | 422.7 | (4676.7) | 3040.6 | (62377.1) |
| Median (IQR) | 468.1 | (200.2;1249.6) | 617.9 | (266.2;1526.2) | 662.0 | (312.6;1581.0) | 50.0 | (50.0;100.1) | 338.9 | (50.0;1031.1) |
| Range [min;max] | | [50.0;2288889] | | [50.0;97316.6] | | [50.0;70566.0] | | [50.0;90118.7] | | [50.0;2288889] |
| Missing data | 16 | | 9 | | 6 | | 15 | | 46 | |
| **Ebola IgG concentration in EU/mL at D28, n** | 393 | | 396 | | 191 | | 373 | | 1353 | |
| Mean (SD) | 4526.6 | (53831.4) | 9354.1 | (17323.3) | 7656.9 | (11623.9) | 206.5 | (793.4) | 5190.4 | (30983.1) |
| Median (IQR) | 930.7 | (442.3;1780.2) | 4710.0 | (1669.1;9740.4) | 3659.2 | (1652.3;9178.3) | 50.0 | (50.0;50.0) | 1121.4 | (120.8;4113.0) |
| Range [min;max] | | [50.0;1063113] | | [50.0;217006.5] | | [50.0;94747.6] | | [50.0;9769.4] | | [50.0;1063113] |
| Missing data | 10 | | 11 | | 11 | | 16 | | 48 | |
| **Ebola IgG concentration in EU/mL at D56, n** | 395 | | 398 | | 195 | | 364 | | 1352 | |
| Mean (SD) | 3478.7 | (17646.5) | 6290.7 | (7858.0) | 5940.3 | (6330.8) | 381.3 | (3393.4) | 3827.6 | (11112.0) |
| Median (IQR) | 1135.1 | (547.7;2305.5) | 4140.2 | (1830.3;8071.5) | 3988.0 | (1939.1;7634.9) | 50.0 | (50.0;75.1) | 1307.9 | (162.0;4100.6) |
| Range [min;max] | | [50.0;296383.4] | | [50.0;74013.5] | | [50.0;38521.2] | | [50.0;63412.4] | | [50.0;296383.4] |
| Missing data | 8 | | 9 | | 7 | | 25 | | 49 | |
| **Ebola IgG concentration in EU/mL at D63, n** | 386 | | 399 | | 188 | | 360 | | 1333 | |
| Mean (SD) | 106586.3 | (168719.7) | 6450.0 | (11200.4) | 56036.0 | (61624.7) | 772.9 | (6941.2) | 40906.9 | (104330.6) |

| | Ad26, MVA Group | | rVSV Group | | rVSV– Booster Group | | Placebo Group | | Total | |
|---|---|---|---|---|---|---|---|---|---|---|
| Median (IQR) | 49740.2 | (9907.4;116512.9) | 3705.7 | (1718.6;7164.7) | 36660.2 | (21356.5;65255.5) | 50.0 | (50.0;50.0) | 4827.6 | (280.6;34907.4) |
| Range [min;max] | | [50.0;1153668] | | [50.0;121207.2] | | [50.0;429759.6] | | [50.0;111677.4] | | [50.0;1153668] |
| Missing data | 17 | | 8 | | 14 | | 29 | | 68 | |
| **Ebola IgG concentration in EU/mL at M3, n** | 384 | | 395 | | 196 | | 366 | | 1341 | |
| Mean (SD) | 80437.2 | (140111.1) | 5119.7 | (7486.8) | 23654.2 | (27438.1) | 212.1 | (758.4) | 28056.7 | (83024.8) |
| Median (IQR) | 38118.3 | (14719.8;89268.1) | 3243.2 | (1587.0;5818.0) | 14900.6 | (9189.5;27036.6) | 50.0 | (50.0;50.0) | 4737.5 | (203.3;21289.4) |
| Range [min;max] | | [50.0;1605717] | | [50.0;75480.1] | | [1806.1;241992.4] | | [50.0;8991.8] | | [50.0;1605717] |
| Missing data | 19 | | 12 | | 6 | | 23 | | 60 | |
| **Ebola IgG concentration in EU/mL at M6, n** | 387 | | 393 | | 193 | | 346 | | 1319 | |
| Mean (SD) | 4811.4 | (8724.2) | 4825.2 | (12560.2) | 7270.7 | (8850.5) | 321.1 | (1367.1) | 3997.5 | (9307.4) |
| Median (IQR) | 2717.9 | (1360.2;5279.8) | 2725.0 | (1235.4;5223.2) | 4899.0 | (2539.2;8122.4) | 50.0 | (50.0;112.9) | 1939.8 | (232.2;4700.8) |
| Range [min;max] | | [50.0;129038.8] | | [50.0;231890.5] | | [241.3;65641.4] | | [50.0;17525.4] | | [50.0;231890.5] |
| Missing data | 16 | | 14 | | 9 | | 43 | | 82 | |
| **Ebola IgG concentration in EU/mL at M12, n** | 386 | | 391 | | 191 | | 367 | | 1335 | |
| Mean (SD) | 2974.2 | (6457.4) | 6126.9 | (10172.8) | 6947.6 | (9062.7) | 170.0 | (555.8) | 3695.2 | (7803.1) |
| Median (IQR) | 1372.6 | (683.8;2896.4) | 3358.3 | (1513.3;6630.1) | 3953.6 | (1871.2;7811.7) | 50.0 | (50.0;50.0) | 1401.6 | (159.2;3986.7) |
| Range [min;max] | | [50.0;81886.6] | | [50.0;95909.8] | | [131.0;56324.0] | | [50.0;5645.6] | | [50.0;95909.8] |
| Missing data | 17 | | 16 | | 11 | | 22 | | 66 | |
| **Ebola IgG concentration in EU/mL at M24, n** | 307 | | 325 | | 158 | | 353 | | 1143 | |
| Mean (SD) | 1386.7 | (1891.9) | 5804.6 | (15145.1) | 4809.4 | (6803.5) | 597.2 | (8721.0) | 2872.2 | (10049.9) |
| Median (IQR) | 700.2 | (287.8;1748.9) | 2303.5 | (1015.6;4578.0) | 2796.9 | (1381.5;5832.1) | 50.0 | (50.0;50.0) | 703.4 | (50.0;2663.2) |
| Range [min;max] | | [50.0;15555.6] | | [50.0;156282.2] | | [50.0;49583.3] | | [50.0;163757.8] | | [50.0;163757.8] |
| Missing data | 96 | | 82 | | 44 | | 36 | | 258 | |
| **Ebola IgG concentration in EU/mL at M36, n** | 303 | | 293 | | 161 | | 348 | | 1105 | |
| Mean (SD) | 1368.2 | (2638.5) | 3444.8 | (5132.7) | 5152.4 | (11051.9) | 179.7 | (981.9) | 2095.9 | (5479.6) |
| Median (IQR) | 604.9 | (290.5;1352.2) | 1930.9 | (853.3;4373.8) | 2678.1 | (1111.4;4725.9) | 50.0 | (50.0;50.0) | 604.9 | (50.0;2174.3) |
| Range [min;max] | | [50.0;31454.6] | | [50.0;56585.1] | | [50.0;101286.2] | | [50.0;16865.9] | | [50.0;101286.2] |
| Missing data | 100 | | 114 | | 41 | | 41 | | 296 | |
| **Ebola IgG concentration in EU/mL at M48, n** | 295 | | 314 | | 148 | | 297 | | 1054 | |

| | Ad26, MVA Group | | rVSV Group | | rVSV− Booster Group | | Placebo Group | | Total | |
|---|---|---|---|---|---|---|---|---|---|---|
| Mean (SD) | 1664.1 | (3898.0) | 3765.2 | (7194.3) | 4289.4 | (8720.6) | 4337.0 | (11540.8) | 3411.9 | (8302.2) |
| Median (IQR) | 639.4 | (281.2;1518.2) | 2102.7 | (891.2;4153.7) | 1927.3 | (1045.9;4251.0) | 1269.3 | (50.0;4673.5) | 1352.7 | (445.3;3424.1) |
| Range [min;max] | | [50.0;34344.9] | | [50.0;69465.8] | | [144.7;74660.4] | | [50.0;141685.9] | | [50.0;141685.9] |
| Missing data | 108 | | 93 | | 54 | | 92 | | 347 | |
| **Ebola IgG concentration in EU/mL at M60, n** | 301 | | 310 | | 150 | | 297 | | 1058 | |
| Mean (SD) | 1504.4 | (2740.2) | 4750.1 | (12727.0) | 4208.6 | (7032.1) | 3200.1 | (6292.7) | 3314.8 | (8318.5) |
| Median (IQR) | 650.0 | (282.1;1668.4) | 2140.2 | (890.0;4814.7) | 2589.0 | (1259.5;4512.8) | 888.1 | (248.1;3446.4) | 1311.5 | (427.7;3461.6) |
| Range [min;max] | | [50.0;31367.5] | | [50.0;176053.8] | | [50.0;77414.2] | | [50.0;62438.3] | | [50.0;176053.8] |
| Missing data | 102 | | 97 | | 52 | | 92 | | 343 | |

**Table S10. Geometric mean antibody concentrations and their 95% confidence interval according to as-treated vaccine strategy and trial visit - Adults**

| | Visit | Geometric Mean | 95% CI | Geometric Mean ($Log_{10}$) | 95% CI |
|---|---|---|---|---|---|
| Ad26, MVA Group | D0 | 78.56 | [71.35 ; 86.49] | 1.86 | [1.83 ; 1.90] |
| | D7 | 88.73 | [79.66 ; 98.84] | 1.91 | [1.87 ; 1.95] |
| | D14 | 276.53 | [238.12 ; 321.14] | 2.37 | [2.31 ; 2.43] |
| | D28 | 435.64 | [382.54 ; 496.11] | 2.58 | [2.53 ; 2.64] |
| | D56 | 537.94 | [479.27 ; 603.79] | 2.69 | [2.64 ; 2.74] |
| | D63 | 5819.26 | [4867.71 ; 6956.81] | 3.68 | [3.60 ; 3.77] |
| | M3 | 8632.60 | [7566.55 ; 9848.85] | 3.89 | [3.83 ; 3.96] |
| | M6 | 1139.61 | [1012.29 ; 1282.95] | 3.02 | [2.96 ; 3.07] |
| | M12 | 514.70 | [455.17 ; 582.03] | 2.66 | [2.61 ; 2.72] |
| | M24 | 250.03 | [216.29 ; 289.03] | 2.33 | [2.27 ; 2.39] |
| | M36 | 253.31 | [218.01 ; 294.33] | 2.34 | [2.28 ; 2.40] |
| | M48 | 224.38 | [193.47 ; 260.22] | 2.29 | [2.23 ; 2.35] |
| | M60 | 252.32 | [215.11 ; 295.95] | 2.33 | [2.26 ; 2.39] |
| rVSV Group | D0 | 78.18 | [71.06 ; 86.01] | 1.86 | [1.83 ; 1.89] |
| | D7 | 94.81 | [85.59 ; 105.01] | 1.94 | [1.90 ; 1.98] |
| | D14 | 574.54 | [500.44 ; 659.60] | 2.70 | [2.64 ; 2.76] |
| | D28 | 2298.24 | [2011.94 ; 2625.26] | 3.31 | [3.25 ; 3.37] |
| | D56 | 2434.11 | [2161.03 ; 2741.69] | 3.35 | [3.30 ; 3.40] |
| | D63 | 2390.23 | [2121.84 ; 2692.57] | 3.34 | [3.29 ; 3.39] |
| | M3 | 2057.14 | [1820.62 ; 2324.38] | 3.27 | [3.22 ; 3.33] |
| | M6 | 1763.01 | [1568.59 ; 1981.53] | 3.20 | [3.15 ; 3.26] |
| | M12 | 1715.51 | [1519.43 ; 1936.89] | 3.19 | [3.13 ; 3.24] |
| | M24 | 1159.80 | [1010.74 ; 1330.85] | 3.01 | [2.95 ; 3.08] |
| | M36 | 1144.62 | [984.37 ; 1330.97] | 3.00 | [2.93 ; 3.07] |
| | M48 | 955.98 | [832.89 ; 1097.25] | 2.93 | [2.87 ; 2.99] |
| | M60 | 1098.97 | [936.58 ; 1289.53] | 2.98 | [2.91 ; 3.05] |
| rVSV─Booster Group | D0 | 93.57 | [79.06 ; 110.74] | 1.92 | [1.86 ; 1.98] |
| | D7 | 100.59 | [83.29 ; 121.48] | 1.95 | [1.88 ; 2.01] |
| | D14 | 599.75 | [482.91 ; 744.85] | 2.71 | [2.62 ; 2.80] |
| | D28 | 2311.75 | [1897.24 ; 2816.83] | 3.31 | [3.22 ; 3.40] |
| | D56 | 2239.06 | [1899.27 ; 2639.64] | 3.31 | [3.24 ; 3.39] |
| | D63 | 13530.00 | [11291.78 ; 16211.88] | 4.09 | [4.01 ; 4.18] |
| | M3 | 8903.92 | [7631.59 ; 10388.36] | 3.92 | [3.84 ; 3.99] |
| | M6 | 2728.36 | [2342.90 ; 3177.24] | 3.41 | [3.34 ; 3.47] |
| | M12 | 1894.73 | [1616.73 ; 2220.54] | 3.24 | [3.17 ; 3.31] |
| | M24 | 1242.58 | [996.80 ; 1548.95] | 3.03 | [2.93 ; 3.14] |
| | M36 | 1182.67 | [976.95 ; 1431.72] | 3.03 | [2.94 ; 3.12] |
| | M48 | 1051.23 | [855.78 ; 1291.32] | 2.97 | [2.88 ; 3.07] |
| | M60 | 1216.29 | [995.22 ; 1486.47] | 3.04 | [2.95 ; 3.13] |
| Placebo Group | D0 | 87.27 | [78.71 ; 96.76] | 1.90 | [1.86 ; 1.94] |
| | D7 | 84.14 | [77.05 ; 91.87] | 1.89 | [1.86 ; 1.93] |
| | D14 | 87.22 | [79.46 ; 95.75] | 1.91 | [1.87 ; 1.94] |
| | D28 | 84.61 | [76.71 ; 93.31] | 1.89 | [1.86 ; 1.93] |
| | D56 | 102.28 | [91.86 ; 113.89] | 1.97 | [1.93 ; 2.01] |
| | D63 | 98.40 | [87.86 ; 110.19] | 1.95 | [1.91 ; 1.99] |
| | M3 | 95.51 | [84.99 ; 107.32] | 1.93 | [1.89 ; 1.97] |
| | M6 | 113.63 | [99.93 ; 129.21] | 2.00 | [1.95 ; 2.05] |
| | M12 | 87.18 | [78.77 ; 96.50] | 1.90 | [1.86 ; 1.94] |
| | M24 | 67.90 | [62.66 ; 73.58] | 1.81 | [1.78 ; 1.84] |
| | M36 | 65.87 | [60.64 ; 71.56] | 1.79 | [1.77 ; 1.82] |

| | Visit | Geometric Mean | 95% CI | Geometric Mean ($Log_{10}$) | 95% CI |
|---|---|---|---|---|---|
| | M48 | 68.13 | [62.31 ; 74.50] | 1.81 | [1.78 ; 1.84] |
| | M60 | 68.89 | [62.79 ; 75.58] | 1.81 | [1.78 ; 1.84] |

**Table S11. Geometric mean antibody concentrations and their 95% confidence interval according to as-treated vaccine strategy and trial visit - Children**

| | Visit | Geometric Mean | 95% CI | Geometric Mean ($Log_{10}$) | 95% CI |
|---|---|---|---|---|---|
| Ad26, MVA Group | D0 | 71.37 | [65.72 ; 77.52] | 1.83 | [1.80 ; 1.86] |
| | D7 | 79.00 | [71.47 ; 87.32] | 1.86 | [1.83 ; 1.90] |
| | D14 | 512.10 | [445.29 ; 588.95] | 2.64 | [2.59 ; 2.70] |
| | D28 | 889.44 | [790.43 ; 1000.85] | 2.90 | [2.85 ; 2.96] |
| | D56 | 1138.56 | [1014.98 ; 1277.18] | 3.02 | [2.97 ; 3.07] |
| | D63 | 35444.61 | [29782.00 ; 42183.87] | 4.48 | [4.40 ; 4.56] |
| | M3 | 34386.18 | [29733.46 ; 39766.96] | 4.49 | [4.42 ; 4.56] |
| | M6 | 2515.70 | [2230.90 ; 2836.86] | 3.36 | [3.30 ; 3.41] |
| | M12 | 1440.00 | [1284.26 ; 1614.63] | 3.12 | [3.07 ; 3.17] |
| | M24 | 672.36 | [581.34 ; 777.62] | 2.77 | [2.70 ; 2.83] |
| | M36 | 629.43 | [546.99 ; 724.30] | 2.75 | [2.68 ; 2.81] |
| | M48 | 636.63 | [547.72 ; 739.97] | 2.74 | [2.68 ; 2.81] |
| | M60 | 644.67 | [554.77 ; 749.14] | 2.75 | [2.68 ; 2.82] |
| rVSV Group | D0 | 74.98 | [68.52 ; 82.04] | 1.84 | [1.81 ; 1.87] |
| | D7 | 82.16 | [74.34 ; 90.80] | 1.88 | [1.84 ; 1.91] |
| | D14 | 645.41 | [563.09 ; 739.78] | 2.74 | [2.68 ; 2.80] |
| | D28 | 3999.29 | [3478.61 ; 4597.90] | 3.54 | [3.48 ; 3.61] |
| | D56 | 3508.99 | [3113.45 ; 3954.77] | 3.50 | [3.44 ; 3.56] |
| | D63 | 3393.48 | [3020.01 ; 3813.13] | 3.49 | [3.43 ; 3.54] |
| | M3 | 2845.08 | [2533.26 ; 3195.27] | 3.41 | [3.36 ; 3.47] |
| | M6 | 2352.72 | [2072.78 ; 2670.47] | 3.32 | [3.26 ; 3.38] |
| | M12 | 3065.91 | [2714.24 ; 3463.15] | 3.44 | [3.38 ; 3.50] |
| | M24 | 2142.19 | [1843.26 ; 2489.60] | 3.27 | [3.20 ; 3.34] |
| | M36 | 1695.29 | [1458.66 ; 1970.32] | 3.17 | [3.10 ; 3.24] |
| | M48 | 1816.69 | [1580.67 ; 2087.95] | 3.21 | [3.14 ; 3.27] |
| | M60 | 1982.33 | [1716.06 ; 2289.92] | 3.25 | [3.18 ; 3.31] |
| rVSV—Booster Group | D0 | 71.29 | [62.27 ; 81.62] | 1.82 | [1.78 ; 1.87] |
| | D7 | 80.26 | [69.84 ; 92.23] | 1.87 | [1.82 ; 1.92] |
| | D14 | 672.44 | [560.89 ; 806.18] | 2.77 | [2.69 ; 2.85] |
| | D28 | 3606.14 | [2985.09 ; 4356.41] | 3.51 | [3.42 ; 3.60] |
| | D56 | 3564.08 | [3035.85 ; 4184.23] | 3.51 | [3.44 ; 3.59] |
| | D63 | 34495.32 | [29443.37 ; 40414.10] | 4.51 | [4.43 ; 4.59] |
| | M3 | 16064.93 | [14237.42 ; 18127.02] | 4.19 | [4.14 ; 4.24] |
| | M6 | 4664.30 | [4072.81 ; 5341.68] | 3.64 | [3.59 ; 3.71] |
| | M12 | 3891.84 | [3330.82 ; 4547.34] | 3.56 | [3.49 ; 3.63] |
| | M24 | 2637.37 | [2196.85 ; 3166.23] | 3.38 | [3.30 ; 3.47] |
| | M36 | 2327.37 | [1910.58 ; 2835.08] | 3.32 | [3.23 ; 3.41] |
| | M48 | 2182.14 | [1828.66 ; 2603.96] | 3.31 | [3.23 ; 3.38] |
| | M60 | 2346.59 | [1950.51 ; 2823.09] | 3.33 | [3.25 ; 3.42] |
| Placebo Group | D0 | 77.61 | [70.56 ; 85.37] | 1.86 | [1.82 ; 1.89] |
| | D7 | 79.95 | [71.85 ; 88.97] | 1.86 | [1.83 ; 1.90] |
| | D14 | 79.25 | [71.62 ; 87.69] | 1.86 | [1.83 ; 1.90] |

| Visit | Geometric Mean | 95% CI | Geometric Mean ($Log_{10}$) | 95% CI |
|---|---|---|---|---|
| D28 | 73.45 | [66.83 ; 80.73] | 1.83 | [1.80 ; 1.87] |
| D56 | 78.89 | [71.15 ; 87.47] | 1.86 | [1.83 ; 1.90] |
| D63 | 82.79 | [73.63 ; 93.09] | 1.87 | [1.84 ; 1.91] |
| M3 | 75.62 | [68.58 ; 83.39] | 1.84 | [1.81 ; 1.88] |
| M6 | 86.60 | [77.16 ; 97.19] | 1.89 | [1.85 ; 1.93] |
| M12 | 74.71 | [68.29 ; 81.73] | 1.84 | [1.81 ; 1.88] |
| M24 | 67.40 | [61.59 ; 73.75] | 1.80 | [1.77 ; 1.83] |
| M36 | 67.59 | [62.00 ; 73.69] | 1.80 | [1.78 ; 1.83] |
| M48 | 69.94 | [63.72 ; 76.78] | 1.82 | [1.78 ; 1.85] |
| M60 | 71.70 | [64.99 ; 79.10] | 1.82 | [1.79 ; 1.86] |

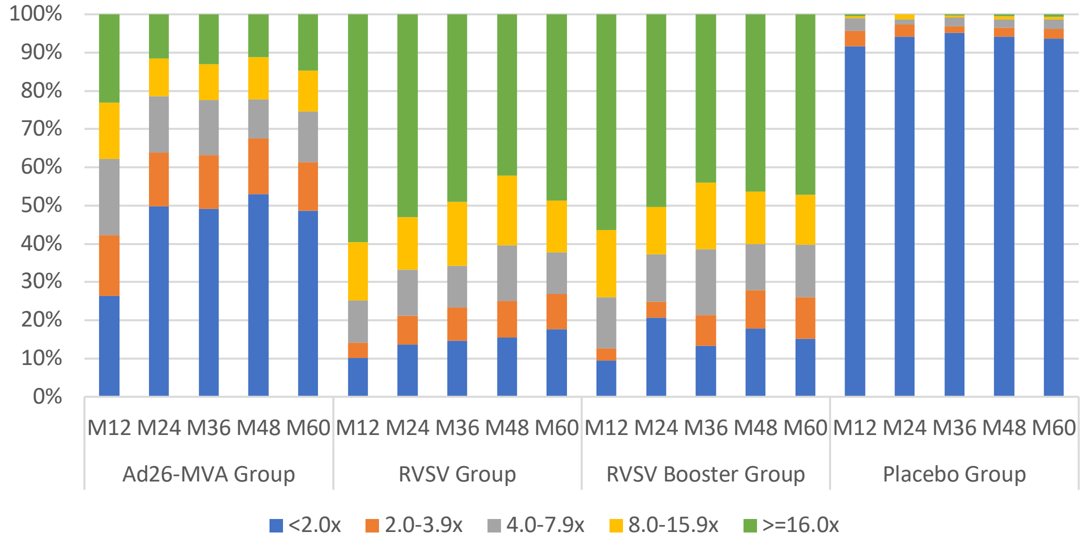

**Figure S7. Proportions of participants with different categories of increase (fold-change) in antibody concentration from baseline, according to vaccine strategy and long-term trial visit: Adults**

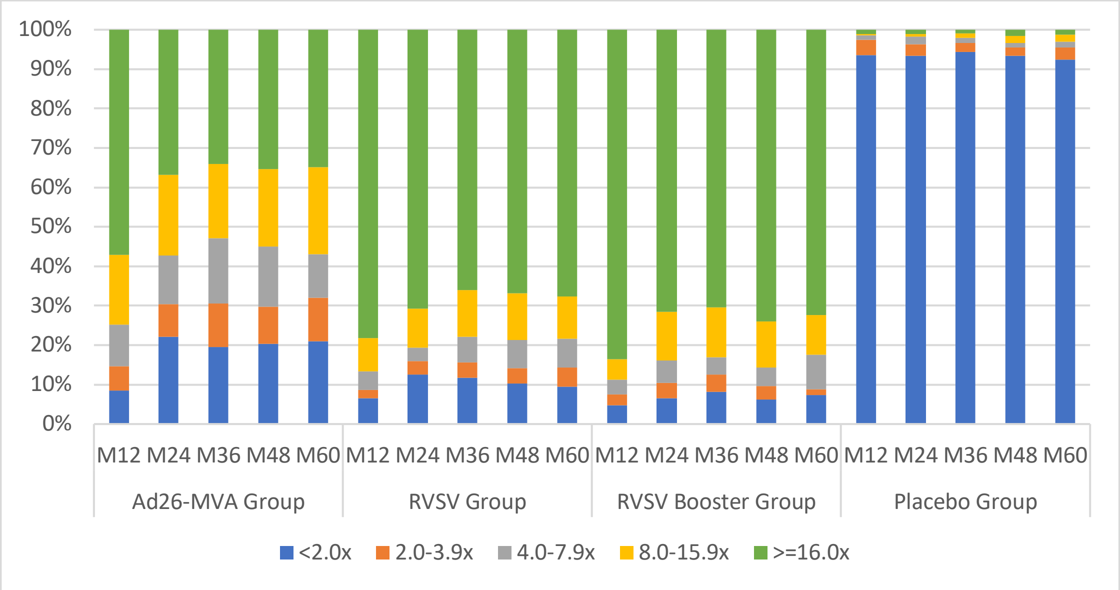

**Figure S8. Proportions of participants with different categories of increase (fold-change) in antibody concentration from baseline, according to vaccine strategy and long-term trial visit: Children**

**Table S12. Number and proportions of participants with different categories of increase (fold-change) in antibody concentration from baseline, according to vaccine strategy and annual trial visit: Adults**

| | Ad26, MVA Group | | rVSV Group | | rVSV—Booster Group | | Placebo Group | |
|---|---|---|---|---|---|---|---|---|
| **M12, n (%)** | 364 | | 376 | | 181 | | 377 | |
| <2.0x | 96 | (26.4) | 38 | (10.1) | 17 | (9.4) | 345 | (91.5) |
| 2.0-3.9x | 58 | (15.9) | 15 | (4.0) | 6 | (3.3) | 15 | (4.0) |
| 4.0-7.9x | 72 | (19.8) | 42 | (11.2) | 24 | (13.3) | 13 | (3.4) |
| 8.0-15.9x | 54 | (14.8) | 57 | (15.2) | 32 | (17.7) | 2 | (0.5) |
| >=16.0x | 84 | (23.1) | 224 | (59.6) | 102 | (56.4) | 2 | (0.5) |
| **M24, n (%)** | 312 | | 307 | | 145 | | 375 | |
| <2.0x | 155 | (49.7) | 42 | (13.7) | 30 | (20.7) | 353 | (94.1) |
| 2.0-3.9x | 44 | (14.1) | 23 | (7.5) | 6 | (4.1) | 12 | (3.2) |
| 4.0-7.9x | 46 | (14.7) | 37 | (12.1) | 18 | (12.4) | 5 | (1.3) |
| 8.0-15.9x | 31 | (9.9) | 42 | (13.7) | 18 | (12.4) | 5 | (1.3) |
| >=16.0x | 36 | (11.5) | 163 | (53.1) | 73 | (50.3) | 0 | (0.0) |
| **M36, n (%)** | 313 | | 292 | | 150 | | 368 | |
| <2.0x | 154 | (49.2) | 43 | (14.7) | 20 | (13.3) | 350 | (95.1) |
| 2.0-3.9x | 44 | (14.1) | 25 | (8.6) | 12 | (8.0) | 6 | (1.6) |
| 4.0-7.9x | 45 | (14.4) | 32 | (11.0) | 26 | (17.3) | 9 | (2.4) |
| 8.0-15.9x | 29 | (9.3) | 49 | (16.8) | 26 | (17.3) | 2 | (0.5) |
| >=16.0x | 41 | (13.1) | 143 | (49.0) | 66 | (44.0) | 1 | (0.3) |
| **M48, n (%)** | 302 | | 303 | | 140 | | 369 | |
| <2.0x | 160 | (53.0) | 47 | (15.5) | 25 | (17.9) | 347 | (94.0) |
| 2.0-3.9x | 44 | (14.6) | 29 | (9.6) | 14 | (10.0) | 9 | (2.4) |
| 4.0-7.9x | 31 | (10.3) | 44 | (14.5) | 17 | (12.1) | 8 | (2.2) |
| 8.0-15.9x | 33 | (10.9) | 55 | (18.2) | 19 | (13.6) | 3 | (0.8) |
| >=16.0x | 34 | (11.3) | 128 | (42.2) | 65 | (46.4) | 2 | (0.5) |
| **M60, n (%)** | 300 | | 279 | | 146 | | 359 | |
| <2.0x | 146 | (48.7) | 49 | (17.6) | 22 | (15.1) | 336 | (93.6) |
| 2.0-3.9x | 38 | (12.7) | 26 | (9.3) | 16 | (11.0) | 9 | (2.5) |
| 4.0-7.9x | 40 | (13.3) | 30 | (10.8) | 20 | (13.7) | 9 | (2.5) |
| 8.0-15.9x | 32 | (10.7) | 38 | (13.6) | 19 | (13.0) | 3 | (0.8) |
| >=16.0x | 44 | (14.7) | 136 | (48.7) | 69 | (47.3) | 2 | (0.6) |

**Table S13. Number and proportions of participants with different categories of increase (fold-change) in antibody concentration from baseline, according to vaccine strategy and annual trial visit: Children**

| | Ad26, MVA Group | | rVSV Group | | rVSV—Booster Group | | Placebo Group | |
|---|---|---|---|---|---|---|---|---|
| **M12, n (%)** | 378 | | 386 | | 188 | | 363 | |
| <2.0x | 32 | (8.5) | 25 | (6.5) | 9 | (4.8) | 340 | (93.7) |
| 2.0-3.9x | 23 | (6.1) | 8 | (2.1) | 5 | (2.7) | 14 | (3.9) |
| 4.0-7.9x | 40 | (10.6) | 18 | (4.7) | 7 | (3.7) | 4 | (1.1) |
| 8.0-15.9x | 67 | (17.7) | 33 | (8.5) | 10 | (5.3) | 1 | (0.3) |
| >=16.0x | 216 | (57.1) | 302 | (78.2) | 157 | (83.5) | 4 | (1.1) |
| **M24, n (%)** | 302 | | 321 | | 155 | | 350 | |
| <2.0x | 67 | (22.2) | 40 | (12.5) | 10 | (6.5) | 327 | (93.4) |
| 2.0-3.9x | 25 | (8.3) | 11 | (3.4) | 6 | (3.9) | 10 | (2.9) |
| 4.0-7.9x | 37 | (12.3) | 11 | (3.4) | 9 | (5.8) | 7 | (2.0) |
| 8.0-15.9x | 62 | (20.5) | 32 | (10.0) | 19 | (12.3) | 2 | (0.6) |
| >=16.0x | 111 | (36.8) | 227 | (70.7) | 111 | (71.6) | 4 | (1.1) |
| **M36, n (%)** | 297 | | 288 | | 159 | | 345 | |
| <2.0x | 58 | (19.5) | 34 | (11.8) | 13 | (8.2) | 326 | (94.5) |
| 2.0-3.9x | 33 | (11.1) | 11 | (3.8) | 7 | (4.4) | 8 | (2.3) |
| 4.0-7.9x | 49 | (16.5) | 19 | (6.6) | 7 | (4.4) | 4 | (1.2) |
| 8.0-15.9x | 56 | (18.9) | 34 | (11.8) | 20 | (12.6) | 4 | (1.2) |
| >=16.0x | 101 | (34.0) | 190 | (66.0) | 112 | (70.4) | 3 | (0.9) |
| **M48, n (%)** | 289 | | 310 | | 146 | | 333 | |
| <2.0x | 59 | (20.4) | 32 | (10.3) | 9 | (6.2) | 311 | (93.4) |
| 2.0-3.9x | 27 | (9.3) | 12 | (3.9) | 5 | (3.4) | 7 | (2.1) |
| 4.0-7.9x | 44 | (15.2) | 22 | (7.1) | 7 | (4.8) | 4 | (1.2) |
| 8.0-15.9x | 57 | (19.7) | 37 | (11.9) | 17 | (11.6) | 6 | (1.8) |
| >=16.0x | 102 | (35.3) | 207 | (66.8) | 108 | (74.0) | 5 | (1.5) |
| **M60, n (%)** | 296 | | 305 | | 148 | | 326 | |
| <2.0x | 62 | (20.9) | 29 | (9.5) | 11 | (7.4) | 301 | (92.3) |
| 2.0-3.9x | 33 | (11.1) | 15 | (4.9) | 2 | (1.4) | 10 | (3.1) |
| 4.0-7.9x | 33 | (11.1) | 22 | (7.2) | 13 | (8.8) | 5 | (1.5) |
| 8.0-15.9x | 65 | (22.0) | 33 | (10.8) | 15 | (10.1) | 6 | (1.8) |
| >=16.0x | 103 | (34.8) | 206 | (67.5) | 107 | (72.3) | 4 | (1.2) |

### S3.3. Factors associated with the antibody concentration

S3.3.1. Description of the antibody concentrations over time by vaccine strategy, according to age, sex, country and antibody level at baseline

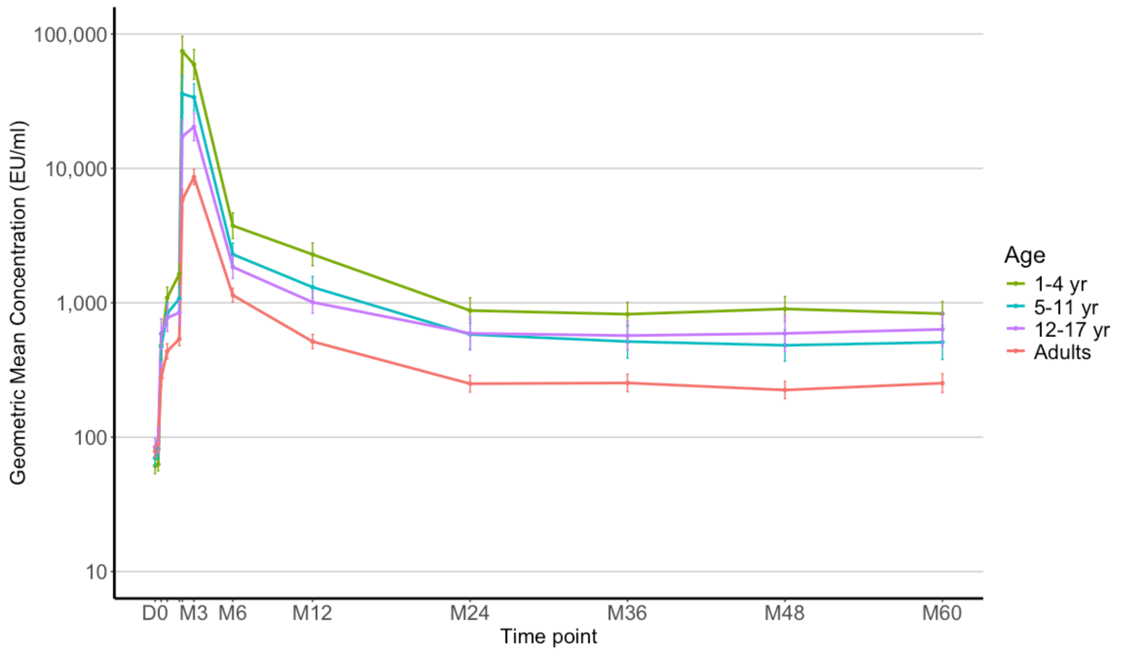

**Figure S9. Antibody concentrations (Geometric Mean Concentrations) over time in the Ad26, MVA group (adults and children), according to age category**
I bars indicate 95% confidence intervals.

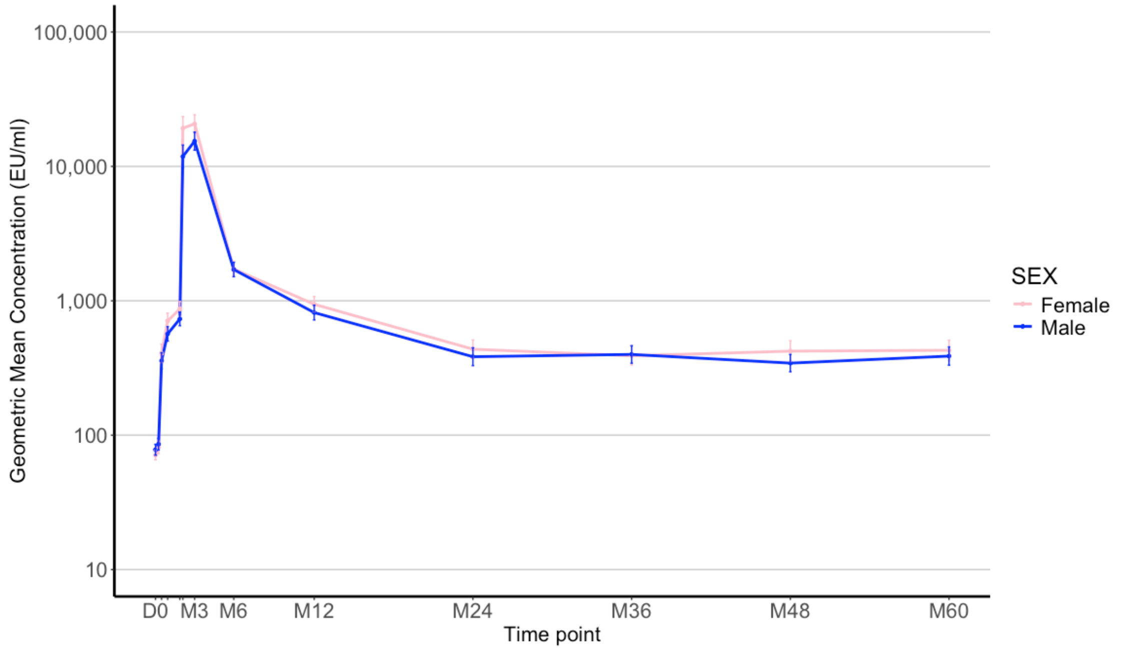

**Figure S10. Antibody concentrations (Geometric Mean Concentrations) over time in the Ad26, MVA strategy (adults and children combined), according to sex**

Ι bars indicate 95% confidence intervals.

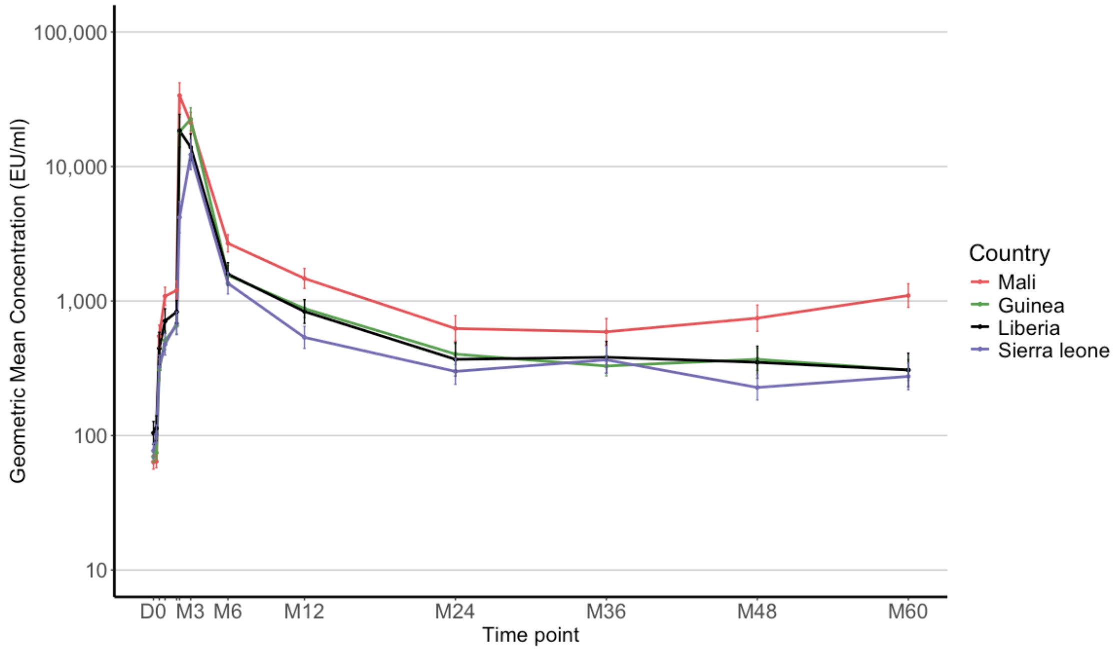

**Figure S11. Antibody concentrations (Geometric Mean Concentrations) over time in the Ad26, MVA group (adults and children combined), according to country**

I bars indicate 95% confidence intervals.

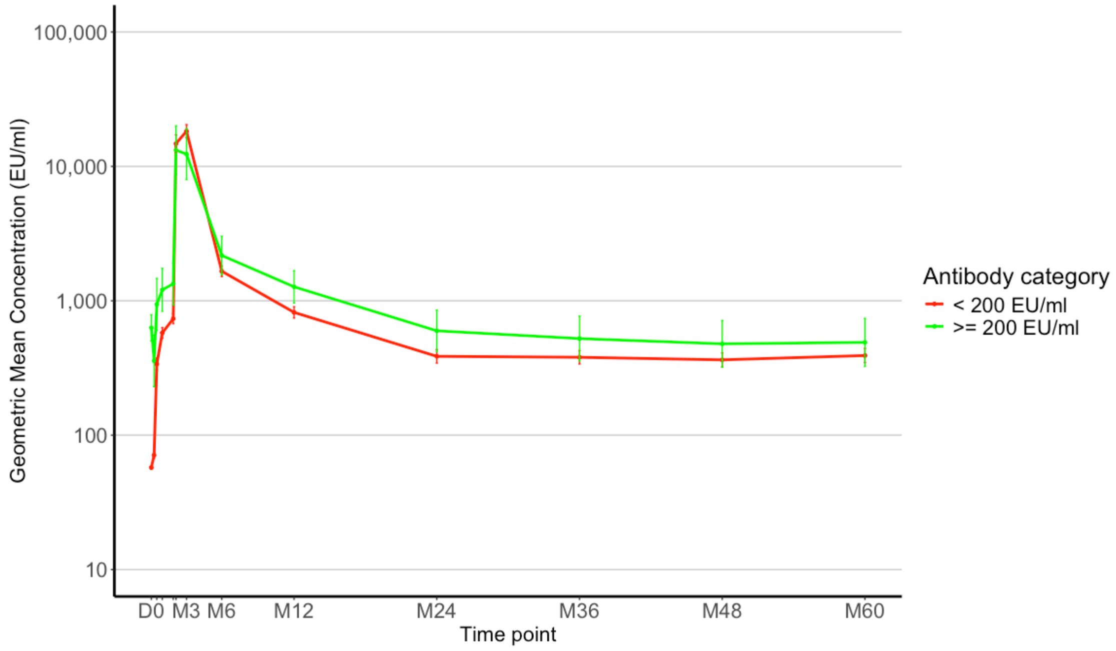

**Figure S12. Antibody concentrations (Geometric Mean Concentrations) over time in the Ad26, MVA group (adults and children combined), according to category of antibody at baseline**

I bars indicate 95% confidence intervals.

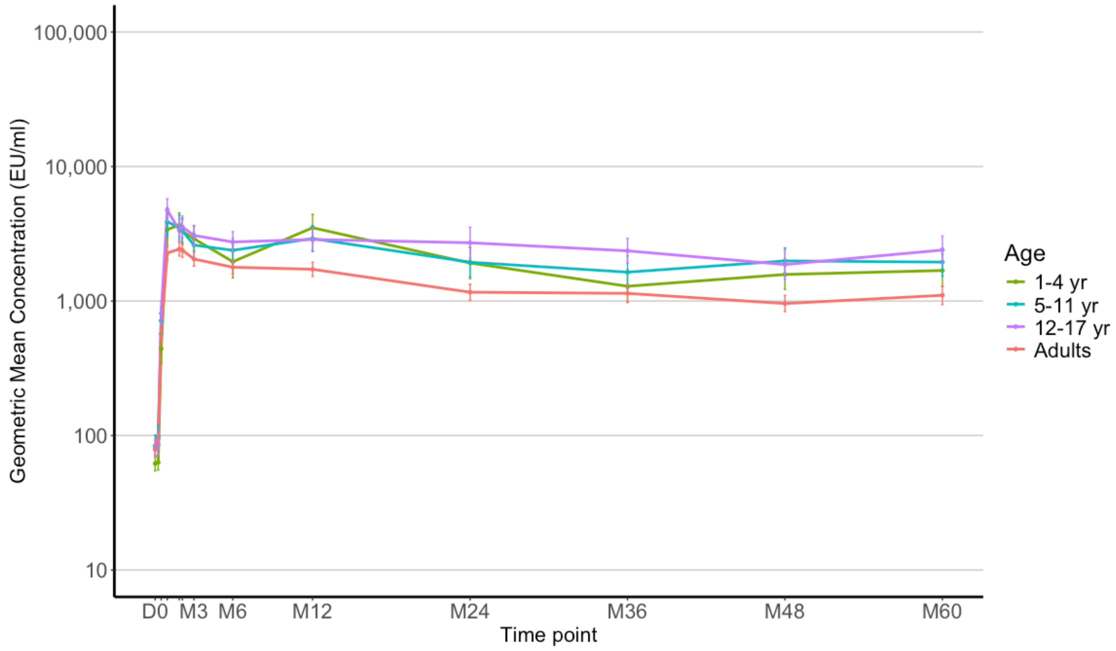

**Figure S13. Antibody concentrations (Geometric Mean Concentrations) over time in the rVSV group (adults and children combined), according to age category**

I bars indicate 95% confidence intervals.

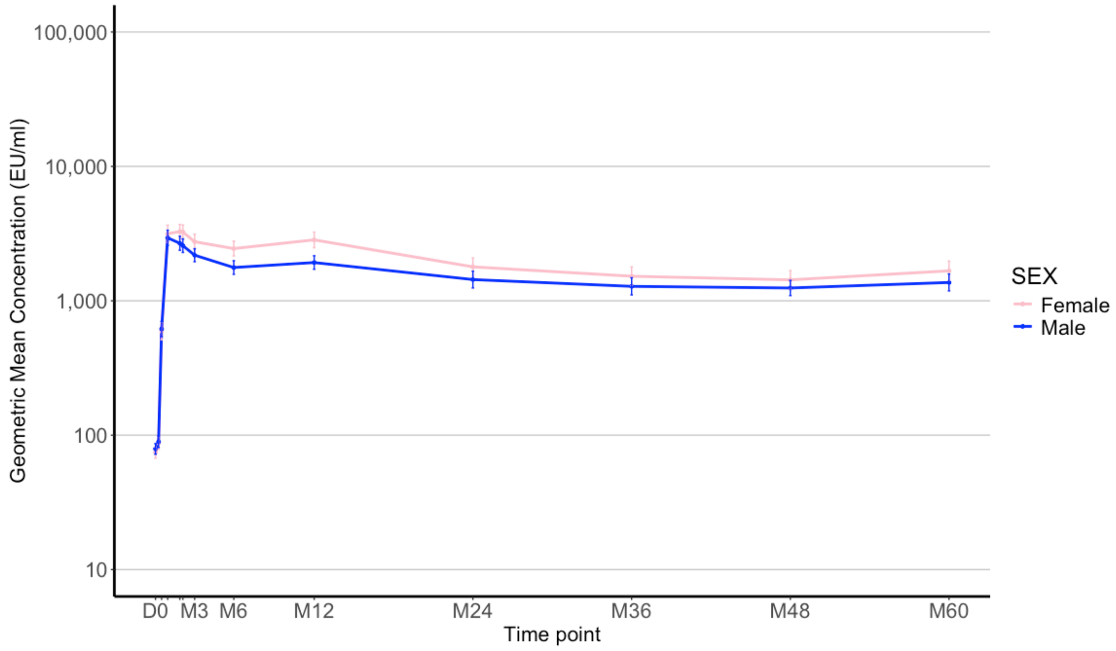

**Figure S14. Antibody concentrations (Geometric Mean Concentrations) over time in the rVSV group (adults and children combined), according to sex**

I bars indicate 95% confidence intervals.

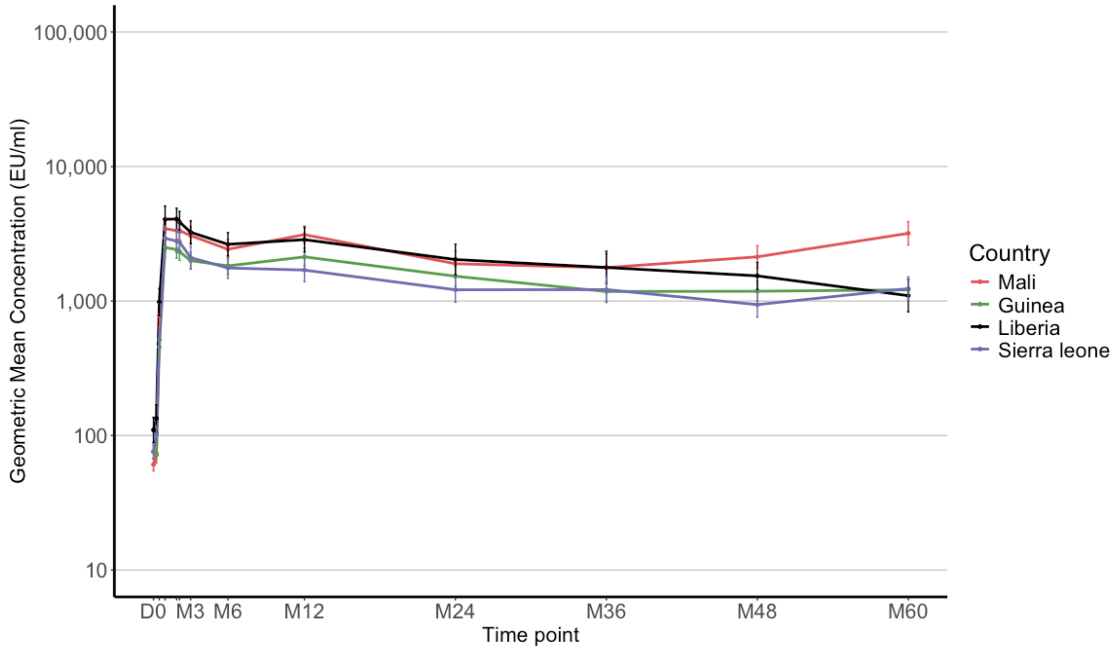

**Figure S15. Antibody concentrations (Geometric Mean Concentrations) over time in the rVSV group (adults and children combined), according to country**

I bars indicate 95% confidence intervals.

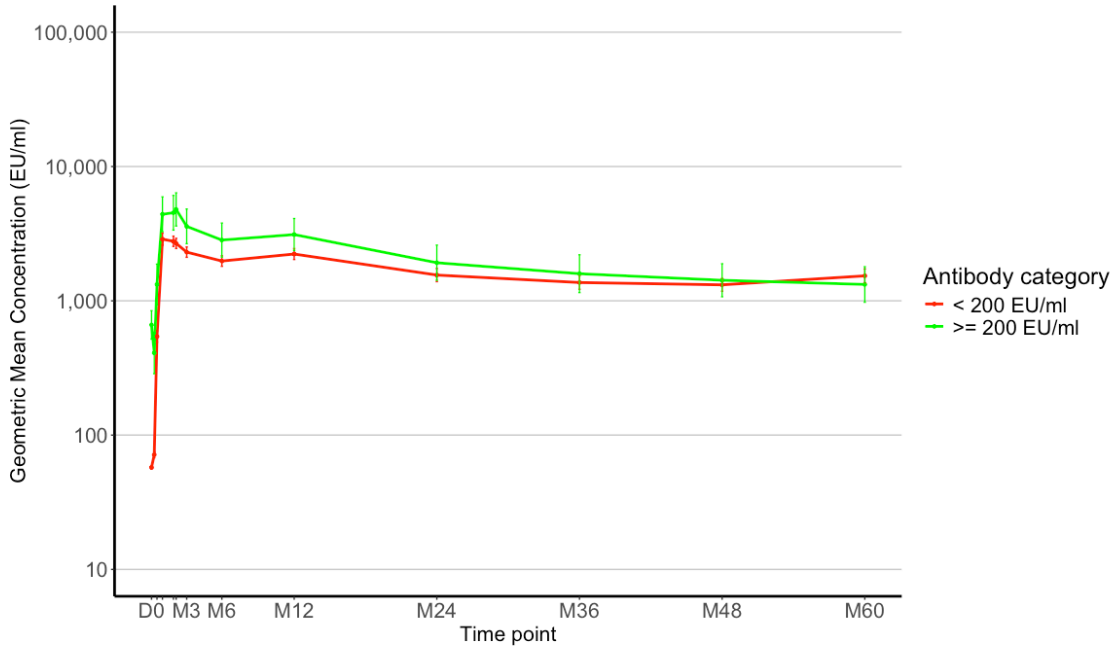

**Figure S16. Antibody concentrations (Geometric Mean Concentrations) over time in the rVSV group (adults and children combined), according to category of antibody at baseline**

I bars indicate 95% confidence intervals.

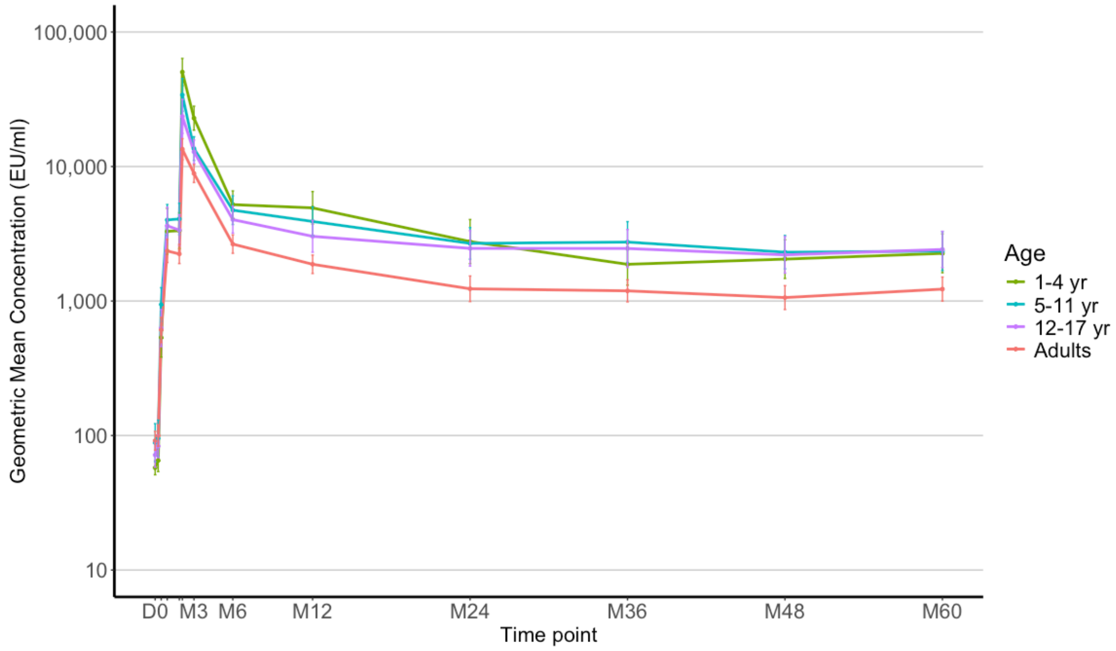

**Figure S17. Antibody concentrations (Geometric Mean Concentrations) over time in the rVSV-booster group (adults and children ombined), according to age category**

I bars indicate 95% confidence intervals.

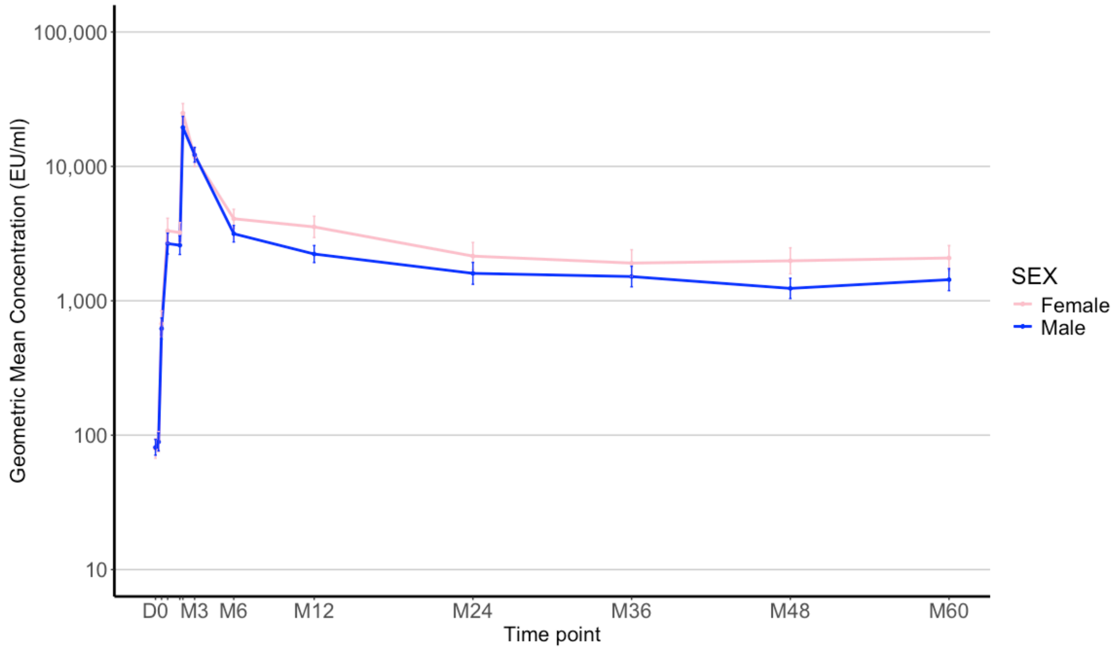

**Figure S18. Antibody concentrations (Geometric Mean Concentrations) over time in the rVSV-booster group (adults and children combined), according to sex**

I bars indicate 95% confidence intervals.

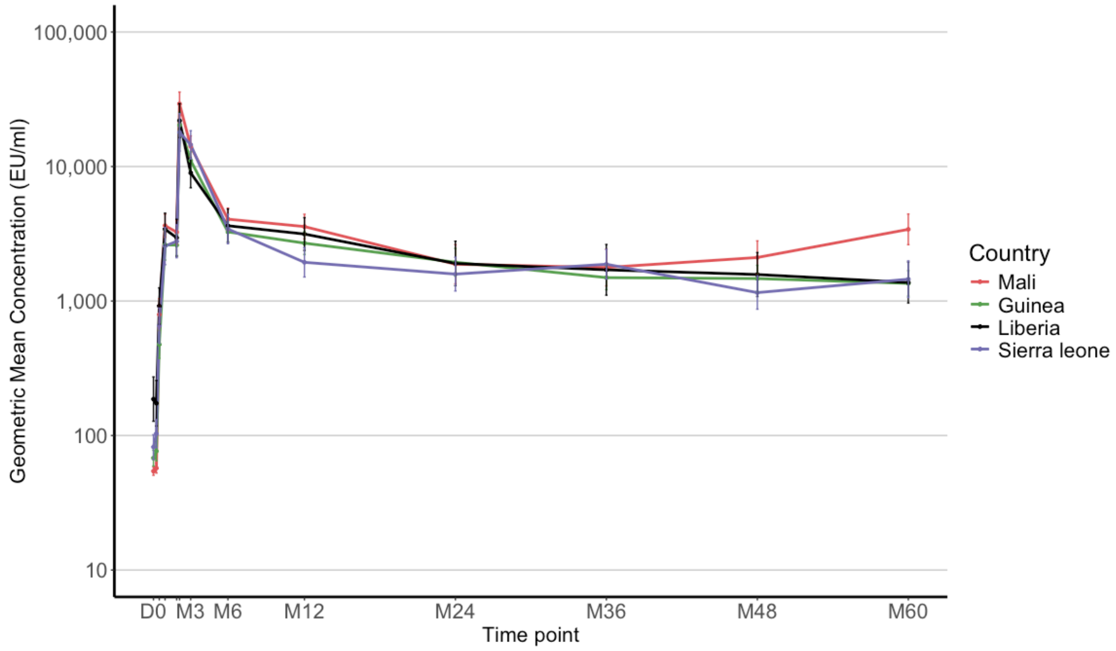

**Figure S19. Antibody concentrations (Geometric Mean Concentrations) over time in the rVSV-booster group (adults and children combined), according to country**

I bars indicate 95% confidence intervals.

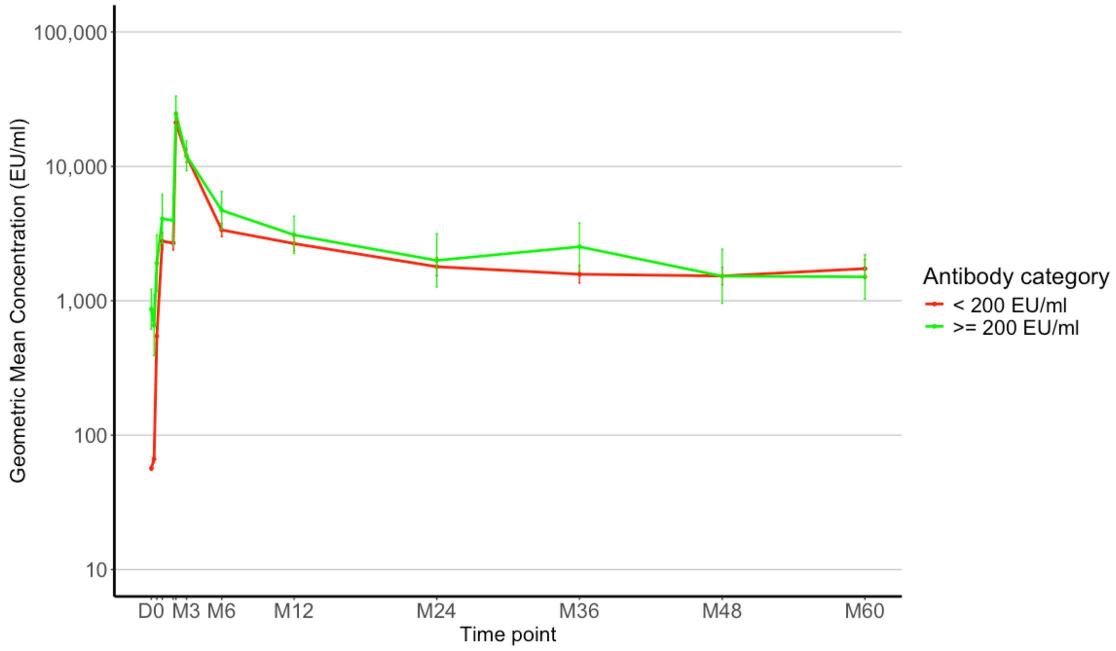

**Figure S20. Antibody concentrations (Geometric Mean Concentrations) over time in the rVSV-booster group (adults and children combined), according to category of antibody at baseline**

I bars indicate 95% confidence intervals.

#### S3.3.2. Linear mixed effect regression model output

| | Estimate | Standard error | P-value |
|---|---|---|---|
| **Fixed effects** | | | |
| $\beta_0$ (intercept) | 2.161e+00 | 1.015e-01 | < 0.0001 |
| $\beta_1$ (slope) | 7.036e-02 | 1.164e-02 | < 0.0001 |
| $\beta_2$ (rVSV arm) | 7.831e-01 | 1.380e-01 | < 0.0001 |
| $\beta_3$(rVSV-booster arm) | 5.312e-01 | 1.638e-01 | 0.001 |
| $\beta_4$ (rVSV arm, slope) | 5.394e-03 | 1.666e-02 | 0.746 |
| $\beta_5$ (rVSV-booster arm, slope) | 3.532e-02 | 2.037e-02 | 0.083 |
| $\beta_6$ (1–4 yr) | 5.020e-01 | 5.193e-02 | < 0.0001 |
| $\beta_7$ (5–11 yr) | 3.325e-01 | 5.162e-02 | < 0.0001 |
| $\beta_8$ (12–17 yr) | 3.822e-01 | 4.981e-02 | < 0.0001 |
| $\beta_9$ (1–4 yr, rVSV arm) | -3.357e-01 | 7.372e-02 | < 0.0001 |
| $\beta_{10}$ (5–11 yr, rVSV arm) | -9.755e-02 | 7.111e-02 | 0.170 |
| $\beta_{11}$ (12–17 yr, rVSV arm) | -7.859e-02 | 7.106e-02 | 0.269 |
| $\beta_{12}$ (1–4 yr, rVSV-booster arm) | -2.044e-01 | 8.875e-02 | 0.021 |
| $\beta_{13}$ (5–11 yr, rVSV-booster arm) | 2.321e-03 | 8.804e-02 | 0.979 |
| $\beta_{14}$ (12–17 yr, rVSV-booster arm) | -5.219e-03 | 8.776e-02 | 0.953 |
| $\beta_{15}$ (female) | 1.582e-02 | 3.637e-02 | 0.664 |
| $\beta_{16}$ (female, rVSV arm) | 7.805e-02 | 5.131e-02 | 0.129 |
| $\beta_{17}$ (female, rVSV-booster arm) | 1.477e-01 | 6.377e-02 | 0.021 |
| $\beta_{18}$ (Guinea) | -1.795e-01 | 5.238e-02 | 0.001 |
| $\beta_{19}$ (Liberia) | -1.663e-01 | 6.371e-02 | 0.009 |
| $\beta_{20}$ (Sierra Leone) | -1.918e-01 | 5.815e-02 | 0.001 |
| $\beta_{21}$ (Guinea, rVSV arm) | 8.438e-02 | 7.436e-02 | 0.257 |
| $\beta_{22}$ (Liberia, rVSV arm) | 2.200e-01 | 9.038e-02 | 0.015 |
| $\beta_{23}$ (Sierra Leone, rVSV arm) | 3.593e-02 | 8.199e-02 | 0.661 |
| $\beta_{24}$ (Guinea, rVSV-booster arm) | 2.164e-01 | 9.135e-02 | 0.018 |
| $\beta_{25}$ (Liberia, rVSV-booster arm) | 1.639e-01 | 1.140e-01 | 0.151 |
| $\beta_{26}$ (Sierra Leone, rVSV-booster arm) | 2.200e-01 | 1.006e-01 | 0.029 |
| $\beta_{27}$ (Guinea, slope) | -9.562e-02 | 1.499e-02 | < 0.0001 |
| $\beta_{28}$ (Liberia, slope) | -1.027e-01 | 1.808e-02 | < 0.0001 |
| $\beta_{29}$ (Sierra Leone, slope) | -9.803e-02 | 1.610e-02 | < 0.0001 |
| $\beta_{30}$ (Guinea, rVSV arm, slope) | -9.293e-03 | 2.137e-02 | 0.664 |
| $\beta_{31}$ (Guinea, rVSV-booster arm, slope) | -5.378e-02 | 2.612e-02 | 0.035 |
| $\beta_{32}$ (Liberia, rVSV arm, slope) | -5.417e-02 | 2.566e-02 | 0.520 |
| $\beta_{33}$ (Liberia, rVSV-booster arm, slope) | -5.042e-02 | 3.110e-02 | 0.040 |
| $\beta_{34}$ (Sierra Leone, rVSV arm, slope) | 1.473e-02 | 2.291e-02 | 0.105 |
| $\beta_{35}$ (Sierra Leone, rVSV-booster arm, slope) | -3.130e-02 | 2.848e-02 | 0.272 |
| $\beta_{36}$ (pre-vaccine IgG antibody concentration) | -1.393e-01 | 6.505e-02 | < 0.0001 |

| | | | |
|---|---|---|---|
| $\beta_{37}$ (pre-vaccine IgG antibody concentration, rVSV arm) | -6.362e-02 | 7.668e-02 | 0.032 |
| $\beta_{38}$ (pre-vaccine IgG antibody concentration, rVSV-booster arm) | 2.966e-4 | 1.082e-4 | 0.407 |
| **Random effects and residual error** | **Variance** | **Std.Dev** | |
| $\omega_0^2$ (variance $\gamma_{0i}$) | 0.203657 | 0.45128 | |
| $\omega_1^2$ (variance $\gamma_{1i}$) | 0.001351 | 0.03675 | |
| $\sigma^2$ (variance of residual error) | 0.082684 | 0.28755 | |

**Table S14. Linear mixed effect regression model parameter estimates**
Parameter estimates of the linear mixed effect regression model, estimated on $\log_{10}$ anti-EBOV $GP_{1,2}$ IgG antibody concentrations in all participant groups (1,837 participants)

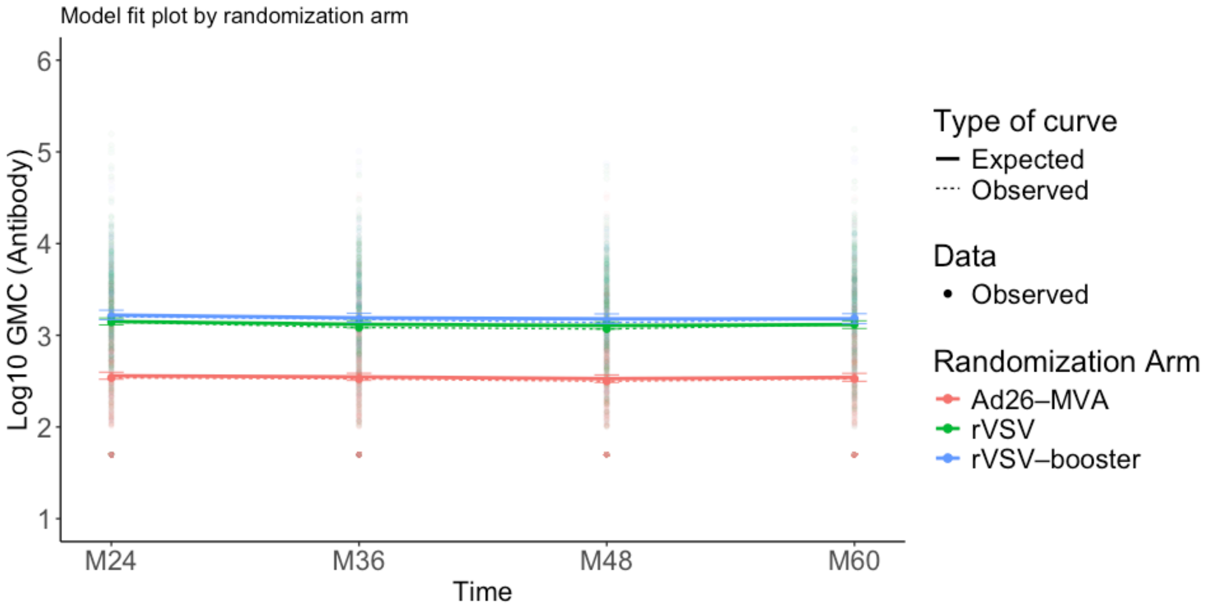

**Figure S21. Modelling of anti-EBOV $GP_{1,2}$ IgG concentration (EU/mL) from 24 months post-vaccination to 60 months according to vaccine group**

Observed and modelled (multivariable model) antibody GMCs over time according to vaccine arm in overall population (adults and children) I bars indicate 95% confidence intervals.

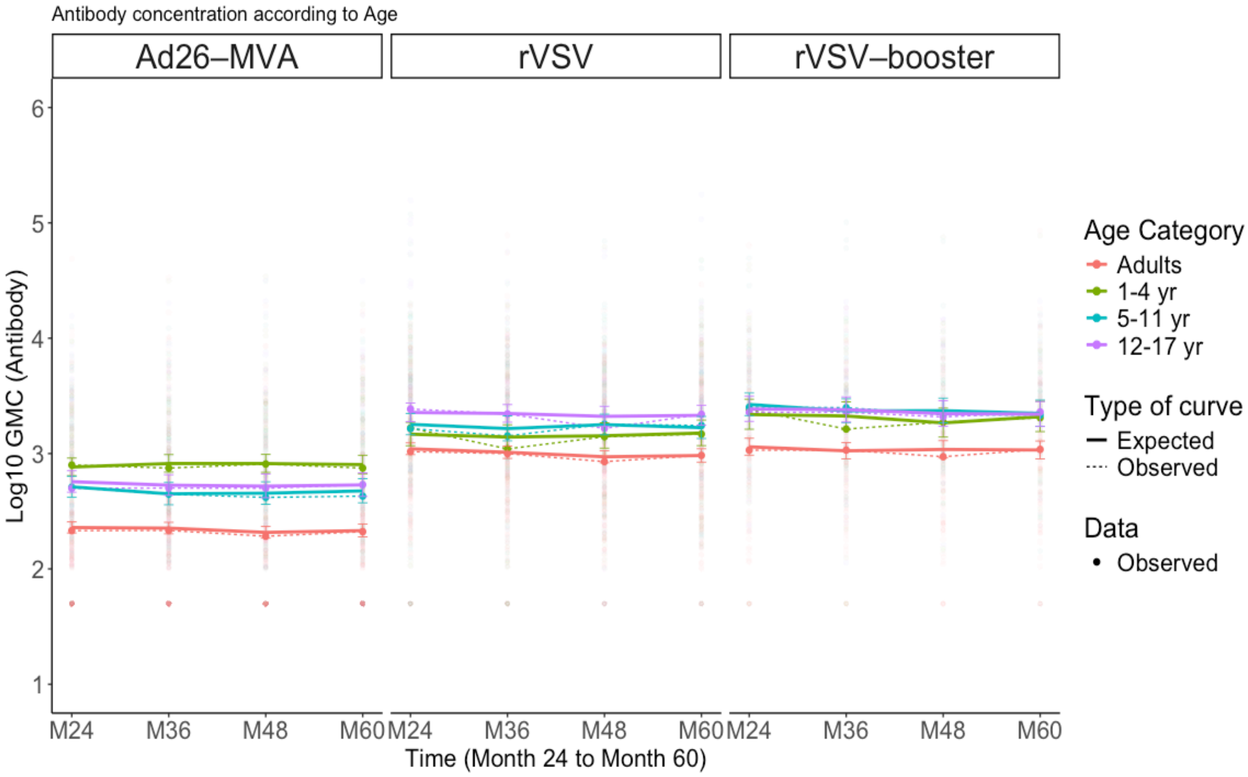

**Figure S22. Modelling of anti-EBOV $GP_{1,2}$ IgG concentration (EU/mL) from 24 months post-vaccination to 60 months, according to age category in Ad26–MVA, rVSV and rVSV–booster groups**

GMC of antibodies observed and modelled (multivariable models) over time in each vaccinated group according to age category. Dots represent individually observed serum samples. I bars indicate 95% confidence intervals.

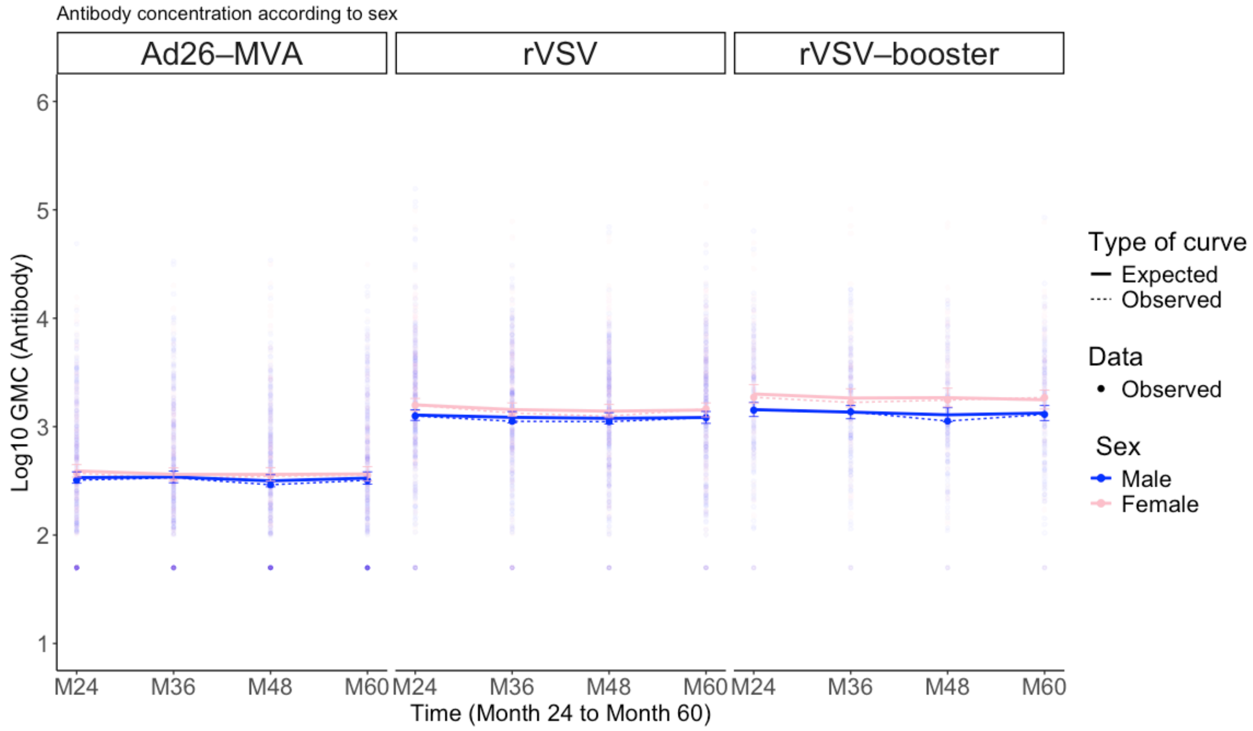

**Figure S23. Modelling of anti-EBOV $GP_{1,2}$ IgG concentration (EU/mL) from 24 months post-vaccination to 60 months, according to sex in Ad26–MVA, rVSV and rVSV–booster groups**

GMC of antibodies observed and modelled (multivariable models) over time in each vaccinated group according to sex. Dots represent individually observed serum samples. I bars indicate 95% confidence intervals.

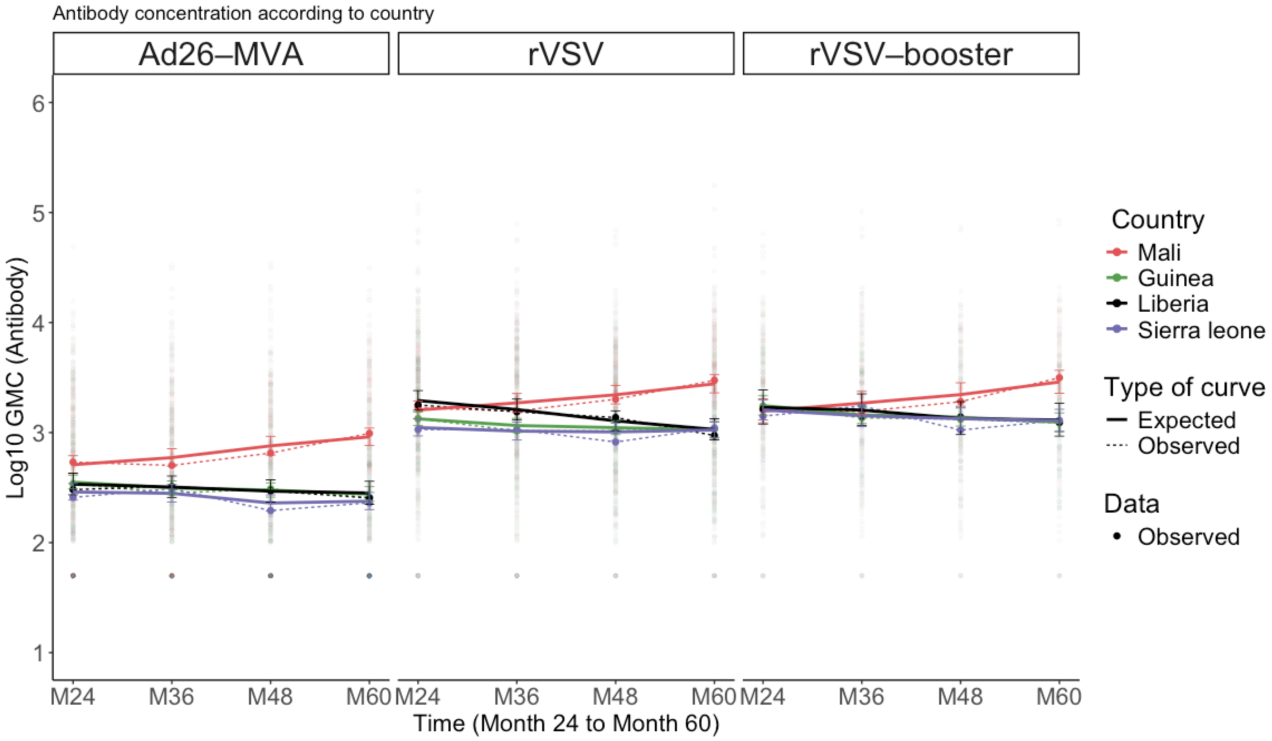

**Figure S24. Modelling of anti-EBOV $GP_{1,2}$ IgG concentration (EU/mL) from 24 months post-vaccination to 60 months, according to country in Ad26–MVA, rVSV and rVSV–booster groups**

GMC of antibodies observed and modelled (multivariable models) over time in each vaccinated group according to country. Dots represent individually observed serum samples. I bars indicate 95% confidence intervals.

#### S3.4. Safety

##### Table S15. Serious Adverse Events or Death in Adults and Children post-M12 to M60, including active vaccinations in the initial placebo group

| | N Pts | N Pts w/ SAE | % Pts w/ SAE | N Pts w/ SAE related to the vaccination | % |
|---|---|---|---|---|---|
| **Adults** | | | | | |
| Ad26, MVA Group | 381 | 15 | 3.94 | .. | .. |
| rVSV Group | 405 | 22 | 5.43 | .. | .. |
| rVSV–Booster Group | 187 | 10 | 5.35 | .. | .. |
| Placebo Group | 412 | 17 | 4.13 | 1 | 0.24 |
| *Not vaccinated* | *74* | *3* | *4.05* | .. | .. |
| *Ad26, MVA** | *204* | *13* | *6.37* | *1* | *0.06* |
| *rVSV** | *134* | *1* | *0.75* | .. | .. |
| **Children** | | | | | |
| Ad26, MVA Group | 396 | 8 | 2.02 | .. | .. |
| rVSV Group | 410 | 8 | 1.95 | .. | .. |
| rVSV–Booster Group | 198 | 5 | 2.53 | .. | .. |
| Placebo Group | 388 | 8 | 2.06 | .. | .. |
| *Not vaccinated* | 71 | 4 | 5.63 | .. | .. |
| *Ad26, MVA** | 199 | 4 | 2.01 | .. | .. |
| *rVSV** | 118 | 0 | 0 | .. | .. |

Pts = participants
* number of participants with SAE post-M12 regardless of the timing of occurrence (prior of after vaccination)

##### Table S16. Number of Serious Adverse Events, post-M12 to M60, before and after vaccination, in the participants vaccinated in the initial placebo group

| | Number of SAE |
|---|---|
| **Adults** | |
| Before vaccination | 12 |
| After rVSV vaccination | 0 |
| After Ad26 vaccination* | 3** |
| **Children** | |
| Before vaccination | 5 |
| After rVSV vaccination | 0 |
| After Ad26 vaccination* | 1 |

*Events occurred after Ad26 vaccination and prior to MVA vaccination
** 1 of these SAEs was considered as possibly related to vaccination (ischaemic stroke in a participant aged aged in their 30s, occurred 49 days after Ad26 administration).

#### Table S17: Incidence of SAEs by MedDRA System Organ Class: Adults post-M12 to M60

| | Ad26, MVA Group | rVSV Group | rVSV Booster Group | Placebo Group |
|---|---|---|---|---|
| **Number of participants by SOC - Adults, n (%)** | 17 | 23 | 10 | 17 |
| Blood and lymphatic system disorders; pregnancy, puerperium and perinatal conditions | 0 (0.0) | 1 (4.3) | 0 (0.0) | 1 (5.9) |
| Cardiac disorders | 0 (0.0) | 0 (0.0) | 1 (10.0) | 1 (5.9) |
| Congenital, familial and genetic disorders | 0 (0.0) | 0 (0.0) | 1 (10.0) | 0 (0.0) |
| Gastrointestinal disorders | 2 (11.8) | 1 (4.3) | 1 (10.0) | 3 (17.6) |
| Gastrointestinal disorders; gastrointestinal disorders | 0 (0.0) | 1 (4.3) | 0 (0.0) | 0 (0.0) |
| Gastrointestinal disorders; gastrointestinal disorders; gastrointestinal disorders | 0 (0.0) | 0 (0.0) | 0 (0.0) | 1 (5.9) |
| Gastrointestinal disorders; infections and infestations | 0 (0.0) | 0 (0.0) | 0 (0.0) | 1 (5.9) |
| Gastrointestinal disorders; infections and infestations; gastrointestinal disorders | 1 (5.9) | 0 (0.0) | 0 (0.0) | 0 (0.0) |
| General disorders and administration site conditions | 2 (11.8) | 3 (13.0) | 0 (0.0) | 0 (0.0) |
| Hepatobiliary disorders | 0 (0.0) | 1 (4.3) | 0 (0.0) | 0 (0.0) |
| Infections and infestations | 2 (11.8) | 4 (17.4) | 0 (0.0) | 2 (11.8) |
| Infections and infestations; gastrointestinal disorders | 1 (5.9) | 0 (0.0) | 0 (0.0) | 0 (0.0) |
| Infections and infestations; injury, poisoning and procedural complications | 0 (0.0) | 0 (0.0) | 0 (0.0) | 1 (5.9) |
| Infections and infestations; reproductive system and breast disorders | 1 (5.9) | 0 (0.0) | 0 (0.0) | 0 (0.0) |
| Injury, poisoning and procedural complications | 1 (5.9) | 1 (4.3) | 2 (20.0) | 3 (17.6) |
| Injury, poisoning and procedural complications; injury, poisoning and procedural complications | 1 (5.9) | 2 (8.7) | 0 (0.0) | 0 (0.0) |
| Neoplasms benign, malignant and unspecified (incl cysts and polyps) | 2 (11.8) | 1 (4.3) | 0 (0.0) | 0 (0.0) |
| Neoplasms benign, malignant and unspecified (incl cysts and polyps); neoplasms benign, malignant and unspecified (incl cysts and polyps) | 1 (5.9) | 0 (0.0) | 0 (0.0) | 0 (0.0) |
| Nervous system disorders | 1 (5.9) | 3 (13.0) | 0 (0.0) | 2 (11.8) |
| Pregnancy, puerperium and perinatal conditions | 0 (0.0) | 2 (8.7) | 4 (40.0) | 1 (5.9) |
| Psychiatric disorders | 0 (0.0) | 0 (0.0) | 0 (0.0) | 1 (5.9) |
| Reproductive system and breast disorders | 0 (0.0) | 1 (4.3) | 0 (0.0) | 0 (0.0) |
| Respiratory, thoracic and mediastinal disorders | 1 (5.9) | 0 (0.0) | 1 (10.0) | 0 (0.0) |
| Surgical and medical procedures | 1 (5.9) | 0 (0.0) | 0 (0.0) | 0 (0.0) |
| Vascular disorders | 0 (0.0) | 2 (8.7) | 0 (0.0) | 0 (0.0) |

#### Table S18: Incidence of SAEs by MedDRA System Organ Class: Children post-M12 to M60

| | Ad26, MVA Group | rVSV Group | rVSV Booster Group | Placebo Group |
|---|---|---|---|---|
| **Number of participants by SOC - Children, n (%)** | 9 | 8 | 5 | 8 |
| Blood and lymphatic system disorders | 0 (0.0) | 0 (0.0) | 0 (0.0) | 1 (12.5) |
| Blood and lymphatic system disorders; infections and infestations | 2 (22.2) | 0 (0.0) | 0 (0.0) | 0 (0.0) |
| Blood and lymphatic system disorders; infections and infestations; nervous system disorders | 0 (0.0) | 1 (12.5) | 0 (0.0) | 0 (0.0) |
| Gastrointestinal disorders | 0 (0.0) | 0 (0.0) | 2 (40.0) | 1 (12.5) |
| General disorders and administration site conditions | 0 (0.0) | 0 (0.0) | 0 (0.0) | 1 (12.5) |
| Infections and infestations | 3 (33.3) | 1 (12.5) | 1 (20.0) | 1 (12.5) |
| Infections and infestations; infections and infestations; blood and lymphatic system disorders | 1 (11.1) | 0 (0.0) | 0 (0.0) | 0 (0.0) |
| Infections and infestations; metabolism and nutrition disorders; metabolism and nutrition disorders | 0 (0.0) | 0 (0.0) | 0 (0.0) | 1 (12.5) |
| Injury, poisoning and procedural complications | 1 (11.1) | 0 (0.0) | 1 (20.0) | 1 (12.5) |
| Injury, poisoning and procedural complications; infections and infestations | 0 (0.0) | 0 (0.0) | 1 (20.0) | 0 (0.0) |
| Injury, poisoning and procedural complications; injury, poisoning and procedural complications | 0 (0.0) | 1 (12.5) | 0 (0.0) | 0 (0.0) |
| Nervous system disorders; injury, poisoning and procedural complications | 0 (0.0) | 1 (12.5) | 0 (0.0) | 0 (0.0) |
| Pregnancy, puerperium and perinatal conditions | 0 (0.0) | 2 (25.0) | 0 (0.0) | 2 (25.0) |
| Psychiatric disorders | 1 (11.1) | 1 (12.5) | 0 (0.0) | 0 (0.0) |
| Psychiatric disorders; psychiatric disorders | 0 (0.0) | 1 (12.5) | 0 (0.0) | 0 (0.0) |
| Respiratory, thoracic and mediastinal disorders | 1 (11.1) | 0 (0.0) | 0 (0.0) | 0 (0.0) |

##### Table S19. Incidence of death during the long-term follow up post-M12 to M60

| | Ad26, MVA Group | | rVSV Group | | rVSV–Booster Group | | Placebo Group | | Total | |
|---|---|---|---|---|---|---|---|---|---|---|
| **Death, n (%)** | | | | | | | | | | |
| Adults | 6 | (1.6) | 6 | (1.5) | 3 | (1.6) | 2 | (0.5) | 17 | (1.2) |
| Children | 2 | (0.5) | 2 | (0.5) | 0 | (0.0) | 1 | (0.3) | 5 | (0.4) |

##### Table S20. Description of causes of death in Adults and Children

| Vaccine group | N | Cause of death (after M12) |
|---|---|---|
| **Adults** | | |
| Ad26, MVA Group | | |
| | 1 | Respiratory distress |
| | 1 | Cerebrovascular accident |
| | 2 | Unknown |
| | 1 | Malaria |
| | 1 | Shortness of breath restlessness |
| Placebo Group | | |
| | 1 | Open fracture of inferior members and head trauma |
| | 1 | Myocardial infarction |
| rVSV Booster Group | | |
| | 1 | Pulmonary oedema |
| | 1 | Road traffic accident |
| | 1 | Terrorist attack |
| rVSV Group | | |
| | 1 | Severe hypoglycemia |
| | 1 | Respiratory failure |
| | 4 | Unknown |
| **Children** | | |
| Ad26, MVA Group | | |
| | 1 | Respiratory distress |
| | 1 | Malaria |
| Placebo Group | | |
| | 1 | Sickle cell crisis |
| | 1 | Drowning |
| rVSV Group | | |
| | 1 | Closed cervical trauma with tetraplegia |

[1] PREVAC Study Team, Kieh M, Richert L, et al. Randomized trial of vaccines for Zaire Ebola virus disease. N Engl J Med. 2022 Dec 29;387(26):2411–2424. doi:10.1056/NEJMoa2200072

[2] Berry IM, Farhat SB, Callier V, et al. Comparison of Four Assays That Measure Antibodies to Ebola Virus Glycoprotein. bioRxiv. Preprint posted online March 20, 2026:2026.03.18.708022. doi:10.64898/2026.03.18.708022
